# DISCO: A Delta-NIHSS-Based Machine Learning Model for Predicting Recurrence, Disability, and Mortality Following Acute Ischemic Stroke

**DOI:** 10.1101/2025.05.11.25326629

**Authors:** Shiyao Cheng, Yuandan Wei, Huaguang Zheng, Qingrong Zhang, Xuanyan Yang, Zhen Lu, Yanfeng Shi, Jie Zhang, Cang Guo, Xiaoling Liao, Jianping Wu, Xia Meng, Zixiao Li, Hao Li, Yongjun Wang, Si Cheng, Siyang Liu

## Abstract

**Background:** An accurate, robust, clinically accessible, and explainable predictive model for post-stroke composite outcomes could identify high-risk patients for targeted interventions. However, such a model is currently lacking. This study leverages artificial intelligence to develop and validate a predictive model for post-stroke outcomes at three months and over five years, leveraging comprehensive data from the third China National Stroke Registry (CNSR-III), one of China’s largest nationwide multi-center ischemic stroke registries with five-year follow-up and the CHANCE-2 trial, a genotype-guided dual anti-platelet therapy trial in China.

**Methods:** We evaluated 309 hospitalization variables, including baseline characteristics, medical history, hospitalization data, biomarkers, geographical factors, NIHSS/mRS scores, and stroke polygenic risk scores (PRS), using an extreme gradient boosting tree model. Feature importance was assessed via Shapley values. Primary outcomes were three-month stroke recurrence (5.6%), disability (mRS > 2, 13.75%), and mortality (1.18%). Secondary outcomes were assessed at six additional time points over five years. A nested cross-validation scheme was employed for feature selection and internal validation in 80% of patients (n=11,313) from CNSR-III cohort. External validation of the model was performed in the remaining 20% patients (n=2,627) from CNSR-III cohort and in CHANCE-2 trail (n=5,158).

**Results:** Global and domain-specific delta-NIHSS_(admission-discharge)_ emerged as the strongest predictor of stroke recurrence, disability and mortality. The Delta-NIHSS-Based Predictor for Post-Stroke Composite Outcomes (DISCO) model, integrating 16 delta-NIHSS_(admission-to-discharge)_ and 8 clinically accessible variables, achieved AUCs > 0.8 for recurrence and disability and > 0.9 for mortality at three months. The highest-risk 1% patients exhibited a >10-fold relative risk (RR) for recurrence, >30-fold RR for disability, and >100-fold RR for mortality at three months. The DISCO model is clinically accessible at http://www.discosysu.cn.

**Conclusion:** The DISCO model, incorporating 24 readily obtainable clinical variables, demonstrates high accuracy, robustness, clinical accessibility, and explainability in predicting post-stroke outcomes. The predictive strength of delta-NIHSS_(admission-discharge)_ provides mechanistic insights into stroke outcomes and informs future acute stroke treatment and rehabilitation strategies.

## Introduction

Stroke is a leading cause of death and long-term disability worldwide, with high rates of recurrence, mortality, and disability, particularly in underdeveloped regions.^1^ Effective prediction models for post-stroke outcomes are crucial for guiding clinical decision-making by identifying high-risk patients who may benefit from targeted evaluation and intervention. Existing statistical tools, such as the Recurrence Risk Estimator (RRE) for early stroke recurrence,^2^ the ASTRIL score for functional outcomes,^3^ the IScore for stroke mortality^4^, and the ABCD score for transient ischemic attack (TIA) patients^5^, rely on clinical, imaging, or laboratory variables and are developed using multivariable regression models. However, regression-based models are susceptible to multicollinearity and fail to optimize the complex, nonlinear relationships between variables and outcomes, which limits their predictive accuracy and robustness. Moreover, these models depend on imaging and laboratory data that may not be readily available in resource-limited settings, thereby restricting their broader clinical applicability.

Machine learning (ML) techniques have gained increasing attention in clinical prediction modelling due to their ability to address nonlinear relationships between variables and outcomes, capture complex variable interactions, and improve predictive accuracy^6,7^. Several ML-based models, such as early warning systems for hypotension^8^, have been successfully implemented in clinical practice with support from clinical trials. However, ensuring the transparency and interpretability of ML models is essential, as their lack thereof may hinder widespread clinical adoption.^9,10^ A systematic review of ML-based models for stroke recurrence in PubMed and Web of Science revealed 12 models (**Expanded Methods, Table S1**). Most exhibited suboptimal predictive performance, with only four achieving an AUC greater than 0.8 (developed by Kai Wang^11^, Ji lv^12^, Hao Wang^13^ and Yu Gao^14^). Notably, none of these models underwent external validation, raising concerns regarding their accuracy and generalizability. Furthermore, clinical accessibility remains a substantial challenge, as many of these models rely on limited feature sets and lack publicly available applications or source codes. Additionally, existing ML-based models focus on stroke recurrence and have not incorporated long-term outcomes such as disability (measured by the mRS) and mortality (**eTable 1**), further compromising their practical applicability for individualized patient care.

In this study, we aim to develop a clinical prediction model with high accuracy, robustness, clinical accessibility, and explainability for predicting post-stroke composite outcomes, including stroke recurrence, disability and mortality at three months and over five years. We employed explainable artificial intelligence (AI) techniques to evaluate the predictive importance of 309 clinical features, most of which are readily available during hospitalization, among 11,813 ischemic stroke patients in the CNSR-III, a nationwide multi-center registry of ischemic stroke and transient ischemic attack (TIA) in China^15,16^. Through this approach, we identified previously unrecognized predictors of post-stroke outcomes.

Based on these findings and a rigorous internal-external validation framework, we developed the DISCO model, which integrates 24 clinically accessible features. The model was externally validated using data from remaining 2,627 ischemic stroke patients in CNSR-III, and in 5,158 ischemic stroke patients enrolled in the CHANCE-2 clinical trial^17^. We assessed the model’s risk stratification capability in both the development and validation cohorts, and developed a user-friendly R Shiny web application (http://www.discosysu.cn/) to provide individualized risk predictions and explanations.

## Methods

### Study population

A total of 15,166 patients were enrolled in CNSR-III, a nationwide registry of acute ischemic stroke or transient ischemic attack (TIA) in China between August 2015 and March 2018.^15^ Exclusion criteria included 7 patients experienced stroke recurrence during hospitalization, 35 patients dead during hospitalization, and 1,184 diagnosed with TIA. The development cohort comprised 11,313 patients, while validation cohort 1 included 2,627 patients, selected randomly based on hospital assignment. Validation cohort 2 involved 5,158 ischemic stroke patients from the CHANCE-2 trial, excluding those diagnosed with TIA.^17^ Polygenic risk scores (PRS) were calculated for 9,402 stroke patients from CNSR-III with whole-genome sequencing (WGS) data (Figure 1).^16^ More detailed information on participant recruitment and informed consent procedures is provided in the CNSR-III and CHANCE-2 studies.^15,17^

**Figure 1.**
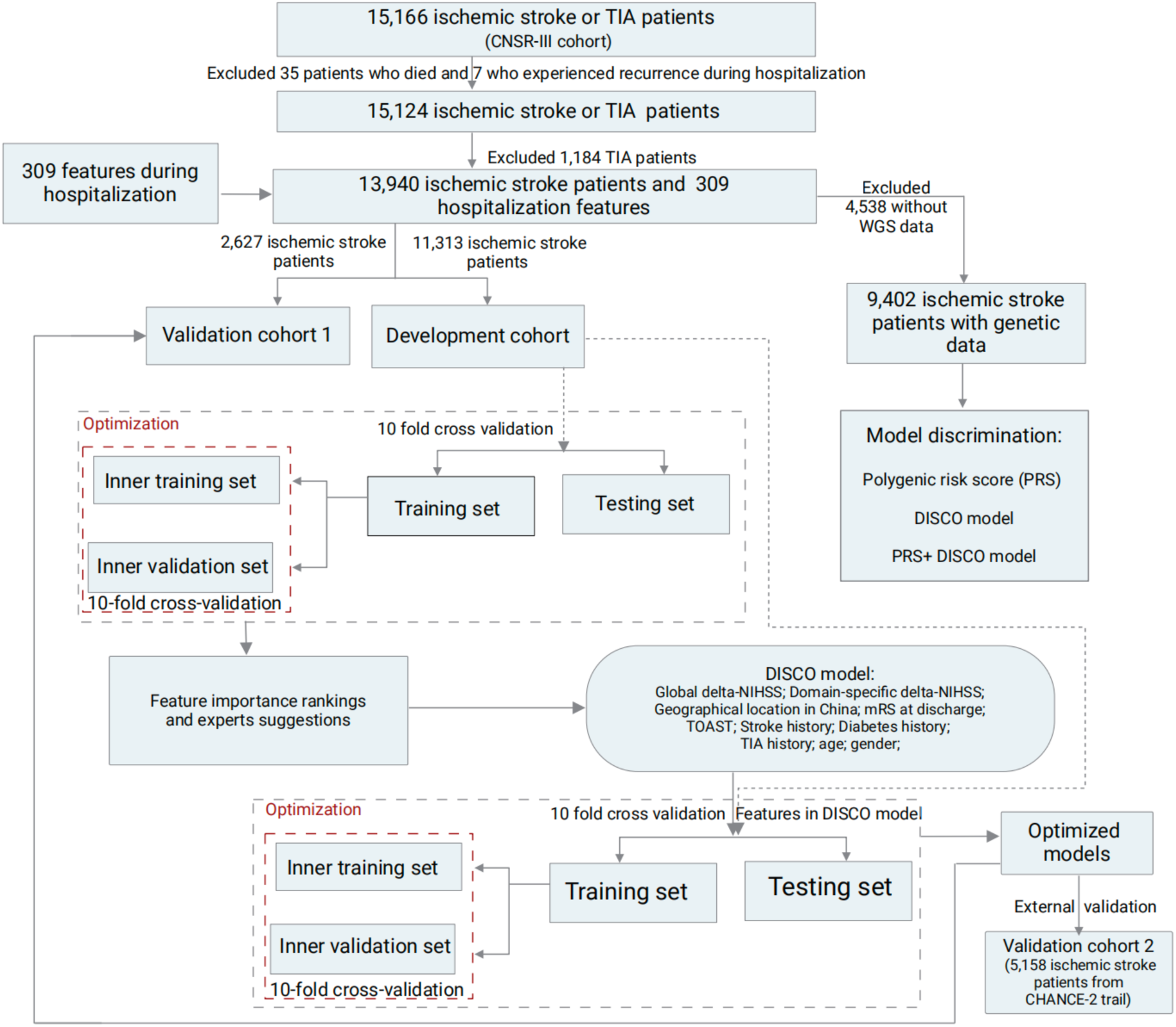
Workflow for Predictive Model Development. TIA refers to transient ischemic attack. In validation cohort 2, the CHANCE-2 trail initially enrolled 6,412 patients with stroke or TIA. After excluding TIA patients, 5,158 stroke patients remained.

### Candidate predictors

Baseline data, along with biological and urine samples, were collected by physicians through face-to-face interview in CNSR-III. A total of 309 candidate predictors were collected, categorized into the following groups: basic information, disease history, hospitalization information, biomarkers, geographical location, NIHSS/mRS score, and PRS. For the disease history variables, missing values were assumed to indicate the absence of the event, while missing values for biomarkers were retained as missing. When developing the model, missing values will be filled by XGBoost’s default direction method. Detailed descriptions and statistical summaries of the 309 predictors are provided in **Table S2**. Additionally, the 3-month recurrence risk was calculated using the RRE model within the CNSR-III cohort (Described in **Table S3**, **Expanded Methods**). Due to the unavailability of genetic scores specific to stroke recurrence, we used the PRS for stroke susceptibility from the GIGASTROKE study of East Asians as a proxy^18^(**Expanded Methods**).

### Definition of post-stroke outcomes

Patients in the CNSR-III were followed up at 3 months, 6 months, and annually from 1 to 5 years, with follow-up completion rates exceeding 90% in 2 years and around 80% in 5 years (**Table S4**). Follow-up assessments were conducted within a 14-day window from each scheduled time point. Patients were interviewed face-to-face by physicians within the first year, followed by telephone interviews conducted by trained clinical research coordinators thereafter. Regardless of the mode of follow-up, patient evaluations adhered to the standardized operational manual of the follow-up case report form (CRF) (Details in **Expanded Methods**).

The primary outcomes included stroke recurrence, all-cause mortality, and functional outcomes at 3 months. Stroke recurrence was defined as any new stroke event identified through readmission records or patient self-reports during follow-up. Functional outcomes were defined using a binary modified Rankin Scale (mRS) classification, with mRS 0-2 considered non-events, and mRS 3-6 deemed as events. Each fatality was confirmed using the Population Mortality Registration and Management Information System within the Life Registration Information Management System of the Chinese Center for Disease Control and Prevention. In the assessment of stroke recurrence, if a patient died from a stroke, the event was classified as stroke recurrence. Death from non-stroke causes, such as cancer, was considered a non-event in the analysis. In the assessment of disability, patients who died during the observation period were excluded from the analysis. In the CHANCE-2 trial, follow-ups were conducted at 3 months and 1 year using similar evaluation methods.

### Model construction, evaluation, and explanation

The extreme gradient boosting tree (XGBoost) algorithm^19^ was used for model construction due to its scalability and effectiveness in handling nonlinear, sparse, and class-imbalanced classification data. To enhance model robustness and prevent overfitting, tenfold cross-validation was employed, along with an inner tenfold cross-validation for hyperparameter tuning through a random grid search. The optimization criterion was the area under the receiver operating curve (AUC) (**Figure 1**). This nested cross-validation approach provides more robust and unbiased results, as demonstrated in previous studies.^19–21^ Using this methodology, models were initially developed using all 309 candidate features. Based on feature importance rankings and expert recommendations, 24 features were selected for the final model, incorporating 16 global and domain-specific delta-NIHSS_(admission-discharge)_ variables. Delta-NIHSS_(admission-discharge)_ is defined as total NIHSS change from admission to discharge. The model was termed DISCO (**D**elta-N**I**HSS-Based Predictor for Post-**S**troke **C**omposite **O**utcomes). To enhance interpretability, Shapley additive explanations (SHAP) values^21^ were used to assess feature importance, complementing XGBoost’s importance scores to guide feature selection and provide individualized explanations for each feature’s contribution.

The DISCO model was trained using the development cohorts and externally validated in validation cohorts 1 and 2. Model performance was evaluated using sensitivity, specificity, F1-score, and AUC, while model calibration was evaluated using the Brier score, with 95% confidence intervals (CIs) derived from 200 bootstrap samples. To benchmark XGBoost’s performance, the DISCO model’s feature set was applied to other algorithms, including logistic regression, naive Bayes, random forest, and support vector machine. A head-to-head comparison with the RRE model in the CNSR-III cohort was also performed (**Expanded Methods**). DeLong’s test was used to compare the AUC between DISCO model and RRE model.^22^ All statistical analyses were performed in R version 4.2.0. A detailed description of the model construction process and software specifications is provided in the **Expanded**

## Methods

### Sensitivity analysis

To evaluate the robustness of the DISCO model across different clinical scenarios, sensitivity analyses were conducted under the following two conditions:

1. **Scenario 1**: Stroke patients admitted to the hospital within 48 hours of stroke onset and with a hospital stay of equal or fewer than 14 days (n=8,262).
2. **Scenario 2**: Stroke patients who did not undergo intravenous thrombolysis with rt-PA (TPA) or mechanical thrombectomy (n=12,681).

### Factors associated with delta-NIHSS

Given the strong predictive power of delta-NIHSS_(admission-discharge)_ for stroke recurrence, disability, and mortality, we examined factors influencing delta-NIHSS_(admission-discharge)_ in the full CNSR-III analysis, and two sensitivity scenarios, using multivariable logistic regression. Delta-NIHSS_(admission-discharge)_ was dichotomized into two categories: delta-NIHSS_(admission-discharge)_ ≥ 0 (non-events) and delta-NIHSS_(admission-discharge)_ < 0 (events).

Associations between baseline clinical variables and both global and domain-specific delta-NIHSS_(admission-discharge)_ were assessed. Additionally, z-transformed biomarkers were analyzed to explore potential biological mechanisms underlying delta-NIHSS_(admission-discharge)_ variation. Biomarkers with significant missingness (>5,000 missing values), and those exhibiting strong collinearity (assessed via variance inflation factor [VIF]) were excluded. A total of 38 biomarkers were included in the final regression analysis. To ensure robustness, sensitivity analyses were conducted within the two predefined subgroups. The detailed workflow is presented in **Figure S1**.

### Risk stratification

Risk stratification for stroke recurrence, mRS, and mortality at 3 months was assessed using relative risks (RRs) across the development cohort and two validation cohorts. RRs were calculated using the formula shown below (Equation 1), with patients in the 0-60% risk category serving as the reference group:

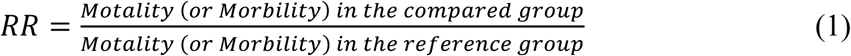

### Development of web application

To facilitate clinical application of the DISCO model, we developed an R Shiny web application, available at http://www.discosysu.cn/. This tool allows users to evaluate individual risk based on the 24 features in the DISCO model and provides personalized explanations of the predicted risk using SHAP values for enhanced interpretability.

### Results Study sample

Among the 11,313 patients in the development cohort, 2,627 patients in validation cohort 1, and 5,158 patients in validation cohort 2, the median ages were 62, 63, and 65 years, respectively. The proportion of female participants was 31.10%, 32.74%, and 32.80%, respectively, and the median NIHSS scores at admission was 3, 3 and 2 (**Table 1**). For mechanical thrombectomy, the proportions were 0.31%, 0.15%, and 0% respectively and they were 8.94%, 8.37%, and 0% for thrombolysis. According to the TOAST classification, most patients were diagnosed with large artery atherosclerosis (LAA), small vessel disease (SVS), or stroke of undetermined cause. At the 3-month follow-up, stroke recurrence rates ranged from 5.60% to 6.90% across the development and validation cohorts. Disability rates ranged from 2.85% to 17.05%, while mortality rates ranged from 0.48% to 1.75%. Rates of these three stroke outcomes during the subsequent 6-month follow-up, and annual follow-ups from years 1 to 5 are detailed in **Table S5**. Cumulative incidences curve for stroke recurrence at 3 months are presented in **Figure S2**.

**Table 1.**
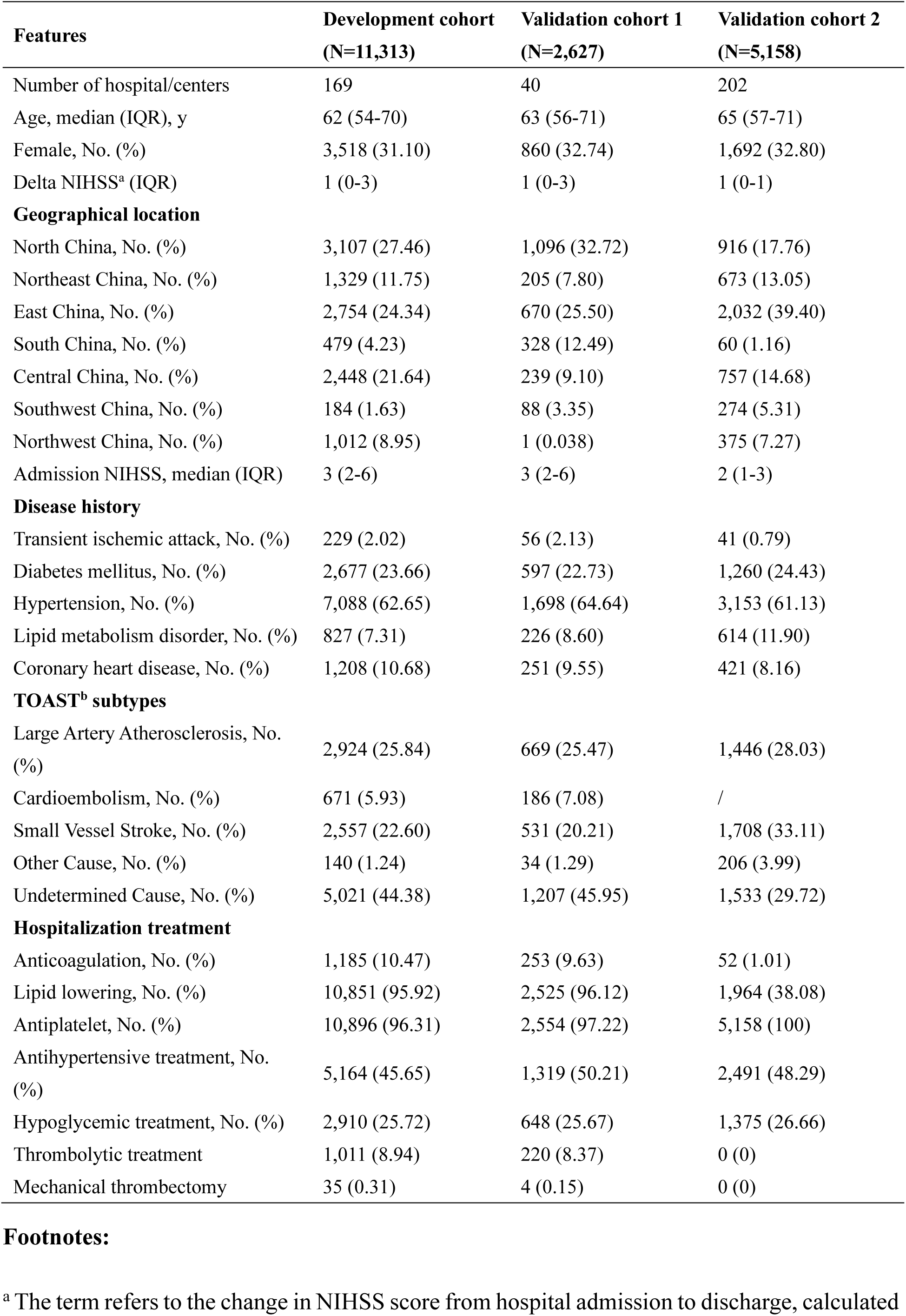
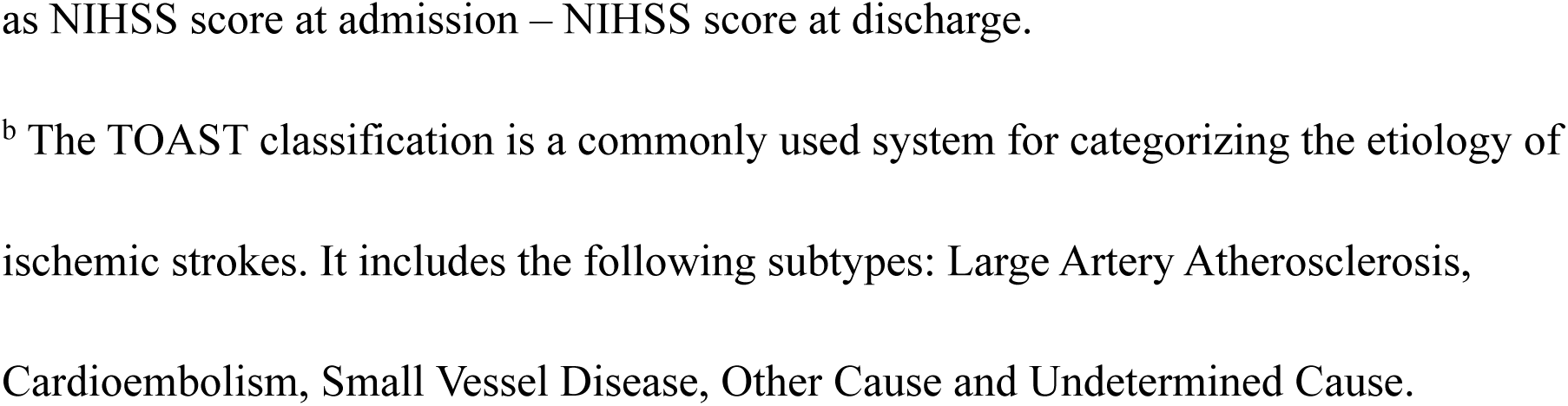
Baseline Characteristics of the Study Cohorts.

| Features | Development cohort<br>(N=11,313) | Validation cohort 1<br>(N=2,627) | Validation cohort 2<br>(N=5,158) |
| --- | --- | --- | --- |
| Number of hospital/centers | 169 | 40 | 202 |
| Age, median (IQR), y | 62 (54-70) | 63 (56-71) | 65 (57-71) |
| Female, No. (%) | 3,518 (31.10) | 860 (32.74) | 1,692 (32.80) |
| Delta NIHSS<sup>a</sup> (IQR) | 1 (0-3) | 1 (0-3) | 1 (0-1) |
| **Geographical location** |  |  |  |
| North China, No. (%) | 3,107 (27.46) | 1,096 (32.72) | 916 (17.76) |
| Northeast China, No. (%) | 1,329 (11.75) | 205 (7.80) | 673 (13.05) |
| East China, No. (%) | 2,754 (24.34) | 670 (25.50) | 2,032 (39.40) |
| South China, No. (%) | 479 (4.23) | 328 (12.49) | 60 (1.16) |
| Central China, No. (%) | 2,448 (21.64) | 239 (9.10) | 757 (14.68) |
| Southwest China, No. (%) | 184 (1.63) | 88 (3.35) | 274 (5.31) |
| Northwest China, No. (%) | 1,012 (8.95) | 1 (0.038) | 375 (7.27) |
| Admission NIHSS, median (IQR) | 3 (2-6) | 3 (2-6) | 2 (1-3) |
| **Disease history** |  |  |  |
| Transient ischemic attack, No. (%) | 229 (2.02) | 56 (2.13) | 41 (0.79) |
| Diabetes mellitus, No. (%) | 2,677 (23.66) | 597 (22.73) | 1,260 (24.43) |
| Hypertension, No. (%) | 7,088 (62.65) | 1,698 (64.64) | 3,153 (61.13) |
| Lipid metabolism disorder, No. (%) | 827 (7.31) | 226 (8.60) | 614 (11.90) |
| Coronary heart disease, No. (%) | 1,208 (10.68) | 251 (9.55) | 421 (8.16) |
| **TOAST<sup>b</sup> subtypes** |  |  |  |
| Large Artery Atherosclerosis, No. (%) | 2,924 (25.84) | 669 (25.47) | 1,446 (28.03) |
| Cardioembolism, No. (%) | 671 (5.93) | 186 (7.08) | / |
| Small Vessel Stroke, No. (%) | 2,557 (22.60) | 531 (20.21) | 1,708 (33.11) |
| Other Cause, No. (%) | 140 (1.24) | 34 (1.29) | 206 (3.99) |
| Undetermined Cause, No. (%) | 5,021 (44.38) | 1,207 (45.95) | 1,533 (29.72) |
| **Hospitalization treatment** |  |  |  |
| Anticoagulation, No. (%) | 1,185 (10.47) | 253 (9.63) | 52 (1.01) |
| Lipid lowering, No. (%) | 10,851 (95.92) | 2,525 (96.12) | 1,964 (38.08) |
| Antiplatelet, No. (%) | 10,896 (96.31) | 2,554 (97.22) | 5,158 (100) |
| Antihypertensive treatment, No. (%) | 5,164 (45.65) | 1,319 (50.21) | 2,491 (48.29) |
| Hypoglycemic treatment, No. (%) | 2,910 (25.72) | 648 (25.67) | 1,375 (26.66) |
| Thrombolytic treatment | 1,011 (8.94) | 220 (8.37) | 0 (0) |
| Mechanical thrombectomy | 35 (0.31) | 4 (0.15) | 0 (0) |

<sup>a</sup> The term refers to the change in NIHSS score from hospital admission to discharge, calculated
as NIHSS score at admission – NIHSS score at discharge.
<sup>b</sup> The TOAST classification is a commonly used system for categorizing the etiology of
ischemic strokes. It includes the following subtypes: Large Artery Atherosclerosis,
Cardioembolism, Small Vessel Disease, Other Cause and Undetermined Cause.

### Feature selection

The feature importance of all 309 candidate predictors, assessed using SHAP values and XGBoost importance scores, is presented in **Table S6,** with the top 20 features visualized in **Figure S3**. Notably, the global delta-NIHSS_(admission-discharge)_ emerged as the strongest predictor of stroke recurrence at 3 months (**Figure S3** and **Table S6**), accounting for more than 30% of the model’s predictive power. Additional key predictors included mRS at discharge, molecular biomarkers such as choline and fasting blood glucose, domain-specific delta-NIHSS_(admission-discharge)_, history of disease, and age (**Figure S3** and **Table S6**). In contrast, the PRS for stroke susceptibility, despite being normally distributed, contributed to less than 0.5% to the prediction of 3-month stroke recurrence (**Figure S4, Table S6**), suggesting that distinct genetic factors and biological mechanisms drive stroke occurrence and recurrence risk.

Based on a combination of feature importance rankings and clinical feasibility, as recommended by stroke specialists, we selected 24 features for the final model. These included the global delta-NIHSS_(admission-discharge)_, 15 domain-specific delta-NIHSS_(admission-discharge)_ features, and 8 core clinical variables: mRS at discharge, geographical location, age, gender, TOAST classification, history of stroke, diabetes and TIA. This final 24-feature model was designated as the DISCO model (**Figure 1**).

### Model performance

In internal cross-validation, the DISCO model for predicting 3-month stroke recurrence achieved an AUC of 0.802 (95% CI: 0.743 - 0.861) and an F1 score of 0.950. In the two validation cohorts, the AUCs were 0.743 (0.694 - 0.791) and 0.803 (0.775 - 0.831), respectively (**Figure 2A**). The DSICO model outperformed the RRE model for 3-month recurrence risk, which yielded an AUC of 0.576 (95% CI: 0.557 - 0.596) and an F1 score of 0.837 in CNSR-III (**Figure 2A**). Additional models using all 309 features, revascularization features, global and domain-specific delta-NIHSS_(admission-discharge)_ features, biomarker features, and the top 20 features were also evaluated. The 24-feature DISCO model demonstrated performance comparable to the full 309-feature model (AUC: 0.815; 95% CI: 0.762 - 0.868) for 3-month stroke recurrence (**Figure S5**). Other models exhibited lower AUCs than the DISCO model.

**Figure 2.**
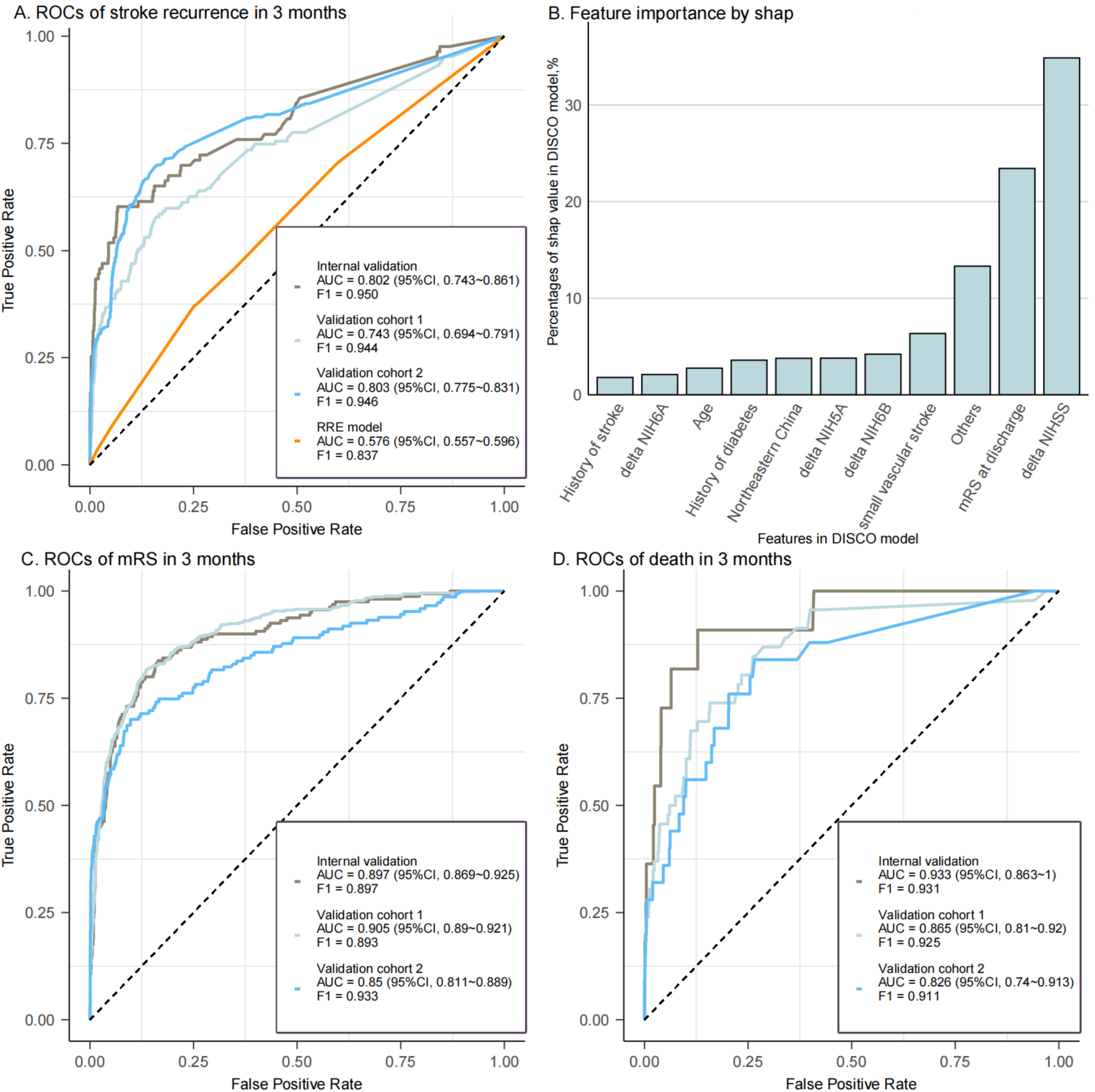
ROC Curves and Feature Importance of the DISCO Model. (A) The ROC curve illustrating the predictive performance of the DISCO model for 3-month stroke recurrence, with the RRE model estimates derived from 13,940 patients in the CNSR-III cohort via the algorithm on the RRE website (https://www.nmr.mgh.harvard.edu/RRE/). (B) Feature importance rankings. Others represents 14 features utilized in DISCO model. (C) The ROC curve for predicting 3-month disability (mRS > 2). (D) The ROC curve for the DISCO model in predicting 3-month mortality.

Delta-NIHSS_(admission-discharge)_ was the most important predictor in the DISCO model, contributing 34.6% to the model’s prediction power (**Figure 2B**). The model showed high calibration performance in validation cohort 1 and 2 (**Figure S6, Figure S7),** though it slightly overestimated 3-month stroke recurrence risk. Decision Curve Analysis indicated a positive net benefit for the DISCO model. The optimal cut-off value in validation cohort 1, based on the maximum Youden index, was 0.106 (sensitivity = 0.585, specificity = 0.833, Youden index = 1.486), higher than that of the RRE model (**Figure S8**). In validation cohort 2, the cut-off value was 0.097 (**Figure S9).** Additionally, the DISCO model demonstrated the highest AUC for predicting 3-month stroke recurrence compared to four other statistical or machine learning algorithms (**Figure S10**). AUCs for internal and external validations, along with parameters for secondary endpoint events, are detailed in **Table S7**.

For predicting 3-month post-discharge disability events, the DISCO model achieved an internal validation AUC of 0.897 (95% CI: 0.869 - 0.925) and external validation AUCs of 0.905 (95% CI: 0.890 – 0.921) and 0.850 (95% CI: 0.811 - 0.889) (**Figure 2C and TableS7**). For predicting 3-month mortality, the DISCO model demonstrated an AUC of 0.933 (95% CI: 0.863 – 1) in internal validation, with AUCs of 0.865 (95% CI: 0.81 - 0.92), and 0.826 (95% CI: 0.74 - 0.913) in the two validation cohorts (**Figure 2D** and **Table S7**).

AUCs decreased with longer follow-up durations (**Table S7**). In the development and the two validation cohorts, AUCs for 1-year stroke recurrence decreased from 0.8 to 0.7. Sensitivity analyses did not show significant AUC reductions for 3-month stroke recurrence in scenarios 1 and 2 (**Figure S11 and Figure S12**). Adding PRS improved AUCs for predicting 3-month stroke recurrence in both development and validation cohort 1 (**Figure S13**).

### Influencing factors of global and domain-specific delta-NIHSS_(admission-discharge)_

To better understand the significant contribution of delta-NIHSS_(admission-discharge)_ in predicting stroke composite outcomes, we investigated the associations between hospitalization factors and both global and domain-specific delta-NIHSS_(admission-discharge)_. In a retrospective analysis, the distribution of NIHSS scores at admission and discharge differed between patients who did not experience stroke recurrence at 3 months (**Figure 3A**) and those who did (**Figure 3B**). Patients without recurrence tended to have a higher proportion of NIHSS scores ≤ 3 at discharge compared to admission, whereas patients with recurrence were more likely to have NIHSS scores > 3 at discharge. In addition, based on NIHSS changes from admission to discharge across different baseline NIHSS scores, patients with worsening delta-NIHSS_(admission-discharge)_ during hospitalization tend to have poorer prognoses (**Table S8**). The distribution of both global and domain-specific delta-NIHSS_(admission-discharge)_ was zero-inflated (**Figure S14**), prompting us to dichotomize patients into events (delta − NIHSS < 0) and non-events (delta − NIHSS ≥ 0).

**Figure 3.**
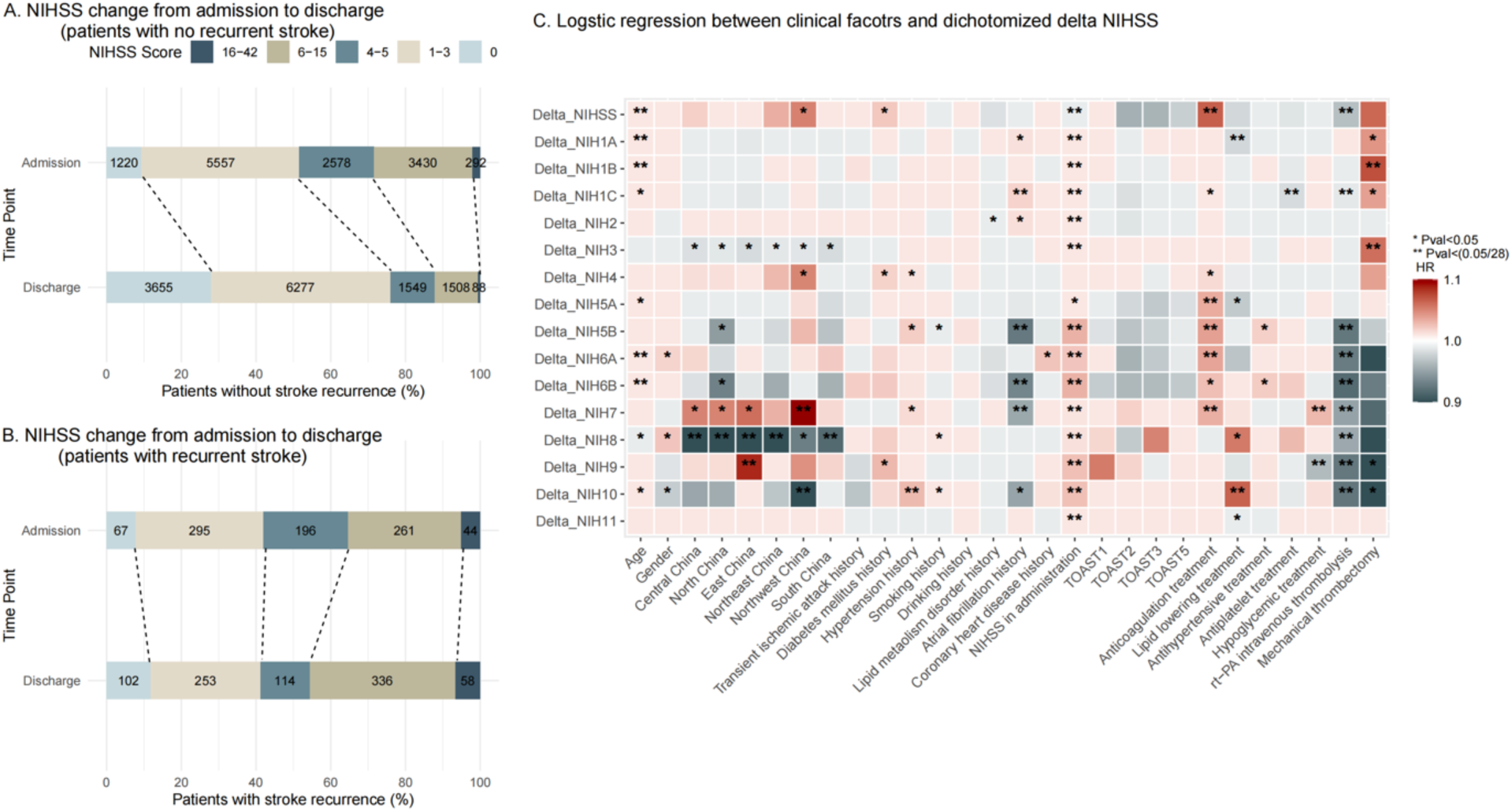
Patterns and Potential Influencing Factors of NIHSS changes in CNSR-III. (A) NIHSS changes from admission to discharge in patients without recurrent stroke. (B) NIHSS changes from admission to discharge in patients with recurrent stroke. (C) Results of multivariable regression analysis showing the associations between baseline clinical variables and dichotomized delta-NIHSS _(admission-discharge)._

When regressing dichotomized global and domain-specific delta-NIHSS_(admission-discharge)_ on 28 hospitalization characteristics, with Bonferroni correction applied, we found that age, geographic location, NIHSS at admission, anticoagulation treatment, TPA treatment, and a history of atrial fibrillation were significantly associated with multiple delta-NIHSS_(admission-discharge)_ domains (**Figure 3C, Table S9**). Additional factors, including lipid-lowering treatments, antiplatelet and hypoglycemic treatments, and mechanical thrombectomy, were significantly associated with specific delta-NIHSS_(admission-discharge)_ domains. In scenario 1 which restricted patients to those who arrived at the hospitals within 48 hours and stayed no more than 14 days, mechanical thrombectomy became more strongly and positively associated with multiple delta-NIHSS_(admission-discharge)_, while rt-PA remained negatively associated with several delta-NIHSS_(admission-discharge)_ (**Figure S15, Table S10**). In scenario 2, after excluding patients who received TPA or mechanical thrombectomy, age, geographical location, NIHSS at admission, anticoagulation treatment, and a history of atrial fibrillation remained the most significant factors associated with dichotomized delta-NIHSS_(admission-discharge)_ (**Figure S16, Table S11**).

At the molecular level, when regressing dichotomized delta-NIHSS_(admission-discharge)_ against 38 biomarkers assayed at the time of patient admission, IL6 demonstrated the most widespread Bonferroni-corrected significance across domain-specific delta-NIHSS_(admission-discharge)_ in all three scenarios (**Figure S17-S19, Table S12-14**).

### Risk stratification and clinical application

The DISCO model demonstrates strong capacity in identifying high-risk patients. For stroke recurrence, disability, and mortality at 3 months, the relative risks (RRs) for patients in the highest risk group (99%-100%) compared to the reference group (0%-60%) were 33.95, 38.07, and 252, respectively (**Figure 4**). The model’s stratification performance was consistent in validation cohort 1 (**Figure S20**), where the RRs for the highest-risk group compared to the reference group patients were 19.71, 32.73, and 166.9, and in validation cohort 2 (**Figure S21**), where the RRs were 35.62, 113.64, and 111.18.

**Figure 4.**
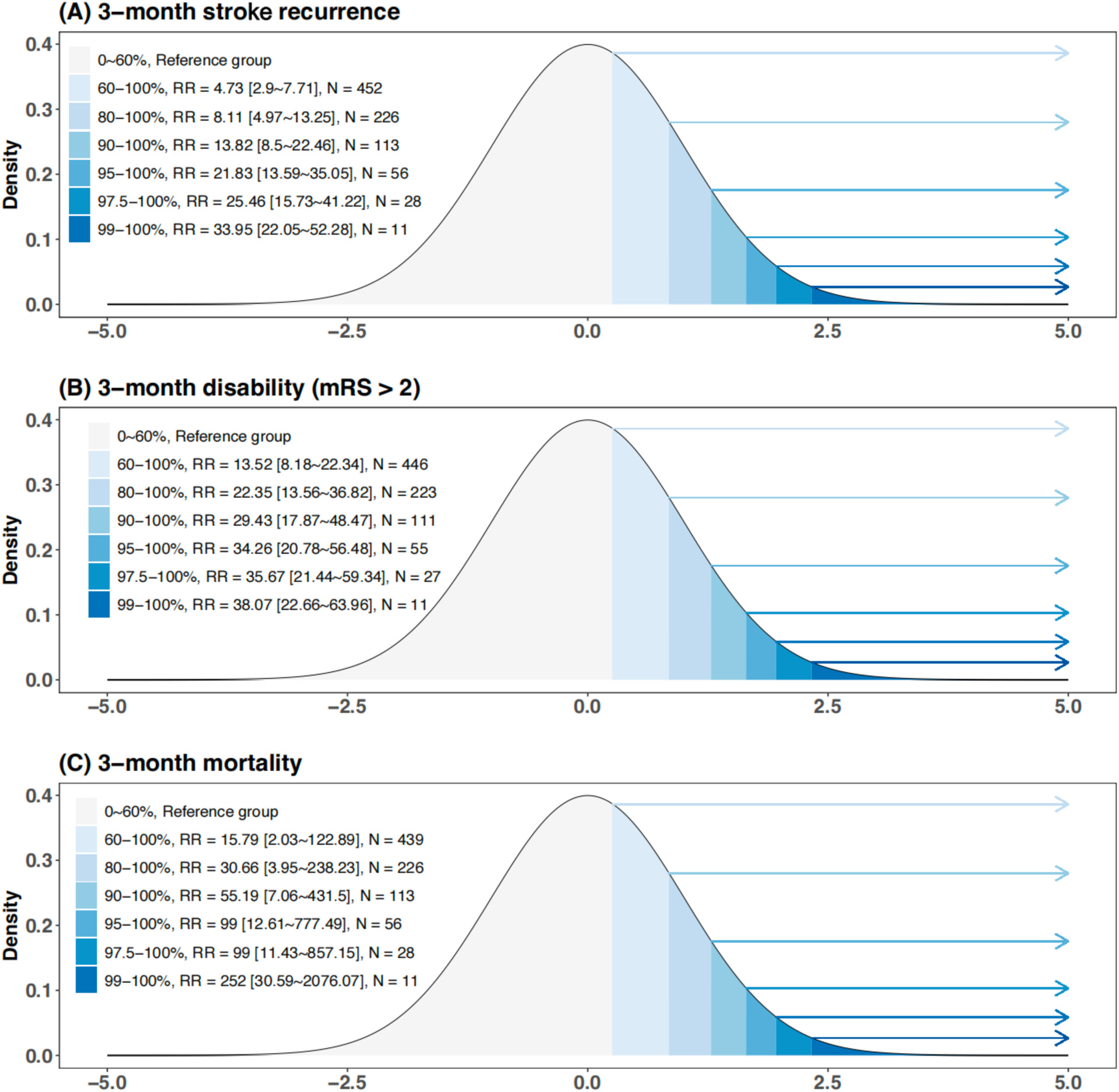
Risk Stratification by the DISCO Model for Three Stroke Outcomes at 3 Months. The figure displays the density distribution of predicted risks across the study population for three stroke outcomes at 3 months. Relative risks (RRs) were calculated using the formula detailed in the Methods section, with the reference group defined as the bottom 60% of patients based on predicted risk.

To illustrate personalized risk assessment, two patients from validation cohort 1 were randomly selected for analysis (**Figure S22**). For Patient A, the delta-NIHSS4 (poor facial palsy, −2) had the highest SHAP value, suggesting the need for focused rehabilitation on facial palsy post-discharge. For Patient B, delta-NIHSS5A (poor left motor arm, −1) had the highest SHAP value, followed by delta-NIHSS6B (poor right motor leg, −1), indicating a need for post-discharge motor rehabilitation. These results illustrate the utility of personalized feature importance explanations to inform tailored clinical management strategies.

To facilitate real-world applications, we developed a web-based application for risk estimation and personalized result explanations, which is accessible at http://www.discosysu.cn.

## Discussion

In this study, we employed a robust machine learning framework, incorporating explainable artificial intelligence techniques, to systematically evaluate the importance of 309 hospitalization features in predicting three critical post-stroke outcomes: recurrence, disability, and mortality at 3 months, as well as across six additional time points over five years. For the first time, we identified clinically accessible global and domain-specific delta-NIHSS_(admission-discharge)_ as key predictors of post-stroke recurrence, disability and mortality at 3 months. Based on these findings, we developed the DISCO model, which demonstrated dramatically improved discriminative ability compared to existing models, achieving AUCs more than 0.8 for 3-month stroke recurrence and disability, 0.9 for mortality.

The NIHSS is a simple, widely used clinical measure that offers valuable insights into stroke severity and recovery.^23^ It is commonly used as an endpoint in randomized controlled trials (RCTs)^23,24^ and has been associated with 90-day functional outcomes^25,26^. Previous observational and genetic studies have suggested that specific NIHSS domains correspond to distinct biological mechanisms and testing modalities,^23,27,28^ such as NIHSS 1A, which aligns with the Montreal Cognitive Assessment Scale (MOCA)^23,29^. Our findings of global and domain-specific delta-NIHSS_(admission-discharge)_ as novel and essential predictive variables for 3-month stroke composite outcomes, are consistent with the biological understanding of NIHSS. Furthermore, these findings provide new insights into the mechanisms of post-stroke recurrence and functional outcomes. Since domain-specific delta-NIHSS_(admission-discharge)_ reflects deficits in specific functional domain, its strong predictive power for 3-month stroke recurrence suggests that recurrence may represent the deterioration or progression of the initial ischemic stroke rather than the onset of a new stroke. In addition, the definitions of stroke recurrence, disability and NIHSS are based on symptomatic observation, which may explain why delta-NIHSS_(admission-discharge)_ is a strong predictor of post-stroke composite outcomes. The predictive contribution of delta-NIHSS to 3-month composite outcomes underscores the importance of recovery during hospitalization for secondary ischemic stroke prevention and improving long-term quality of life.

For future risk stratification and personalized management, the DISCO model robustly identified high-risk patients with markedly elevated relative risks in both the internal and external validation. Identifying these high-risk patients allows healthcare providers and caregivers to implement targeted interventions aimed at preventing recurrence and reducing adverse post-discharge outcomes. To facilitate clinical implementation, we developed a web application that enables clinicians to calculate individual risk estimates for stroke recurrence, disability, and mortality at three months.

Delivering AI-assisted personalized risk information and expert recommendations has been proven to be an effective non-pharmacological intervention in multicenter RCTs, such as those focused on parental check-ups for term planned delivery^30^ and primary diabetes care^31^. The DISCO model, with its predictive power and domain-specific explainability, provides a solid foundation for guiding targeted neurorehabilitation interventions in stroke care.

### Limitations

The study has several limitations. First, although the DISCO model underwent a rigorous validation process to ensure generalizability, its performance in populations beyond the Chinese cohort remains to be established. Given the model’s emphasis on clinical accessibility, the inclusion of 24 easily assessable features facilitates its validation in independent, multi-ancestry cohorts, which would help evaluate and extend its applicability. Second, the PRS used in this study were developed for ischemic stroke occurrence rather than recurrence, as no PRS models specifically designed for stroke outcomes currently exist. Therefore, we cannot rule out the potential contribution of genetic factors to stroke outcome prediction. Further investigations are warranted when genetic studies specifically targeting secondary stroke prevention become available. Third, the mechanistic understanding of global and domain-specific delta-NIHSS_(admission-discharge)_ remains limited. While our observational analysis identified several factors significantly associated with delta-NIHSS_(admission-discharge),_ future research is needed to explore their causal effects. Such investigations may provide insights into more precise treatment and intervention strategies for acute stroke management and post-stroke neurorehabilitation.

## Conclusion

The DISCO model, an AI-driven, delta-NIHSS-based predictor of composite ischemic stroke outcomes, demonstrates high accuracy, robustness, clinical accessibility, and explainability. By highlighting the significance of domain-specific recovery during hospitalization, it provides a valuable tool for personalized risk prediction, supporting targeted neurorehabilitation interventions for ischemic stroke patients.

## Supporting information

Expanded Methods

TRIPOD checklist

## Data Availability

All data produced in the present study are available upon reasonable request to the authors

## Acknowledgement

The Study was supported by Grants from National Key Research and Development Program of China (2022YFE0209600, 2022YFC2502400, 2022YFC2502402, 2022YFC2502404), the National Natural Science Foundation of China (82471304), the Young Elite Scientists Sponsorship Program by CAST (2023QNRC001) and Young Talents Supporting Program of Capital Medical University (B2417).

## References

1. Steinmetz JD, Seeher KM, Schiess N, et al. Global, regional, and national burden of disorders affecting the nervous system, 1990–2021: a systematic analysis for the Global Burden of Disease Study 2021. The Lancet Neurology. 2024;23(4):344–381. doi:10.1016/S1474-4422(24)00038-3

2. Arsava EM, Kim GM, Oliveira-Filho J, et al. Prediction of Early Recurrence After Acute Ischemic Stroke. JAMA Neurology. 2016;73(4):396–401. doi:10.1001/jamaneurol.2015.4949

3. Ntaios G, Faouzi M, Ferrari J, Lang W, Vemmos K, Michel P. An integer-based score to predict functional outcome in acute ischemic stroke. Published online 2012.

4. Saposnik G, Kapral MK, Liu Y, et al. IScore: a risk score to predict death early after hospitalization for an acute ischemic stroke. Circulation. 2011;123(7):739–749. doi:10.1161/CIRCULATIONAHA.110.983353

5. Rothwell PM, Giles MF, Flossmann E, et al. A simple score (ABCD) to identify individuals at high early risk of stroke after transient ischaemic attack. Lancet. 2005;366(9479):29–36. doi:10.1016/S0140-6736(05)66702-5

6. Beam AL, Kohane IS. Big Data and Machine Learning in Health Care. JAMA. 2018;319(13):1317–1318. doi:10.1001/jama.2017.18391

7. Heo J, Yoon JG, Park H, Kim YD, Nam HS, Heo JH. Machine Learning–Based Model for Prediction of Outcomes in Acute Stroke. Stroke. 2019;50(5):1263–1265. doi:10.1161/STROKEAHA.118.024293

8. Wijnberge M, Geerts BF, Hol L, et al. Effect of a Machine Learning-Derived Early Warning System for Intraoperative Hypotension vs Standard Care on Depth and Duration of Intraoperative Hypotension During Elective Noncardiac Surgery: The HYPE Randomized Clinical Trial. JAMA. 2020;323(11):1052–1060. doi:10.1001/jama.2020.0592

9. Kundu S. AI in medicine must be explainable. Nat Med. 2021;27(8):1328. doi:10.1038/s41591-021-01461-z

10. Van Calster B, Steyerberg EW, Collins GS. Artificial Intelligence Algorithms for Medical Prediction Should Be Nonproprietary and Readily Available. JAMA Intern Med. 2019;179(5):731. doi:10.1001/jamainternmed.2019.0597

11. Wang K, Shi Q, Sun C, et al. A machine learning model for visualization and dynamic clinical prediction of stroke recurrence in acute ischemic stroke patients: A real-world retrospective study. Front Neurosci. 2023;17:1130831. doi:10.3389/fnins.2023.1130831

12. Lv J, Zhang M, Fu Y, et al. An interpretable machine learning approach for predicting 30-day readmission after stroke. International Journal of Medical Informatics. 2023;174:105050. doi:10.1016/j.ijmedinf.2023.105050

13. Wang H, Sun Y, Zhu J, Zhuang Y, Song B. Diffusion-weighted imaging-based radiomics for predicting 1-year ischemic stroke recurrence. Front Neurol. 2022;13:1012896. doi:10.3389/fneur.2022.1012896

14. Gao Y, Li Zang, Zhai X yang, et al. An interpretable machine learning model for stroke recurrence in patients with symptomatic intracranial atherosclerotic arterial stenosis. Front Neurosci. 2024;17:1323270. doi:10.3389/fnins.2023.1323270

15. Wang Y, Jing J, Meng X, et al. The Third China National Stroke Registry (CNSR-III) for patients with acute ischaemic stroke or transient ischaemic attack: design, rationale and baseline patient characteristics. Stroke Vasc Neurol. 2019;4(3). doi:10.1136/svn-2019-000242

16. Cheng S, Xu Z, Bian S, et al. The STROMICS genome study: deep whole-genome sequencing and analysis of 10K Chinese patients with ischemic stroke reveal complex genetic and phenotypic interplay. Cell Discov. 2023;9(1):75. doi:10.1038/s41421-023-00582-8

17. Wang Yongjun, Meng Xia, Wang Anxin, et al. Ticagrelor versus Clopidogrel in CYP2C19 Loss-of-Function Carriers with Stroke or TIA. New England Journal of Medicine. 2021;385(27):2520–2530. doi:10.1056/NEJMoa2111749

18. Mishra A, Malik R, Hachiya T, et al. Stroke genetics informs drug discovery and risk prediction across ancestries. Nature. 2022;611(7934):115–123. doi:10.1038/s41586-022-05165-3

19. Chen T, Guestrin C. XGBoost: A Scalable Tree Boosting System. In: Proceedings of the 22nd ACM SIGKDD International Conference on Knowledge Discovery and Data Mining.; 2016:785–794. doi:10.1145/2939672.2939785

20. Cawley GC, Talbot NLC. On Over-fitting in Model Selection and Subsequent Selection Bias in Performance Evaluation.

21. Lundberg SM, Erion G, Chen H, et al. From local explanations to global understanding with explainable AI for trees. Nat Mach Intell. 2020;2(1):56–67. doi:10.1038/s42256-019-0138-9

22. DeLong ER, DeLong DM, Clarke-Pearson DL. Comparing the areas under two or more correlated receiver operating characteristic curves: a nonparametric approach. Biometrics. 1988;44(3):837–845.

23. Braun RG, Heitsch L, Cole JW, et al. Domain-Specific Outcomes for Stroke Clinical Trials: What the Modified Rankin Isn’t Ranking. Neurology. 2021;97(8):367–377. doi:10.1212/WNL.0000000000012231

24. Hong Y, LaBresh KA. Overview of the American Heart Association “Get with the Guidelines” programs: coronary heart disease, stroke, and heart failure. Crit Pathw Cardiol. 2006;5(4):179–186. doi:10.1097/01.hpc.0000243588.00012.79

25. Heitsch L, Ibanez L, Carrera C, et al. Early Neurological Change After Ischemic Stroke Is Associated With 90-Day Outcome. Stroke. 2021;52(1):132–141. doi:10.1161/STROKEAHA.119.028687

26. Hendrix P, Melamed I, Collins M, et al. NIHSS 24 h After Mechanical Thrombectomy Predicts 90-Day Functional Outcome. Clin Neuroradiol. 2022;32(2):401–406. doi:10.1007/s00062-021-01068-4

27. Aldridge CM, Braun R, Lohse K, et al. Genome-Wide Association Studies of 3 Distinct Recovery Phenotypes in Mild Ischemic Stroke. Neurology. 2024;102(3):e208011. doi:10.1212/WNL.0000000000208011

28. Cramer SC, Wolf SL, Adams HP, et al. Stroke Recovery and Rehabilitation Research: Issues, Opportunities, and the National Institutes of Health StrokeNet. Stroke. 2017;48(3):813–819. doi:10.1161/STROKEAHA.116.015501

29. Nasreddine ZS, Phillips NA, Bédirian V, et al. The Montreal Cognitive Assessment, MoCA: a brief screening tool for mild cognitive impairment. J Am Geriatr Soc. 2005;53(4):695–699. doi:10.1111/j.1532-5415.2005.53221.x

30. Rial-Crestelo M, Lubusky M, Parra-Cordero M, et al. Term planned delivery based on fetal growth assessment with or without the cerebroplacental ratio in low-risk pregnancies (RATIO37): an international, multicentre, open-label, randomised controlled trial. Lancet. 2024;403(10426):545–553. doi:10.1016/S0140-6736(23)02228-6

31. Li J, Guan Z, Wang J, et al. Integrated image-based deep learning and language models for primary diabetes care. Nat Med. 2024;30(10):2886–2896. doi:10.1038/s41591-024-03139-8

