## Supplemental Table S1-S14 and Supplementary Figure S1-S18 for "DISCO: A Delta-NIHSS-Based Machine Learning Model for Predicting Recurrence, Disability, and Mortality Following Acute Ischemic Stroke"

### Supplemental Figures and Tables

**Figure S1**. Workflow of Delta-NIHSS(admission-discharge) Influence Factors Exploration in Whole Analysis Sample and Two Scenarios

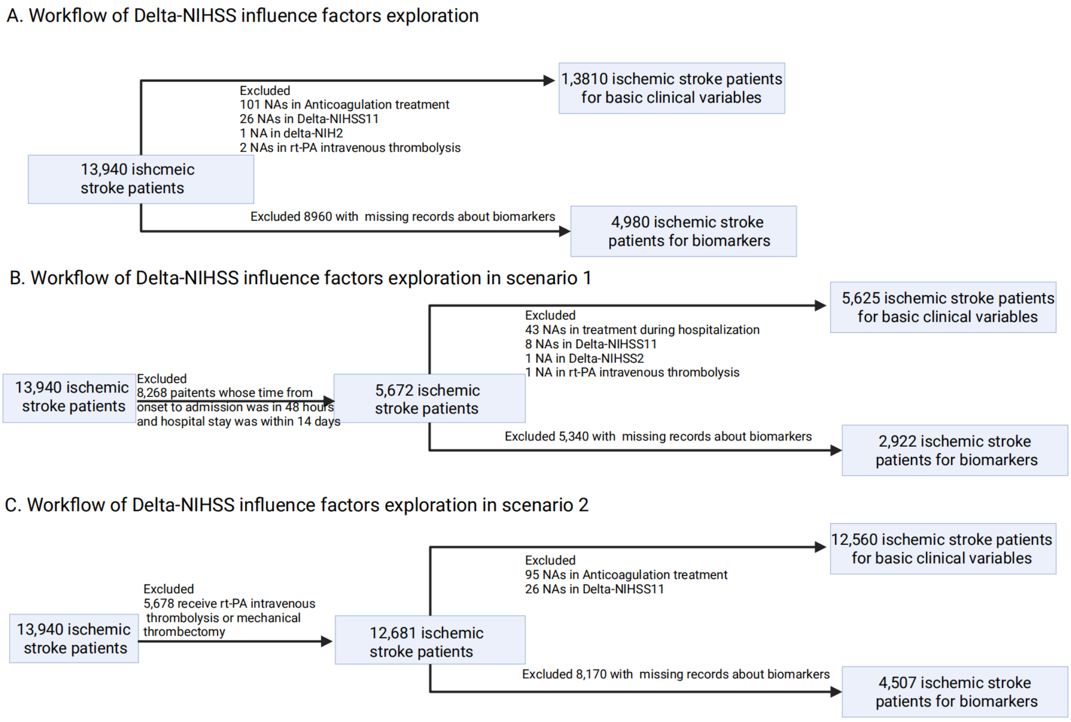

Part A of the figure outlines the patient recruitment workflow for exploring the factors influencing delta-NIHSS(admission-discharge). In this process, 13,810 ischemic stroke patients were included for a multivariable analysis of basic clinical variables, while 4,980 ischemic stroke patients were utilized for biomarker analysis in the entire CNSR-III cohort. Part B presents the patients in Scenario 1, which includes those with a hospital stay within 14 days or a time from stroke onset to admission of less than 48 hours. In this group, 5,625 patients were analyzed for clinical variables, and 2,922 for biomarkers. Part C focuses on Scenario 2, consisting of patients who did not receive rt-PA intravenous thrombolysis. This group included 12,560 ischemic stroke patients for clinical variables and 4,507 for biomarkers.

**Figure S2. The cumulative incidence of stroke recurrence in all patients of CNSR-III and analysis group**

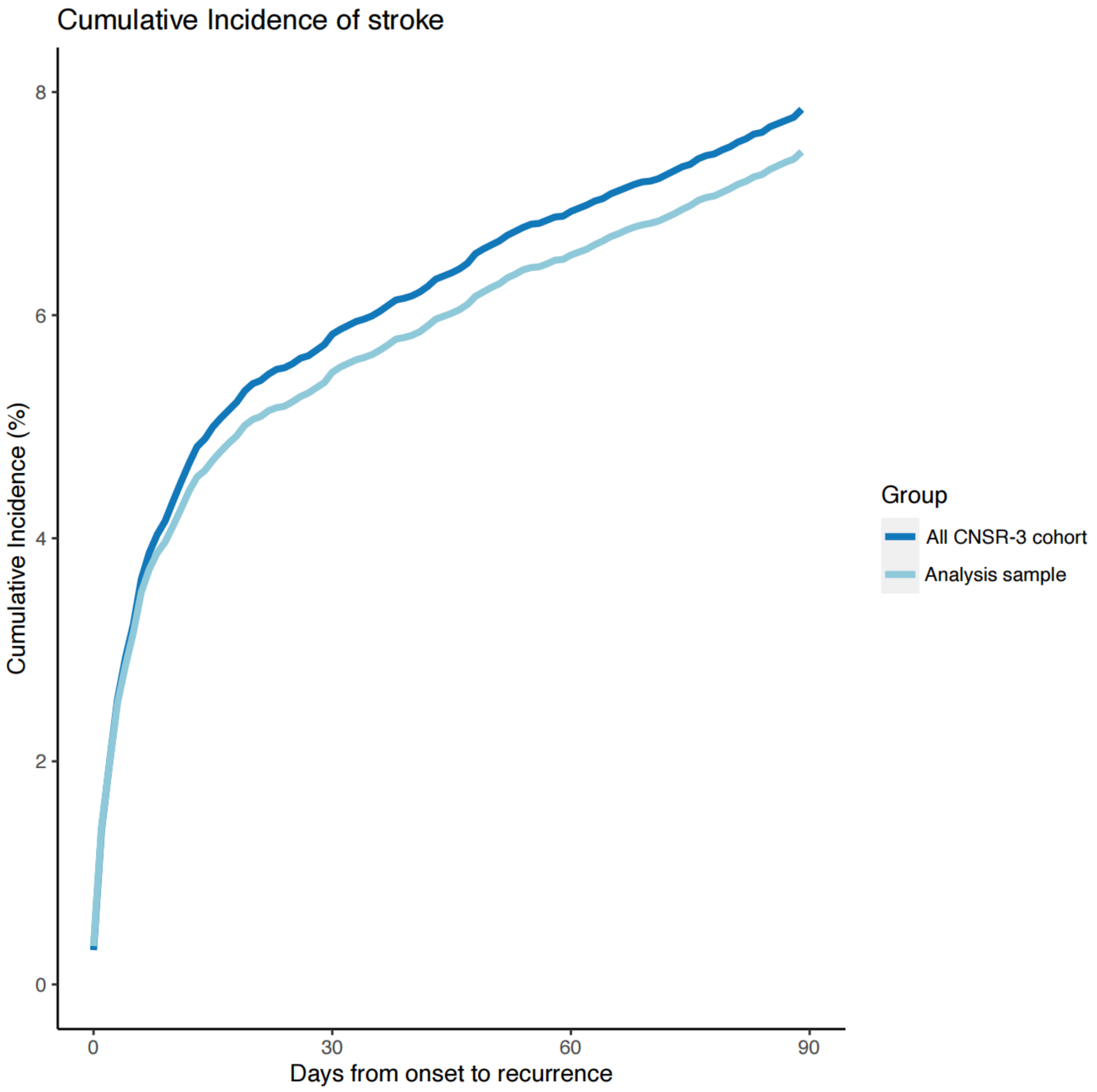

This figure presents the cumulative incidence of stroke recurrence in 3 months in all CNSR-III cohort and analysis group. Most stroke recurrence happens in days and weeks and it's similar in all CNSR-III cohort and analysis group.

**Figure S3. The feature importance rankings of all features in the model predicting stroke recurrence in 3 months (Top 20)**

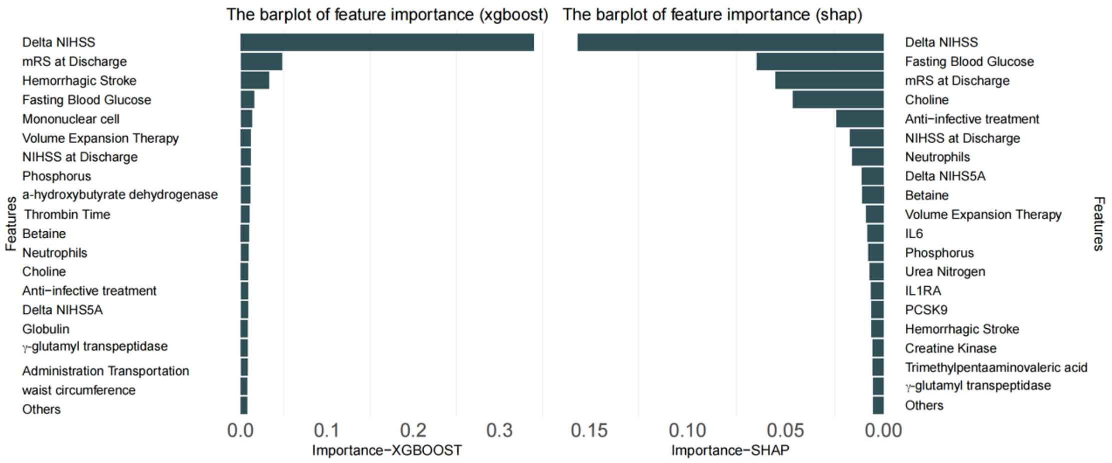

The left barplot is the rankings of feature importance given by XGBoost and the right barplot is the feature importance given by SHAP. The top 3 features given by XGBoost and Shap are same, which are delta-NIHSS$_{(admission-discharge)}$ and modified Rankin score.

Mononuclear Cell means Absolute value of mononuclear cell population.

Neutrophils: Absolute value of neutrophils

Delta NIHS5A: Delta NIHS5A is an ordinary variable to assess the left arm movement change between administration and discharge.

**Figure S4. The distribution of ischemic stroke susceptibility's PRS in CNSR-III cohort**

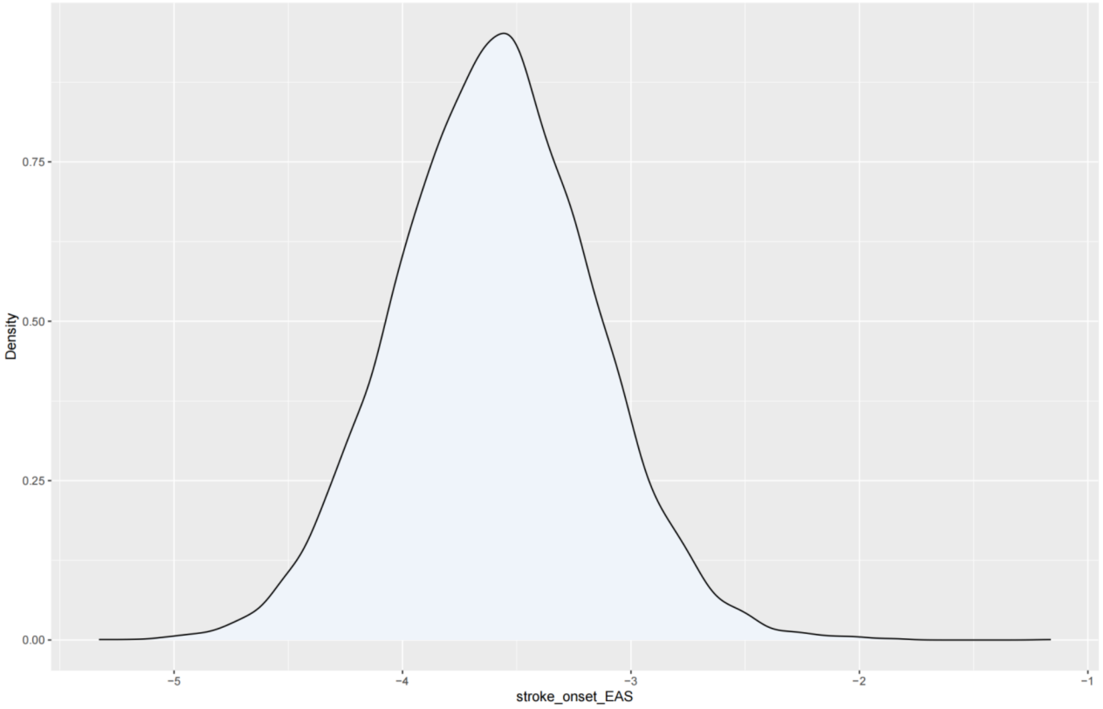

The figure above presented the Polygenic Risk Scores (PRS) based on the 9,402 ischemic stroke patients in CNSR-III cohort and the utilized PRS model is proposed by GIGASTROKE consortium for Eastern Asian.

**Figure S5. Receiver Operating Characteristic (ROC) Curves of 7 models in internal validation**

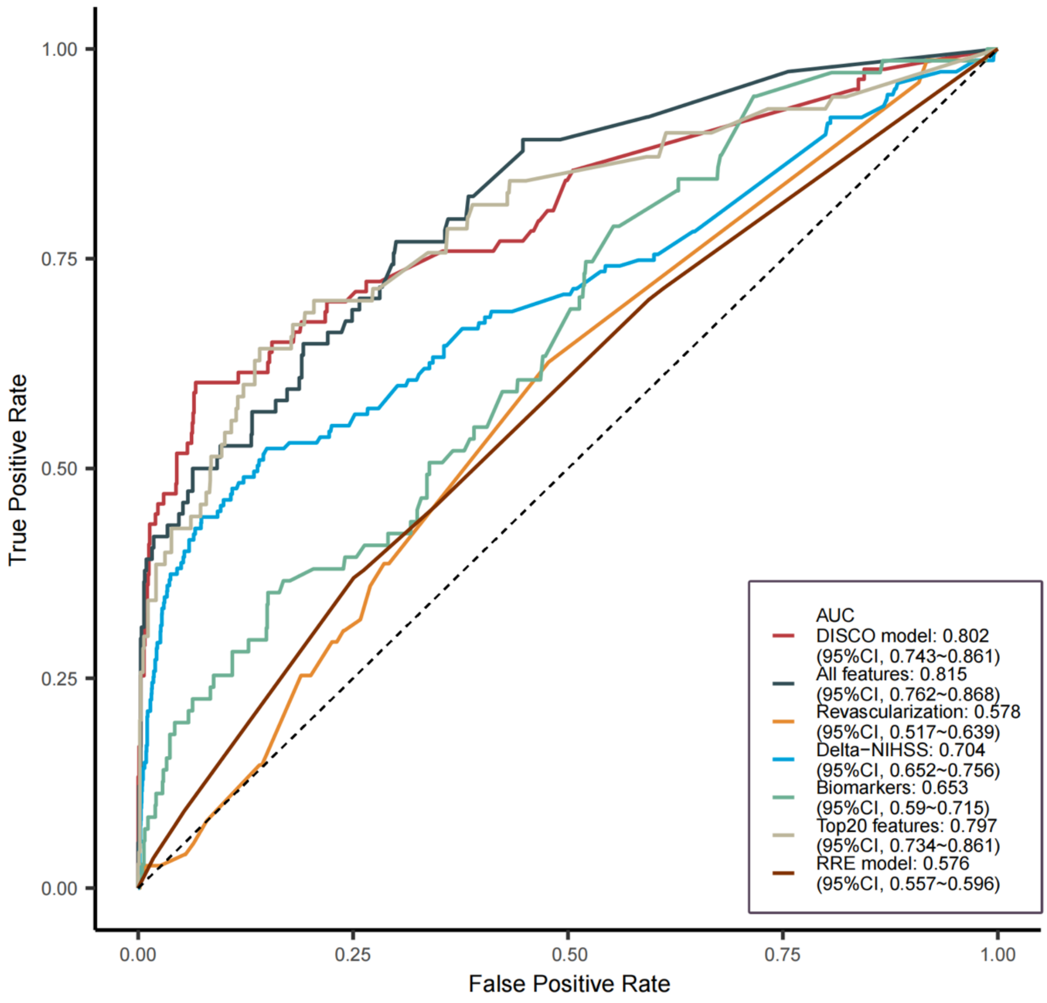

AUC: Area Under the Curve

Top20 model is constructed by the ranking Xgboost ranging from 1 to 20, age and gender to predict stroke recurrence in 3 months in CNSR-III cohort. All features model is the model contributed by 309 variables during hospital stay to predict the stroke recurrence in 3 months in CNSR-III cohort.

Revascularization model is the model constructed by age, gender, rt-PA intravenous thrombectomy and mechanical retrieval of the thrombus to predict the stroke recurrence in 3 months in CNSR-III cohort.

delta-NIHSS$_{\text{(admission-discharge)}}$. model is the model constructed by age, gender, global delta-NIHSS$_{\text{(admission-discharge)}}$. and domain-specific delta-NIHSS$_{\text{(admission-discharge)}}$. to predict the stroke recurrence in 3 months in CNSR-III cohort.

Biomarkers is the model constructed by the biomarkers described in **Table S2** to predict stroke recurrence in 3 months in CNSR-III cohort.

Among the seven models described above, the performance of model only slightly weaker than the model constructed with all features.

**Figure S6. The Calibration Curve of DISCO Model in validation cohort 1**

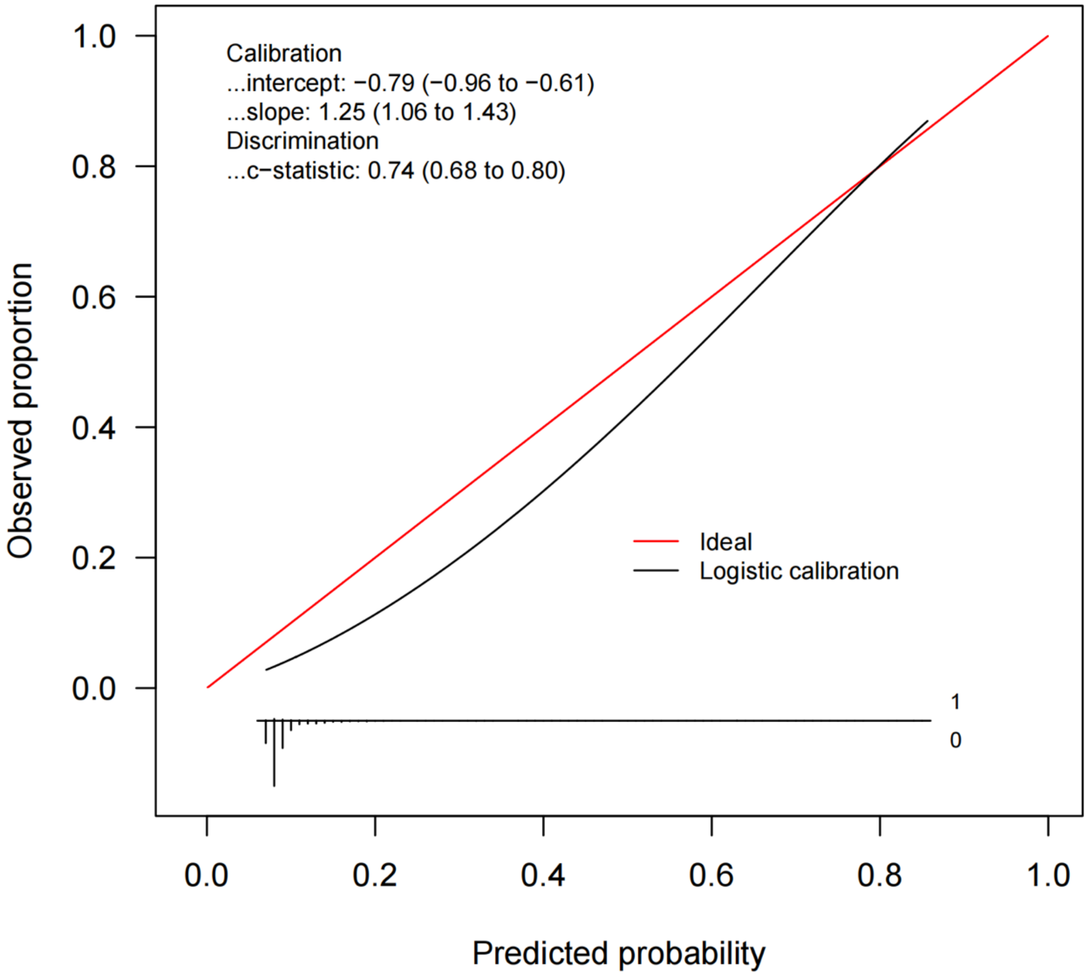

Calibration curves constructed by the bootstrap approach in the validation cohort1 set. The apparent curve and bias-corrected curve mildly deviated from the reference line, but good conformity between the observation and prediction was observed.

**Figure S7. The Calibration Curve of Main Model in validation cohort 2**

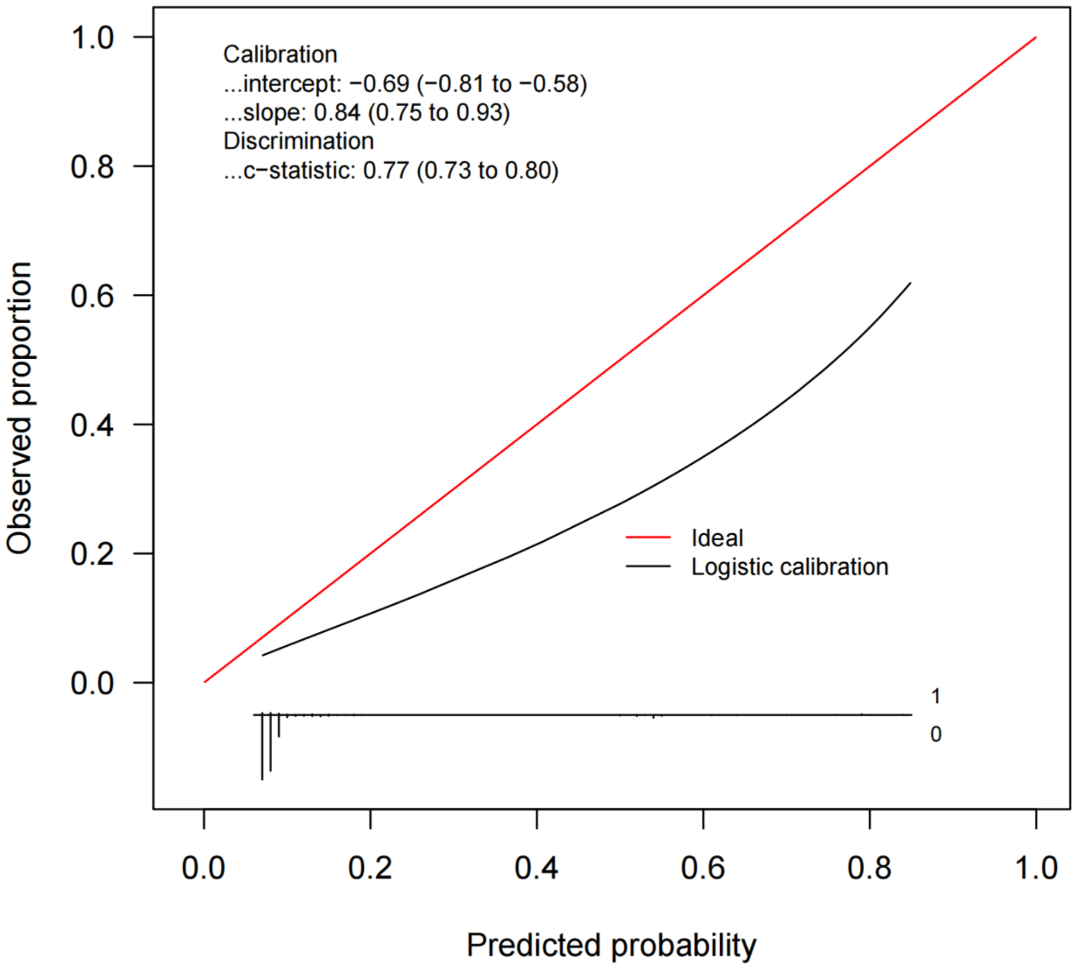

Calibration curves constructed by the bootstrap approach in the validation cohort 2. The apparent curve and bias-corrected curve mildly deviated from the reference line, but good conformity between the observation and prediction was observed.

**Figure S8. The Decision Curve of DISCO model in validation cohort 1**

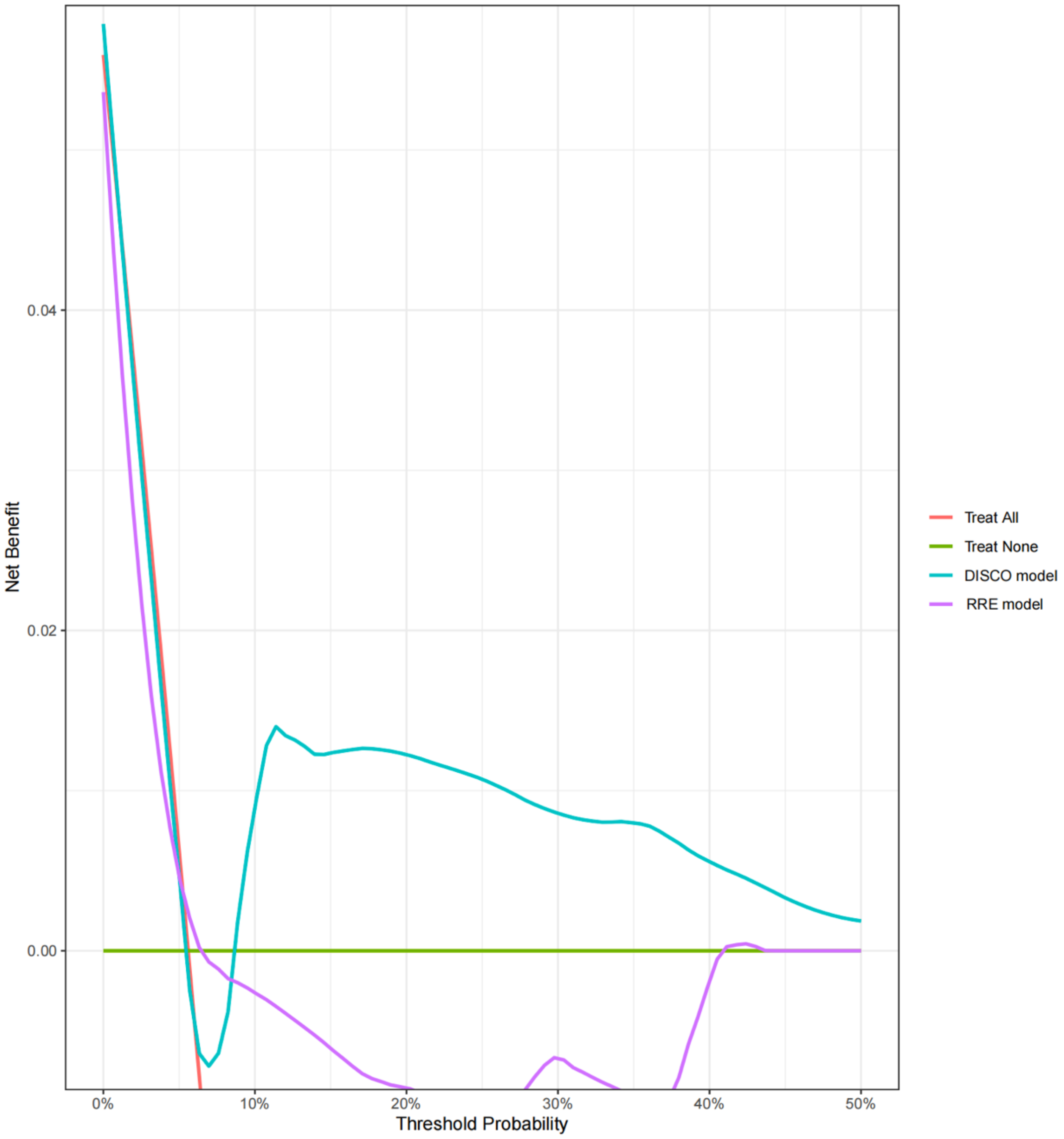

DISCO model is the result of decision curve analysis of DISCO model in the dataset of validation cohort 1.

RRE model is the result of decision curve analysis of RRE model in the dataset of validation cohort 1.

**Figure S9. The Decision Curve of DISCO model in validation cohort 2**

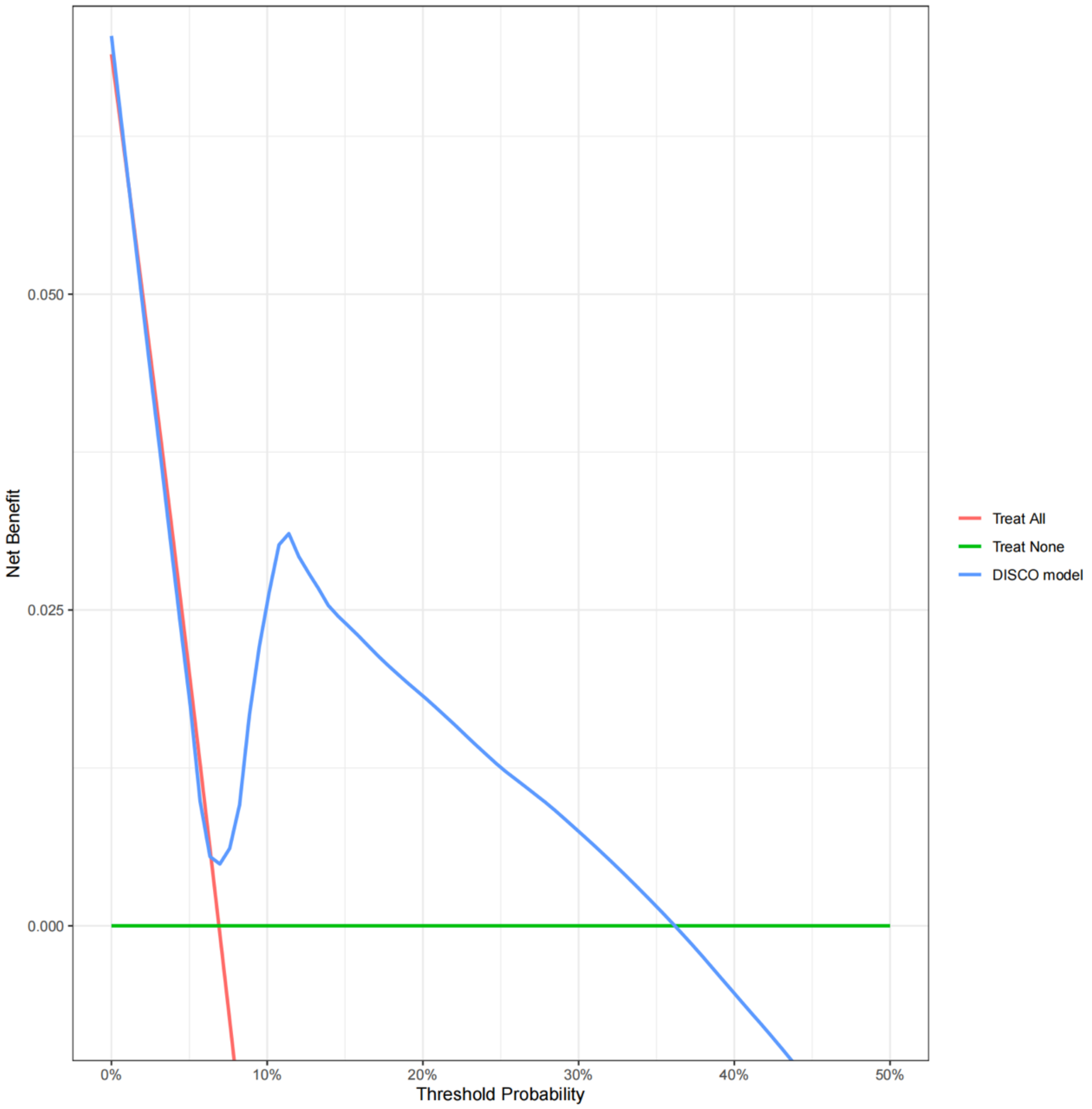

DISCO model is the result of decision curve analysis of DISCO model in the validation cohort 2.

**Figure S10. The ROCs of five different methods in the construction of DISCO model (validation cohort1)**

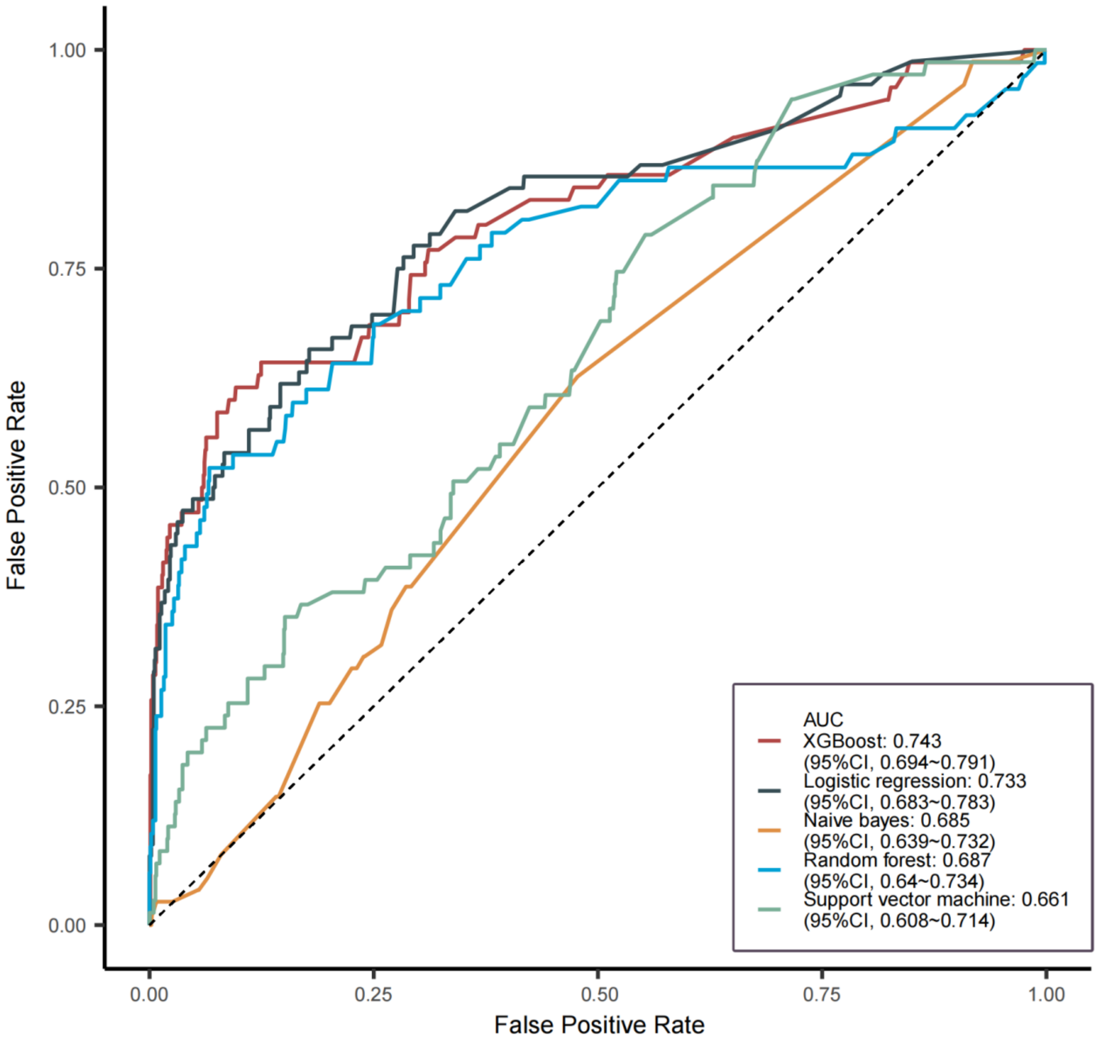

We compared the model constructed by the selected features of DISCO model by different machine learning algorithms and logistic regression in the validation cohort 1. Then, we found that the model constructed by XGBoost had highest AUC.

**Figure S11. ROCs of DISCO model performance in scenario 1**

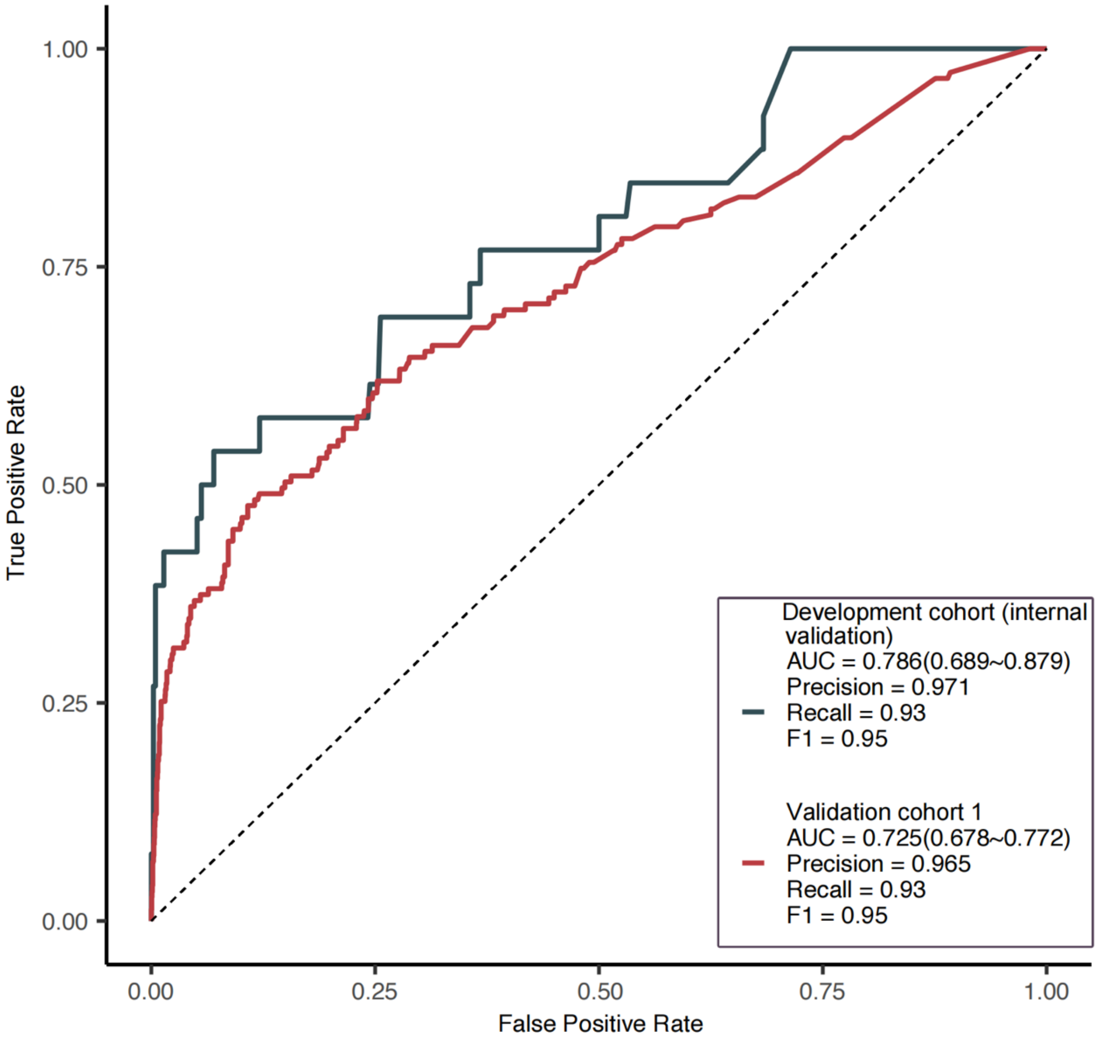

The figure presents the AUC in internal validation and external validation for DISCO model to predict stroke recurrence in 3 months when the sample was limited with ischemic stroke patients whose hospital stay within 14 days and the time from stroke onset to admission equal or fewer than 2 days. The AUC is similar to the DISCO model in whole analysis group.

**Figure S12. ROCs of DISCO DISCO performance in scenario 2**

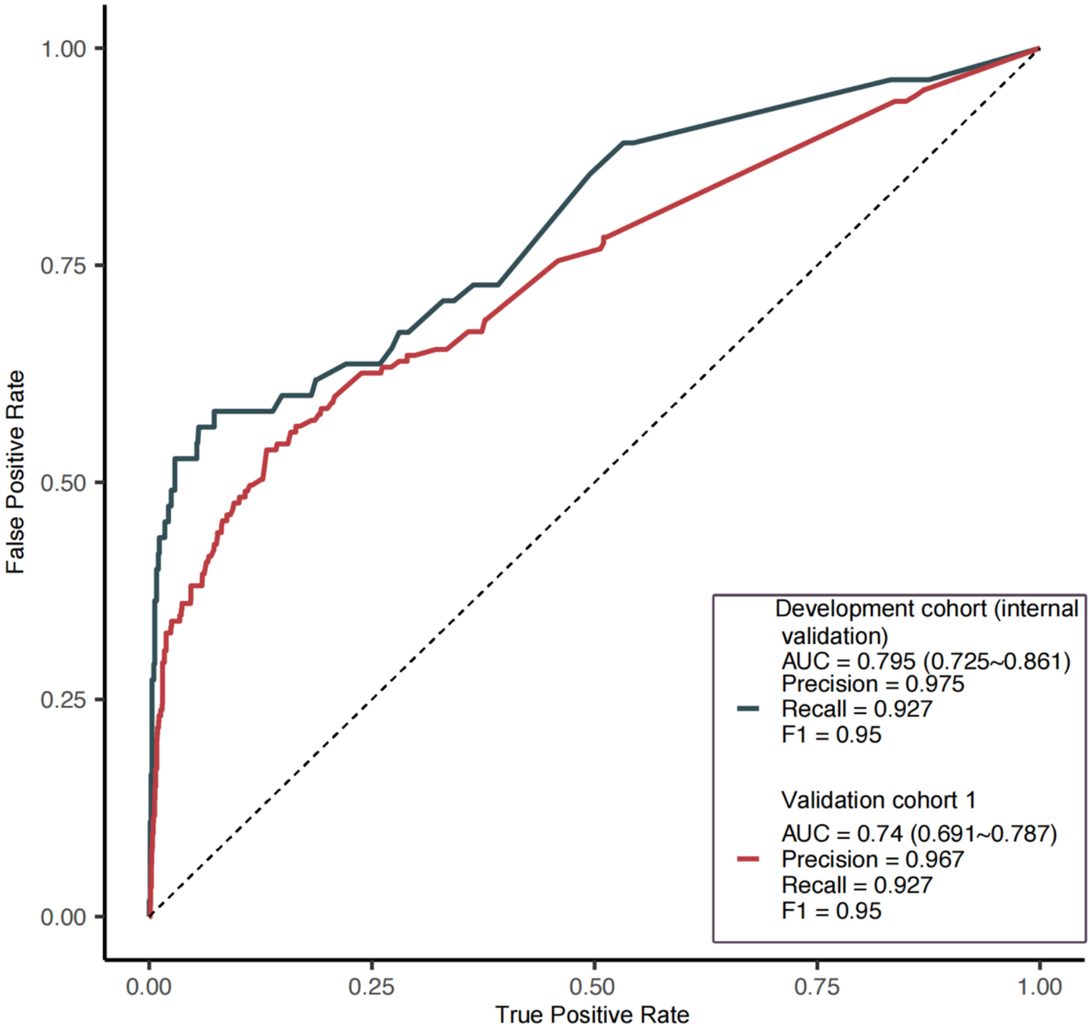

The figure presents the AUC in internal validation and external validation for DISCO model to predict stroke recurrence in 3 months when the sample was limited with stroke patients whose didn't undergo rt-PA intravenous thrombolysis or mechanical thrombectomy.

**Figure S13. ROC curves for the DISCO model adding polygenic risk scores**

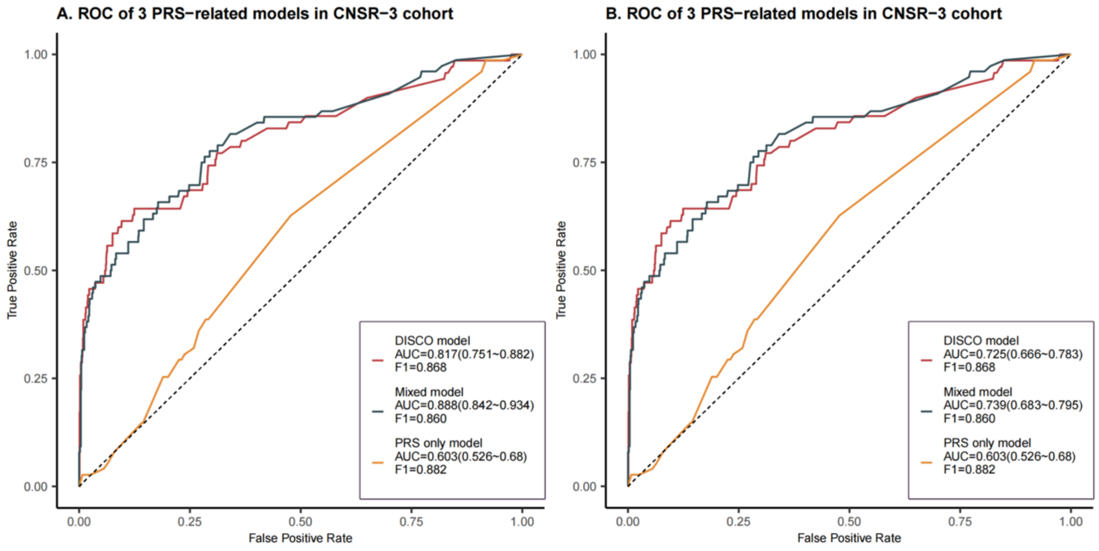

The figure illustrates the AUCs for the DISCO model, the DISCO model with PRS (mixed model), and the PRS model constructed by the feature of age, gender and PRS in predicting 3-month stroke recurrence. The evaluations were performed using data from 9,402 ischemic stroke patients in the CNSR-III cohort, which includes WGS data, with results presented for development cohort (A, [internal validation]) and validation cohort 1 (B).

**Figure S14. The Distribution of delta NIHSS in all analysis group**

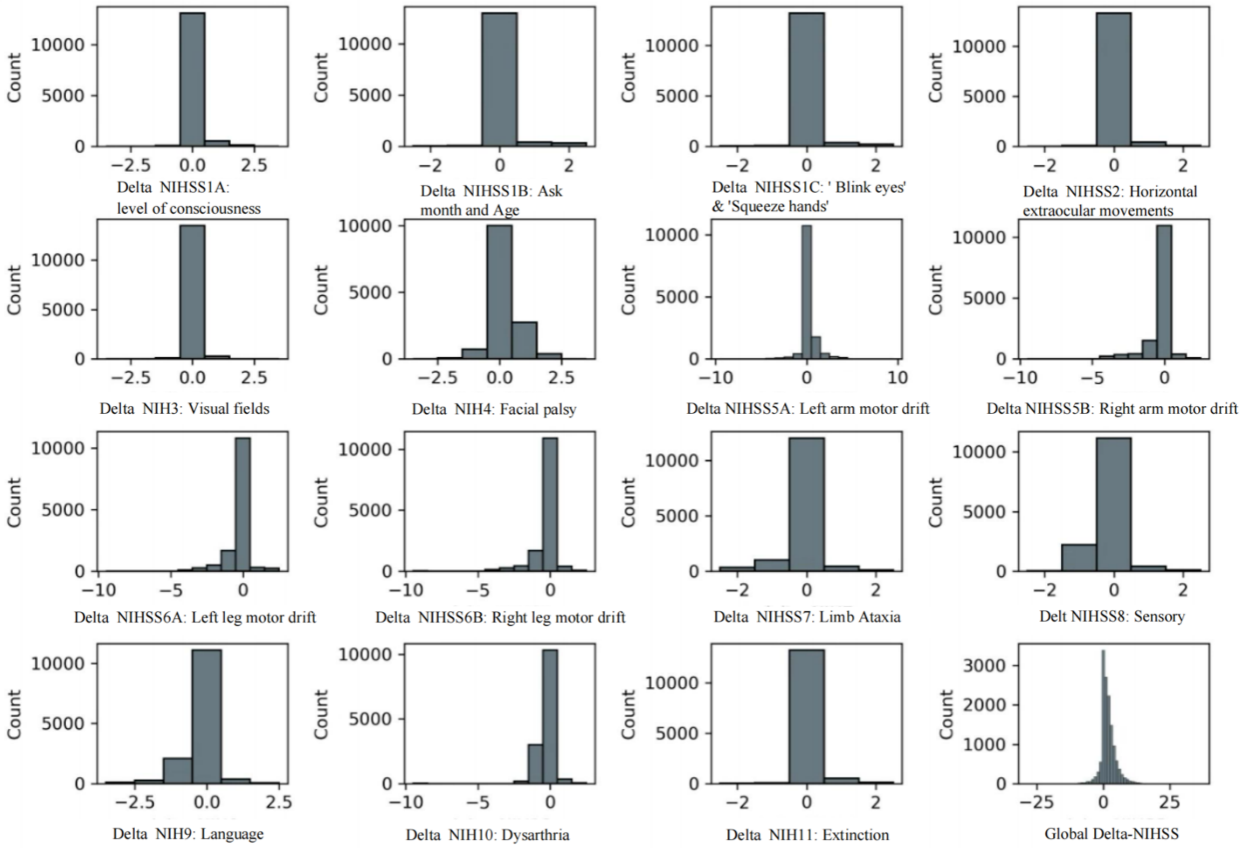

The distribution of global nad domain-specific delta-NIHSS$_{\text{(admission-discharge)}}$ is zero-inflated.

**Figure S15. Multi-variable regression Results of Basic Clinical Variables and Delta-NIHSS(admission-discharge) in Scenario1**

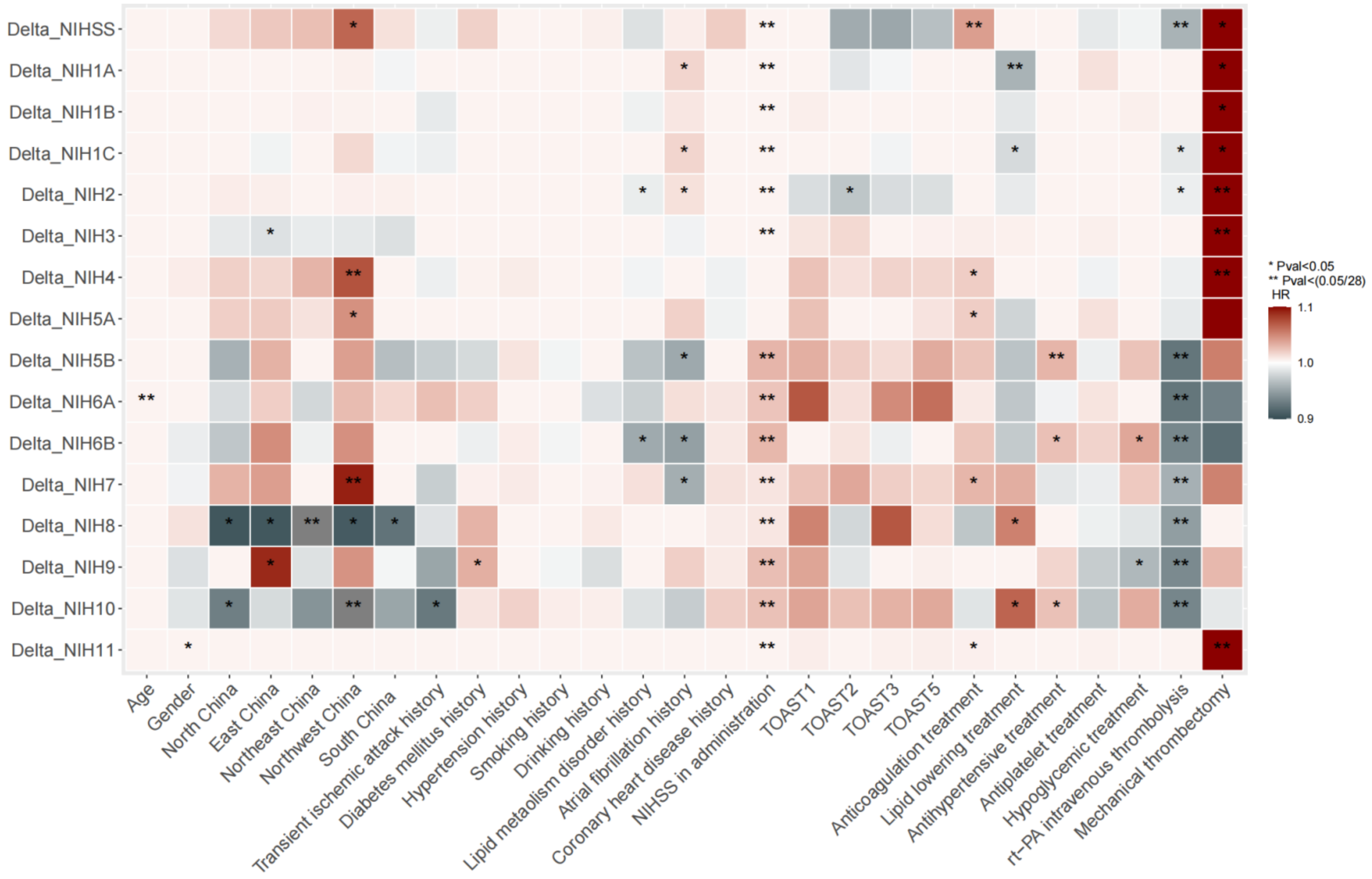

This figure presents the results of multi-variable regression between clinical variables and global and domain-specific delta-NIHSS(admission-discharge) in scenario 1.

**Figure S16. Multi-variable Regression Results of Basic Clinical Variables and Delta-NIHSS(admission-discharge) in Scenario2**

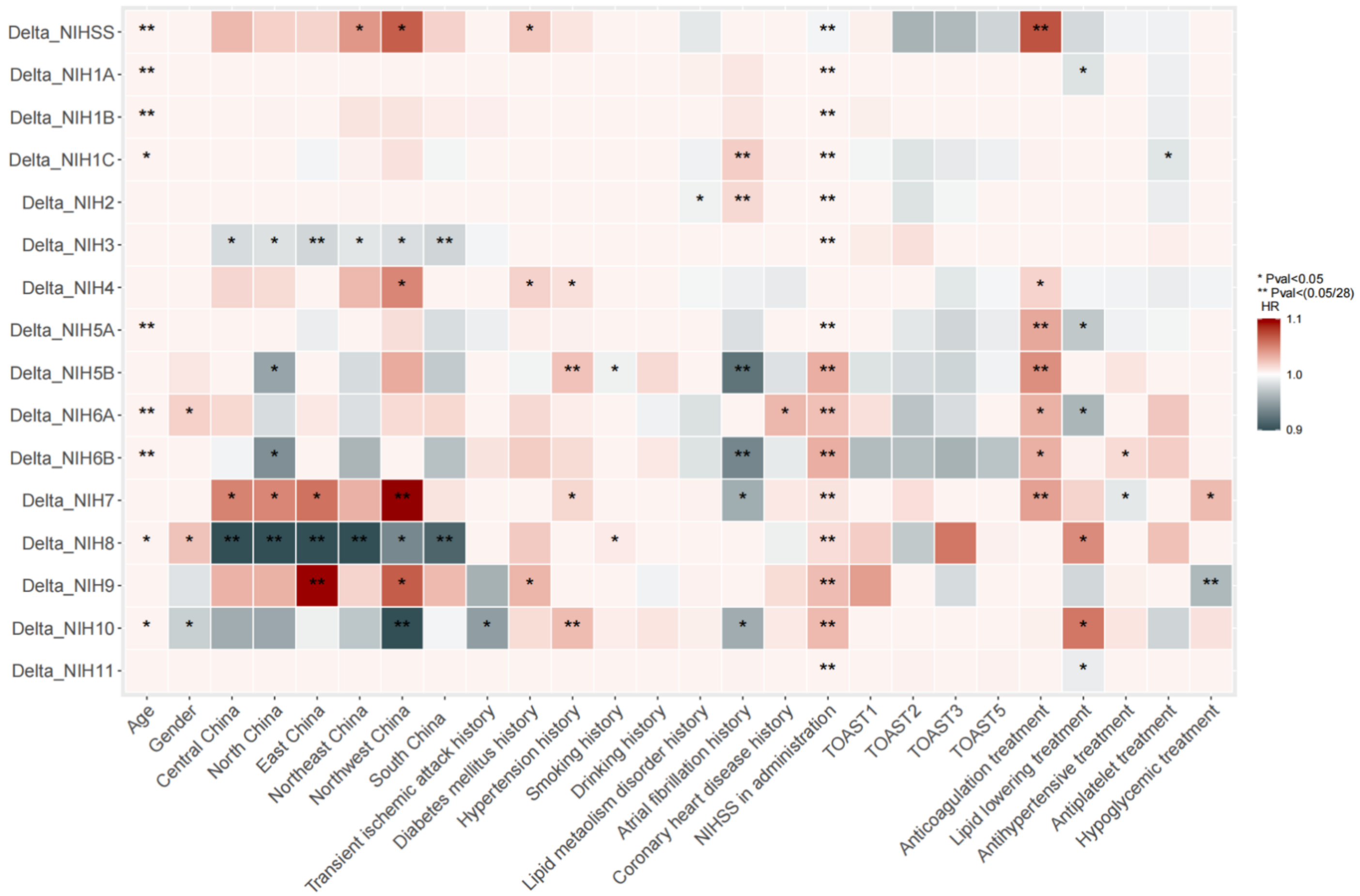

This figure presents the results of multi-variable regression between clinical variables and global and domain-specific delta-NIHSS(admission-discharge) in scenario2.

**Figure S17. Multi-variables regression Results of biomarkers and Delta-NIHSS(admission-discharge)**

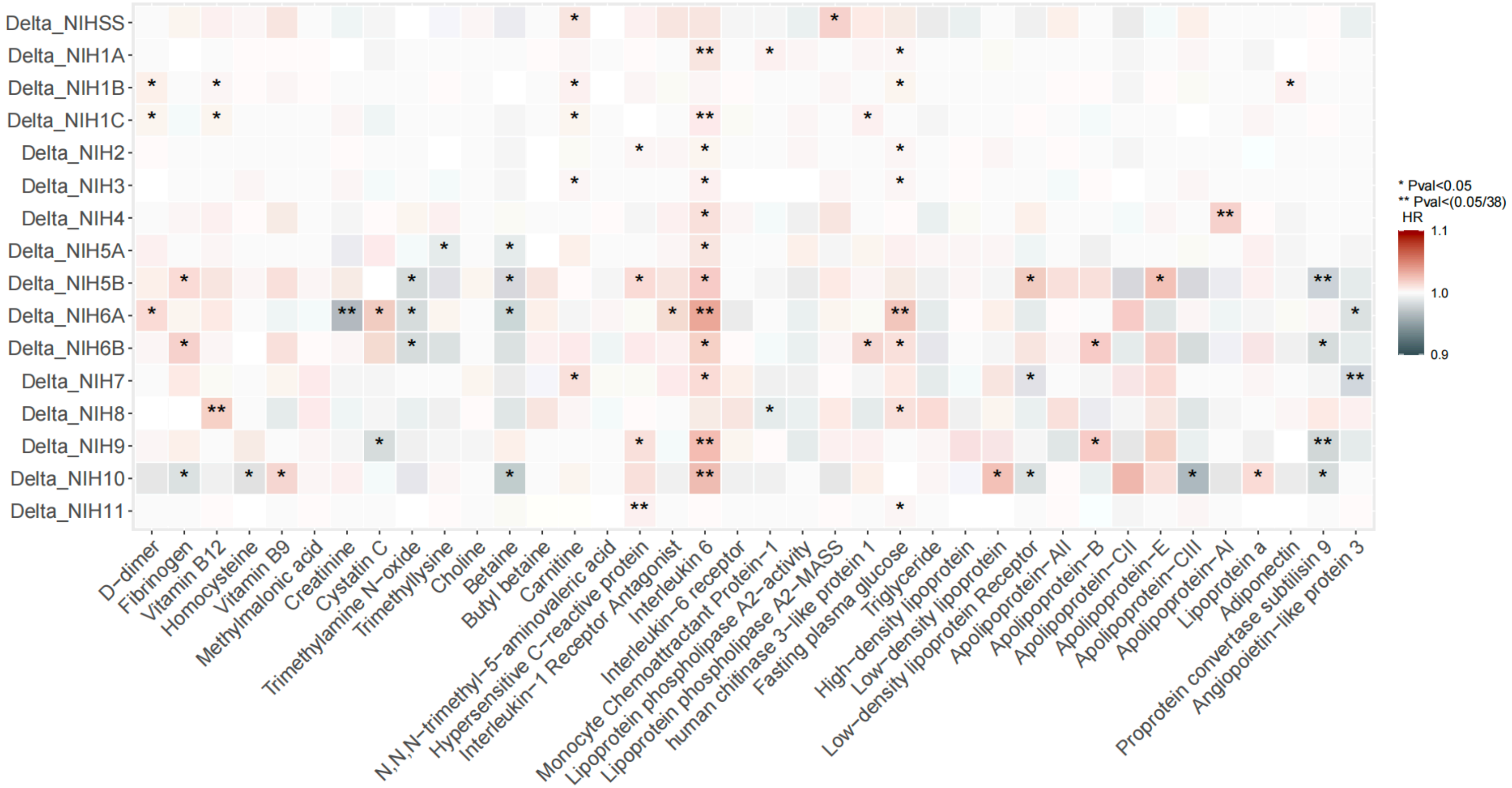

This figure presents the results of multi-variable regression between biomarkers and global and domain-specific delta-NIHSS(admission-discharge).

**Figure S18. Multi-variable regression Results of Biomarkers and Delta-NIHSS$_{(admission-discharge)}$ in Scenario 1**

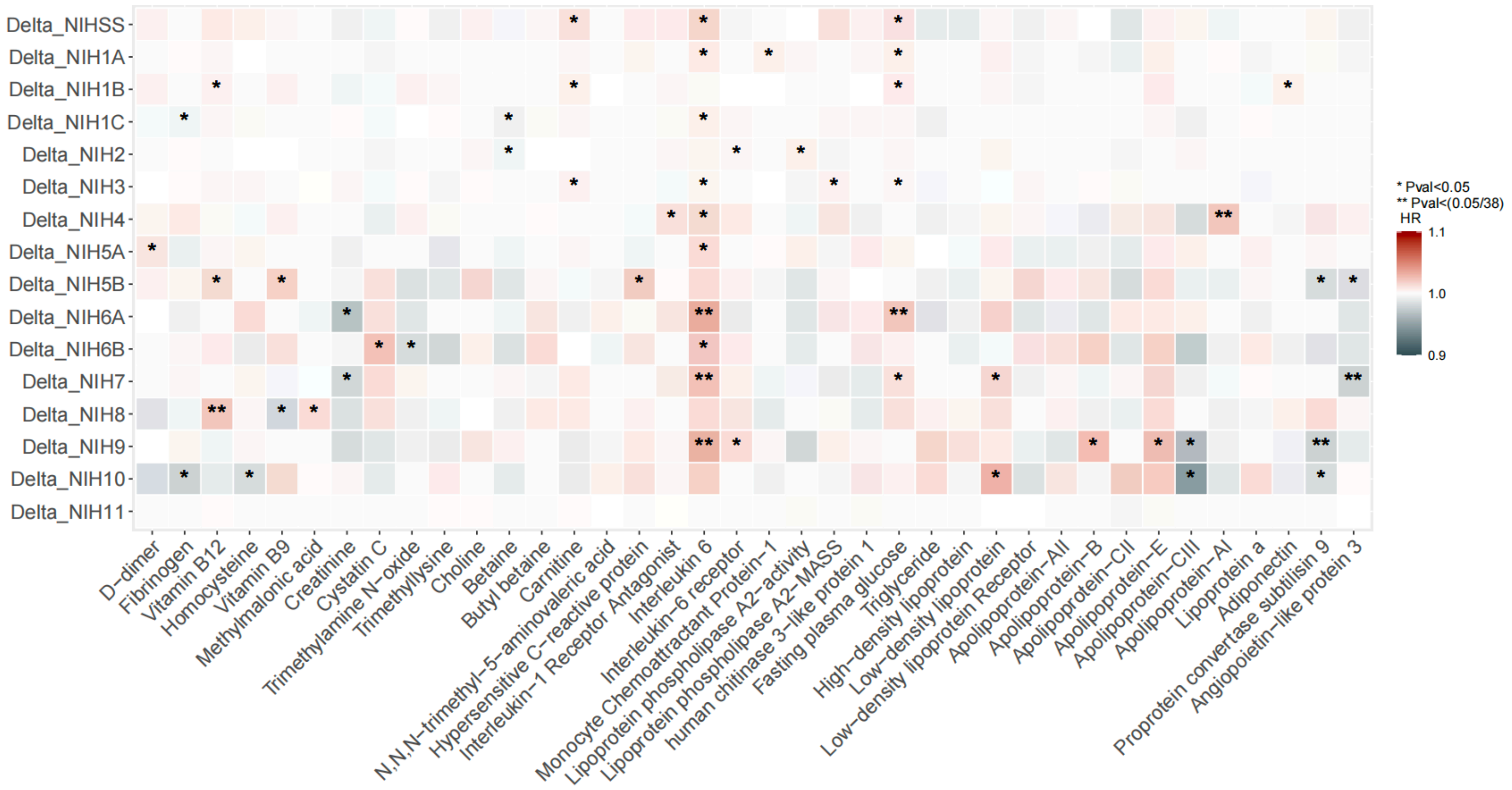

This figure presents the results of multi-variable regression between biomarkers and global and domain-specific delta-NIHSS$_{(admission-discharge)}$ in scenario1.

**Figure S19. Multi-variable regression Results of Biomarkers and Delta-NIHSS$_{(admission-discharge)}$ in Scenario 2**

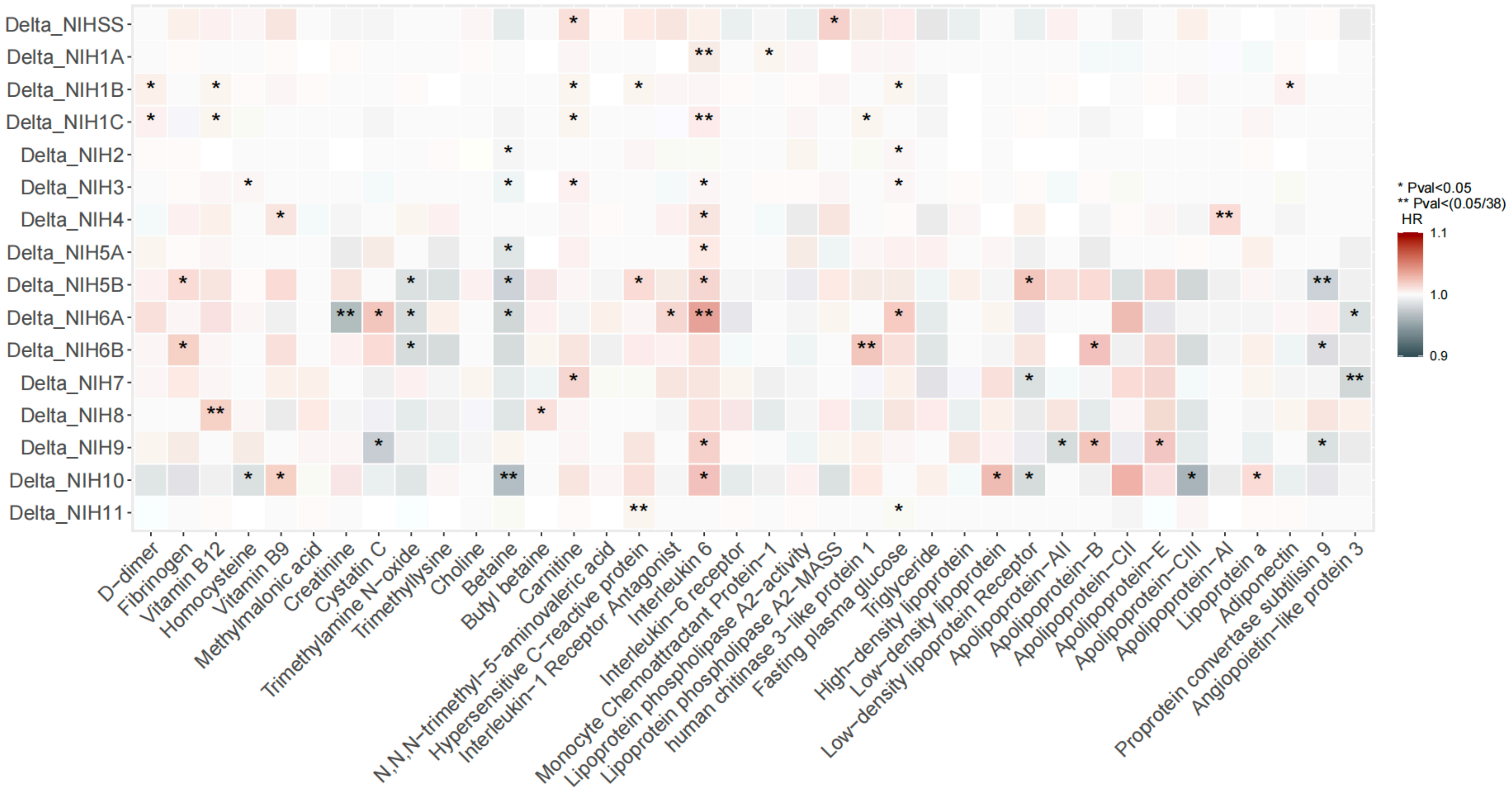

This figure presents the results of multi-variable regression between biomarkers and global and domain-specific delta-NIHSS$_{(admission-discharge)}$ in scenario2

**Figure S20. The risk stratification of DISCO model for 3-month recurrence, disability and mortality in the validation cohort 1**

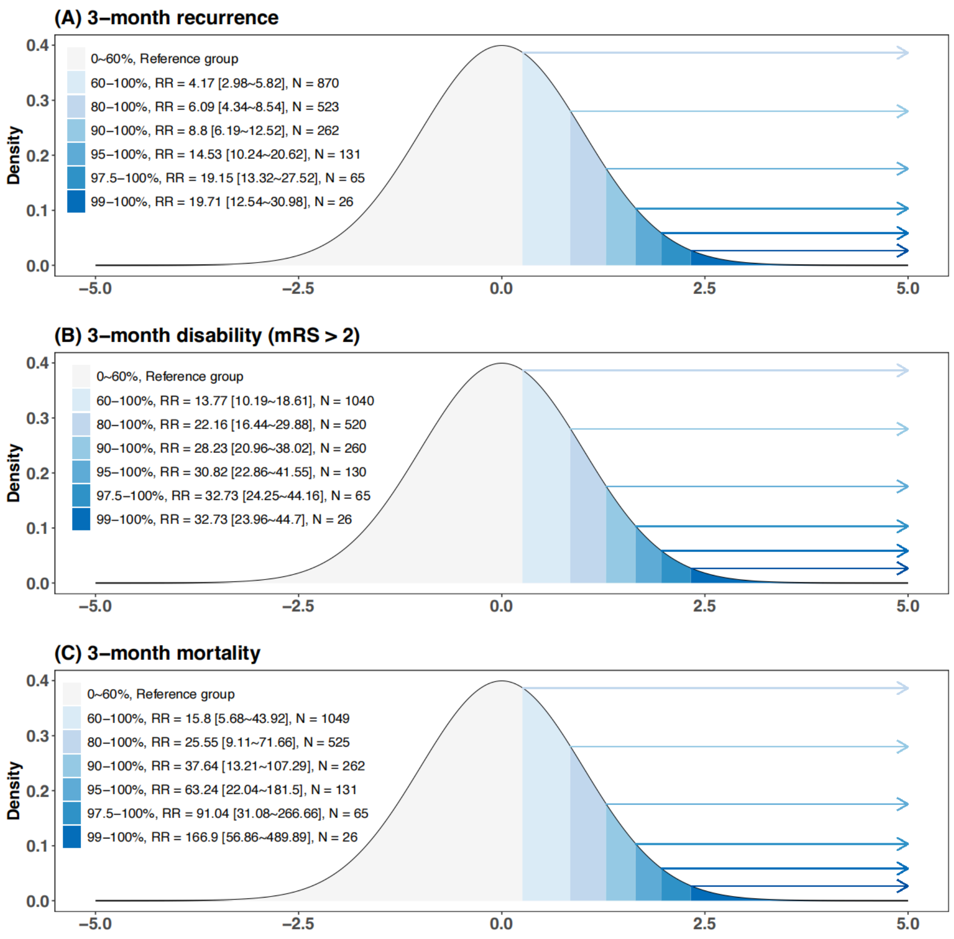

**Figure S21. The risk stratification of main model for 3-month recurrence, disability and mortality in validation cohort 2**

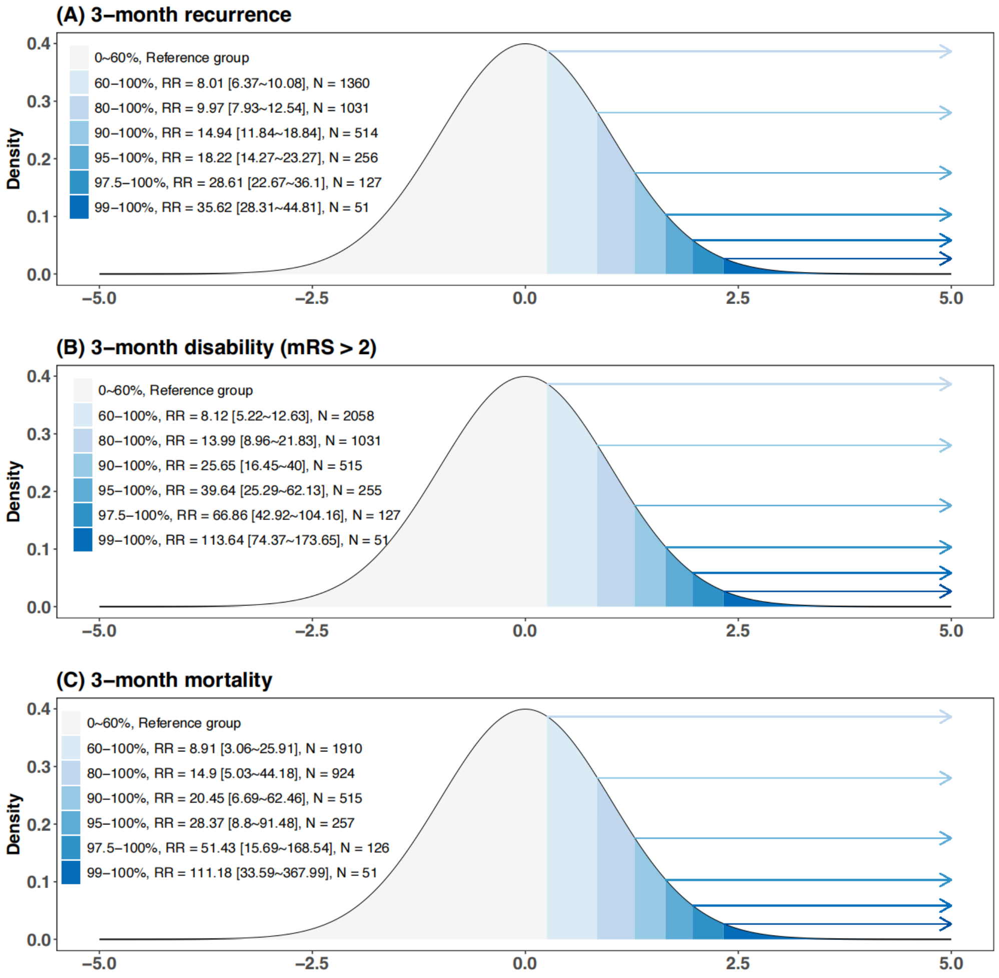

**Figure S22. The individual explanation of randomly selected patient A and B**

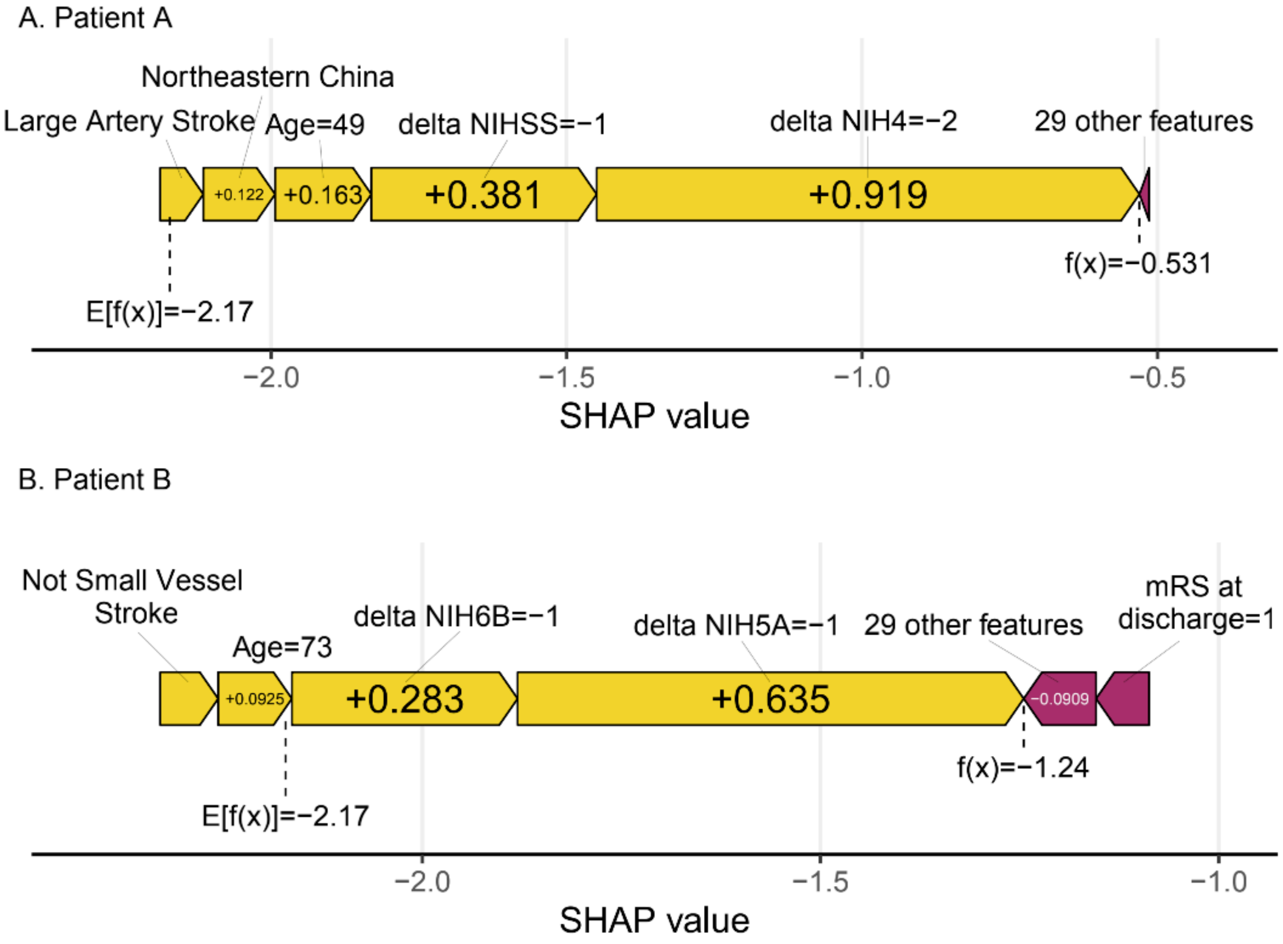

**Table S1. Literature review of stroke outcome prediction by machine learning method**

| Paper author | Year | Dataset Size (N=patients) | Geographic area | Outcomes | Method of model | Main recurrence predictors | AUROC | external validation |
|---|---|---|---|---|---|---|---|---|
| Vida Abedi | 2021 | 2091 | USA | Stroke recurrence prediction of 1, 2, 3, 4, and 5 years | LR, XGC, GBM, RF, SVM, DT | age, body mass index, and laboratory-based features (such as high-density lipoprotein, hemoglobin A1c, and creatinine) | 0.79(1-year) 0.69(5-year) | No |
| Kai Wang | 2023 | 645 | China | Stroke recurrence prediction within 1 year | RF, NBC, DT, XGB, GBM, LR | right hemisphere, homocysteine (HCY), C-reactive protein (CRP), and stroke severity (SS) | 0.946 | Yes |
| Ji Lv | 2023 | 6558 | China | Stroke recurrence prediction within 30 days | XGB | severe carotid artery stenosis, fresh cerebral infarction, weakness, urine glucose, glycosylated hemoglobin and sex | 0.8 | No |
| Negar Darabi | 2021 | 3184 | USA | Stroke recurrence prediction within 30 days | RF, GBM, XGB, SVM, and LR | the National Institutes of Health Stroke Scale score above 24, insert indwelling urinary catheter, hypercoagulable state, and percutaneous gastrostomy | 0.74 | No |

| | | | | | | | | |
|---|---|---|---|---|---|---|---|---|
| Christina M. Lineback | 2021 | 2855 | USA | Stroke recurrence prediction within 30 days | LR, NB, SVM, RF, GBM, XGB | “stenosis,” “encephalomalacia,” “craniectomy,” “encephalomalacia,” “mild calcified atherosclerotic,” “hypoattenuation white matter,” and “chiari ii malformation.” | 0.62 | No |
| Haifeng Xu | 2021 | 3159 | China | Long-term stroke recurrence prediction | AdaB, TrAdaB | BMI, age, minor stroke, hypertension, drinking, rule 3, sex, area, HbA1c, smoking | 0.758 | No |
| Jianmo Liu | 2023 | 612 | China | Stroke recurrence prediction within 1 year | LR, SVC, LightGBM, RF | 20 radiomics features | 0.789 | No |
| Hao Wang | 2022 | 1003 | China | Stroke recurrence prediction within 1 year | RNN | 513 radiomics features and clinical features | 0.847 | No |
| Yu Gao | 2023 | 180 | China | Stroke recurrence prediction in SICAS | LR, SVM, GNB, CNB, kNN | “history of hypertension,” “homocysteine level,” “NWI value,” “stenosis rate,” “intracranial hemorrhage,” “positive remodeling,” and “enhancement grade” | 0.912 | No |
| Kenneth J Ottenbacher | 2001 | 9584 | USA | Stroke recurrence prediction within 3–6 months | LR and neural networks | sphincter control, self-care ability, age, marital status, ethnicity and length of stay | 0.74 | No |

| | | | | | | | | |
|---|---|---|---|---|---|---|---|---|
| Giovanna Mercurio | 2023 | 3699 | Rome | Stroke recurrence prediction within 30 days | LR, decision tree, RF,XGB | hemoglobin level, atrial fibrillation, brain hemorrhage, discharge home, chronic obstructive pulmonary disease, one and more than one hospitalization | 0.61 | No |
| Yuan Xu | 2019 | 6070 | China | Stroke recurrence prediction within 90 days | LR, XGB | hypertension, RDW (CV), direct bilirubin, LOS, pneumonia, ALP, C-reactive protein, Aspartate aminotransferase, diabetes mellitus, and glycosylated hemoglobin. | 0.782 | No |

**Table S2. Data description of variables in CNSR-III**

| Number of Feature | Feature name | Feature description | Feature statistics | Number of NAs | feature type |
|---|---|---|---|---|---|
| 1 | Female | 4378 | counts | 0 | Basic information |
| 2 | Ethnic (Han) | 13522 | counts | 0 | Basic information |
| 3 | AGE | 63(54-70) | media(IQR) | 0 | Basic information |
| 4 | Height | 166.92;7.46 | mean;SD | 0 | Basic information |
| 5 | Weight | 68.87;10.99 | mean;SD | 23 | Basic information |
| 6 | BMI | 24.69;3.32 | mean;SD | 0 | Basic information |
| 7 | Waist circumference (cm) | 81.99;11.67 | mean;SD | 155 | Basic information |
| 8 | Marital states | | | 26 | Basic information |
| | unmarried | 88 | counts | | |
| | married | 13073 | counts | | |
| | divorced | 117 | counts | | |
| | widowed | 620 | counts | | |
| | remarried | 16 | counts | | |
| 9 | Living condition | | | 0 | Basic information |
| | living alone | 709 | | | |
| | living with others | 13231 | | | |
| 10 | Education level | | | 2034 | Basic information |
| | college or above | 1123 | counts | | |
| | high school | 2719 | counts | | |
| | junior school | 4058 | counts | | |
| | primary school | 2988 | counts | | |
| | illiteracy | 1018 | counts | | |
| 11 | Family monthly income per capita | | | 3520 | Basic information |
| | <700 Yuan | 786 | counts | | |
| | 700~1500 Yuan | 1937 | counts | | |

| | | | | | |
|---|---|---|---|---|---|
| | 1501~2300 Yuan | 2981 | counts | | |
| | >2300 Yuan | 4716 | counts | | |
| 12 | Occupation | | | 971 | Basic information |
| | leaders | 369 | counts | | |
| | Technician | 764 | counts | | |
| | clerical staff | 378 | counts | | |
| | Businessman | 433 | counts | | |
| | Production staff | 3988 | counts | | |
| | Transport staff | 536 | counts | | |
| | soldier | 22 | counts | | |
| | other | 606 | counts | | |
| | unemployment | 1572 | counts | | |
| | Retirement | 4291 | counts | | |
| | Students | 10 | counts | | |
| 13 | First department | | | 148 | Basic information |
| | Neurology Emergency | 4130 | counts | | |
| | Medical emergency | 4153 | counts | | |
| | Neurosurgery Emergency | 98 | counts | | |
| | Outpatient clinic | 5411 | counts | | |
| 14 | Antiplatelet agents | 2348 | counts | 0 | Disease history |
| 15 | Anticoagulant agents | 136 | counts | 0 | Disease history |
| 16 | Lipid-lowering drugs | 1483 | counts | 0 | Disease history |
| 17 | Antioxidant against lipid peroxidation | 45 | counts | 0 | Disease history |
| 18 | Hypoglycemic drugs | 2594 | counts | 0 | Disease history |
| 19 | Antihypertensive drugs | 6271 | counts | 0 | Disease history |
| 20 | History: Smoking | 4420 | counts | 0 | Disease history |
| 21 | Cumulative smoking (years) | 0 (0-20) | median(IQR) | | |
| 22 | History: Drinking | 1976 | counts | 0 | Disease history |
| 23 | History: Ischemic stroke | 2916 | counts | 0 | Disease history |

| | | | | | |
|---|---|---|---|---|---|
| 24 | History: Ischemic stroke times | | | 0 | |
| | Once | 2363 | counts | | Disease history |
| | Twice | 382 | counts | | Disease history |
| | 3 Times | 170 | counts | | Disease history |
| 25 | History: Intracranial hemorrhage | 245 | counts | | Disease history |
| 26 | History: ICH times | | | 0 | |
| | Once | 230 | counts | | Disease history |
| | Twice | 15 | counts | | Disease history |
| | Three Times | 0 | counts | | Disease history |
| 27 | History: Subarachnoid hemorrhage | 21 | counts | | Disease history |
| 28 | History: SAH times | | | 0 | Disease history |
| | Once | 21 | counts | | |
| | Twice | 0 | counts | | |
| | 3 Times | 0 | counts | | |
| 29 | Stroke History | 3111 | counts | 0 | Disease history |
| 30 | Transient ischemic attack | 285 | counts | 0 | Disease history |
| 31 | Lipid metabolism disorders | 1053 | counts | 0 | Disease history |
| 32 | Years of lipid metabolism disorders | 0(0-0) | Median (IQR) | 0 | Disease history |
| 33 | Diabetes | 3274 | counts | 0 | Disease history |
| 34 | Years of diabetes | 0(0-0) | Median (IQR) | | |
| 35 | Hypertension | 8786 | counts | 0 | Disease history |
| 36 | Years of hypertension | 3(0-10) | Median (IQR) | 0 | Disease history |
| 37 | Heart disease | 1940 | counts | 0 | Disease history |

| | | | | | |
|---|---|---|---|---|---|
| 38 | Coronary heart disease | 1459 | counts | 0 | Disease history |
| 39 | Atrial fibrillation | 948 | counts | 0 | Disease history |
| 40 | Physical activity | 7767 | counts | 0 | Disease history |
| 41 | Heart failure | 92 | counts | 0 | Disease history |
| 42 | Heart valve disease | 53 | counts | 0 | Disease history |
| 43 | Infective endocarditis | 2 | counts | 0 | Disease history |
| 44 | Non-bacterial thrombotic endocarditis | 2 | counts | 0 | Disease history |
| 45 | Dilated cardiomyopathy | 18 | counts | 0 | Disease history |
| 46 | Rheumatic heart disease | 47 | counts | 0 | Disease history |
| 47 | Migraine | 13573 | counts | 0 | Disease history |
| 48 | Peripheral arterial disease | 102 | counts | 0 | Disease history |
| 49 | Carotid artery stenosis | 116 | counts | 0 | Disease history |
| 50 | Venous thromboembolism | 39 | counts | 0 | Disease history |
| 51 | Pulmonary embolism | 21 | counts | 0 | Disease history |
| 52 | Cancer | 124 | counts | 0 | Disease history |
| 53 | Dementia | 44 | counts | 0 | Disease history |
| 54 | Mental disorders | 34 | counts | 0 | Disease history |
| 55 | Epilepsy | 46 | counts | 0 | Disease history |
| 56 | Sickle cell disease | 11 | counts | 0 | Disease history |
| 57 | Pregnancy or postpartum within 6 weeks | 6 | counts | 0 | Disease history |
| 58 | Chronic obstructive pulmonary disease | 122 | counts | 0 | Disease history |

| | | | | | |
|---|---|---|---|---|---|
| 59 | Sleep apnea | 107 | counts | 0 | Disease history |
| 60 | Asthma | 63 | counts | 0 | Disease history |
| 61 | Liver dysfunction | 74 | counts | 0 | Disease history |
| 62 | Cirrhosis | 20 | counts | 0 | Disease history |
| 63 | Gastrointestinal ulcers; | 150 | counts | 0 | Disease history |
| 64 | Gastrointestinal bleeding | 74 | counts | 0 | Disease history |
| 65 | Arthritis | 259 | counts | 0 | Disease history |
| 66 | Hypertension | 2959 | counts | 0 | Family history |
| 67 | Hyperlipidemia | 3010 | counts | 0 | Family history |
| 68 | Diabetes | 912 | counts | 0 | Family history |
| 69 | Stroke | 1845 | counts | 0 | Family history |
| 70 | Coronary heart disease | 665 | counts | 0 | Family history |
| 71 | Cancer | 484 | counts | 0 | Family history |
| 72 | Migraine with visual aura | 27 | counts | 0 | Family history |
| 73 | Renal insufficiency | 122 | counts | 0 | Disease history |
| 74 | Migraine within 1 year | 37 | counts | 0 | Disease history |
| 75 | Adiponectin | 2.35;2.06 | mean; SD | 4067 | Biomarkers |
| 76 | Albumin/Globulin | 1.6;0.33 | mean; SD | 2447 | Biomarkers |
| 77 | Advanced glycation end products (AGEs) | 218.71; 69.08 | mean; SD | 10067 | Biomarkers |
| 78 | Biomarkers: Albumin | 40.42;4.07 | mean; SD | 1047 | Biomarkers |
| 79 | Biomarkers: Alkaline Phosphatase | 79.68; 86.64 | mean; SD | 2520 | Biomarkers |
| 80 | Alanine Aminotransferase | 22.1;17.37 | mean; SD | 842 | Biomarkers |
| 81 | Angptl3 | 98.76;48.83 | mean; SD | 4012 | Biomarkers |
| 82 | Aspartate Aminotransferase | 21.62;13.07 | mean; SD | 937 | Biomarkers |
| 83 | Vitamin B12 | 355.33; 338.88 | mean; SD | 5217 | Biomarkers |
| 84 | Folic Acid | 13.08;12.73 | mean; SD | 4500 | Biomarkers |
| 85 | Creatine Kinase | 108.21;229.2 | mean; SD | 5542 | Biomarkers |

| | | | | | |
|---|---|---|---|---|---|
| 86 | Creatine Kinase-Mb Quality | 1.53;2.08 | mean; SD | 13612 | Biomarkers |
| 87 | Creatine Kinase-Mb | 14.02;8.49 | mean; SD | 6655 | Biomarkers |
| 88 | Absolute Value of Basophils | 0.03;0.19 | mean; SD | 442 | Biomarkers |
| 89 | Absolute Value of Eosinophils | 0.15;1.49 | mean; SD | 369 | Biomarkers |
| 90 | Hematocrit | 36.13;15.21 | mean; SD | 187 | Biomarkers |
| 91 | Hemoglobin | 140.79;16.98 | mean; SD | 199 | Biomarkers |
| 92 | Absolute Value of Lymphocyte Population | 1.79;0.82 | mean; SD | 233 | Biomarkers |
| 93 | Average Hemoglobin Level | 30.56;4.6 | mean; SD | 184 | Biomarkers |
| 94 | Average Hemoglobin Concentration | 334.69;15.64 | mean; SD | 194 | Biomarkers |
| 95 | Mean Volume of Red Blood Cells | 91.04;6.01 | mean; SD | 296 | Biomarkers |
| 96 | Absolute Value of Mononuclear Cell Population | 0.45;0.21 | mean; SD | 273 | Biomarkers |
| 97 | Average Platelet Volume | 10.03;1.41 | mean; SD | 335 | Biomarkers |
| 98 | Absolute Value of Neutrophils | 4.93;2.27 | mean; SD | 186 | Biomarkers |
| 99 | Platelet Volume | 0.5;3.11 | mean; SD | 1246 | Biomarkers |
| 100 | Platelet Distribution Width | 13.5;3.8 | mean; SD | 6097 | Biomarkers |
| 101 | Platelet Volume Distribution Width | 15.31;7.33 | mean; SD | 8518 | Biomarkers |
| 102 | Ratio of Large Platelets | 27.45;10.08 | mean; SD | 4637 | Biomarkers |
| 103 | Absolute Value of Platelets | 217.59;64.23 | mean; SD | 209 | Biomarkers |
| 104 | Absolute Value of Red Blood Cells | 4.66;3.97 | mean; SD | 176 | Biomarkers |
| 105 | Red Blood Cell Distribution Width Sd | 42.3;5.93 | mean; SD | 3739 | Biomarkers |
| 106 | Red Blood Cell Distribution Width CV | 12.81;2.13 | mean; SD | 544 | Biomarkers |
| 107 | Absolute Value of White Blood Cells | 7.55;26.4 | mean; SD | 179 | Biomarkers |
| 108 | Calcium | 2.26;1.05 | mean; SD | 4095 | Biomarkers |

| | | | | | |
|---|---|---|---|---|---|
| 109 | Partial Thromboplastin Time | 30.06;5.93 | mean; SD | 424 | Biomarkers |
| 110 | International Normalized Ratio | 1;0.13 | mean; SD | 406 | Biomarkers |
| 111 | Prothrombin Time | 11.95;1.62 | mean; SD | 357 | Biomarkers |
| 112 | Thrombin Time | 16.95;4.68 | mean; SD | 1046 | Biomarkers |
| 113 | Cholinesterase | 8738.35; 70913.16 | mean; SD | 7310 | Biomarkers |
| 114 | Total Cholesterol | 4.12; 1.23 | mean; SD | 4079 | Biomarkers |
| 115 | Chlorine | 104.08; 3.85 | mean; SD | 2997 | Biomarkers |
| 116 | Carbon Dioxide Binding Capacity | 24.9; 3.14 | mean; SD | 4933 | Biomarkers |
| 117 | C-Peptide | 813.01; 521.58 | mean; SD | 7769 | Biomarkers |
| 118 | Creatinine | 73.22; 32.56 | mean; SD | 4107 | Biomarkers |
| 119 | C-Reactive Protein | 7.25; 14.97 | mean; SD | 11401 | Biomarkers |
| 120 | Cystatin C | 1; 0.3 | mean; SD | 3929 | Biomarkers |
| 121 | Biomarkers: Direct Bilirubin | 4.31; 2.45 | mean; SD | 1240 | Biomarkers |
| 122 | Biomarkers: D-Dimer Dd | 1.73; 2.59 | mean; SD | 4312 | Biomarkers |
| 123 | Biomarkers: Esrage | 0.41; 0.28 | mean; SD | 10069 | Biomarkers |
| 124 | Biomarkers: Iron | 15.6; 8.3 | mean; SD | 12689 | Biomarkers |
| 125 | Biomarkers: Free Fatty Acid | 0.77; 0.36 | mean; SD | 10422 | Biomarkers |
| 126 | Biomarkers: Fibrinogen Fib | 394.23; 138.57 | mean; SD | 4307 | Biomarkers |
| 127 | Biomarkers: Fasting Blood Glucose | 6.45; 2.62 | mean; SD | 2812 | Biomarkers |
| 128 | Biomarkers: Glycated Albumin | 16.86; 5.74 | mean; SD | 10405 | Biomarkers |
| 129 | γ-glutamyl Transpeptidase | 35; 40.27 | mean; SD | 2127 | Biomarkers |
| 130 | Globulin | 26.07; 4.61 | mean; SD | 2251 | Biomarkers |
| 131 | Random Blood Glucose | 8.32; 4.19 | mean; SD | 12294 | Biomarkers |
| 132 | Glycosylated Hemoglobin | 6.54; 1.83 | mean; SD | 5867 | Biomarkers |
| 133 | A-Hydroxybutyrate Dehydrogenase | 142.76; 43.97 | mean; SD | 8245 | Biomarkers |
| 134 | Homocysteine Hcy | 19.11; 12.46 | mean; SD | 4329 | Biomarkers |
| 135 | High-Density Lipoprotein | 0.96; 0.3 | mean; SD | 4079 | Biomarkers |
| 136 | High-Sensitivity C-Reactive Protein | 8.04; 26.92 | mean; SD | 4067 | Biomarkers |
| 137 | Indirect Bilirubin | 10.43; 5.63 | mean; SD | 3374 | Biomarkers |
| 138 | Interleukin-1 Receptor Antagonist | 505.02; 548.19 | mean; SD | 3766 | Biomarkers |
| 139 | Interleukin 6 | 4.5;4.7 | mean;SD | 3817 | Biomarkers |

| | | | | | |
|---|---|---|---|---|---|
| 140 | Interleukin-6 Receptor | 41567.06; 15520.12 | mean; SD | 3752 | Biomarkers |
| 141 | Insulin | 86.52; 187.25 | mean; SD | 7743 | Biomarkers |
| 142 | Potassium | 3.92; 0.4 | mean; SD | 2535 | Biomarkers |
| 143 | Lactate Dehydrogenase | 190.04; 70.85 | mean; SD | 4659 | Biomarkers |
| 144 | Low-Density Lipoprotein | 2.45; 1.08 | mean; SD | 4079 | Biomarkers |
| 145 | LDL Receptor | 17.59; 11.76 | mean; SD | 4064 | Biomarkers |
| 146 | Lipoprotein Phospholipase A2-Activity | 162.5; 50.48 | mean; SD | 3872 | Biomarkers |
| 147 | Lipoprotein Phospholipase A2-Mass | 182.1; 76.66 | mean; SD | 3744 | Biomarkers |
| 148 | Lipoprotein A | 228.1; 256.38 | mean; SD | 4137 | Biomarkers |
| 149 | Monocyte Chemoattractant Protein-1 | 312.55; 363.24 | mean; SD | 3789 | Biomarkers |
| 150 | Magnesium | 0.88; 0.51 | mean; SD | 7774 | Biomarkers |
| 151 | Methylmalonic Acid | 332.49; 653.82 | mean; SD | 4554 | Biomarkers |
| 152 | Sodium | 140.61; 3.28 | mean; SD | 2553 | Biomarkers |
| 153 | Ngal | 95940.68; 102304.32 | mean; SD | 9258 | Biomarkers |
| 154 | Oxidized LDL | 104069.05; 45681.34 | mean; SD | 11292 | Biomarkers |
| 155 | OXLDL Receptor 1 | 797.81; 623.85 | mean; SD | 11258 | Biomarkers |
| 156 | Phosphorus | 1.07; 0.21 | mean; SD | 6674 | Biomarkers |
| 157 | Prealbumin | 930.85; 13379.73 | mean; SD | 7865 | Biomarkers |
| 158 | Proprotein Convertase Subtilisin 9 | 370.81; 133.14 | mean; SD | 3747 | Biomarkers |
| 159 | sRAGE | 1101.36; 658.08 | mean; SD | 10086 | Biomarkers |
| 160 | Total Bile Acids | 5.15; 7.82 | mean; SD | 4604 | Biomarkers |
| 161 | Total Bilirubin | 14.6; 7.04 | mean; SD | 1091 | Biomarkers |
| 162 | Triglyceride | 1.6; 0.92 | mean; SD | 4079 | Biomarkers |
| 163 | Total Protein | 66.42; 6.18 | mean; SD | 1165 | Biomarkers |
| 164 | Uric Acid | 301.91; 91.41 | mean; SD | 5394 | Biomarkers |

| | | | | | |
|---|---|---|---|---|---|
| 165 | Urea | 5.26;2.3 | mean; SD | 9118 | Biomarkers |
| 166 | Human Chitinase 3-like | 88949.97; 65468.28 | mean; SD | 3752 | Biomarkers |
| 167 | Apo-A1 | 0.8;3.6 | mean; SD | 4513 | Biomarkers |
| 168 | Apo-C3 | 0.25;0.13 | mean; SD | 4500 | Biomarkers |
| 169 | Apo-E | 0.05;0.02 | mean; SD | 4500 | Biomarkers |
| 170 | Apo-A2 | 0.3;0.09 | mean; SD | 4501 | Biomarkers |
| 171 | Apo-B | 1.56;0.78 | mean; SD | 4499 | Biomarkers |
| 172 | Apo-C2 | 0.13;0.08 | mean; SD | 4500 | Biomarkers |
| 173 | Betaine | 47.22;25.19 | mean; SD | 3891 | Biomarkers |
| 174 | Trimethylamine oxide | 2.35;3.19 | mean; SD | 3880 | Biomarkers |
| 175 | Choline | 14.04;4.76 | mean; SD | 3889 | Biomarkers |
| 176 | Carnitine | 50.67;14.06 | mean; SD | 3887 | Biomarkers |
| 177 | Trimethylpentaaminovaleric acid | 0.47;0.79 | mean; SD | 3893 | Biomarkers |
| 178 | Butyl betaine | 1.05;0.49 | mean; SD | 3887 | Biomarkers |
| 179 | Trimethyllysine | 1.13;0.71 | mean; SD | 4750 | Biomarkers |
| 180 | Cholesterol efflux capacity | 20.69;26.54 | mean; SD | 8964 | Biomarkers |
| 181 | Acetylcholine | 1.26;0.49 | mean; SD | 5356 | Biomarkers |
| 182 | Phosphocholine | 0.99;0.55 | mean; SD | 5339 | Biomarkers |
| 183 | Northeast China | 1705 | counts | 0 | Geographic |
| 184 | East China | 3642 | counts | 0 | Geographic |
| 185 | South China | 839 | counts | 0 | Geographic |
| 186 | Central China | 2933 | counts | 0 | Geographic |
| 187 | Southwest China | 287 | counts | 0 | Geographic |
| 188 | Northwest China | 1061 | counts | 0 | Geographic |
| 189 | Northern China | 3473 | counts | 0 | Geographic |
| 190 | Large artery atherosclerosis Stroke | 3845 | counts | 0 | Toast |
| 191 | Cardiac embolic Stroke | 911 | counts | 0 | Toast |
| 192 | Small vascular occlusion Stroke | 3164 | counts | 0 | Toast |
| 193 | Other determination Stroke | 182 | counts | 0 | Toast |
| 194 | Undetermined Stroke | 7022 | counts | 0 | Toast |
| 195 | NIH1A at administration | 0(0-0) | Media (IQR) | 0 | NIHSS/mRS information |
| 196 | NIH1B at administration | 0(0-0) | Media (IQR) | 0 | NIHSS/mRS information |
| 197 | NIH1C at administration | 0(0-0) | Media (IQR) | 0 | NIHSS/mRS information |

| | | | | | |
|---|---|---|---|---|---|
| 198 | NIH2 at administration | 0(0-0) | Media (IQR) | 0 | NIHSS/mRS information |
| 199 | NIH3 at administration | 0(0-0) | Media (IQR) | 0 | NIHSS/mRS information |
| 200 | NIH4 at administration | 1(0-1) | Media (IQR) | 0 | NIHSS/mRS information |
| 201 | NIH5A at administration | 0(0-1) | Media (IQR) | 0 | NIHSS/mRS information |
| 202 | NIH5B at administration | 0(0-1) | Media (IQR) | 0 | NIHSS/mRS information |
| 203 | NIH6A at administration | 0(0-1) | Media (IQR) | 0 | NIHSS/mRS information |
| 204 | NIH6B at administration | 0(0-1) | Media (IQR) | 0 | NIHSS/mRS information |
| 205 | NIH7 at administration | 0(0-0) | Media (IQR) | 1 | NIHSS/mRS information |
| 206 | NIH8 at administration | 0(0-1) | Media (IQR) | 1 | NIHSS/mRS information |
| 207 | NIH9 at administration | 0(0-1) | Media (IQR) | 0 | NIHSS/mRS information |
| 208 | NIH10 at administration | 0(0-1) | Media (IQR) | 1 | NIHSS/mRS information |
| 209 | NIH11 at administration | 0(0-0) | Media (IQR) | 19 | NIHSS/mRS information |
| 210 | NIHSS at administration | 3(2-6) | Media (IQR) | 0 | NIHSS/mRS information |
| 211 | Delta NIHSS (NIHSS at administration - NIHSS at discharge) | 1(0-3) | Media (IQR) | 0 | NIHSS/mRS information |
| 212 | Delta NIH1A (NIHS1A at administration - NIHS1A at discharge) | 0(0-0) | Media (IQR) | 0 | NIHSS/mRS information |
| 213 | Delta NIH1B (NIHS1B at administration - NIHS1B at discharge) | 0(0-0) | Media (IQR) | 0 | NIHSS/mRS information |
| 214 | Delta NIH1C (NIHS1B at administration - NIHS1B at discharge) | 0(0-0) | Media (IQR) | 0 | NIHSS/mRS information |
| 215 | Delta NIH2 (NIHS2 at administration - NIHS2 at discharge) | 0(0-0) | Media (IQR) | 1 | NIHSS/mRS information |

| | | | | | |
|---|---|---|---|---|---|
| 216 | Delta NIHS3 (NIHS3 at administration - NIHS3 at discharge) | 0(0-0) | Media (IQR) | 0 | NIHSS/mRS information |
| 217 | Delta NIH4 (NIHS4 at administration - NIHS4 at discharge) | 0(0-0) | Media (IQR) | 0 | NIHSS/mRS information |
| 218 | Delta NIHS5A (NIHS5A at administration - NIHS5A at discharge) | 0(0-0) | Media (IQR) | 0 | NIHSS/mRS information |
| 219 | Delta NIHS5B (NIHS5B at administration - NIHS5B at discharge) | 0(0-0) | Media (IQR) | 0 | NIHSS/mRS information |
| 220 | Delta NIHS6A (NIHS6A at administration - NIHS6A at discharge) | 0(0-0) | Media (IQR) | 0 | NIHSS/mRS information |
| 221 | Delta NIHS6B (NIHS6B at administration - NIHS6B at discharge) | 0(0-0) | Media (IQR) | 0 | NIHSS/mRS information |
| 222 | Delta NIHS7 (NIHS7 at administration - NIHS7 at discharge) | 0(0-0) | Media (IQR) | 0 | NIHSS/mRS information |
| 223 | Delta NIHS8 (NIHS8 at administration - NIHS8 at discharge) | 0(0-0) | Media (IQR) | 0 | NIHSS/mRS information |
| 224 | Delta NIHS9 (NIHS9 at administration - NIHS9 at discharge) | 0(0-0) | Media (IQR) | 0 | NIHSS/mRS information |
| 225 | Delta NIHS10 (NIHS10 at administration - NIHS10 at discharge) | 0(0-0) | Media (IQR) | 0 | NIHSS/mRS information |
| 226 | Delta NIHS11 (NIHS11 at administration - NIHS11 at discharge) | 0(0-0) | Media (IQR) | 0 | NIHSS/mRS information |
| 227 | NIH1A at discharge | 0(0-0) | Media (IQR) | 0 | NIHSS/mRS information |
| 228 | NIH1B at discharge | 0(0-0) | Media (IQR) | 0 | NIHSS/mRS information |
| 229 | NIH1C at discharge | 0(0-0) | Media (IQR) | 0 | NIHSS/mRS information |
| 230 | NIH2 at discharge | 0(0-0) | Media (IQR) | 1 | NIHSS/mRS information |

| | | | | | |
|---|---|---|---|---|---|
| 231 | NIH3 at discharge | 0(0-0) | Media (IQR) | 0 | NIHSS/mRS information |
| 232 | NIH4 at discharge | 0(0-1) | Media (IQR) | 0 | NIHSS/mRS information |
| 233 | NIH5A at discharge | 0(0-0) | Media (IQR) | 0 | NIHSS/mRS information |
| 234 | NIH5B at discharge | 0(0-0) | Media (IQR) | 0 | NIHSS/mRS information |
| 235 | NIH6A at discharge | 0(0-0) | Media (IQR) | 0 | NIHSS/mRS information |
| 236 | NIH6B at discharge | 0(0-0) | Media (IQR) | 0 | NIHSS/mRS information |
| 237 | NIH7 at discharge | 0(0-0) | Media (IQR) | 0 | NIHSS/mRS information |
| 238 | NIH8 at discharge | 0(0-0) | Media (IQR) | 0 | NIHSS/mRS information |
| 239 | NIH9 at discharge | 0(0-0) | Media (IQR) | 0 | NIHSS/mRS information |
| 240 | NIH10 at discharge | 0(0-0) | Media (IQR) | 0 | NIHSS/mRS information |
| 241 | NIH11 at discharge | 0(0-0) | Media (IQR) | 26 | NIHSS/mRS information |
| 242 | NIHSS at discharge | 2(0-4) | Media (IQR) | 0 | NIHSS/mRS information |
| 243 | History: mRS (before onset) | | | | |
| | 0-No symptoms at all | 10243 | counts | 0 | NIHSS/mRS information |
| | 1-No significant disability despite symptoms | 2393 | counts | 0 | NIHSS/mRS information |
| | 2-Slight disability | 664 | counts | 0 | NIHSS/mRS information |
| | 3-Moderate disability | 302 | counts | 0 | NIHSS/mRS information |
| | 4-Moderately severe disability | 292 | counts | 0 | NIHSS/mRS information |
| | 5-Severe disability | 46 | counts | 0 | NIHSS/mRS information |
| 244 | mRS score at administration | | | | |
| | 0-No symptoms at all | 1401 | counts | 0 | NIHSS/mRS information |
| | 1-No significant disability despite symptoms | 5016 | counts | 0 | NIHSS/mRS information |

| | | | | | |
|---|---|---|---|---|---|
| | 2-Slight disability | 2794 | counts | 0 | NIHSS/mRS information |
| | 3-Moderate disability | 1777 | counts | 0 | NIHSS/mRS information |
| | 4-Moderately severe disability | 2588 | counts | 0 | NIHSS/mRS information |
| | 5-Severe disability | 364 | counts | 0 | NIHSS/mRS information |
| 245 | mRS score at discharge | | | | |
| | 0-No symptoms at all | 2845 | counts | 0 | NIHSS/mRS information |
| | 1-No significant disability despite symptoms | 6296 | counts | 0 | NIHSS/mRS information |
| | 2-Slight disability | 2159 | counts | 0 | NIHSS/mRS information |
| | 3-Moderate disability | 1204 | counts | 0 | NIHSS/mRS information |
| | 4-Moderately severe disability | 1242 | counts | 0 | NIHSS/mRS information |
| | 5-Severe disability | 194 | counts | 0 | NIHSS/mRS information |
| 246 | Ambulance informed the hospital in advance | 792 | counts | 12696 | Hospitalization information |
| 247 | Admitting Diagnosis: | 13940 | counts | 0 | Hospitalization information |
| | Cerebral infarction | 13940 | counts | 0 | Hospitalization information |
| | Transient Cerebral ischemia | 0 | Counts | 0 | Hospitalization information |
| 248 | History of sleep disorders | 215 | counts | 11554 | Hospitalization information |
| 249 | Sleep apnea-syndrome | 45 | counts | 13736 | Hospitalization information |
| 250 | Transient ischemic attack | 45 | counts | 85 | Hospitalization information |
| 251 | Hemorrhagic stroke | 26 | counts | 71 | Hospitalization information |
| 252 | Systemic embolic events | 27 | counts | 106 | Hospitalization information |

| | | | | | |
|---|---|---|---|---|---|
| 253 | Vascular related operation and surgical treatment | 284 | counts | 0 | Hospitalization information |
| 254 | Carotid artery stenting | 35 | counts | 13656 | Hospitalization information |
| 255 | Carotid endarterectomy | 7 | counts | 13656 | Hospitalization information |
| 256 | Intracranial arterial stenting | 22 | counts | 13656 | Hospitalization information |
| 257 | Decompressive craniectomy | 6 | counts | 13656 | Hospitalization information |
| 258 | Deep venous thrombosis prevention within 48 hours of admission | 1652 | counts | 10845 | Hospitalization information |
| 259 | Swallowing function: Evaluation before oral feeding, drinking, medication within 48 hours of admission | 12512 | counts | 0 | Hospitalization information |
| 260 | Evaluation during hospitalization | 10207 | counts | 0 | Hospitalization information |
| 261 | Cerebral infarction | 153 | counts | 0 | Hospitalization information |
| 262 | Hypertension | 10295 | counts | 0 | Hospitalization information |
| 263 | Pathoglycemia | 4522 | counts | 0 | Hospitalization information |
| 264 | Lipid metabolism disorder | 5127 | counts | 0 | Hospitalization information |
| 265 | Coronary heart disease | 2105 | counts | 0 | Hospitalization information |
| 266 | Atrial fibrillation | 919 | counts | 0 | Hospitalization information |
| 267 | Respiratory disease | 790 | counts | 0 | Hospitalization information |
| 268 | Liver disease | 845 | counts | 0 | Hospitalization information |
| 269 | Urinary system diseases | 397 | counts | 0 | Hospitalization information |
| 270 | Peripheral arterial disease | 779 | counts | 0 | Hospitalization information |
| 271 | Deep venous thrombosis | 77 | counts | 0 | Hospitalization information |

| | | | | | |
|---|---|---|---|---|---|
| 272 | Bleeding | 202 | counts | 0 | Hospitalization information |
| 273 | Epilepsy | 64 | counts | 0 | Hospitalization information |
| 274 | Smoking cessation mission | 11031 | counts | 0 | Hospitalization information |
| 275 | Whereabouts after discharge | 233 | counts | 220 | Hospitalization information |
| 276 | Antiplatelet agents (Discharge medication) | 12665 | counts | 0 | Hospitalization information |
| 277 | Anticoagulant drugs (Discharge medication) | 420 | counts | 0 | Hospitalization information |
| 278 | Lipid-lowering drugs (Discharge medication) | 12790 | counts | 0 | Hospitalization information |
| 279 | Hypoglycemic therapy (Discharge medication) | 3341 | counts | 0 | Hospitalization information |
| 280 | Antihypertensive therapy (Discharge medication) | 6901 | counts | 0 | Hospitalization information |
| 281 | Antioxidant against lipid peroxidation (Discharge medication) | 896 | counts | 0 | Hospitalization information |
| 282 | Antidepressant treatment in discharge description | 86 | counts | 11399 | Hospitalization information |
| 283 | Cognitive impairment treatment in discharge description | 23 | counts | 11399 | Hospitalization information |
| 284 | Anti-insomnia treatment in discharge description | 60 | counts | 11403 | Hospitalization information |
| 285 | Sleep disorder | 253 | counts | 11413 | Hospitalization information |
| 286 | Post-stroke depression | 117 | counts | 11416 | Hospitalization information |
| 287 | Mechanical thrombectomy | 39 | counts | 0 | Hospitalization information |
| 288 | Stent Therapy | 29 | counts | 0 | Hospitalization information |
| 289 | Anti-platelet therapy | 13450 | counts | 101 | Hospitalization information |
| 290 | Anticoagulation treatment during hospitalization | 1438 | counts | 101 | Hospitalization information |
| 291 | Lipid-lowering drugs | 13376 | counts | 101 | Hospitalization information |

| | | | | | |
|---|---|---|---|---|---|
| 292 | Antioxidant treatment during hospitalization | 2570 | counts | 101 | Hospitalization information |
| 293 | Hypoglycemic treatment | 3558 | counts | 101 | Hospitalization information |
| 294 | Antihypertensive treatment | 6483 | counts | 101 | Hospitalization information |
| 295 | Volume expansion therapy | 2255 | counts | 103 | Hospitalization information |
| 296 | Dehydration treatment to reduce intracranial pressure | 759 | counts | 102 | Hospitalization information |
| 297 | Anti-infective treatment during hospitalization | 989 | counts | 102 | Hospitalization information |
| 298 | Traditional Chinese medicine therapy during hospitalization | 8137 | counts | 103 | Hospitalization information |
| 299 | Intravenous Thrombolysis: rt-PA intravenous thrombolytic | 1231 | | 2 | Hospitalization information |
| 300 | Pre-hospital transportation | | | 3861 | Hospitalization information |
| | 1-Ambulance | 1491 | counts | | Hospitalization information |
| | 2-Private car | 7008 | counts | | Hospitalization information |
| | 3-Taxi | 1580 | counts | | Hospitalization information |
| 301 | Heart rate | 76(68-80) | Media (IQR) | 146 | Hospitalization information |
| 302 | Blood pressure at admission | 150(136-165) | Media (IQR) | 14 | Hospitalization information |
| 303 | Blood pressure at admission (R) | 87(80-96) | Media (IQR) | 373 | Hospitalization information |
| 304 | Blood pressure at admission (L) | 149(135-163) | Media (IQR) | 14 | Hospitalization information |
| 305 | Blood pressure at admission (L) | 85(79-95) | Media (IQR) | 361 | Hospitalization information |
| 306 | mRS score (within 24 hours after admission) | 2(1-3) | Media (IQR) | 0 | Hospitalization information |
| 307 | Blood pressure at admission: SBP | 149(135-164) | Media (IQR) | 10 | Hospitalization information |

| | | | | | |
|---|---|---|---|---|---|
| 308 | Blood pressure at admission: DBP | 86(79-95) | Media (IQR) | 273 | Hospitalization information |
| 309 | Polygenic Risk Score Calculated by GIGASTROKE | / | / | / | Genetic Information |

**Table S3. Stroke recurrence risk in 3 months evaluated by RRE model in CNSR- III cohort**

| Recurrence risk | 0.007 | 0.011 | 0.034 | 0.039 | 0.042 | 0.059 | 0.191 | 0.273 | 0.388 | 0.453 |
|---|---|---|---|---|---|---|---|---|---|---|
| Frequency | 5,328 | 242 | 3,453 | 1,158 | 163 | 2,819 | 159 | 355 | 257 | 6 |

**Table S4. The method and rate of follow-ups in development and validation cohort1 of CNSR-III**

| | Development Cohort (N=11,313) | | | | Validation Cohort 1 (N=2,627) | | | |
|---|---|---|---|---|---|---|---|---|
| | Face-to-face | Telephone | Lost | follow-up rate (%) | Face-to-face | telephone | Lost | follow-up rate (%) |
| 3 months | 7,455 | 3,724 | 134 | 98.82 | 1,653 | 950 | 24 | 99.09 |
| 6 months | 1,089 | 9,934 | 290 | 97.44 | 210 | 2,348 | 69 | 97.37 |
| 12 months | 6,009 | 4,815 | 489 | 95.68 | 1,437 | 1,072 | 118 | 95.51 |
| 2 years | 372 | 10,109 | 832 | 92.65 | 94 | 2,347 | 186 | 92.92 |
| 3 years | 145 | 9,939 | 1,229 | 89.14 | 42 | 2,260 | 319 | 87.86 |
| 4 years | 61 | 9,466 | 1,786 | 84.21 | 34 | 2,184 | 409 | 84.43 |
| 5 years | 250 | 8,757 | 2,306 | 79.62 | 29 | 2,072 | 526 | 79.98 |

**Table S5. The description of endpoint events in development, validation cohort 1 and 2.**

| | Development cohort | Validation cohort 1 | Validation cohort 2 |
|---|---|---|---|
| 3 months | | | |
| recurrence | 716 (6.33%) | 147 (5.60%) | 356 (6.90%) |
| death | 134 (1.18%) | 46 (1.75%) | 25 (0.48%) |
| disability | 1556 (13.75%) | 448 (17.05%) | 147 (2.85%) |
| 6 months | | | |
| recurrence | 895 (7.91%) | 193 (7.35%) | / |
| death | 216 (1.91%) | 68 (2.59%) | / |
| disability | 1411 (12.47%) | 408 (15.53%) | / |
| 1 year | | | |
| recurrence | 1098 (9.71%) | 260 (9.90%) | 460 (8.92%) |
| death | 351 (3.10%) | 98 (3.73%) | 78 (1.51%) |
| disability | 1468 (12.98%) | 432 (16.44%) | 223 (3.48%) |
| 2 years | | | |
| recurrence | 1397 (12.35%) | 330 (12.56%) | / |
| death | 629 (5.56%) | 170 (6.47%) | / |
| disability | 1028 (9.09%) | 264 (10.05%) | / |
| 3 years | | | |
| recurrence | 1459 (12.90%) | 392 (14.92%) | / |
| death | 866 (7.65%) | 231 (8.79%) | / |
| disability | 992 (8.77%) | 269 (10.24%) | / |
| 4 years | | | |
| recurrence | 1514 (13.38%) | 447 (17.02%) | / |
| death | 1114 (9.84%) | 288 (10.96%) | / |
| disability | 927 (8.19%) | 247 (9.40%) | / |
| 5 years | | | |
| recurrence | 1549 (13.69%) | 482 (18.35%) | / |
| death | 1024 (9.05%) | 341 (12.98%) | / |
| disability | 892 (7.88%) | 239 (9.10%) | / |

Stroke recurrence was defined as any new stroke event identified through readmission records or patient self-reports during follow-up.

Death was defined as all-cause mortality and each fatality was confirmed using the Population Mortality Registration and Management Information System within the Life Registration Information Management System of the Chinese Center for Disease Control and Prevention.

Disability was defined using a binary modified Rankin Scale (mRS) classification, with mRS 0-2 considered non-events, and mRS 3-6 deemed as events.

Due to the cohort design, the follow-up of validation cohort 2 is limited in 3 months and 1 year.

**Table S6. The feature importance ranking based on SHAP and XGBoost**

| Feature name | Shap score | Ranking (shap) | Xgboost score | Ranking (xgboost) |
|---|---|---|---|---|
| NIHSS change from admission to discharge: Total score of NIHSS at discharge | 0.199 | 1 | 0.346 | 1 |
| Discharge status: mRS score at discharge | 0.080 | 2 | 0.058 | 2 |
| Choline | 0.062 | 3 | 0.014 | 4 |
| Discharge NIHSS: Total score of NIHSS at discharge | 0.030 | 4 | 0.012 | 7 |
| Inpatient Therapy: Anti-infective treatment during hospitalization | 0.027 | 5 | 0.010 | 12 |
| fasting blood glucose | 0.021 | 6 | 0.009 | 15 |
| NIHSS change from admission to discharge: NIHSS5a: Motor Arm-L | 0.013 | 7 | 0.010 | 11 |
| Inpatient Therapy: Volume expansion therapy | 0.012 | 8 | 0.014 | 5 |
| aspartate aminotransferase | 0.012 | 9 | 0.008 | 17 |
| Inpatient Event: Hemorrhagic stroke | 0.011 | 10 | 0.046 | 3 |
| Bbetaine | 0.010 | 11 | 0.002 | 97 |
| Monocyte Chemoattractant Protein-1 | 0.008 | 12 | 0.008 | 18 |
| phosphorus | 0.006 | 13 | 0.012 | 8 |
| lipoprotein phospholipase A2-activity | 0.006 | 14 | 0.010 | 10 |
| Discharge Medication: Lipid-lowering drugs | 0.006 | 15 | 0.006 | 26 |
| globulin | 0.005 | 16 | 0.009 | 13 |
| bsolute value of lymphocyte population | 0.005 | 17 | 0.011 | 9 |
| Inpatient Therapy: Anticoagulation treatment during hospitalization | 0.005 | 18 | 0.004 | 57 |
| urea nitrogen | 0.005 | 19 | 0.006 | 29 |
| Trimethylpentaaminovaleric acid | 0.005 | 20 | 0.007 | 19 |
| D-dimer DD (μg/ml); | 0.005 | 21 | 0.004 | 47 |
| Phosphcholine | 0.005 | 22 | 0.003 | 60 |
| Absolute value of mononuclear cell population | 0.004 | 23 | 0.014 | 6 |
| creatinine | 0.004 | 24 | 0.005 | 39 |
| absolute value of red blood cells | 0.004 | 25 | 0.005 | 42 |
| platelet volume | 0.004 | 26 | 0.009 | 16 |
| Pre-Hospital: Transportation | 0.004 | 27 | 0.006 | 33 |
| Absolute value of neutrophils | 0.003 | 28 | 0.003 | 77 |
| Discharge NIHSS1c: ’Blink eyes’ & ‘Squeeze hands’ | 0.003 | 29 | 0.006 | 25 |
| Inpatient Therapy: Dehydration treatment to reduce intracranial pressure | 0.003 | 30 | 0.001 | 127 |

| | | | | |
|---|---|---|---|---|
| Absolute value of platelets | 0.003 | 31 | 0.009 | 14 |
| Red blood cell distribution width CV | 0.003 | 32 | 0.003 | 83 |
| calcium | 0.003 | 33 | 0.006 | 31 |
| Vitamin B12 | 0.003 | 34 | 0.007 | 22 |
| thrombin time (s) | 0.003 | 35 | 0.007 | 23 |
| Waist circumference (cm) | 0.003 | 36 | 0.004 | 54 |
| Butyl betaine | 0.003 | 37 | 0.006 | 27 |
| ICONS: Neck circumference(cm) | 0.003 | 38 | 0.003 | 84 |
| sodium | 0.003 | 39 | 0.002 | 98 |
| interleukin 6 | 0.003 | 40 | 0.007 | 21 |
| creatine kinase | 0.003 | 41 | 0.005 | 35 |
| Fibrinogen Fib (mg/dl); | 0.003 | 42 | 0.003 | 61 |
| direct bilirubin | 0.003 | 43 | 0.006 | 30 |
| a-hydroxybutyrate dehydrogenase | 0.003 | 44 | 0.004 | 50 |
| total bilirubin | 0.002 | 45 | 0.005 | 45 |
| Inpatient Therapy: Deep venous thrombosis prevention within 48 hours of admission | 0.002 | 46 | 0.003 | 78 |
| History: Cumulative smoking(years) | 0.002 | 47 | 0.001 | 118 |
| Trimethylamine oxide | 0.002 | 48 | 0.004 | 48 |
| methylmalonic acid (nmol/L); | 0.002 | 49 | 0.003 | 68 |
| Occupation: leaders; technician; Clerical staff; Businessman; Production staff ; transport staff ; Soldier; other; unemployed; Retirement; Students; Unknown | 0.002 | 50 | 0.003 | 71 |
| C-reactive protein (CRP) | 0.002 | 51 | 0.004 | 46 |
| interleukin-6 receptor | 0.002 | 52 | 0.006 | 28 |
| WEIGHT1 | 0.002 | 53 | 0.004 | 56 |
| Baseline: Plasma: Apo-C3 | 0.002 | 54 | 0.003 | 67 |
| total protein | 0.002 | 55 | 0.005 | 43 |
| Carnitine | 0.002 | 56 | 0.003 | 66 |
| Baseline: Plasma: Apo-E | 0.002 | 57 | 0.004 | 52 |
| albumin/globulin | 0.002 | 58 | 0.003 | 76 |
| average hemoglobin concentration | 0.002 | 59 | 0.004 | 49 |
| triglyceride | 0.002 | 60 | 0.005 | 36 |
| Homocysteine HCY (μmol/L); | 0.002 | 61 | 0.004 | 51 |
| γ-glutamyl transpeptidase | 0.002 | 62 | 0.005 | 41 |
| magnesium | 0.002 | 63 | 0.002 | 100 |
| Inpatient Therapy: Antihypertensive treatment | 0.002 | 64 | 0.002 | 88 |
| ICONS: History of sleep disorders | 0.002 | 65 | 0.006 | 32 |
| hemoglobin | 0.002 | 66 | 0.005 | 37 |

| | | | | |
|---|---|---|---|---|
| Proprotein Convertase Subtilisin 9 | 0.002 | 67 | 0.003 | 59 |
| alkaline phosphatase | 0.002 | 68 | 0.005 | 40 |
| albumin | 0.002 | 69 | 0.007 | 24 |
| International Normalized Ratio | 0.002 | 70 | 0.004 | 58 |
| ICONS: Cognitive impairment treatment in discharge description | 0.002 | 71 | 0.001 | 129 |
| Physical examination: Heart rate | 0.002 | 72 | 0.003 | 75 |
| human chitinase 3-like protein 1 | 0.002 | 73 | 0.003 | 82 |
| adiponectin | 0.002 | 74 | 0.003 | 64 |
| glycosylated hemoglobin | 0.001 | 75 | 0.005 | 34 |
| Physical examination: Blood pressure at admission: systolic blood pressure | 0.001 | 76 | 0.003 | 72 |
| Final diagnosis: bleeding | 0.001 | 77 | 0.001 | 148 |
| ratio of large platelets | 0.001 | 78 | 0.002 | 93 |
| absolute value of white blood cells | 0.001 | 79 | 0.001 | 120 |
| Demography: Family monthly income per capita: <700 Yuan; 700~1500Yuan; 1501~2300Yuan; ＞2300Yuan;not known | 0.001 | 80 | 0.007 | 20 |
| Potassium | 0.001 | 81 | 0.005 | 38 |
| folic acid | 0.001 | 82 | 0.003 | 70 |
| high-sensitivity C-reactive protein (mg/L); | 0.001 | 83 | 0.004 | 55 |
| average hemoglobin level | 0.001 | 84 | 0.005 | 44 |
| total cholesterol | 0.001 | 85 | 0.003 | 81 |
| Iron | 0.001 | 86 | 0.002 | 85 |
| cystatin C (mg/L); | 0.001 | 87 | 0.002 | 114 |
| prealbumin | 0.001 | 88 | 0.003 | 63 |
| Platelet distribution width | 0.001 | 89 | 0.002 | 96 |
| Geographic location: Northeastern China | 0.001 | 90 | 0.001 | 146 |
| Chlorine | 0.001 | 91 | 0.003 | 73 |
| Discharge status: whereabouts after discharge | 0.001 | 92 | 0.002 | 111 |
| creatine kinase-MB | 0.001 | 93 | 0.003 | 74 |
| lipoprotein phospholipase A2-MASS | 0.001 | 94 | 0.002 | 95 |
| Glycated Albumin (%); | 0.001 | 95 | 0.003 | 79 |
| Admitting NIHSS6a: Motor Leg-Left | 0.001 | 96 | 0.002 | 101 |
| Final diagnosis: liver disease | 0.001 | 97 | 0.002 | 106 |
| Baseline: Plasma: Apo-C2 | 0.001 | 98 | 0.001 | 117 |
| partial thromboplastin time (s) | 0.001 | 99 | 0.004 | 53 |
| alanine aminotransferase | 0.001 | 100 | 0.001 | 142 |
| lipoprotein a | 0.001 | 101 | 0.003 | 62 |
| AGEs | 0.001 | 102 | 0.002 | 103 |

| | | | | |
|---|---|---|---|---|
| Admitting NIHSS9: Language | 0.001 | 103 | 0.001 | 116 |
| Limb rehabilitation: Evaluation during hospitalization | 0.001 | 104 | 0.002 | 105 |
| Physical examination: Blood pressure at admission: Right | 0.001 | 105 | 0.002 | 107 |
| Demography: Education level: college or above; high school; junior school; primary school; illiteracy; not known. | 0.001 | 106 | 0.002 | 115 |
| indirect bilirubin | 0.001 | 107 | 0.002 | 92 |
| Physical examination: Blood pressure at admission: Left systolic blood pressure | 0.001 | 108 | 0.002 | 90 |
| History: Heart disease category; Heart failure | 0.001 | 109 | 0.001 | 144 |
| Cholinesterase | 0.001 | 110 | 0.002 | 102 |
| Oxidized LDL (mU/L); | 0.001 | 111 | 0.001 | 122 |
| lactate dehydrogenase | 0.001 | 112 | 0.002 | 87 |
| total bile acids | 0.001 | 113 | 0.002 | 108 |
| Mean volume of red blood cells | 0.001 | 114 | 0.002 | 89 |
| NIHSS change from admission to discharge: NIHSS6b: Motor Leg-R | 0.001 | 115 | 0.001 | 133 |
| Discharge NIHSS6b: Motor Leg-R | 0.001 | 116 | 0.001 | 149 |
| Red blood cell distribution width SD | 0.001 | 117 | 0.003 | 65 |
| Absolute value of eosinophils | 0.001 | 118 | 0.003 | 80 |
| History: Ischemic stroketimes; Once; Twice; >= 3 times | 0.001 | 119 | 0.002 | 86 |
| Physical examination: mRS score (within 24 hours after admission) | 0.001 | 120 | 0.001 | 132 |
| Physical examination: Blood pressure at admission: Left | 0.001 | 121 | 0.002 | 110 |
| Age | 0.001 | 122 | 0.002 | 113 |
| OxLDL Receptor 1 (pg/ml); | 0.001 | 123 | 0.001 | 141 |
| Acetylcholine | 0.001 | 124 | 0.001 | 131 |
| Physical examination: Blood pressure at admission: diastolic blood pressure | 0.001 | 125 | 0.001 | 126 |
| Physical examination: Blood pressure at admission: Right | 0.001 | 126 | 0.003 | 69 |
| Discharge NIHSS5b: Motor Arm-R | 0.001 | 127 | 0.001 | 154 |
| apolipoprotein-A1 | 0.001 | 128 | 0.002 | 91 |
| Inpatient Therapy: Antioxidant treatment during hospitalization | 0.001 | 129 | 0.001 | 125 |
| Creatine kinase-MB quality | 0.001 | 130 | 0.001 | 130 |
| ANGPTL3 | 0.001 | 131 | 0.002 | 99 |

| | | | | |
|---|---|---|---|---|
| low-density lipoprotein | 0.001 | 132 | 0.002 | 94 |
| Discharge NIHSS1a: level of consciousness | 0.001 | 133 | 0.001 | 124 |
| Discharge NIHSS9: Language | 0.001 | 134 | 0.001 | 121 |
| esRAGE (ng/ml); | 0.001 | 135 | 0.001 | 128 |
| Discharge status: smoking cessation mission | 0.001 | 136 | 0.001 | 134 |
| LDL Receptor | 0.000 | 137 | 0.001 | 123 |
| carbon dioxide binding capacity | 0.000 | 138 | 0.001 | 137 |
| Platelet volume distribution width | 0.000 | 139 | 0.002 | 112 |
| ICONS: Sleep disorder | 0.000 | 140 | 0.001 | 138 |
| C-peptide (pmol/L); | 0.000 | 141 | 0.002 | 109 |
| Geographic location: Northern China | 0.000 | 142 | 0.001 | 119 |
| Average platelet volume | 0.000 | 143 | 0.002 | 104 |
| Admitting NIHSS7: Limb Ataxia | 0.000 | 144 | 0.001 | 151 |
| NIHSS change from admission to discharge: NIHSS4: Facial Palsy | 0.000 | 145 | 0.001 | 143 |
| ICONS: Sleep apnea-syndrome | 0.000 | 146 | 0.001 | 155 |
| high-density lipoprotein | 0.000 | 147 | 0.001 | 152 |
| History: Heart disease category; Atrial fibrillation | 0.000 | 148 | 0.001 | 150 |
| Height | 0.000 | 149 | 0.001 | 135 |
| Physical examination: Body mass index | 0.000 | 150 | 0.001 | 145 |
| Inpatient Therapy: Anti-platelet therapy | 0.000 | 151 | 0.001 | 136 |
| Baseline: Plasma: Apo-A2 | 0.000 | 152 | 0.000 | 162 |
| Trimethyllysine | 0.000 | 153 | 0.001 | 153 |
| NGAL (ng/ml); | 0.000 | 154 | 0.001 | 156 |
| NIHSS change from admission to discharge: NIHSS9: Language | 0.000 | 155 | 0.000 | 158 |
| prothrombin time (s) | 0.000 | 156 | 0.001 | 147 |
| random blood glucose | 0.000 | 157 | 0.001 | 139 |
| Absolute value of basophils | 0.000 | 158 | 0.001 | 157 |
| sRAGE (pg/ml); | 0.000 | 159 | 0.001 | 140 |
| Admitting NIHSS: Total score | 0.000 | 160 | 0.000 | 159 |
| interleukin-1 Receptor Antagonist | 0.000 | 161 | 0.000 | 161 |
| Etiology according to TOAST system: arge artery atherosclerosis | 0.000 | 162 | 0.000 | 160 |
| Hematocrit | 0.000 | 163 | 0.000 | 163 |
| Basic Information: Gender; male; female | 0.000 | 164 | 0.000 | 164 |
| Demography: Race: Han; others | 0.000 | 164 | 0.000 | 164 |
| Demography: Marital status: 1-unmarried; 2-Married; 3-divorced; 4-widowed; 5-remarried; 98-Unknown | 0.000 | 164 | 0.000 | 164 |

| | | | | |
|---|---|---|---|---|
| Demography: Living conditions: 1-living alone; 2-living with others | 0.000 | 164 | 0.000 | 164 |
| Pre-Hospital: First department: Neurology Emergency; medical emergency; Neurosurgery Emergency;outpatient clinic; unknown | 0.000 | 164 | 0.000 | 164 |
| Medication history: Antiplatelet agents | 0.000 | 164 | 0.000 | 164 |
| Medication history: Anticoagulant agents; | 0.000 | 164 | 0.000 | 164 |
| Medication history: Lipid-lowering drugs | 0.000 | 164 | 0.000 | 164 |
| Medication history: Antioxidant against lipid peroxidation | 0.000 | 164 | 0.000 | 164 |
| Medication history: Hypoglycemic drugs | 0.000 | 164 | 0.000 | 164 |
| Medication history: Antihypertensive drugs | 0.000 | 164 | 0.000 | 164 |
| History: Other disease; mental disorders; | 0.000 | 164 | 0.000 | 164 |
| History: Other disease; mental disorders; | 0.000 | 164 | 0.000 | 164 |
| History: ICH times; Once; Twice; >= 3 times | 0.000 | 164 | 0.000 | 164 |
| History: SAH times; Once; Twice; 3 times | 0.000 | 164 | 0.000 | 164 |
| History: Stroke History; | 0.000 | 164 | 0.000 | 164 |
| History: Transient ischemic attack (TIA); | 0.000 | 164 | 0.000 | 164 |
| History: Years of lipid metabolism disorders | 0.000 | 164 | 0.000 | 164 |
| History: Years of diabetes | 0.000 | 164 | 0.000 | 164 |
| History: Years of hypertension (99 for UK) | 0.000 | 164 | 0.000 | 164 |
| History: Heart disease; | 0.000 | 164 | 0.000 | 164 |
| History: Heart disease category; Coronary heart disease | 0.000 | 164 | 0.000 | 164 |
| History: Physical activity; | 0.000 | 164 | 0.000 | 164 |
| History: Heart valve disease | 0.000 | 164 | 0.000 | 164 |
| History: Heart disease category; Infective endocarditis | 0.000 | 164 | 0.000 | 164 |
| History: Heart disease category; non-bacterial thrombotic endocarditis | 0.000 | 164 | 0.000 | 164 |
| History: Heart disease category; Dilated cardiomyopathy | 0.000 | 164 | 0.000 | 164 |
| History: Heart disease category; Rheumatic heart disease | 0.000 | 164 | 0.000 | 164 |
| History: Migraine with visual aura; | 0.000 | 164 | 0.000 | 164 |
| History: Other disease; peripheral arterial disease | 0.000 | 164 | 0.000 | 164 |
| History: Other disease; carotid artery stenosis | 0.000 | 164 | 0.000 | 164 |
| History: venous thromboembolism | 0.000 | 164 | 0.000 | 164 |
| History: Other disease; pulmonary embolism | 0.000 | 164 | 0.000 | 164 |
| History: Other disease; cancer | 0.000 | 164 | 0.000 | 164 |
| History: Other disease; dementia | 0.000 | 164 | 0.000 | 164 |

| | | | | |
|---|---|---|---|---|
| History: Other disease; mental disorders | 0.000 | 164 | 0.000 | 164 |
| History: Other disease; epilepsy | 0.000 | 164 | 0.000 | 164 |
| History: Other disease; sickle cell disease | 0.000 | 164 | 0.000 | 164 |
| History: Other disease; Pregnancy or postpartum within 6 weeks | 0.000 | 164 | 0.000 | 164 |
| History: Other disease; Chronic obstructive pulmonary disease (COPD) | 0.000 | 164 | 0.000 | 164 |
| History: Other disease; Sleep apnea | 0.000 | 164 | 0.000 | 164 |
| History: Other disease; asthma | 0.000 | 164 | 0.000 | 164 |
| History: Other disease; liver dysfunction | 0.000 | 164 | 0.000 | 164 |
| History: Other disease; cirrhosis | 0.000 | 164 | 0.000 | 164 |
| History: Other disease; gastrointestinal ulcers | 0.000 | 164 | 0.000 | 164 |
| History: Other disease; gastrointestinal bleeding | 0.000 | 164 | 0.000 | 164 |
| History: Other disease; arthritis | 0.000 | 164 | 0.000 | 164 |
| Family History: Hypertension; | 0.000 | 164 | 0.000 | 164 |
| Family History: Hyperlipidemia; | 0.000 | 164 | 0.000 | 164 |
| Family History: Diabetes; | 0.000 | 164 | 0.000 | 164 |
| Family History: Stroke; | 0.000 | 164 | 0.000 | 164 |
| Family History: Coronary heart disease; | 0.000 | 164 | 0.000 | 164 |
| Family History: Cancer; | 0.000 | 164 | 0.000 | 164 |
| History: Migraine within 1 year | 0.000 | 164 | 0.000 | 164 |
| History: Diabetes; | 0.000 | 164 | 0.000 | 164 |
| History: Hypertension | 0.000 | 164 | 0.000 | 164 |
| History: Lipid metabolism disorders | 0.000 | 164 | 0.000 | 164 |
| History: Stroke type; Ischemic stroke | 0.000 | 164 | 0.000 | 164 |
| History: Stroke type; Intracranial hemorrhage | 0.000 | 164 | 0.000 | 164 |
| History: Stroke type; Subarachnoid hemorrhage (SAH) | 0.000 | 164 | 0.000 | 164 |
| History: Migraine; | 0.000 | 164 | 0.000 | 164 |
| History: Other disease; renal insufficiency | 0.000 | 164 | 0.000 | 164 |
| Pre-Hospital: Ambulance informed the hospital in advance | 0.000 | 164 | 0.000 | 164 |
| Admitting Diagnosis: 1-cerebral infarction; 2-transient cerebral ischemia | 0.000 | 164 | 0.000 | 164 |
| Inpatient Event: Transient ischemic attack (TIA) | 0.000 | 164 | 0.000 | 164 |
| Inpatient Event: systemic embolic events | 0.000 | 164 | 0.000 | 164 |
| Inpatient Therapy: Vascular related operation and surgical treatment | 0.000 | 164 | 0.000 | 164 |
| Inpatient Therapy: Carotid artery stenting | 0.000 | 164 | 0.000 | 164 |
| Inpatient Therapy: Carotid endarterectomy (CEA) | 0.000 | 164 | 0.000 | 164 |

| | | | | |
|---|---|---|---|---|
| Inpatient Therapy: Intracranial arterial stenting | 0.000 | 164 | 0.000 | 164 |
| Inpatient Therapy: Decompressive craniectomy | 0.000 | 164 | 0.000 | 164 |
| Inpatient Therapy: Walk within 48 hours of admission | 0.000 | 164 | 0.000 | 164 |
| Final diagnosis: cerebral infarction or transient cerebral ischemia | 0.000 | 164 | 0.000 | 164 |
| Final diagnosis: Hypertension | 0.000 | 164 | 0.000 | 164 |
| Final diagnosis: pathoglycemia | 0.000 | 164 | 0.000 | 164 |
| Final diagnosis: lipid metabolism disorder | 0.000 | 164 | 0.000 | 164 |
| Final diagnosis: coronary heart disease | 0.000 | 164 | 0.000 | 164 |
| Final diagnosis: atrial fibrillation | 0.000 | 164 | 0.000 | 164 |
| Final diagnosis: Respiratory disease | 0.000 | 164 | 0.000 | 164 |
| Final diagnosis: urinary system diseases | 0.000 | 164 | 0.000 | 164 |
| Final diagnosis: peripheral arterial disease | 0.000 | 164 | 0.000 | 164 |
| Final diagnosis: Deep venous thrombosis | 0.000 | 164 | 0.000 | 164 |
| Final diagnosis: Epilepsy | 0.000 | 164 | 0.000 | 164 |
| Discharge medication: Antiplatelet agents | 0.000 | 164 | 0.000 | 164 |
| Discharge Medication: Anticoagulant drugs | 0.000 | 164 | 0.000 | 164 |
| Discharge Medication: Hypoglycemic therapy | 0.000 | 164 | 0.000 | 164 |
| Discharge Medication: Antihypertensive therapy | 0.000 | 164 | 0.000 | 164 |
| Discharge Medication: antioxidant against lipid peroxidation | 0.000 | 164 | 0.000 | 164 |
| Antidepressant treatment in discharge description | 0.000 | 164 | 0.000 | 164 |
| Anti-insomnia treatment in discharge description | 0.000 | 164 | 0.000 | 164 |
| ICONS: Post-stroke depression | 0.000 | 164 | 0.000 | 164 |
| Endovascular Therapy: Mechanical thrombectomy | 0.000 | 164 | 0.000 | 164 |
| Endovascular Therapy: Stent Therapy | 0.000 | 164 | 0.000 | 164 |
| Inpatient Therapy: Lipid-lowering drugs | 0.000 | 164 | 0.000 | 164 |
| Inpatient Therapy: Hypoglycemic treatment | 0.000 | 164 | 0.000 | 164 |
| Inpatient Therapy: Traditional Chinese medicine therapy during hospitalization | 0.000 | 164 | 0.000 | 164 |
| Intravenous Thrombolysis: rt-PA intravenous thrombolytic | 0.000 | 164 | 0.000 | 164 |
| apolipoprotein-B | 0.000 | 164 | 0.000 | 164 |
| free fatty acid (mmol/L); | 0.000 | 164 | 0.000 | 164 |
| Insulin (pmol/L); | 0.000 | 164 | 0.000 | 164 |
| uric acid (μmol/L); | 0.000 | 164 | 0.000 | 164 |
| Cholesterol efflux capacity | 0.000 | 164 | 0.000 | 164 |
| Etiology according to TOAST system: cardiogenic embolism | 0.000 | 164 | 0.000 | 164 |

| | | | | |
|---|---|---|---|---|
| Etiology according to TOAST system: small artery occlusion | 0.000 | 164 | 0.000 | 164 |
| Etiology according to TOAST system: stroke of another determined cause | 0.000 | 164 | 0.000 | 164 |
| Etiology according to TOAST system: stroke of an undetermined cause | 0.000 | 164 | 0.000 | 164 |
| Geographic location: eastern China | 0.000 | 164 | 0.000 | 164 |
| Geographic location: Southern China | 0.000 | 164 | 0.000 | 164 |
| Geographic location: centeral China | 0.000 | 164 | 0.000 | 164 |
| Geographic location: Sourthwestern China | 0.000 | 164 | 0.000 | 164 |
| Geographic location: Northwestern China | 0.000 | 164 | 0.000 | 164 |
| History: mRS score( before onset) | 0.000 | 164 | 0.000 | 164 |
| Admitting NIHSS1a: Level of consciousness | 0.000 | 164 | 0.000 | 164 |
| Admitting NIHSS1b: Ask month and Age | 0.000 | 164 | 0.000 | 164 |
| Admitting NIHSS1c: ’Blink eyes’ & ‘Squeeze hands’ | 0.000 | 164 | 0.000 | 164 |
| Admitting NIHSS2: Horizontal extraocular movements | 0.000 | 164 | 0.000 | 164 |
| Admitting NIHSS3: Visual Field | 0.000 | 164 | 0.000 | 164 |
| Admitting NIHSS4: Facial Palsy | 0.000 | 164 | 0.000 | 164 |
| Admitting NIHSS5a: Motor Arm-Left | 0.000 | 164 | 0.000 | 164 |
| Admitting NIHSS5b: Motor Arm-Right | 0.000 | 164 | 0.000 | 164 |
| Admitting NIHSS6b: Motor Leg-Right | 0.000 | 164 | 0.000 | 164 |
| Admitting NIHSS8: Sensation | 0.000 | 164 | 0.000 | 164 |
| Admitting NIHSS10: Dysarthria | 0.000 | 164 | 0.000 | 164 |
| Admitting NIHSS11: Extinction | 0.000 | 164 | 0.000 | 164 |
| Discharge NIHSS1b: Ask month and Age | 0.000 | 164 | 0.000 | 164 |
| Discharge NIHSS2: Horizontal extraocular movements | 0.000 | 164 | 0.000 | 164 |
| Discharge NIHSS3: Visual Field | 0.000 | 164 | 0.000 | 164 |
| Discharge NIHSS4: Facial Palsy | 0.000 | 164 | 0.000 | 164 |
| Discharge NIHSS5a: Motor Arm-L | 0.000 | 164 | 0.000 | 164 |
| Discharge NIHSS6a: Motor Leg-L | 0.000 | 164 | 0.000 | 164 |
| Discharge NIHSS7: Limb Ataxia | 0.000 | 164 | 0.000 | 164 |
| Discharge NIHSS8: Sensation | 0.000 | 164 | 0.000 | 164 |
| Discharge NIHSS10: Dysarthria | 0.000 | 164 | 0.000 | 164 |
| Discharge NIHSS11: Extinction | 0.000 | 164 | 0.000 | 164 |
| NIHSS change from admission to discharge: NIHSS1a: level of consciousness | 0.000 | 164 | 0.000 | 164 |
| NIHSS change from admission to discharge: NIHSS1b: Ask month and Age | 0.000 | 164 | 0.000 | 164 |

| | | | | |
|---|---|---|---|---|
| NIHSS change from admission to discharge: NIHSS1c:'Blink eyes' & 'Squeeze hands' | 0.000 | 164 | 0.000 | 164 |
| NIHSS change from admission to discharge: NIHSS2: Horizontal extraocular movements | 0.000 | 164 | 0.000 | 164 |
| NIHSS change from admission to discharge: NIHSS3: Visual Field | 0.000 | 164 | 0.000 | 164 |
| NIHSS change from admission to discharge: NIHSS5b: Motor Arm-R | 0.000 | 164 | 0.000 | 164 |
| NIHSS change from admission to discharge: NIHSS6b: Motor Leg-R | 0.000 | 164 | 0.000 | 164 |
| NIHSS change from admission to discharge: NIHSS7: Limb Ataxia | 0.000 | 164 | 0.000 | 164 |
| NIHSS change from admission to discharge: NIHSS8: Sensation | 0.000 | 164 | 0.000 | 164 |
| NIHSS change from admission to discharge: NIHSS10: Dysarthria | 0.000 | 164 | 0.000 | 164 |
| NIHSS change from admission to discharge: NIHSS11: Extinction | 0.000 | 164 | 0.000 | 164 |

**Table S7. The AUCs, precision, recall and F1 of primary and secondary models**

| | Development cohort | | | | Validation cohort 1 | | | | Validation cohort 2 | | | |
|---|---|---|---|---|---|---|---|---|---|---|---|---|
| | AUC | precision | recall | F1 | AUC | precision | recall | F1 | AUC | precision | recall | F1 |
| 3 months | | | | | | | | | | | | |
| recurrence | 0.802 (0.743~0.861) | 0.976 | 0.933 | 0.950 | 0.743 (0.694~0.791) | 0.964 | 0.925 | 0.944 | 0.803 (0.775~0.831) | 0.963 | 0.929 | 0.946 |
| disability | 0.897 (0.869~0.925) | 0.969 | 0.836 | 0.897 | 0.905 (0.89~0.921) | 0.959 | 0.836 | 0.893 | 0.85 (0.811~0.889) | 0.99 | 0.882 | 0.933 |
| death | 0.933 (0.863~1） | 0.999 | 0.872 | 0.931 | 0.865 (0.81~0.92) | 0.994 | 0.865 | 0.925 | 0.826 (0.74~0.913) | 0.998 | 0.839 | 0.911 |
| 6 months | | | | | | | | | | | | |
| recurrence | 0.785 (0.733~0.84) | 0.970 | 0.745 | 0.843 | 0.684 (0.64~0.729) | 0.954 | 0.734 | 0.829 | / | / | / | / |
| disability | 0.89 (0.855~0.918) | 0.975 | 0.797 | 0.877 | 0.88 (0.858~0.901) | 0.96 | 0.797 | 0.877 | / | / | / | / |
| death | 0.82 (0.77~0.869) | 0.994 | 0.881 | 0.934 | 0.82 (0.77~0.869) | 0.986 | 0.865 | 0.922 | / | / | / | / |
| 1 year | | | | | | | | | | | | |
| recurrence | 0.737 (0.677~0.79) | 0.947 | 0.823 | 0.881 | 0.671 (0.631~0.709) | 0.929 | 0.830 | 0.877 | 0.732 (0.706~0.759) | 0.944 | 0.86 | 0.9 |
| disability | 0.866 (0.832~0.894) | 0.977 | 0.711 | 0.823 | 0.872 (0.855~0.89) | 0.956 | 0.841 | 0.895 | 0.769 (0.727~0.806) | 0.983 | 0.737 | 0.843 |
| death | 0.849 (0.782~0.905) | 0.990 | 0.813 | 0.893 | 0.796 (0.742~0.844) | 0.984 | 0.807 | 0.887 | 0.714 (0.651~0.78) | 0.992 | 0.789 | 0.879 |
| 2 years | | | | | | | | | | | | |
| recurrence | 0.666 (0.563~0.774) | 0.984 | 0.731 | 0.839 | 0.617 (0.557~0.68) | 0.979 | 0.781 | 0.869 | / | / | / | / |
| disability | 0.846 (0.804~0.887) | 0.965 | 0.795 | 0.872 | 0.822 (0.793~0.85) | 0.956 | 0.795 | 0.872 | / | / | / | / |
| death | 0.822 (0.755~0.877) | 0.128 | 0.655 | 0.213 | 0.754 (0.702~0.805) | 0.089 | 0.486 | 0.150 | / | / | / | / |
| 3 years | | | | | | | | | | | | |
| recurrence | 0.696 (0.579~0.806) | 0.988 | 0.744 | 0.849 | 0.546 (0.48~0.623) | 0.976 | 0.761 | 0.855 | / | / | / | / |
| disability | 0.819 (0.769~0.867) | 0.962 | 0.831 | 0.892 | 0.803 (0.773~0.831) | 0.942 | 0.831 | 0.892 | / | / | / | / |

| | | | | | | | | | | | | |
|---|---|---|---|---|---|---|---|---|---|---|---|---|
| death | 0.797 (0.704~0.87) | 0.055 | 0.826 | 0.104 | 0.73 (0.661~0.794) | 0.051 | 0.738 | 0.095 | / | / | / | / |
| 4 years | | | | | | | | | | | | |
| recurrence | 0.678 (0.515~0.817) | 0.995 | 0.711 | 0.830 | 0.523 (0.449~0.598) | 0.973 | 0.657 | 0.784 | / | / | / | / |
| disability | 0.804 (0.751~0.849) | 0.965 | 0.776 | 0.86 | 0.793 (0.762~0.823) | 0.95 | 0.776 | 0.86 | / | / | / | / |
| death | 0.815 (0.728~0.888) | 0.047 | 0.938 | 0.089 | 0.786 (0.725~0.841) | 0.055 | 0.772 | 0.102 | / | / | / | / |
| 5 years | | | | | | | | | | | | |
| recurrence | 0.704 (0.568~0.848) | 1.000 | 0.480 | 0.649 | 0.573 (0.483~0.668) | 0.988 | 0.457 | 0.625 | / | / | / | / |
| disability | 0.806 (0.756~0.854) | 0.969 | 0.734 | 0.835 | 0.775 (0.742~0.809) | 0.949 | 0.734 | 0.835 | / | / | / | / |
| death | 0.839 (0.754~0.901) | 0.061 | 0.955 | 0.115 | 0.699 (0.63~0.763) | 0.043 | 0.698 | 0.081 | / | / | / | / |

Precision is True Positive/ (True Positives +False Positives)

Recall is True Positive/ (True Positives + False Negatives)

F1 is (2*precision*Recall)/ (Precision + Recall)

**Table S8. Distribution of NIHSS at admission (row) and discharge (column) in CNSR-III**

| | 0 | 1 | 2 | 3 | 4 | 5 | 6 | 7 | 8 | 9 | 10 | 11 | 12 | 13 | 14 | 15 | 16 | 17 | 18 | 19 | 20 | 21+ |
|---|---|---|---|---|---|---|---|---|---|---|---|---|---|---|---|---|---|---|---|---|---|---|
| 0 | 1061 | 115 | 48 | 19 | 17 | 4 | 5 | 5 | 3 | 2 | 2 | 3 | 0 | 2 | 0 | 0 | 0 | 0 | 0 | 1 | 0 | 0 |
| 1 | 851 | 794 | 105 | 46 | 26 | 14 | 12 | 13 | 5 | 4 | 2 | 2 | 0 | 0 | 1 | 0 | 0 | 0 | 0 | 0 | 0 | 0 |
| 2 | 668 | 669 | 623 | 90 | 50 | 23 | 17 | 12 | 8 | 6 | 6 | 1 | 1 | 1 | 2 | 1 | 0 | 0 | 0 | 0 | 0 | 0 |
| 3 | 420 | 465 | 438 | 324 | 66 | 23 | 19 | 9 | 9 | 8 | 6 | 4 | 2 | 0 | 0 | 2 | 0 | 0 | 2 | 0 | 0 | 2 |
| 4 | 286 | 323 | 406 | 266 | 196 | 53 | 37 | 20 | 12 | 9 | 6 | 1 | 3 | 3 | 0 | 2 | 0 | 0 | 0 | 0 | 0 | 2 |
| 5 | 156 | 152 | 236 | 214 | 147 | 121 | 38 | 34 | 17 | 14 | 2 | 8 | 2 | 1 | 1 | 1 | 1 | 0 | 0 | 0 | 0 | 4 |
| 6 | 99 | 105 | 168 | 173 | 171 | 106 | 73 | 23 | 18 | 17 | 5 | 1 | 1 | 2 | 1 | 0 | 0 | 0 | 1 | 0 | 0 | 5 |
| 7 | 63 | 55 | 97 | 103 | 107 | 82 | 83 | 50 | 11 | 12 | 13 | 8 | 1 | 0 | 2 | 1 | 0 | 2 | 1 | 0 | 0 | 2 |
| 8 | 35 | 44 | 61 | 58 | 85 | 69 | 72 | 40 | 36 | 14 | 8 | 6 | 6 | 2 | 1 | 1 | 1 | 1 | 0 | 0 | 0 | 2 |
| 9 | 29 | 18 | 32 | 46 | 47 | 54 | 32 | 38 | 27 | 27 | 7 | 10 | 5 | 2 | 0 | 2 | 0 | 1 | 1 | 0 | 0 | 0 |
| 10 | 18 | 24 | 33 | 28 | 32 | 39 | 32 | 34 | 32 | 28 | 21 | 7 | 5 | 3 | 4 | 2 | 0 | 0 | 0 | 1 | 0 | 1 |
| 11 | 13 | 12 | 9 | 13 | 23 | 21 | 24 | 23 | 20 | 29 | 15 | 21 | 9 | 2 | 2 | 1 | 0 | 3 | 0 | 0 | 0 | 1 |
| 12 | 16 | 15 | 10 | 11 | 15 | 10 | 11 | 17 | 23 | 25 | 19 | 12 | 14 | 5 | 1 | 0 | 3 | 1 | 0 | 0 | 1 | 1 |
| 13 | 7 | 3 | 6 | 6 | 14 | 11 | 7 | 8 | 15 | 13 | 6 | 12 | 6 | 5 | 2 | 1 | 3 | 2 | 0 | 0 | 0 | 4 |
| 14 | 6 | 3 | 6 | 4 | 5 | 2 | 7 | 12 | 13 | 5 | 13 | 10 | 5 | 5 | 6 | 0 | 1 | 1 | 1 | 1 | 0 | 2 |
| 15 | 5 | 2 | 2 | 3 | 2 | 3 | 5 | 8 | 8 | 6 | 7 | 2 | 9 | 3 | 3 | 2 | 2 | 0 | 1 | 0 | 0 | 2 |
| 16 | 4 | 1 | 2 | 4 | 1 | 3 | 1 | 2 | 5 | 7 | 5 | 3 | 4 | 5 | 8 | 1 | 3 | 0 | 0 | 0 | 0 | 2 |
| 17 | 1 | 1 | 4 | 3 | 5 | 1 | 1 | 5 | 3 | 5 | 3 | 1 | 2 | 0 | 1 | 2 | 3 | 1 | 2 | 0 | 0 | 1 |
| 18 | 2 | 0 | 2 | 1 | 0 | 0 | 3 | 3 | 2 | 1 | 1 | 1 | 4 | 1 | 6 | 7 | 1 | 2 | 1 | 0 | 0 | 4 |
| 19 | 4 | 0 | 3 | 0 | 0 | 3 | 3 | 1 | 1 | 1 | 3 | 1 | 1 | 4 | 2 | 1 | 4 | 0 | 1 | 0 | 1 | 4 |
| 20 | 3 | 0 | 5 | 1 | 0 | 2 | 1 | 0 | 1 | 2 | 1 | 0 | 1 | 1 | 1 | 0 | 2 | 2 | 0 | 0 | 3 | 2 |

| | | | | | | | | | | | | | | | | | | | | | | |
|---|---|---|---|---|---|---|---|---|---|---|---|---|---|---|---|---|---|---|---|---|---|---|
| 21+ | 10 | 5 | 9 | 6 | 7 | 3 | 2 | 1 | 1 | 2 | 3 | 3 | 7 | 5 | 8 | 4 | 2 | 3 | 7 | 5 | 2 | 27 |

Notes:

The “21+” in the last colnames and rownames means the NIHSS is 21 or more than 21.

**Table S9a. Multi-variables regression of basic clinical variables and Delta-NIHSS$_{(admission-discharge)}$**

| | Delta-NIHSS | | | Delta-NIH1A | | | Delta-NIH1B | | | Delta-NIH1C | | |
|---|---|---|---|---|---|---|---|---|---|---|---|---|
| variables | Estimate | SE | P-value | Estimate | SE | P-value | Estimate | SE | P-value | Estimate | SE | P-value |
| (Intercept) | -1.663 | 0.632 | 0.009 | -0.163 | 0.079 | 0.040 | 0.030 | 0.089 | 0.737 | -0.120 | 0.074 | 0.108 |
| AGE | -0.011 | 0.002 | 0.000 | 0.000 | 0.000 | 0.075 | 0.000 | 0.000 | 0.388 | 0.000 | 0.000 | 0.386 |
| GENDER | -0.070 | 0.055 | 0.199 | -0.003 | 0.007 | 0.621 | -0.012 | 0.008 | 0.127 | -0.002 | 0.006 | 0.716 |
| Central China | 0.093 | 0.161 | 0.564 | 0.001 | 0.020 | 0.973 | -0.008 | 0.023 | 0.717 | -0.046 | 0.019 | 0.014 |
| Northeast China | -0.019 | 0.159 | 0.906 | 0.002 | 0.020 | 0.903 | -0.003 | 0.022 | 0.877 | -0.027 | 0.019 | 0.147 |
| Northwest china | 0.145 | 0.158 | 0.358 | 0.010 | 0.020 | 0.621 | -0.001 | 0.022 | 0.963 | -0.024 | 0.019 | 0.192 |
| East china | -0.063 | 0.166 | 0.704 | -0.001 | 0.021 | 0.964 | 0.000 | 0.023 | 0.989 | -0.022 | 0.020 | 0.251 |
| Northwest china | 0.054 | 0.173 | 0.757 | 0.022 | 0.022 | 0.313 | -0.012 | 0.024 | 0.624 | -0.038 | 0.020 | 0.059 |
| South china | -0.092 | 0.177 | 0.603 | -0.014 | 0.022 | 0.528 | -0.044 | 0.025 | 0.077 | -0.061 | 0.021 | 0.003 |
| History of TIA | 0.034 | 0.128 | 0.789 | 0.025 | 0.016 | 0.128 | 0.006 | 0.018 | 0.746 | 0.027 | 0.015 | 0.078 |
| History of Diabetes | -0.137 | 0.078 | 0.078 | -0.002 | 0.010 | 0.802 | 0.010 | 0.011 | 0.345 | -0.008 | 0.009 | 0.363 |
| History of Hypertension | -0.132 | 0.048 | 0.006 | -0.006 | 0.006 | 0.311 | -0.005 | 0.007 | 0.433 | -0.017 | 0.006 | 0.003 |
| History of Smoke | -0.037 | 0.022 | 0.091 | -0.002 | 0.003 | 0.370 | -0.002 | 0.003 | 0.493 | -0.003 | 0.003 | 0.241 |
| History of Drink | -0.060 | 0.064 | 0.348 | -0.001 | 0.008 | 0.922 | -0.003 | 0.009 | 0.743 | -0.002 | 0.008 | 0.838 |
| History of lipid metabolism disorder | 0.142 | 0.079 | 0.071 | 0.024 | 0.010 | 0.013 | 0.017 | 0.011 | 0.118 | 0.012 | 0.009 | 0.215 |
| History of Atrial Fibrillation | -0.193 | 0.113 | 0.088 | 0.045 | 0.014 | 0.001 | 0.042 | 0.016 | 0.008 | 0.024 | 0.013 | 0.077 |
| NIHSS at administration | 0.459 | 0.005 | 0.000 | 0.027 | 0.001 | 0.000 | 0.030 | 0.001 | 0.000 | 0.027 | 0.001 | 0.000 |
| History of Coronary Heart Disease | 0.020 | 0.070 | 0.773 | -0.002 | 0.009 | 0.841 | -0.001 | 0.010 | 0.883 | 0.002 | 0.008 | 0.822 |
| large artery atherosclerosis | -0.051 | 0.196 | 0.794 | 0.021 | 0.025 | 0.403 | 0.011 | 0.028 | 0.688 | 0.007 | 0.023 | 0.768 |

| | | | | | | | | | | | | |
|---|---|---|---|---|---|---|---|---|---|---|---|---|
| cardiogenic embolism | 0.849 | 0.222 | 0.000 | 0.010 | 0.028 | 0.714 | 0.051 | 0.031 | 0.099 | 0.018 | 0.026 | 0.484 |
| Small artery occlusion | 0.448 | 0.197 | 0.023 | 0.021 | 0.025 | 0.395 | 0.004 | 0.028 | 0.883 | 0.002 | 0.023 | 0.938 |
| stroke of an undetermined cause | 0.347 | 0.194 | 0.074 | 0.027 | 0.024 | 0.260 | 0.027 | 0.027 | 0.320 | 0.015 | 0.023 | 0.511 |
| Anticoagulation treatment | -0.413 | 0.074 | 0.000 | 0.005 | 0.009 | 0.573 | -0.015 | 0.010 | 0.149 | -0.017 | 0.009 | 0.047 |
| Lipid-lowering drugs | 0.098 | 0.114 | 0.388 | -0.011 | 0.014 | 0.430 | -0.015 | 0.016 | 0.351 | -0.041 | 0.013 | 0.002 |
| Antihypertensive treatment | 0.028 | 0.046 | 0.553 | 0.003 | 0.006 | 0.588 | 0.003 | 0.007 | 0.700 | 0.010 | 0.005 | 0.078 |
| Anti-platelet therapy | 0.228 | 0.131 | 0.082 | -0.021 | 0.016 | 0.197 | 0.003 | 0.018 | 0.862 | -0.008 | 0.015 | 0.588 |
| Hypoglycemic treatment | -0.040 | 0.075 | 0.596 | -0.005 | 0.009 | 0.597 | -0.028 | 0.011 | 0.007 | -0.008 | 0.009 | 0.394 |
| rt-PA intravenous thrombolytic | 1.094 | 0.078 | 0.000 | 0.020 | 0.010 | 0.037 | 0.045 | 0.011 | 0.000 | 0.024 | 0.009 | 0.008 |
| Mechanical thrombectomy | 0.634 | 0.416 | 0.128 | 0.144 | 0.052 | 0.006 | -0.059 | 0.058 | 0.316 | 0.184 | 0.049 | 0.000 |

**Table S9b. Multi-variables regression of basic clinical variables and Delta-NIHSS(admission-discharge)**

| | Delta-NIH2 | | | Delta-NIH3 | | | Delta-NIH4 | | | Delta-NIH5A | | |
|---|---|---|---|---|---|---|---|---|---|---|---|---|
| variables | Estimate | SE | P-value | Estimate | SE | P-value | Estimate | SE | P-value | Estimate | SE | P-value |
| (Intercept) | -0.112 | 0.060 | 0.061 | 0.081 | 0.060 | 0.177 | 0.062 | 0.140 | 0.656 | 0.196 | 0.209 | 0.348 |
| AGE | 0.000 | 0.000 | 0.453 | 0.000 | 0.000 | 0.931 | -0.002 | 0.000 | 0.000 | -0.002 | 0.001 | 0.005 |
| GENDER | -0.009 | 0.005 | 0.084 | -0.004 | 0.005 | 0.458 | 0.005 | 0.012 | 0.670 | -0.002 | 0.018 | 0.909 |
| Central China | 0.021 | 0.015 | 0.173 | 0.010 | 0.015 | 0.517 | -0.083 | 0.036 | 0.019 | 0.100 | 0.053 | 0.059 |
| Northeast China | 0.014 | 0.015 | 0.338 | 0.013 | 0.015 | 0.383 | -0.101 | 0.035 | 0.004 | 0.086 | 0.053 | 0.104 |
| Northwest china | 0.021 | 0.015 | 0.160 | 0.007 | 0.015 | 0.665 | -0.120 | 0.035 | 0.001 | 0.102 | 0.052 | 0.052 |
| East china | 0.017 | 0.016 | 0.278 | 0.011 | 0.016 | 0.482 | -0.159 | 0.037 | 0.000 | 0.055 | 0.055 | 0.319 |
| Northwest china | 0.012 | 0.016 | 0.454 | 0.008 | 0.016 | 0.628 | -0.149 | 0.038 | 0.000 | 0.108 | 0.057 | 0.060 |
| South china | -0.012 | 0.017 | 0.471 | 0.002 | 0.017 | 0.928 | -0.094 | 0.039 | 0.016 | 0.152 | 0.059 | 0.010 |
| History of TIA | 0.005 | 0.012 | 0.665 | 0.010 | 0.012 | 0.437 | -0.001 | 0.028 | 0.965 | -0.002 | 0.043 | 0.968 |
| History of Diabetes | -0.003 | 0.007 | 0.644 | 0.001 | 0.007 | 0.941 | -0.028 | 0.017 | 0.104 | 0.010 | 0.026 | 0.692 |
| History of Hypertension | -0.005 | 0.005 | 0.245 | -0.003 | 0.005 | 0.578 | -0.008 | 0.011 | 0.445 | -0.015 | 0.016 | 0.338 |
| History of Smoke | -0.001 | 0.002 | 0.635 | 0.001 | 0.002 | 0.503 | 0.000 | 0.005 | 0.990 | -0.010 | 0.007 | 0.168 |
| History of Drink | -0.009 | 0.006 | 0.126 | -0.003 | 0.006 | 0.679 | 0.014 | 0.014 | 0.334 | -0.003 | 0.021 | 0.904 |
| History of lipid metabolism disorder | 0.015 | 0.007 | 0.050 | 0.005 | 0.008 | 0.475 | 0.002 | 0.017 | 0.888 | -0.011 | 0.026 | 0.665 |
| History of Atrial Fibrillation | 0.037 | 0.011 | 0.000 | 0.015 | 0.011 | 0.163 | 0.001 | 0.025 | 0.957 | -0.016 | 0.037 | 0.672 |
| NIHSS at administration | 0.017 | 0.001 | 0.000 | 0.007 | 0.001 | 0.000 | 0.029 | 0.001 | 0.000 | 0.051 | 0.002 | 0.000 |
| History of Coronary Heart Disease | -0.002 | 0.007 | 0.709 | -0.003 | 0.007 | 0.664 | 0.016 | 0.016 | 0.296 | 0.046 | 0.023 | 0.047 |
| large artery atherosclerosis | 0.003 | 0.019 | 0.862 | 0.015 | 0.019 | 0.415 | -0.030 | 0.043 | 0.492 | 0.014 | 0.065 | 0.832 |
| cardiogenic embolism | 0.010 | 0.021 | 0.637 | 0.016 | 0.021 | 0.461 | 0.040 | 0.049 | 0.419 | 0.204 | 0.073 | 0.006 |

| | | | | | | | | | | | | |
|---|---|---|---|---|---|---|---|---|---|---|---|---|
| Small artery occlusion | -0.006 | 0.019 | 0.762 | 0.013 | 0.019 | 0.494 | 0.027 | 0.044 | 0.539 | 0.087 | 0.065 | 0.180 |
| stroke of an undetermined cause | -0.006 | 0.018 | 0.750 | 0.015 | 0.019 | 0.425 | 0.000 | 0.043 | 0.993 | 0.074 | 0.064 | 0.248 |
| Anticoagulation treatment | -0.007 | 0.007 | 0.291 | -0.002 | 0.007 | 0.764 | -0.009 | 0.016 | 0.585 | -0.087 | 0.025 | 0.000 |
| Lipid-lowering drugs | -0.016 | 0.011 | 0.143 | 0.002 | 0.011 | 0.873 | 0.018 | 0.025 | 0.467 | 0.058 | 0.038 | 0.122 |
| Antihypertensive treatment | -0.005 | 0.004 | 0.285 | -0.001 | 0.004 | 0.811 | -0.013 | 0.010 | 0.190 | 0.024 | 0.015 | 0.113 |
| Anti-platelet therapy | -0.009 | 0.012 | 0.490 | -0.003 | 0.012 | 0.833 | 0.070 | 0.029 | 0.015 | 0.001 | 0.043 | 0.977 |
| Hypoglycemic treatment | -0.004 | 0.007 | 0.603 | 0.003 | 0.007 | 0.698 | 0.016 | 0.017 | 0.338 | -0.021 | 0.025 | 0.391 |
| rt-PA intravenous thrombolytic | 0.056 | 0.007 | 0.000 | -0.005 | 0.007 | 0.495 | 0.141 | 0.017 | 0.000 | 0.148 | 0.026 | 0.000 |
| Mechanical thrombectomy | 0.092 | 0.039 | 0.020 | -0.109 | 0.040 | 0.006 | -0.108 | 0.092 | 0.240 | -0.444 | 0.138 | 0.001 |

**Table S9c. Multi -variables regression of basic clinical variables and delta-NIHSS$_{(admission-discharge)}$**

| | Delta-NIH5B | | | Delta-NIH6A | | | Delta-NIH6B | | | Delta-NIH7 | | |
|---|---|---|---|---|---|---|---|---|---|---|---|---|
| variables | Estimate | SE | P-value | Estimate | SE | P-value | Estimate | SE | P-value | Estimate | SE | P-value |
| (Intercept) | 0.100 | 0.206 | 0.626 | 0.047 | 0.193 | 0.809 | 0.071 | 0.189 | 0.708 | -0.217 | 0.112 | 0.053 |
| AGE | -0.001 | 0.001 | 0.063 | -0.002 | 0.001 | 0.000 | -0.003 | 0.001 | 0.000 | 0.000 | 0.000 | 0.468 |
| GENDER | -0.032 | 0.018 | 0.075 | -0.037 | 0.017 | 0.028 | -0.026 | 0.016 | 0.110 | 0.002 | 0.010 | 0.806 |
| Central China | -0.025 | 0.052 | 0.630 | -0.064 | 0.049 | 0.193 | 0.009 | 0.048 | 0.856 | -0.069 | 0.029 | 0.016 |
| Northeast China | 0.000 | 0.052 | 0.994 | -0.026 | 0.049 | 0.599 | 0.024 | 0.048 | 0.621 | -0.040 | 0.028 | 0.161 |
| Northwest china | 0.043 | 0.051 | 0.403 | -0.013 | 0.048 | 0.785 | 0.051 | 0.047 | 0.278 | -0.058 | 0.028 | 0.039 |
| East china | -0.040 | 0.054 | 0.457 | -0.043 | 0.051 | 0.400 | -0.033 | 0.050 | 0.510 | -0.083 | 0.029 | 0.005 |
| Northwest china | -0.056 | 0.056 | 0.321 | -0.032 | 0.053 | 0.549 | -0.006 | 0.052 | 0.901 | -0.179 | 0.031 | 0.000 |
| South china | -0.045 | 0.058 | 0.436 | -0.112 | 0.054 | 0.038 | -0.035 | 0.053 | 0.505 | -0.054 | 0.031 | 0.086 |
| History of TIA | -0.026 | 0.042 | 0.532 | 0.045 | 0.039 | 0.247 | -0.023 | 0.038 | 0.557 | 0.009 | 0.023 | 0.709 |
| History of Diabetes | -0.011 | 0.025 | 0.662 | -0.016 | 0.024 | 0.492 | -0.051 | 0.023 | 0.029 | 0.003 | 0.014 | 0.854 |
| History of Hypertension | -0.013 | 0.016 | 0.400 | -0.028 | 0.015 | 0.060 | 0.002 | 0.014 | 0.871 | -0.018 | 0.009 | 0.033 |
| History of Smoke | 0.008 | 0.007 | 0.252 | -0.014 | 0.007 | 0.036 | -0.004 | 0.007 | 0.525 | -0.006 | 0.004 | 0.102 |
| History of Drink | -0.049 | 0.021 | 0.018 | 0.006 | 0.020 | 0.769 | -0.020 | 0.019 | 0.292 | -0.003 | 0.011 | 0.813 |
| History of lipid metabolism disorder | 0.014 | 0.026 | 0.576 | 0.031 | 0.024 | 0.200 | 0.015 | 0.024 | 0.521 | 0.000 | 0.014 | 0.993 |
| History of Atrial Fibrillation | 0.151 | 0.037 | 0.000 | -0.020 | 0.034 | 0.565 | 0.089 | 0.034 | 0.009 | 0.131 | 0.020 | 0.000 |
| NIHSS at administration | -0.057 | 0.002 | 0.000 | -0.020 | 0.002 | 0.000 | -0.050 | 0.002 | 0.000 | 0.004 | 0.001 | 0.000 |
| History of Coronary Heart Disease | 0.025 | 0.023 | 0.280 | -0.028 | 0.021 | 0.196 | 0.032 | 0.021 | 0.125 | -0.006 | 0.012 | 0.640 |
| large artery atherosclerosis | -0.071 | 0.064 | 0.263 | 0.040 | 0.060 | 0.505 | 0.051 | 0.059 | 0.384 | -0.021 | 0.035 | 0.549 |
| cardiogenic embolism | 0.014 | 0.072 | 0.843 | 0.193 | 0.068 | 0.004 | 0.121 | 0.066 | 0.068 | -0.052 | 0.039 | 0.185 |

| | | | | | | | | | | | | |
|---|---|---|---|---|---|---|---|---|---|---|---|---|
| Small artery occlusion | -0.002 | 0.064 | 0.974 | 0.111 | 0.060 | 0.064 | 0.107 | 0.059 | 0.070 | -0.037 | 0.035 | 0.292 |
| stroke of an undetermined cause | -0.036 | 0.063 | 0.571 | 0.091 | 0.059 | 0.125 | 0.087 | 0.058 | 0.132 | -0.024 | 0.034 | 0.492 |
| Anticoagulation treatment | -0.104 | 0.024 | 0.000 | -0.092 | 0.023 | 0.000 | -0.059 | 0.022 | 0.008 | -0.055 | 0.013 | 0.000 |
| Lipid-lowering drugs | -0.005 | 0.037 | 0.898 | 0.013 | 0.035 | 0.718 | 0.001 | 0.034 | 0.984 | -0.042 | 0.020 | 0.036 |
| Antihypertensive treatment | -0.037 | 0.015 | 0.013 | 0.008 | 0.014 | 0.579 | -0.034 | 0.014 | 0.015 | -0.001 | 0.008 | 0.933 |
| Anti-platelet therapy | 0.013 | 0.043 | 0.758 | 0.002 | 0.040 | 0.964 | -0.036 | 0.039 | 0.364 | 0.013 | 0.023 | 0.579 |
| Hypoglycemic treatment | 0.022 | 0.024 | 0.374 | -0.037 | 0.023 | 0.107 | 0.026 | 0.022 | 0.254 | -0.044 | 0.013 | 0.001 |
| rt-PA intravenous thrombolytic | 0.169 | 0.025 | 0.000 | 0.121 | 0.024 | 0.000 | 0.172 | 0.023 | 0.000 | 0.120 | 0.014 | 0.000 |
| Mechanical thrombectomy | -0.037 | 0.135 | 0.783 | -0.006 | 0.127 | 0.965 | -0.025 | 0.125 | 0.844 | 0.296 | 0.074 | 0.000 |

**Table S9d. Multi-variables regression of basic clinical variables and Delta-NIHSS(admission-discharge)**

| | Delta-NIH8 | | | Delta-NIH9 | | | Delta-NIH10 | | | Delta-NIH11 | | |
|---|---|---|---|---|---|---|---|---|---|---|---|---|
| variables | Estimate | SE | P-value | Estimate | SE | P-value | Estimate | SE | P-value | Estimate | SE | P-value |
| (Intercept) | -0.202 | 0.109 | 0.064 | -0.307 | 0.128 | 0.016 | 0.257 | 0.172 | 0.134 | -0.074 | 0.062 | 0.231 |
| AGE | 0.001 | 0.000 | 0.051 | -0.001 | 0.000 | 0.045 | -0.001 | 0.001 | 0.055 | 0.000 | 0.000 | 0.680 |
| GENDER | -0.028 | 0.009 | 0.003 | 0.008 | 0.011 | 0.458 | 0.032 | 0.015 | 0.030 | -0.013 | 0.005 | 0.015 |
| Central China | 0.142 | 0.028 | 0.000 | 0.007 | 0.032 | 0.826 | 0.054 | 0.044 | 0.217 | 0.024 | 0.016 | 0.127 |
| Northeast China | 0.137 | 0.027 | 0.000 | 0.005 | 0.032 | 0.875 | 0.033 | 0.043 | 0.452 | 0.025 | 0.016 | 0.110 |
| Northwest china | 0.140 | 0.027 | 0.000 | 0.008 | 0.032 | 0.810 | 0.029 | 0.043 | 0.499 | 0.031 | 0.015 | 0.043 |
| East china | 0.134 | 0.029 | 0.000 | -0.071 | 0.034 | 0.036 | -0.008 | 0.045 | 0.854 | 0.023 | 0.016 | 0.161 |
| Northwest china | 0.087 | 0.030 | 0.003 | -0.040 | 0.035 | 0.255 | 0.157 | 0.047 | 0.001 | 0.016 | 0.017 | 0.348 |
| South china | 0.077 | 0.030 | 0.012 | -0.051 | 0.036 | 0.158 | -0.026 | 0.048 | 0.592 | -0.012 | 0.017 | 0.485 |
| History of TIA | 0.023 | 0.022 | 0.297 | 0.018 | 0.026 | 0.495 | 0.002 | 0.035 | 0.959 | -0.001 | 0.013 | 0.946 |
| History of Diabetes | -0.023 | 0.013 | 0.079 | -0.030 | 0.016 | 0.057 | -0.014 | 0.021 | 0.516 | -0.002 | 0.008 | 0.792 |
| History of Hypertension | -0.006 | 0.008 | 0.439 | -0.008 | 0.010 | 0.419 | -0.021 | 0.013 | 0.109 | 0.000 | 0.005 | 0.929 |
| History of Smoke | -0.008 | 0.004 | 0.041 | 0.000 | 0.004 | 0.974 | -0.012 | 0.006 | 0.051 | 0.000 | 0.002 | 0.853 |
| History of Drink | -0.014 | 0.011 | 0.212 | -0.006 | 0.013 | 0.624 | -0.008 | 0.017 | 0.648 | -0.011 | 0.006 | 0.073 |
| History of lipid metabolism disorder | 0.016 | 0.014 | 0.228 | 0.015 | 0.016 | 0.338 | 0.017 | 0.021 | 0.417 | 0.010 | 0.008 | 0.206 |
| History of Atrial Fibrillation | 0.061 | 0.019 | 0.002 | 0.022 | 0.023 | 0.338 | 0.001 | 0.031 | 0.971 | 0.067 | 0.011 | 0.000 |
| NIHSS at administration | 0.004 | 0.001 | 0.000 | -0.027 | 0.001 | 0.000 | -0.028 | 0.001 | 0.000 | 0.021 | 0.001 | 0.000 |
| History of Coronary Heart Disease | 0.010 | 0.012 | 0.425 | 0.003 | 0.014 | 0.808 | -0.018 | 0.019 | 0.345 | 0.002 | 0.007 | 0.754 |
| large artery atherosclerosis | -0.011 | 0.034 | 0.747 | -0.082 | 0.040 | 0.038 | -0.014 | 0.053 | 0.796 | -0.024 | 0.019 | 0.214 |
| cardiogenic embolism | 0.025 | 0.038 | 0.514 | -0.032 | 0.045 | 0.481 | 0.036 | 0.060 | 0.548 | -0.044 | 0.022 | 0.043 |

| | | | | | | | | | | | | |
|---|---|---|---|---|---|---|---|---|---|---|---|---|
| Small artery occlusion | -0.060 | 0.034 | 0.077 | -0.016 | 0.040 | 0.685 | -0.024 | 0.054 | 0.655 | -0.036 | 0.019 | 0.058 |
| stroke of an undetermined cause | 0.002 | 0.033 | 0.952 | -0.025 | 0.039 | 0.519 | -0.012 | 0.053 | 0.815 | -0.032 | 0.019 | 0.087 |
| Anticoagulation treatment | -0.003 | 0.013 | 0.836 | -0.001 | 0.015 | 0.950 | -0.031 | 0.020 | 0.127 | -0.010 | 0.007 | 0.153 |
| Lipid-lowering drugs | -0.078 | 0.020 | 0.000 | -0.003 | 0.023 | 0.903 | -0.095 | 0.031 | 0.002 | -0.016 | 0.011 | 0.163 |
| Antihypertensive treatment | -0.003 | 0.008 | 0.672 | -0.007 | 0.009 | 0.452 | -0.017 | 0.013 | 0.175 | -0.005 | 0.005 | 0.233 |
| Anti-platelet therapy | -0.048 | 0.023 | 0.034 | -0.003 | 0.026 | 0.903 | 0.014 | 0.036 | 0.701 | -0.024 | 0.013 | 0.056 |
| Hypoglycemic treatment | -0.010 | 0.013 | 0.439 | 0.042 | 0.015 | 0.005 | -0.012 | 0.020 | 0.548 | -0.004 | 0.007 | 0.610 |
| rt-PA intravenous thrombolytic | 0.084 | 0.013 | 0.000 | 0.134 | 0.016 | 0.000 | 0.153 | 0.021 | 0.000 | 0.044 | 0.008 | 0.000 |
| Mechanical thrombectomy | 0.148 | 0.072 | 0.039 | 0.178 | 0.084 | 0.035 | -0.253 | 0.113 | 0.025 | 0.113 | 0.041 | 0.006 |

**Table S10a. Multi-variables regression of basic clinical variables and Delta-NIHSS$_{(admission-discharge)}$ in scenario 1**

| | Delta-NIHSS | | | Delta-NIH1A | | | Delta-NIH1B | | | Delta-NIH1C | | |
|---|---|---|---|---|---|---|---|---|---|---|---|---|
| variables | Estimate | SE | P-value | Estimate | SE | P-value | Estimate | SE | P-value | Estimate | SE | P-value |
| (Intercept) | -0.379 | 0.894 | 0.671 | -0.164 | 0.112 | 0.144 | -0.213 | 0.124 | 0.085 | -0.342 | 0.103 | 0.001 |
| AGE | -0.009 | 0.002 | 0.000 | 0.000 | 0.000 | 0.218 | 0.000 | 0.000 | 0.967 | 0.000 | 0.000 | 0.175 |
| GENDER | 0.000 | 0.067 | 0.997 | 0.002 | 0.008 | 0.780 | -0.004 | 0.009 | 0.658 | 0.003 | 0.008 | 0.695 |
| Central China | 0.067 | 0.211 | 0.753 | -0.005 | 0.026 | 0.841 | -0.018 | 0.029 | 0.547 | -0.061 | 0.024 | 0.013 |
| Northeast China | -0.017 | 0.210 | 0.935 | -0.020 | 0.026 | 0.437 | -0.017 | 0.029 | 0.562 | -0.054 | 0.024 | 0.026 |
| Northwest china | 0.024 | 0.209 | 0.908 | -0.024 | 0.026 | 0.353 | -0.021 | 0.029 | 0.470 | -0.054 | 0.024 | 0.026 |
| East china | -0.304 | 0.217 | 0.160 | -0.024 | 0.027 | 0.377 | -0.020 | 0.030 | 0.503 | -0.052 | 0.025 | 0.037 |
| West china | -0.163 | 0.227 | 0.473 | -0.015 | 0.028 | 0.593 | -0.027 | 0.031 | 0.393 | -0.065 | 0.026 | 0.013 |
| South china | -0.027 | 0.231 | 0.909 | -0.041 | 0.029 | 0.157 | -0.042 | 0.032 | 0.188 | -0.075 | 0.027 | 0.005 |
| History of TIA | 0.098 | 0.160 | 0.541 | 0.037 | 0.020 | 0.067 | 0.030 | 0.022 | 0.177 | 0.027 | 0.019 | 0.139 |
| History of Diabetes | -0.127 | 0.097 | 0.191 | -0.009 | 0.012 | 0.461 | 0.014 | 0.014 | 0.306 | -0.013 | 0.011 | 0.262 |
| History of Hypertension | -0.106 | 0.059 | 0.073 | 0.009 | 0.007 | 0.236 | -0.005 | 0.008 | 0.565 | -0.008 | 0.007 | 0.237 |
| History of Smoke | -0.021 | 0.027 | 0.438 | -0.003 | 0.003 | 0.435 | 0.001 | 0.004 | 0.790 | -0.002 | 0.003 | 0.436 |
| History of Drink | 0.019 | 0.079 | 0.808 | 0.002 | 0.010 | 0.838 | 0.006 | 0.011 | 0.603 | 0.003 | 0.009 | 0.717 |
| History of lipid metabolism disorder | 0.279 | 0.100 | 0.005 | 0.034 | 0.013 | 0.008 | 0.016 | 0.014 | 0.236 | 0.011 | 0.012 | 0.322 |
| History of Atrial Fibrillation | -0.364 | 0.146 | 0.012 | -0.009 | 0.018 | 0.608 | 0.022 | 0.020 | 0.283 | 0.000 | 0.017 | 0.992 |
| NIHSS at administration | 0.474 | 0.007 | 0.000 | 0.023 | 0.001 | 0.000 | 0.029 | 0.001 | 0.000 | 0.025 | 0.001 | 0.000 |
| History of Coronary Heart Disease | -0.057 | 0.088 | 0.516 | -0.022 | 0.011 | 0.047 | 0.002 | 0.012 | 0.851 | 0.003 | 0.010 | 0.769 |
| large artery atherosclerosis | -0.125 | 0.267 | 0.639 | 0.021 | 0.034 | 0.536 | 0.013 | 0.037 | 0.722 | 0.006 | 0.031 | 0.833 |
| cardiogenic embolism | 0.658 | 0.296 | 0.026 | 0.021 | 0.037 | 0.565 | 0.073 | 0.041 | 0.075 | 0.016 | 0.034 | 0.632 |

| | | | | | | | | | | | | |
|---|---|---|---|---|---|---|---|---|---|---|---|---|
| Small artery occlusion | 0.220 | 0.267 | 0.410 | 0.024 | 0.034 | 0.480 | 0.007 | 0.037 | 0.854 | 0.002 | 0.031 | 0.957 |
| stroke of an undetermined cause | 0.160 | 0.264 | 0.544 | 0.034 | 0.033 | 0.303 | 0.033 | 0.037 | 0.370 | 0.018 | 0.030 | 0.546 |
| Anticoagulation treatment | -0.159 | 0.097 | 0.103 | 0.021 | 0.012 | 0.079 | -0.008 | 0.014 | 0.559 | -0.007 | 0.011 | 0.519 |
| Lipid-lowering drugs | 0.113 | 0.136 | 0.407 | 0.008 | 0.017 | 0.621 | -0.003 | 0.019 | 0.887 | -0.005 | 0.016 | 0.739 |
| Antihypertensive treatment | 0.009 | 0.057 | 0.881 | -0.005 | 0.007 | 0.530 | -0.002 | 0.008 | 0.816 | 0.001 | 0.007 | 0.898 |
| Anti-platelet therapy | 0.288 | 0.165 | 0.080 | -0.013 | 0.021 | 0.530 | 0.008 | 0.023 | 0.731 | -0.002 | 0.019 | 0.934 |
| Hypoglycemic treatment | -0.077 | 0.094 | 0.415 | 0.000 | 0.012 | 0.988 | -0.028 | 0.013 | 0.031 | -0.007 | 0.011 | 0.518 |
| rt-PA intravenous thrombolytic | 1.102 | 0.089 | 0.000 | 0.025 | 0.011 | 0.027 | 0.033 | 0.012 | 0.007 | 0.015 | 0.010 | 0.142 |
| Mechanical thrombectomy | -0.975 | 0.664 | 0.142 | 0.089 | 0.083 | 0.282 | 0.148 | 0.092 | 0.108 | 0.364 | 0.077 | 0.000 |

**Table S10b. Multi-variables regression of basic clinical variables and Delta-NIHSS$_{(admission-discharge)}$ in scenario 1**

| | Delta-NIH2 | | | Delta-NIH3 | | | Delta-NIH4 | | | Delta-NIH5A | | |
|---|---|---|---|---|---|---|---|---|---|---|---|---|
| variables | Estimate | SE | P-value | Estimate | SE | P-value | Estimate | SE | P-value | Estimate | SE | P-value |
| (Intercept) | 0.115 | 0.082 | 0.160 | 0.263 | 0.084 | 0.002 | 0.335 | 0.200 | 0.093 | 0.862 | 0.295 | 0.004 |
| AGE | 0.000 | 0.000 | 0.660 | 0.000 | 0.000 | 0.941 | -0.001 | 0.001 | 0.051 | -0.002 | 0.001 | 0.011 |
| GENDER | -0.006 | 0.006 | 0.331 | -0.013 | 0.006 | 0.036 | 0.003 | 0.015 | 0.842 | 0.014 | 0.022 | 0.526 |
| Central China | 0.007 | 0.019 | 0.727 | 0.038 | 0.020 | 0.057 | -0.085 | 0.047 | 0.072 | 0.025 | 0.070 | 0.726 |
| Northeast China | 0.006 | 0.019 | 0.773 | 0.046 | 0.020 | 0.021 | -0.104 | 0.047 | 0.027 | 0.006 | 0.070 | 0.929 |
| Northwest china | 0.006 | 0.019 | 0.768 | 0.038 | 0.020 | 0.051 | -0.129 | 0.047 | 0.006 | 0.004 | 0.069 | 0.952 |
| East china | 0.001 | 0.020 | 0.968 | 0.038 | 0.020 | 0.063 | -0.180 | 0.048 | 0.000 | -0.066 | 0.072 | 0.354 |
| West china | 0.001 | 0.021 | 0.969 | 0.047 | 0.021 | 0.027 | -0.194 | 0.051 | 0.000 | 0.019 | 0.075 | 0.805 |
| South china | -0.019 | 0.021 | 0.374 | 0.035 | 0.022 | 0.107 | -0.085 | 0.052 | 0.102 | 0.091 | 0.076 | 0.236 |
| History of TIA | 0.004 | 0.015 | 0.796 | 0.014 | 0.015 | 0.361 | -0.007 | 0.036 | 0.836 | -0.064 | 0.053 | 0.225 |
| History of Diabetes | 0.000 | 0.009 | 0.977 | 0.000 | 0.009 | 0.968 | 0.006 | 0.022 | 0.771 | -0.013 | 0.032 | 0.695 |
| History of Hypertension | -0.003 | 0.005 | 0.599 | -0.006 | 0.006 | 0.279 | -0.016 | 0.013 | 0.236 | -0.025 | 0.019 | 0.201 |
| History of Smoke | -0.001 | 0.002 | 0.818 | -0.002 | 0.003 | 0.460 | 0.003 | 0.006 | 0.584 | -0.014 | 0.009 | 0.112 |
| History of Drink | 0.001 | 0.007 | 0.921 | -0.001 | 0.007 | 0.906 | 0.029 | 0.018 | 0.096 | -0.001 | 0.026 | 0.962 |
| History of lipid metabolism disorder | 0.017 | 0.009 | 0.058 | 0.002 | 0.009 | 0.851 | 0.020 | 0.022 | 0.366 | 0.008 | 0.033 | 0.802 |
| History of Atrial Fibrillation | 0.013 | 0.013 | 0.346 | 0.024 | 0.014 | 0.085 | 0.020 | 0.033 | 0.549 | -0.118 | 0.048 | 0.014 |
| NIHSS at administration | 0.014 | 0.001 | 0.000 | 0.007 | 0.001 | 0.000 | 0.032 | 0.002 | 0.000 | 0.052 | 0.002 | 0.000 |
| History of Coronary Heart Disease | -0.007 | 0.008 | 0.388 | 0.007 | 0.008 | 0.414 | 0.016 | 0.020 | 0.415 | 0.007 | 0.029 | 0.819 |
| large artery atherosclerosis | -0.005 | 0.025 | 0.835 | 0.012 | 0.025 | 0.638 | -0.085 | 0.060 | 0.154 | -0.034 | 0.088 | 0.698 |
| cardiogenic embolism | 0.011 | 0.027 | 0.676 | 0.003 | 0.028 | 0.911 | -0.009 | 0.066 | 0.896 | 0.195 | 0.098 | 0.047 |

| | | | | | | | | | | | | |
|---|---|---|---|---|---|---|---|---|---|---|---|---|
| Small artery occlusion | -0.016 | 0.025 | 0.526 | 0.009 | 0.025 | 0.733 | -0.029 | 0.060 | 0.625 | -0.007 | 0.088 | 0.938 |
| stroke of an undetermined cause | -0.013 | 0.024 | 0.584 | 0.012 | 0.025 | 0.614 | -0.041 | 0.059 | 0.483 | -0.002 | 0.087 | 0.982 |
| Anticoagulation treatment | -0.012 | 0.009 | 0.191 | -0.003 | 0.009 | 0.727 | -0.006 | 0.022 | 0.796 | -0.035 | 0.032 | 0.277 |
| Lipid-lowering drugs | -0.002 | 0.013 | 0.898 | 0.002 | 0.013 | 0.902 | -0.004 | 0.031 | 0.884 | 0.037 | 0.045 | 0.408 |
| Antihypertensive treatment | -0.008 | 0.005 | 0.114 | 0.005 | 0.005 | 0.396 | -0.016 | 0.013 | 0.224 | 0.042 | 0.019 | 0.027 |
| Anti-platelet therapy | 0.001 | 0.015 | 0.967 | -0.012 | 0.015 | 0.453 | 0.079 | 0.037 | 0.032 | 0.017 | 0.054 | 0.757 |
| Hypoglycemic treatment | -0.002 | 0.009 | 0.862 | 0.001 | 0.009 | 0.903 | -0.016 | 0.021 | 0.461 | -0.005 | 0.031 | 0.882 |
| rt-PA intravenous thrombolytic | 0.054 | 0.008 | 0.000 | -0.007 | 0.008 | 0.413 | 0.150 | 0.020 | 0.000 | 0.142 | 0.030 | 0.000 |
| Mechanical thrombectomy | -0.152 | 0.061 | 0.012 | -0.283 | 0.062 | 0.000 | -0.319 | 0.148 | 0.032 | -1.006 | 0.219 | 0.000 |

**Table S10c. Multi-variables regression of basic clinical variables and Delta-NIHSS$_{(admission-discharge)}$ in scenario 1**

| | Delta-NIH5B | | | Delta-NIH6A | | | Delta-NIH6B | | | Delta-NIH7 | | |
|---|---|---|---|---|---|---|---|---|---|---|---|---|
| variables | Estimate | SE | P-value | Estimate | SE | P-value | Estimate | SE | P-value | Estimate | SE | P-value |
| (Intercept) | 0.422 | 0.278 | 0.129 | -0.017 | 0.258 | 0.949 | 0.247 | 0.264 | 0.349 | 0.094 | 0.156 | 0.547 |
| AGE | -0.001 | 0.001 | 0.375 | -0.003 | 0.001 | 0.000 | -0.002 | 0.001 | 0.011 | 0.000 | 0.000 | 0.512 |
| GENDER | -0.005 | 0.021 | 0.826 | -0.023 | 0.019 | 0.234 | 0.001 | 0.020 | 0.977 | 0.004 | 0.012 | 0.741 |
| Central China | -0.039 | 0.066 | 0.554 | -0.041 | 0.061 | 0.498 | -0.011 | 0.062 | 0.855 | -0.039 | 0.037 | 0.288 |
| Northeast China | -0.015 | 0.066 | 0.825 | 0.002 | 0.061 | 0.976 | 0.003 | 0.062 | 0.966 | -0.008 | 0.037 | 0.825 |
| Northwest china | 0.016 | 0.065 | 0.810 | 0.000 | 0.060 | 0.997 | 0.019 | 0.062 | 0.763 | -0.039 | 0.036 | 0.279 |
| East china | -0.100 | 0.068 | 0.139 | -0.048 | 0.063 | 0.442 | -0.077 | 0.064 | 0.227 | -0.068 | 0.038 | 0.071 |
| West china | -0.090 | 0.071 | 0.203 | -0.043 | 0.065 | 0.512 | -0.069 | 0.067 | 0.300 | -0.131 | 0.039 | 0.001 |
| South china | -0.015 | 0.072 | 0.839 | -0.069 | 0.067 | 0.302 | -0.045 | 0.068 | 0.508 | -0.030 | 0.040 | 0.453 |
| History of TIA | 0.028 | 0.050 | 0.581 | 0.007 | 0.046 | 0.877 | -0.003 | 0.047 | 0.946 | 0.024 | 0.028 | 0.393 |
| History of Diabetes | 0.024 | 0.030 | 0.437 | -0.034 | 0.028 | 0.231 | -0.027 | 0.029 | 0.342 | 0.003 | 0.017 | 0.870 |
| History of Hypertension | -0.007 | 0.018 | 0.707 | -0.011 | 0.017 | 0.535 | 0.024 | 0.017 | 0.176 | -0.012 | 0.010 | 0.258 |
| History of Smoke | 0.007 | 0.008 | 0.410 | -0.011 | 0.008 | 0.169 | -0.002 | 0.008 | 0.762 | -0.003 | 0.005 | 0.492 |
| History of Drink | -0.018 | 0.025 | 0.465 | 0.014 | 0.023 | 0.550 | 0.002 | 0.023 | 0.924 | 0.003 | 0.014 | 0.823 |
| History of lipid metabolism disorder | 0.044 | 0.031 | 0.156 | 0.030 | 0.029 | 0.300 | 0.034 | 0.030 | 0.246 | -0.002 | 0.017 | 0.908 |
| History of Atrial Fibrillation | 0.132 | 0.045 | 0.004 | -0.043 | 0.042 | 0.302 | 0.098 | 0.043 | 0.023 | 0.091 | 0.025 | 0.000 |
| NIHSS at administration | -0.052 | 0.002 | 0.000 | -0.019 | 0.002 | 0.000 | -0.045 | 0.002 | 0.000 | 0.002 | 0.001 | 0.053 |
| History of Coronary Heart Disease | -0.023 | 0.027 | 0.410 | -0.002 | 0.025 | 0.934 | -0.007 | 0.026 | 0.799 | -0.005 | 0.015 | 0.761 |
| large artery atherosclerosis | -0.127 | 0.083 | 0.126 | -0.050 | 0.077 | 0.518 | 0.081 | 0.079 | 0.306 | -0.080 | 0.047 | 0.085 |
| cardiogenic embolism | -0.120 | 0.092 | 0.193 | 0.061 | 0.085 | 0.474 | 0.087 | 0.087 | 0.317 | -0.084 | 0.052 | 0.104 |

| | | | | | | | | | | | | |
|---|---|---|---|---|---|---|---|---|---|---|---|---|
| Small artery occlusion | -0.079 | 0.083 | 0.341 | -0.012 | 0.077 | 0.879 | 0.119 | 0.079 | 0.133 | -0.093 | 0.047 | 0.046 |
| stroke of an undetermined cause | -0.103 | 0.082 | 0.212 | -0.016 | 0.076 | 0.839 | 0.102 | 0.078 | 0.194 | -0.068 | 0.046 | 0.143 |
| Anticoagulation treatment | -0.076 | 0.030 | 0.013 | -0.019 | 0.028 | 0.508 | -0.047 | 0.029 | 0.105 | -0.048 | 0.017 | 0.005 |
| Lipid-lowering drugs | 0.060 | 0.043 | 0.157 | 0.064 | 0.039 | 0.107 | 0.065 | 0.040 | 0.109 | -0.051 | 0.024 | 0.031 |
| Antihypertensive treatment | -0.027 | 0.018 | 0.126 | -0.003 | 0.017 | 0.870 | -0.034 | 0.017 | 0.045 | -0.005 | 0.010 | 0.594 |
| Anti-platelet therapy | 0.087 | 0.051 | 0.092 | 0.002 | 0.048 | 0.961 | 0.033 | 0.049 | 0.496 | 0.017 | 0.029 | 0.555 |
| Hypoglycemic treatment | -0.023 | 0.029 | 0.436 | -0.036 | 0.027 | 0.189 | -0.001 | 0.028 | 0.975 | -0.036 | 0.016 | 0.029 |
| rt-PA intravenous thrombolytic | 0.147 | 0.028 | 0.000 | 0.118 | 0.026 | 0.000 | 0.142 | 0.026 | 0.000 | 0.101 | 0.016 | 0.000 |
| Mechanical thrombectomy | -0.585 | 0.207 | 0.005 | -0.010 | 0.192 | 0.957 | -0.509 | 0.196 | 0.009 | 0.032 | 0.116 | 0.784 |

**Table S10d. Multi-variables regression of basic clinical variables and Delta-NIHSS$_{(admission-discharge)}$ in scenario 1**

| | Delta-NIH8 | | | Delta-NIH9 | | | Delta-NIH10 | | | Delta-NIH11 | | |
|---|---|---|---|---|---|---|---|---|---|---|---|---|
| variables | Estimate | SE | P-value | Estimate | SE | P-value | Estimate | SE | P-value | Estimate | SE | P-value |
| (Intercept) | 0.070 | 0.153 | 0.650 | -0.007 | 0.174 | 0.967 | 0.899 | 0.250 | 0.000 | 0.217 | 0.086 | 0.012 |
| AGE | 0.000 | 0.000 | 0.660 | -0.001 | 0.000 | 0.136 | -0.001 | 0.001 | 0.400 | 0.000 | 0.000 | 0.731 |
| GENDER | -0.011 | 0.012 | 0.345 | 0.013 | 0.013 | 0.310 | 0.025 | 0.019 | 0.187 | -0.013 | 0.006 | 0.051 |
| Central China | 0.117 | 0.036 | 0.001 | 0.024 | 0.041 | 0.564 | 0.096 | 0.059 | 0.104 | 0.013 | 0.020 | 0.526 |
| Northeast China | 0.122 | 0.036 | 0.001 | 0.024 | 0.041 | 0.565 | 0.087 | 0.059 | 0.138 | 0.019 | 0.020 | 0.338 |
| Northwest china | 0.111 | 0.036 | 0.002 | 0.010 | 0.041 | 0.804 | 0.070 | 0.058 | 0.230 | 0.023 | 0.020 | 0.262 |
| East china | 0.098 | 0.037 | 0.008 | -0.083 | 0.042 | 0.048 | 0.028 | 0.060 | 0.648 | 0.012 | 0.021 | 0.554 |
| West china | 0.096 | 0.039 | 0.014 | -0.058 | 0.044 | 0.186 | 0.195 | 0.063 | 0.002 | 0.012 | 0.022 | 0.576 |
| South china | 0.058 | 0.040 | 0.142 | -0.020 | 0.045 | 0.661 | 0.047 | 0.065 | 0.463 | -0.009 | 0.022 | 0.688 |
| History of TIA | 0.031 | 0.027 | 0.254 | 0.031 | 0.031 | 0.324 | 0.038 | 0.045 | 0.395 | 0.000 | 0.015 | 0.979 |
| History of Diabetes | -0.019 | 0.017 | 0.252 | -0.033 | 0.019 | 0.084 | 0.007 | 0.027 | 0.801 | 0.005 | 0.009 | 0.580 |
| History of Hypertension | -0.005 | 0.010 | 0.645 | -0.010 | 0.011 | 0.393 | -0.033 | 0.016 | 0.046 | 0.002 | 0.006 | 0.759 |
| History of Smoke | -0.002 | 0.005 | 0.723 | 0.003 | 0.005 | 0.528 | -0.012 | 0.008 | 0.103 | -0.001 | 0.003 | 0.794 |
| History of Drink | -0.004 | 0.013 | 0.782 | 0.004 | 0.015 | 0.790 | 0.010 | 0.022 | 0.646 | -0.001 | 0.008 | 0.881 |
| History of lipid metabolism disorder | 0.015 | 0.017 | 0.387 | 0.019 | 0.020 | 0.326 | 0.046 | 0.028 | 0.100 | 0.005 | 0.010 | 0.580 |
| History of Atrial Fibrillation | 0.033 | 0.025 | 0.185 | -0.031 | 0.028 | 0.277 | -0.061 | 0.041 | 0.131 | 0.042 | 0.014 | 0.003 |
| NIHSS at administration | 0.003 | 0.001 | 0.031 | -0.027 | 0.001 | 0.000 | -0.031 | 0.002 | 0.000 | 0.019 | 0.001 | 0.000 |
| History of Coronary Heart Disease | -0.003 | 0.015 | 0.847 | -0.018 | 0.017 | 0.294 | -0.037 | 0.025 | 0.128 | -0.002 | 0.008 | 0.802 |
| large artery atherosclerosis | -0.073 | 0.046 | 0.113 | -0.053 | 0.052 | 0.313 | -0.047 | 0.075 | 0.533 | -0.056 | 0.026 | 0.031 |
| cardiogenic embolism | -0.008 | 0.051 | 0.875 | 0.017 | 0.058 | 0.766 | 0.075 | 0.083 | 0.361 | -0.053 | 0.028 | 0.064 |

| | | | | | | | | | | | | |
|---|---|---|---|---|---|---|---|---|---|---|---|---|
| Small artery occlusion | -0.112 | 0.046 | 0.015 | -0.014 | 0.052 | 0.793 | -0.049 | 0.075 | 0.514 | -0.065 | 0.026 | 0.011 |
| stroke of an undetermined cause | -0.043 | 0.045 | 0.345 | -0.021 | 0.052 | 0.679 | -0.038 | 0.074 | 0.608 | -0.059 | 0.025 | 0.020 |
| Anticoagulation treatment | 0.014 | 0.017 | 0.395 | 0.004 | 0.019 | 0.844 | -0.047 | 0.027 | 0.084 | -0.012 | 0.009 | 0.184 |
| Lipid-lowering drugs | -0.066 | 0.023 | 0.005 | -0.012 | 0.027 | 0.643 | -0.095 | 0.038 | 0.012 | -0.002 | 0.013 | 0.908 |
| Antihypertensive treatment | -0.009 | 0.010 | 0.380 | -0.006 | 0.011 | 0.620 | -0.014 | 0.016 | 0.390 | -0.004 | 0.006 | 0.503 |
| Anti-platelet therapy | -0.026 | 0.028 | 0.357 | 0.034 | 0.032 | 0.285 | 0.001 | 0.046 | 0.987 | -0.004 | 0.016 | 0.795 |
| Hypoglycemic treatment | -0.002 | 0.016 | 0.926 | 0.053 | 0.018 | 0.004 | -0.032 | 0.026 | 0.223 | -0.004 | 0.009 | 0.635 |
| rt-PA intravenous thrombolytic | 0.080 | 0.015 | 0.000 | 0.131 | 0.017 | 0.000 | 0.149 | 0.025 | 0.000 | 0.037 | 0.009 | 0.000 |
| Mechanical thrombectomy | -0.132 | 0.114 | 0.245 | -0.222 | 0.129 | 0.086 | -0.859 | 0.185 | 0.000 | -0.196 | 0.064 | 0.002 |

**Table S11a. Multi-variables regression of basic clinical variables and Delta-NIHSS$_{(admission-discharge)}$ in scenario 2**

| | Delta-NIHSS | | | Delta-NIH1A | | | Delta-NIH1B | | | Delta-NIH1C | | |
|---|---|---|---|---|---|---|---|---|---|---|---|---|
| variables | Estimate | SE | P-value | Estimate | SE | P-value | Estimate | SE | P-value | Estimate | SE | P-value |
| (Intercept) | 0.632 | 0.464 | 0.173 | 0.025 | 0.057 | 0.664 | 0.062 | 0.063 | 0.331 | 0.099 | 0.054 | 0.064 |
| AGE | -0.012 | 0.002 | 0.000 | -0.001 | 0.000 | 0.023 | 0.000 | 0.000 | 0.180 | 0.000 | 0.000 | 0.381 |
| GENDER | -0.064 | 0.055 | 0.244 | -0.004 | 0.007 | 0.544 | -0.010 | 0.008 | 0.204 | -0.002 | 0.006 | 0.714 |
| Central China | -0.023 | 0.159 | 0.884 | -0.001 | 0.020 | 0.978 | 0.005 | 0.022 | 0.817 | -0.032 | 0.018 | 0.080 |
| Northeast China | -0.113 | 0.158 | 0.475 | 0.004 | 0.020 | 0.829 | 0.007 | 0.022 | 0.732 | -0.015 | 0.018 | 0.414 |
| Northwest china | 0.016 | 0.157 | 0.920 | 0.011 | 0.019 | 0.565 | 0.008 | 0.021 | 0.704 | -0.014 | 0.018 | 0.431 |
| East china | -0.173 | 0.165 | 0.295 | 0.001 | 0.020 | 0.977 | 0.009 | 0.023 | 0.683 | -0.016 | 0.019 | 0.413 |
| West china | -0.064 | 0.171 | 0.711 | 0.023 | 0.021 | 0.284 | 0.001 | 0.023 | 0.953 | -0.032 | 0.020 | 0.111 |
| South china | -0.163 | 0.176 | 0.354 | -0.006 | 0.022 | 0.783 | -0.031 | 0.024 | 0.194 | -0.047 | 0.020 | 0.020 |
| History of TIA | 0.122 | 0.128 | 0.341 | 0.025 | 0.016 | 0.117 | 0.012 | 0.018 | 0.495 | 0.029 | 0.015 | 0.053 |
| History of Diabetes | -0.127 | 0.077 | 0.102 | 0.001 | 0.010 | 0.953 | 0.013 | 0.011 | 0.229 | -0.005 | 0.009 | 0.542 |
| History of Hypertension | -0.155 | 0.048 | 0.001 | -0.011 | 0.006 | 0.074 | -0.009 | 0.007 | 0.178 | -0.019 | 0.006 | 0.001 |
| History of Smoke | -0.033 | 0.022 | 0.137 | -0.002 | 0.003 | 0.433 | -0.003 | 0.003 | 0.356 | -0.004 | 0.003 | 0.101 |
| History of Drink | -0.072 | 0.065 | 0.265 | -0.003 | 0.008 | 0.709 | -0.001 | 0.009 | 0.927 | 0.006 | 0.007 | 0.426 |
| History of lipid metabolism disorder | 0.089 | 0.079 | 0.257 | 0.013 | 0.010 | 0.189 | 0.012 | 0.011 | 0.276 | 0.010 | 0.009 | 0.255 |
| History of Atrial Fibrillation | -0.281 | 0.117 | 0.017 | 0.028 | 0.014 | 0.058 | 0.028 | 0.016 | 0.083 | 0.009 | 0.014 | 0.502 |
| NIHSS at administration | 0.433 | 0.006 | 0.000 | 0.024 | 0.001 | 0.000 | 0.027 | 0.001 | 0.000 | 0.024 | 0.001 | 0.000 |
| History of Coronary Heart Disease | 0.040 | 0.071 | 0.575 | -0.002 | 0.009 | 0.790 | -0.002 | 0.010 | 0.834 | -0.004 | 0.008 | 0.630 |
| large artery atherosclerosis | -0.116 | 0.197 | 0.556 | 0.028 | 0.024 | 0.248 | 0.011 | 0.027 | 0.687 | 0.008 | 0.023 | 0.730 |
| cardiogenic embolism | 0.634 | 0.225 | 0.005 | 0.017 | 0.028 | 0.539 | 0.031 | 0.031 | 0.320 | 0.020 | 0.026 | 0.445 |

| | | | | | | | | | | | | |
|---|---|---|---|---|---|---|---|---|---|---|---|---|
| Small artery occlusion | 0.345 | 0.198 | 0.081 | 0.027 | 0.024 | 0.261 | 0.001 | 0.027 | 0.978 | -0.002 | 0.023 | 0.943 |
| stroke of an undetermined cause | 0.235 | 0.195 | 0.228 | 0.032 | 0.024 | 0.188 | 0.023 | 0.027 | 0.396 | 0.012 | 0.022 | 0.583 |
| Anticoagulation treatment | -0.427 | 0.075 | 0.000 | 0.007 | 0.009 | 0.456 | -0.010 | 0.010 | 0.355 | -0.013 | 0.009 | 0.122 |
| Lipid-lowering drugs | 0.064 | 0.114 | 0.576 | -0.011 | 0.014 | 0.446 | -0.021 | 0.016 | 0.169 | -0.038 | 0.013 | 0.004 |
| Antihypertensive treatment | 0.085 | 0.047 | 0.067 | 0.007 | 0.006 | 0.198 | 0.006 | 0.006 | 0.367 | 0.009 | 0.005 | 0.102 |
| Anti-platelet therapy | 0.134 | 0.139 | 0.333 | -0.025 | 0.017 | 0.140 | -0.008 | 0.019 | 0.681 | -0.013 | 0.016 | 0.405 |
| Hypoglycemic treatment | -0.038 | 0.075 | 0.609 | -0.010 | 0.009 | 0.261 | -0.033 | 0.010 | 0.001 | -0.011 | 0.009 | 0.215 |

**Table S11b. Multi-variables regression of basic clinical variables and Delta-NIHSS$_{(admission-discharge)}$ in scenario 2**

| | Delta-NIH2 | | | Delta-NIH3 | | | Delta-NIH4 | | | Delta-NIH5A | | |
|---|---|---|---|---|---|---|---|---|---|---|---|---|
| variables | Estimate | SE | P-value | Estimate | SE | P-value | Estimate | SE | P-value | Estimate | SE | P-value |
| (Intercept) | 0.014 | 0.042 | 0.738 | -0.018 | 0.045 | 0.688 | 0.063 | 0.103 | 0.542 | 0.043 | 0.153 | 0.777 |
| AGE | 0.000 | 0.000 | 0.559 | 0.000 | 0.000 | 0.584 | -0.002 | 0.000 | 0.000 | -0.002 | 0.001 | 0.001 |
| GENDER | -0.010 | 0.005 | 0.040 | -0.003 | 0.005 | 0.569 | 0.008 | 0.012 | 0.529 | -0.004 | 0.018 | 0.811 |
| Central China | 0.025 | 0.015 | 0.087 | 0.017 | 0.016 | 0.279 | -0.085 | 0.036 | 0.017 | 0.076 | 0.053 | 0.150 |
| Northeast China | 0.020 | 0.014 | 0.160 | 0.015 | 0.015 | 0.329 | -0.099 | 0.035 | 0.005 | 0.071 | 0.052 | 0.172 |
| Northwest china | 0.025 | 0.014 | 0.081 | 0.016 | 0.015 | 0.291 | -0.125 | 0.035 | 0.000 | 0.080 | 0.052 | 0.122 |
| East china | 0.020 | 0.015 | 0.175 | 0.021 | 0.016 | 0.195 | -0.148 | 0.037 | 0.000 | 0.035 | 0.054 | 0.520 |
| West china | 0.017 | 0.016 | 0.272 | 0.017 | 0.017 | 0.299 | -0.141 | 0.038 | 0.000 | 0.081 | 0.057 | 0.150 |
| South china | 0.002 | 0.016 | 0.920 | 0.012 | 0.017 | 0.491 | -0.097 | 0.039 | 0.014 | 0.122 | 0.058 | 0.035 |
| History of TIA | 0.011 | 0.012 | 0.346 | 0.008 | 0.013 | 0.501 | 0.008 | 0.029 | 0.783 | 0.023 | 0.042 | 0.591 |
| History of Diabetes | -0.002 | 0.007 | 0.808 | 0.003 | 0.008 | 0.697 | -0.037 | 0.017 | 0.034 | 0.019 | 0.026 | 0.466 |
| History of Hypertension | 0.000 | 0.004 | 0.951 | -0.004 | 0.005 | 0.362 | -0.008 | 0.011 | 0.440 | -0.011 | 0.016 | 0.495 |
| History of Smoke | 0.000 | 0.002 | 0.937 | 0.002 | 0.002 | 0.321 | -0.001 | 0.005 | 0.821 | -0.007 | 0.007 | 0.327 |
| History of Drink | -0.007 | 0.006 | 0.246 | 0.002 | 0.006 | 0.746 | 0.005 | 0.014 | 0.751 | -0.009 | 0.021 | 0.677 |
| History of lipid metabolism disorder | 0.007 | 0.007 | 0.300 | 0.007 | 0.008 | 0.355 | -0.001 | 0.018 | 0.975 | -0.032 | 0.026 | 0.219 |
| History of Atrial Fibrillation | 0.022 | 0.011 | 0.040 | 0.017 | 0.011 | 0.126 | 0.016 | 0.026 | 0.549 | 0.011 | 0.039 | 0.781 |
| NIHSS at administration | 0.014 | 0.001 | 0.000 | 0.007 | 0.001 | 0.000 | 0.029 | 0.001 | 0.000 | 0.049 | 0.002 | 0.000 |
| History of Coronary Heart Disease | -0.008 | 0.006 | 0.242 | -0.007 | 0.007 | 0.324 | 0.020 | 0.016 | 0.203 | 0.046 | 0.023 | 0.049 |
| large artery atherosclerosis | 0.006 | 0.018 | 0.728 | 0.015 | 0.019 | 0.448 | -0.020 | 0.044 | 0.641 | 0.007 | 0.065 | 0.920 |
| cardiogenic embolism | 0.012 | 0.020 | 0.562 | 0.005 | 0.022 | 0.803 | 0.015 | 0.050 | 0.772 | 0.168 | 0.074 | 0.023 |

| | | | | | | | | | | | | |
|---|---|---|---|---|---|---|---|---|---|---|---|---|
| Small artery occlusion | -0.002 | 0.018 | 0.911 | 0.012 | 0.019 | 0.533 | 0.033 | 0.044 | 0.455 | 0.086 | 0.065 | 0.188 |
| stroke of an undetermined cause | -0.003 | 0.018 | 0.876 | 0.015 | 0.019 | 0.430 | 0.005 | 0.043 | 0.906 | 0.075 | 0.064 | 0.243 |
| Anticoagulation treatment | 0.003 | 0.007 | 0.700 | 0.004 | 0.007 | 0.549 | -0.019 | 0.017 | 0.268 | -0.110 | 0.025 | 0.000 |
| Lipid-lowering drugs | -0.022 | 0.010 | 0.031 | 0.001 | 0.011 | 0.901 | 0.021 | 0.025 | 0.401 | 0.039 | 0.038 | 0.300 |
| Antihypertensive treatment | -0.003 | 0.004 | 0.481 | 0.000 | 0.005 | 0.976 | -0.007 | 0.010 | 0.478 | 0.033 | 0.015 | 0.034 |
| Anti-platelet therapy | 0.002 | 0.013 | 0.897 | -0.013 | 0.014 | 0.328 | 0.080 | 0.031 | 0.009 | -0.027 | 0.046 | 0.560 |
| Hypoglycemic treatment | -0.004 | 0.007 | 0.515 | -0.001 | 0.007 | 0.930 | 0.022 | 0.017 | 0.196 | -0.019 | 0.025 | 0.432 |

**Table S11c. Multi-variables regression of basic clinical variables and Delta-NIHSS$_{(admission-discharge)}$ in scenario 2**

| | Delta-NIH5B | | | Delta-NIH6A | | | Delta-NIH6B | | | Delta-NIH7 | | |
|---|---|---|---|---|---|---|---|---|---|---|---|---|
| variables | Estimate | SE | P-value | Estimate | SE | P-value | Estimate | SE | P-value | Estimate | SE | P-value |
| (Intercept) | 0.225 | 0.153 | 0.142 | 0.166 | 0.142 | 0.241 | 0.208 | 0.139 | 0.134 | 0.183 | 0.084 | 0.030 |
| AGE | -0.002 | 0.001 | 0.020 | -0.003 | 0.001 | 0.000 | -0.003 | 0.001 | 0.000 | 0.000 | 0.000 | 0.409 |
| GENDER | -0.030 | 0.018 | 0.103 | -0.037 | 0.017 | 0.029 | -0.023 | 0.016 | 0.170 | 0.003 | 0.010 | 0.779 |
| Central China | -0.031 | 0.053 | 0.551 | -0.072 | 0.049 | 0.142 | -0.011 | 0.048 | 0.823 | -0.070 | 0.029 | 0.017 |
| Northeast China | -0.007 | 0.052 | 0.889 | -0.029 | 0.048 | 0.547 | 0.006 | 0.048 | 0.907 | -0.039 | 0.029 | 0.179 |
| Northwest china | 0.034 | 0.052 | 0.514 | -0.022 | 0.048 | 0.646 | 0.035 | 0.047 | 0.461 | -0.057 | 0.029 | 0.045 |
| East china | -0.049 | 0.054 | 0.372 | -0.053 | 0.050 | 0.292 | -0.048 | 0.050 | 0.328 | -0.077 | 0.030 | 0.010 |
| West china | -0.064 | 0.057 | 0.259 | -0.054 | 0.052 | 0.307 | -0.018 | 0.051 | 0.727 | -0.181 | 0.031 | 0.000 |
| South china | -0.048 | 0.058 | 0.412 | -0.121 | 0.054 | 0.024 | -0.034 | 0.053 | 0.517 | -0.037 | 0.032 | 0.251 |
| History of TIA | 0.009 | 0.042 | 0.835 | 0.045 | 0.039 | 0.251 | 0.008 | 0.038 | 0.839 | 0.016 | 0.023 | 0.490 |
| History of Diabetes | -0.002 | 0.026 | 0.936 | -0.010 | 0.024 | 0.664 | -0.044 | 0.023 | 0.060 | 0.004 | 0.014 | 0.752 |
| History of Hypertension | -0.017 | 0.016 | 0.278 | -0.032 | 0.015 | 0.032 | -0.001 | 0.014 | 0.951 | -0.016 | 0.009 | 0.063 |
| History of Smoke | 0.011 | 0.007 | 0.134 | -0.014 | 0.007 | 0.039 | -0.001 | 0.007 | 0.876 | -0.005 | 0.004 | 0.261 |
| History of Drink | -0.046 | 0.021 | 0.030 | 0.013 | 0.020 | 0.515 | -0.020 | 0.019 | 0.302 | -0.001 | 0.012 | 0.952 |
| History of lipid metabolism disorder | -0.003 | 0.026 | 0.906 | 0.036 | 0.024 | 0.138 | 0.000 | 0.024 | 0.987 | -0.011 | 0.014 | 0.446 |
| History of Atrial Fibrillation | 0.136 | 0.039 | 0.000 | -0.042 | 0.036 | 0.244 | 0.038 | 0.035 | 0.287 | 0.102 | 0.021 | 0.000 |
| NIHSS at administration | -0.062 | 0.002 | 0.000 | -0.028 | 0.002 | 0.000 | -0.055 | 0.002 | 0.000 | 0.000 | 0.001 | 0.844 |
| History of Coronary Heart Disease | 0.023 | 0.023 | 0.320 | -0.039 | 0.022 | 0.072 | 0.035 | 0.021 | 0.095 | -0.010 | 0.013 | 0.460 |
| large artery atherosclerosis | -0.086 | 0.065 | 0.187 | 0.004 | 0.060 | 0.944 | 0.056 | 0.059 | 0.339 | -0.007 | 0.036 | 0.851 |
| cardiogenic embolism | -0.035 | 0.074 | 0.634 | 0.141 | 0.069 | 0.041 | 0.118 | 0.067 | 0.081 | -0.036 | 0.041 | 0.376 |

| | | | | | | | | | | | | |
|---|---|---|---|---|---|---|---|---|---|---|---|---|
| Small artery occlusion | -0.024 | 0.065 | 0.708 | 0.067 | 0.060 | 0.267 | 0.105 | 0.059 | 0.077 | -0.021 | 0.036 | 0.555 |
| stroke of an undetermined cause | -0.057 | 0.064 | 0.376 | 0.050 | 0.060 | 0.405 | 0.089 | 0.058 | 0.130 | -0.009 | 0.036 | 0.805 |
| Anticoagulation treatment | -0.111 | 0.025 | 0.000 | -0.068 | 0.023 | 0.003 | -0.059 | 0.023 | 0.009 | -0.055 | 0.014 | 0.000 |
| Lipid-lowering drugs | -0.004 | 0.037 | 0.915 | 0.038 | 0.035 | 0.270 | 0.000 | 0.034 | 0.999 | -0.048 | 0.021 | 0.020 |
| Antihypertensive treatment | -0.022 | 0.015 | 0.147 | 0.008 | 0.014 | 0.581 | -0.019 | 0.014 | 0.174 | 0.003 | 0.008 | 0.725 |
| Anti-platelet therapy | 0.044 | 0.046 | 0.338 | 0.005 | 0.042 | 0.904 | -0.015 | 0.042 | 0.726 | 0.023 | 0.025 | 0.359 |
| Hypoglycemic treatment | 0.013 | 0.025 | 0.587 | -0.038 | 0.023 | 0.101 | 0.026 | 0.022 | 0.255 | -0.046 | 0.014 | 0.001 |

**Table S11d. Multi-variables regression of basic clinical variables and Delta-NIHSS$_{(admission-discharge)}$ in scenario 2**

| | Delta-NIH8 | | | Delta-NIH9 | | | Delta-NIH10 | | | Delta-NIH11 | | |
|---|---|---|---|---|---|---|---|---|---|---|---|---|
| variables | Estimate | SE | P-value | Estimate | SE | P-value | Estimate | SE | P-value | Estimate | SE | P-value |
| (Intercept) | 0.014 | 0.081 | 0.861 | 0.019 | 0.095 | 0.844 | 0.135 | 0.129 | 0.295 | 0.026 | 0.043 | 0.546 |
| AGE | 0.000 | 0.000 | 0.195 | -0.001 | 0.000 | 0.027 | -0.001 | 0.001 | 0.027 | 0.000 | 0.000 | 0.370 |
| GENDER | -0.029 | 0.010 | 0.003 | 0.009 | 0.011 | 0.408 | 0.035 | 0.015 | 0.022 | -0.013 | 0.005 | 0.012 |
| Central China | 0.140 | 0.028 | 0.000 | -0.011 | 0.033 | 0.730 | 0.038 | 0.044 | 0.390 | 0.023 | 0.015 | 0.122 |
| Northeast China | 0.143 | 0.028 | 0.000 | -0.013 | 0.032 | 0.680 | 0.027 | 0.044 | 0.545 | 0.027 | 0.015 | 0.070 |
| Northwest china | 0.139 | 0.027 | 0.000 | -0.014 | 0.032 | 0.661 | 0.023 | 0.043 | 0.597 | 0.033 | 0.015 | 0.025 |
| East china | 0.133 | 0.029 | 0.000 | -0.080 | 0.034 | 0.017 | -0.006 | 0.046 | 0.888 | 0.029 | 0.015 | 0.063 |
| West china | 0.080 | 0.030 | 0.008 | -0.062 | 0.035 | 0.076 | 0.156 | 0.048 | 0.001 | 0.018 | 0.016 | 0.262 |
| South china | 0.086 | 0.031 | 0.005 | -0.049 | 0.036 | 0.169 | -0.016 | 0.049 | 0.746 | 0.002 | 0.017 | 0.917 |
| History of TIA | 0.030 | 0.022 | 0.178 | 0.045 | 0.026 | 0.089 | 0.039 | 0.036 | 0.276 | 0.006 | 0.012 | 0.644 |
| History of Diabetes | -0.027 | 0.014 | 0.045 | -0.032 | 0.016 | 0.043 | -0.015 | 0.021 | 0.498 | 0.000 | 0.007 | 0.988 |
| History of Hypertension | -0.002 | 0.008 | 0.825 | -0.008 | 0.010 | 0.435 | -0.019 | 0.013 | 0.145 | 0.003 | 0.005 | 0.467 |
| History of Smoke | -0.008 | 0.004 | 0.037 | 0.002 | 0.005 | 0.658 | -0.009 | 0.006 | 0.130 | 0.001 | 0.002 | 0.761 |
| History of Drink | -0.012 | 0.011 | 0.289 | -0.001 | 0.013 | 0.945 | -0.005 | 0.018 | 0.774 | -0.007 | 0.006 | 0.256 |
| History of lipid metabolism disorder | 0.009 | 0.014 | 0.514 | -0.001 | 0.016 | 0.938 | -0.001 | 0.022 | 0.962 | 0.000 | 0.007 | 0.977 |
| History of Atrial Fibrillation | 0.034 | 0.021 | 0.096 | -0.010 | 0.024 | 0.680 | -0.050 | 0.033 | 0.122 | 0.047 | 0.011 | 0.000 |
| NIHSS at administration | 0.000 | 0.001 | 0.936 | -0.030 | 0.001 | 0.000 | -0.034 | 0.002 | 0.000 | 0.018 | 0.001 | 0.000 |
| History of Coronary Heart Disease | 0.004 | 0.012 | 0.739 | -0.002 | 0.014 | 0.885 | -0.008 | 0.020 | 0.686 | -0.004 | 0.007 | 0.546 |
| large artery atherosclerosis | -0.015 | 0.034 | 0.655 | -0.061 | 0.040 | 0.127 | -0.005 | 0.055 | 0.934 | -0.017 | 0.018 | 0.354 |
| cardiogenic embolism | 0.021 | 0.039 | 0.589 | 0.002 | 0.046 | 0.961 | 0.036 | 0.062 | 0.561 | -0.035 | 0.021 | 0.096 |

| | | | | | | | | | | | | |
|---|---|---|---|---|---|---|---|---|---|---|---|---|
| Small artery occlusion | -0.066 | 0.035 | 0.057 | 0.004 | 0.040 | 0.921 | -0.018 | 0.055 | 0.745 | -0.030 | 0.019 | 0.106 |
| stroke of an undetermined cause | -0.004 | 0.034 | 0.896 | -0.011 | 0.040 | 0.782 | -0.008 | 0.054 | 0.883 | -0.025 | 0.018 | 0.170 |
| Anticoagulation treatment | 0.003 | 0.013 | 0.808 | 0.001 | 0.015 | 0.972 | -0.028 | 0.021 | 0.178 | -0.002 | 0.007 | 0.773 |
| Lipid-lowering drugs | -0.069 | 0.020 | 0.001 | 0.007 | 0.023 | 0.778 | -0.091 | 0.032 | 0.004 | -0.009 | 0.011 | 0.412 |
| Antihypertensive treatment | -0.001 | 0.008 | 0.927 | -0.002 | 0.009 | 0.827 | -0.015 | 0.013 | 0.241 | -0.004 | 0.004 | 0.342 |
| Anti-platelet therapy | -0.038 | 0.024 | 0.120 | -0.016 | 0.028 | 0.564 | 0.029 | 0.038 | 0.449 | -0.006 | 0.013 | 0.655 |
| Hypoglycemic treatment | -0.007 | 0.013 | 0.590 | 0.041 | 0.015 | 0.007 | -0.011 | 0.021 | 0.587 | -0.005 | 0.007 | 0.508 |

**Table S12a. Multi-variables regression of 38 biomarkers and Delta-NIHSS(admission-discharge)**

| | Delta-NIHSS | | | Delta-NIH1A | | | Delta-NIH1B | | | Delta-NIH1C | | |
|---|---|---|---|---|---|---|---|---|---|---|---|---|
| variables | Estimate | SE | P-value | Estimate | SE | P-value | Estimate | SE | P-value | Estimate | SE | P-value |
| (Intercept) | 0.095 | 0.004 | 0.000 | 0.012 | 0.002 | 0.000 | 0.015 | 0.002 | 0.000 | 0.012 | 0.002 | 0.000 |
| Adiponectin | -0.002 | 0.004 | 0.630 | 0.000 | 0.002 | 0.920 | 0.005 | 0.002 | 0.003 | 0.001 | 0.002 | 0.410 |
| ANGPTL3 | -0.007 | 0.005 | 0.107 | -0.002 | 0.002 | 0.275 | -0.001 | 0.002 | 0.451 | 0.001 | 0.002 | 0.587 |
| Vitamin B12 | 0.005 | 0.004 | 0.254 | 0.001 | 0.002 | 0.551 | 0.004 | 0.002 | 0.044 | 0.004 | 0.002 | 0.024 |
| Folic acid | 0.009 | 0.005 | 0.083 | 0.000 | 0.002 | 0.902 | 0.003 | 0.002 | 0.127 | -0.002 | 0.002 | 0.338 |
| Creatinine | -0.005 | 0.008 | 0.518 | 0.000 | 0.003 | 0.963 | -0.002 | 0.003 | 0.526 | 0.002 | 0.003 | 0.398 |
| Cystatin C | -0.003 | 0.007 | 0.665 | -0.003 | 0.003 | 0.239 | -0.001 | 0.003 | 0.705 | -0.005 | 0.003 | 0.066 |
| D-dimer DD | 0.001 | 0.005 | 0.831 | 0.001 | 0.002 | 0.433 | 0.005 | 0.002 | 0.004 | 0.004 | 0.002 | 0.021 |
| Fibrinogen Fib | 0.004 | 0.005 | 0.423 | 0.000 | 0.002 | 0.923 | -0.001 | 0.002 | 0.795 | -0.003 | 0.002 | 0.115 |
| Fasting blood glucose | 0.008 | 0.004 | 0.077 | 0.003 | 0.002 | 0.044 | 0.004 | 0.002 | 0.035 | 0.002 | 0.002 | 0.317 |
| Homocysteine HCY | 0.003 | 0.005 | 0.461 | 0.001 | 0.002 | 0.686 | 0.000 | 0.002 | 0.871 | 0.002 | 0.002 | 0.311 |
| High-density lipoprotein | -0.008 | 0.005 | 0.141 | 0.001 | 0.002 | 0.651 | -0.001 | 0.002 | 0.687 | 0.000 | 0.002 | 0.854 |
| High-sensitivity C-reactive protein | 0.005 | 0.004 | 0.240 | 0.000 | 0.002 | 0.816 | 0.003 | 0.002 | 0.069 | 0.000 | 0.002 | 0.913 |
| Interleukin-1 Receptor Antagonist | 0.010 | 0.005 | 0.053 | 0.000 | 0.002 | 0.845 | 0.002 | 0.002 | 0.300 | 0.002 | 0.002 | 0.413 |
| Interleukin-6 receptor Antagonist | 0.009 | 0.005 | 0.061 | 0.009 | 0.002 | 0.000 | 0.003 | 0.002 | 0.140 | 0.006 | 0.002 | 0.001 |
| Interleukin-6 receptor | -0.005 | 0.004 | 0.265 | -0.001 | 0.002 | 0.723 | 0.000 | 0.002 | 0.801 | 0.002 | 0.002 | 0.177 |
| Low-density lipoprotein | -0.002 | 0.006 | 0.753 | 0.002 | 0.002 | 0.345 | 0.000 | 0.002 | 0.895 | -0.001 | 0.002 | 0.751 |
| LDL Receptor | -0.006 | 0.005 | 0.257 | -0.001 | 0.002 | 0.648 | -0.001 | 0.002 | 0.505 | 0.000 | 0.002 | 0.833 |
| Lipoprotein phospholipase A2-activity | -0.008 | 0.007 | 0.262 | 0.003 | 0.002 | 0.208 | -0.001 | 0.003 | 0.649 | 0.003 | 0.003 | 0.187 |
| Lipoprotein phospholipase A2-MASS | 0.017 | 0.006 | 0.009 | -0.002 | 0.002 | 0.521 | 0.003 | 0.003 | 0.291 | -0.001 | 0.002 | 0.732 |

| | | | | | | | | | | | | |
|---|---|---|---|---|---|---|---|---|---|---|---|---|
| Monocyte Chemoattractant Protein-1 | -0.003 | 0.004 | 0.444 | 0.005 | 0.002 | 0.003 | -0.001 | 0.002 | 0.758 | -0.001 | 0.002 | 0.657 |
| Lipoprotein a | -0.001 | 0.004 | 0.857 | -0.003 | 0.002 | 0.060 | -0.002 | 0.002 | 0.344 | 0.002 | 0.002 | 0.162 |
| Methylmalonic acid | 0.001 | 0.004 | 0.706 | 0.000 | 0.001 | 0.720 | 0.001 | 0.001 | 0.613 | 0.001 | 0.001 | 0.565 |
| Proprotein Convertase Subtilisin 9 | 0.001 | 0.004 | 0.889 | 0.000 | 0.002 | 0.792 | -0.001 | 0.002 | 0.504 | 0.000 | 0.002 | 0.780 |
| Triglyceride | -0.009 | 0.008 | 0.216 | -0.002 | 0.003 | 0.549 | -0.002 | 0.003 | 0.430 | -0.004 | 0.003 | 0.167 |
| Human chitinase 3-like protein 1 | 0.008 | 0.005 | 0.103 | 0.003 | 0.002 | 0.141 | -0.001 | 0.002 | 0.482 | 0.004 | 0.002 | 0.015 |
| Apo-A2 | -0.001 | 0.006 | 0.815 | -0.001 | 0.002 | 0.800 | -0.002 | 0.003 | 0.466 | -0.001 | 0.002 | 0.606 |
| Apo-C3 | 0.006 | 0.011 | 0.599 | 0.003 | 0.004 | 0.468 | 0.002 | 0.005 | 0.632 | 0.000 | 0.004 | 0.989 |
| Apo-E | -0.003 | 0.008 | 0.719 | 0.002 | 0.003 | 0.558 | 0.000 | 0.003 | 0.892 | 0.000 | 0.003 | 0.868 |
| Apo-A2 | 0.006 | 0.006 | 0.372 | -0.001 | 0.002 | 0.591 | 0.001 | 0.003 | 0.840 | 0.001 | 0.002 | 0.734 |
| Apo-B | 0.002 | 0.006 | 0.797 | -0.001 | 0.002 | 0.600 | 0.001 | 0.003 | 0.770 | -0.003 | 0.002 | 0.241 |
| Apo-C2 | -0.009 | 0.010 | 0.375 | -0.005 | 0.004 | 0.215 | -0.003 | 0.004 | 0.417 | -0.002 | 0.004 | 0.541 |
| Betaine | -0.008 | 0.007 | 0.198 | -0.001 | 0.002 | 0.637 | 0.000 | 0.003 | 0.969 | -0.003 | 0.002 | 0.194 |
| Trimethylamine oxide | 0.000 | 0.005 | 0.964 | -0.001 | 0.002 | 0.732 | 0.001 | 0.002 | 0.608 | -0.001 | 0.002 | 0.439 |
| Choline | 0.004 | 0.006 | 0.535 | 0.000 | 0.002 | 0.825 | -0.002 | 0.002 | 0.503 | -0.001 | 0.002 | 0.619 |
| Carnitine | 0.010 | 0.005 | 0.022 | 0.003 | 0.002 | 0.055 | 0.005 | 0.002 | 0.015 | 0.004 | 0.002 | 0.024 |
| Trimethylpentaaminovaleric acid | 0.000 | 0.004 | 0.996 | -0.001 | 0.001 | 0.595 | 0.000 | 0.001 | 0.926 | -0.001 | 0.001 | 0.536 |
| Butyl betaine | 0.002 | 0.004 | 0.701 | 0.000 | 0.002 | 0.797 | 0.000 | 0.002 | 0.853 | 0.002 | 0.002 | 0.337 |
| tml | -0.003 | 0.006 | 0.657 | -0.001 | 0.002 | 0.681 | 0.000 | 0.003 | 0.899 | -0.001 | 0.002 | 0.641 |

**Table S12b. Multi-variables regression of 38 biomarkers and Delta-NIHSS$_{(admission-discharge)}$**

| | Delta-NIH2 | | | Delta-NIH3 | | | Delta-NIH4 | | | Delta-NIH5A | | |
|---|---|---|---|---|---|---|---|---|---|---|---|---|
| variables | Estimate | SE | P-value | Estimate | SE | P-value | Estimate | SE | P-value | Estimate | SE | P-value |
| (Intercept) | 0.009 | 0.001 | 0.000 | 0.010 | 0.001 | 0.000 | 0.062 | 0.003 | 0.000 | 0.047 | 0.003 | 0.000 |
| Adiponectin | -0.001 | 0.001 | 0.592 | 0.001 | 0.001 | 0.419 | -0.005 | 0.004 | 0.172 | -0.003 | 0.003 | 0.324 |
| ANGPTL3 | -0.001 | 0.001 | 0.401 | -0.002 | 0.002 | 0.293 | -0.001 | 0.004 | 0.840 | -0.004 | 0.003 | 0.194 |
| Vitamin B12 | 0.000 | 0.001 | 0.814 | 0.001 | 0.001 | 0.314 | 0.005 | 0.004 | 0.193 | 0.003 | 0.003 | 0.304 |
| Folic acid | -0.002 | 0.002 | 0.346 | -0.001 | 0.002 | 0.740 | 0.008 | 0.004 | 0.055 | -0.001 | 0.004 | 0.805 |
| Creatinine | 0.000 | 0.002 | 0.840 | 0.002 | 0.003 | 0.341 | -0.003 | 0.006 | 0.620 | -0.008 | 0.005 | 0.127 |
| Cystatin C | -0.001 | 0.002 | 0.569 | -0.003 | 0.002 | 0.247 | 0.001 | 0.006 | 0.916 | 0.007 | 0.005 | 0.182 |
| D-dimer DD | 0.000 | 0.001 | 0.766 | 0.000 | 0.002 | 0.895 | -0.001 | 0.004 | 0.739 | 0.005 | 0.003 | 0.167 |
| Fibrinogen Fib | 0.000 | 0.001 | 0.772 | 0.000 | 0.002 | 0.841 | 0.004 | 0.004 | 0.361 | 0.000 | 0.003 | 0.892 |
| Fasting blood glucose | 0.003 | 0.001 | 0.015 | 0.003 | 0.002 | 0.042 | 0.001 | 0.004 | 0.869 | 0.005 | 0.003 | 0.116 |
| Homocysteine HCY | -0.001 | 0.001 | 0.358 | 0.003 | 0.002 | 0.054 | 0.000 | 0.004 | 0.912 | -0.001 | 0.003 | 0.842 |
| High-density lipoprotein | 0.001 | 0.002 | 0.684 | 0.001 | 0.002 | 0.690 | 0.002 | 0.004 | 0.630 | 0.003 | 0.004 | 0.475 |
| High-sensitivity C-reactive protein | 0.003 | 0.001 | 0.015 | 0.001 | 0.001 | 0.332 | -0.004 | 0.004 | 0.296 | -0.001 | 0.003 | 0.647 |
| Interleukin-1 Receptor Antagonist | 0.002 | 0.002 | 0.138 | -0.002 | 0.002 | 0.158 | 0.005 | 0.004 | 0.262 | 0.003 | 0.004 | 0.359 |
| Interleukin-6 receptor Antagonist | 0.004 | 0.002 | 0.020 | 0.005 | 0.002 | 0.002 | 0.012 | 0.004 | 0.004 | 0.010 | 0.004 | 0.004 |
| Interleukin-6 receptor | 0.002 | 0.001 | 0.222 | 0.000 | 0.001 | 0.911 | 0.003 | 0.004 | 0.388 | -0.001 | 0.003 | 0.784 |
| Low-density lipoprotein | 0.002 | 0.002 | 0.200 | -0.001 | 0.002 | 0.516 | -0.002 | 0.005 | 0.702 | 0.003 | 0.004 | 0.436 |
| LDL Receptor | -0.001 | 0.002 | 0.739 | 0.001 | 0.002 | 0.615 | 0.006 | 0.004 | 0.184 | -0.005 | 0.004 | 0.167 |
| Lipoprotein phospholipase A2-activity | 0.003 | 0.002 | 0.119 | 0.000 | 0.002 | 0.960 | -0.007 | 0.006 | 0.229 | 0.006 | 0.005 | 0.240 |
| Lipoprotein phospholipase A2-MASS | -0.002 | 0.002 | 0.399 | 0.003 | 0.002 | 0.157 | 0.010 | 0.005 | 0.069 | -0.004 | 0.005 | 0.408 |

| | | | | | | | | | | | | |
|---|---|---|---|---|---|---|---|---|---|---|---|---|
| Monocyte Chemoattractant Protein-1 | -0.001 | 0.001 | 0.304 | 0.000 | 0.001 | 0.981 | -0.003 | 0.003 | 0.412 | -0.001 | 0.003 | 0.661 |
| Lipoprotein a | 0.000 | 0.001 | 0.864 | -0.001 | 0.001 | 0.470 | 0.001 | 0.004 | 0.886 | 0.005 | 0.003 | 0.111 |
| Methylmalonic acid | 0.001 | 0.001 | 0.418 | -0.001 | 0.001 | 0.651 | -0.002 | 0.003 | 0.447 | -0.001 | 0.003 | 0.635 |
| Proprotein Convertase Subtilisin 9 | -0.001 | 0.001 | 0.347 | 0.001 | 0.002 | 0.440 | 0.002 | 0.004 | 0.604 | -0.001 | 0.003 | 0.814 |
| Triglyceride | -0.001 | 0.002 | 0.696 | -0.001 | 0.003 | 0.652 | -0.008 | 0.006 | 0.205 | 0.003 | 0.006 | 0.609 |
| Human chitinase 3-like protein 1 | 0.002 | 0.001 | 0.232 | -0.001 | 0.002 | 0.403 | -0.002 | 0.004 | 0.560 | 0.004 | 0.003 | 0.273 |
| Apo-A2 | -0.001 | 0.002 | 0.682 | -0.001 | 0.002 | 0.762 | 0.017 | 0.005 | 0.001 | 0.000 | 0.005 | 0.925 |
| Apo-C3 | 0.001 | 0.003 | 0.698 | 0.003 | 0.004 | 0.483 | -0.002 | 0.009 | 0.794 | 0.002 | 0.008 | 0.797 |
| Apo-E | -0.001 | 0.002 | 0.640 | -0.002 | 0.003 | 0.476 | 0.003 | 0.006 | 0.593 | -0.002 | 0.006 | 0.751 |
| Apo-A2 | 0.000 | 0.002 | 0.890 | -0.003 | 0.002 | 0.214 | -0.001 | 0.005 | 0.859 | -0.002 | 0.005 | 0.646 |
| Apo-B | -0.001 | 0.002 | 0.538 | 0.000 | 0.002 | 0.868 | -0.002 | 0.005 | 0.701 | -0.007 | 0.005 | 0.145 |
| Apo-C2 | -0.003 | 0.003 | 0.321 | 0.000 | 0.003 | 0.986 | -0.003 | 0.008 | 0.737 | -0.002 | 0.007 | 0.754 |
| Betaine | -0.003 | 0.002 | 0.100 | -0.004 | 0.002 | 0.097 | 0.002 | 0.005 | 0.690 | -0.010 | 0.005 | 0.042 |
| Trimethylamine oxide | -0.001 | 0.002 | 0.362 | -0.001 | 0.002 | 0.465 | 0.004 | 0.004 | 0.343 | -0.003 | 0.004 | 0.428 |
| Choline | -0.001 | 0.002 | 0.671 | 0.001 | 0.002 | 0.512 | -0.002 | 0.005 | 0.716 | 0.001 | 0.004 | 0.783 |
| Carnitine | 0.002 | 0.001 | 0.140 | 0.003 | 0.002 | 0.039 | 0.001 | 0.004 | 0.792 | 0.006 | 0.003 | 0.071 |
| Trimethylpentaaminovaleric acid | 0.000 | 0.001 | 0.764 | -0.001 | 0.001 | 0.531 | 0.001 | 0.003 | 0.799 | -0.001 | 0.003 | 0.660 |
| Butyl betaine | 0.000 | 0.001 | 0.948 | 0.000 | 0.001 | 0.970 | 0.002 | 0.004 | 0.595 | 0.000 | 0.003 | 0.983 |
| tml | 0.000 | 0.002 | 0.946 | -0.003 | 0.002 | 0.217 | 0.005 | 0.005 | 0.367 | -0.010 | 0.005 | 0.031 |

**Table S12c. Multi-variables regression of 38 biomarkers and Delta-NIHSS$_{(admission-discharge)}$**

| | Delta-NIH5B | | | Delta-NIH6A | | | Delta-NIH6B | | | Delta-NIH7 | | |
|---|---|---|---|---|---|---|---|---|---|---|---|---|
| variables | Estimate | SE | P-value | Estimate | SE | P-value | Estimate | SE | P-value | Estimate | SE | P-value |
| (Intercept) | 0.179 | 0.005 | 0.000 | 0.185 | 0.006 | 0.000 | 0.183 | 0.006 | 0.000 | 0.097 | 0.004 | 0.000 |
| Adiponectin | -0.004 | 0.006 | 0.499 | 0.002 | 0.006 | 0.712 | -0.007 | 0.006 | 0.244 | -0.002 | 0.004 | 0.613 |
| ANGPTL3 | -0.010 | 0.006 | 0.094 | -0.016 | 0.006 | 0.009 | -0.010 | 0.006 | 0.107 | -0.019 | 0.005 | 0.000 |
| Vitamin B12 | 0.010 | 0.006 | 0.079 | 0.007 | 0.006 | 0.194 | 0.004 | 0.006 | 0.525 | 0.002 | 0.004 | 0.620 |
| Folic acid | 0.012 | 0.007 | 0.080 | -0.005 | 0.007 | 0.463 | 0.012 | 0.007 | 0.078 | 0.004 | 0.005 | 0.430 |
| Creatinine | 0.008 | 0.010 | 0.401 | -0.036 | 0.010 | 0.000 | 0.003 | 0.010 | 0.725 | -0.004 | 0.008 | 0.577 |
| Cystatin C | 0.000 | 0.009 | 0.981 | 0.021 | 0.010 | 0.031 | 0.015 | 0.010 | 0.125 | -0.005 | 0.007 | 0.517 |
| D-dimer DD | 0.006 | 0.006 | 0.350 | 0.014 | 0.006 | 0.026 | 0.002 | 0.006 | 0.694 | 0.001 | 0.005 | 0.765 |
| Fibrinogen Fib | 0.014 | 0.006 | 0.023 | 0.004 | 0.006 | 0.504 | 0.017 | 0.006 | 0.007 | 0.007 | 0.005 | 0.128 |
| Fasting blood glucose | 0.008 | 0.006 | 0.171 | 0.023 | 0.006 | 0.000 | 0.012 | 0.006 | 0.039 | 0.008 | 0.005 | 0.083 |
| Homocysteine HCY | 0.003 | 0.006 | 0.555 | 0.002 | 0.006 | 0.757 | 0.000 | 0.006 | 0.995 | -0.001 | 0.005 | 0.770 |
| High-density lipoprotein | 0.003 | 0.007 | 0.661 | 0.001 | 0.007 | 0.940 | -0.001 | 0.007 | 0.855 | -0.003 | 0.005 | 0.610 |
| High-sensitivity C-reactive protein | 0.013 | 0.006 | 0.021 | 0.002 | 0.006 | 0.723 | 0.007 | 0.006 | 0.230 | 0.002 | 0.004 | 0.613 |
| Interleukin-1 Receptor Antagonist | 0.010 | 0.006 | 0.113 | 0.014 | 0.006 | 0.026 | 0.005 | 0.006 | 0.456 | 0.007 | 0.005 | 0.155 |
| Interleukin-6 receptor Antagonist | 0.020 | 0.006 | 0.002 | 0.035 | 0.007 | 0.000 | 0.018 | 0.007 | 0.006 | 0.014 | 0.005 | 0.006 |
| Interleukin-6 receptor | -0.002 | 0.006 | 0.730 | -0.008 | 0.006 | 0.128 | 0.000 | 0.006 | 0.932 | 0.006 | 0.004 | 0.144 |
| Low-density lipoprotein | -0.004 | 0.007 | 0.609 | 0.006 | 0.008 | 0.426 | -0.005 | 0.008 | 0.544 | 0.010 | 0.006 | 0.079 |
| LDL Receptor | 0.020 | 0.007 | 0.002 | -0.010 | 0.007 | 0.128 | 0.010 | 0.007 | 0.126 | -0.014 | 0.005 | 0.007 |
| Lipoprotein phospholipase A2-activity | -0.009 | 0.009 | 0.324 | -0.007 | 0.009 | 0.400 | -0.007 | 0.009 | 0.413 | -0.006 | 0.007 | 0.355 |
| Lipoprotein phospholipase A2-MASS | 0.007 | 0.008 | 0.386 | 0.004 | 0.008 | 0.624 | 0.002 | 0.008 | 0.776 | 0.000 | 0.007 | 0.945 |

| | | | | | | | | | | | | |
|---|---|---|---|---|---|---|---|---|---|---|---|---|
| Monocyte Chemoattractant Protein-1 | -0.004 | 0.005 | 0.420 | 0.000 | 0.005 | 0.942 | -0.003 | 0.005 | 0.616 | -0.006 | 0.004 | 0.141 |
| Lipoprotein a | 0.005 | 0.006 | 0.393 | -0.004 | 0.006 | 0.527 | 0.009 | 0.006 | 0.125 | 0.003 | 0.004 | 0.525 |
| Methylmalonic acid | 0.002 | 0.005 | 0.596 | -0.003 | 0.005 | 0.555 | 0.001 | 0.005 | 0.888 | 0.007 | 0.004 | 0.064 |
| Proprotein Convertase Subtilisin 9 | -0.022 | 0.006 | 0.000 | 0.003 | 0.006 | 0.667 | -0.017 | 0.006 | 0.004 | -0.003 | 0.005 | 0.439 |
| Triglyceride | -0.007 | 0.010 | 0.502 | -0.011 | 0.010 | 0.291 | -0.011 | 0.010 | 0.273 | -0.010 | 0.008 | 0.181 |
| Human chitinase 3-like protein 1 | 0.003 | 0.006 | 0.605 | 0.002 | 0.006 | 0.723 | 0.016 | 0.006 | 0.010 | -0.004 | 0.005 | 0.432 |
| Apo-A2 | -0.008 | 0.008 | 0.301 | -0.005 | 0.008 | 0.514 | -0.006 | 0.008 | 0.498 | -0.001 | 0.006 | 0.926 |
| Apo-C3 | -0.019 | 0.014 | 0.188 | 0.002 | 0.015 | 0.872 | -0.018 | 0.014 | 0.211 | -0.002 | 0.011 | 0.842 |
| Apo-E | 0.024 | 0.010 | 0.019 | -0.012 | 0.010 | 0.228 | 0.017 | 0.010 | 0.089 | 0.013 | 0.008 | 0.091 |
| Apo-A2 | 0.011 | 0.008 | 0.175 | 0.001 | 0.008 | 0.941 | 0.004 | 0.008 | 0.675 | -0.002 | 0.006 | 0.737 |
| Apo-B | 0.012 | 0.008 | 0.149 | 0.000 | 0.008 | 0.961 | 0.019 | 0.008 | 0.023 | -0.003 | 0.006 | 0.602 |
| Apo-C2 | -0.019 | 0.013 | 0.153 | 0.019 | 0.013 | 0.156 | -0.010 | 0.013 | 0.457 | 0.009 | 0.010 | 0.394 |
| Betaine | -0.019 | 0.008 | 0.025 | -0.021 | 0.009 | 0.012 | -0.015 | 0.009 | 0.088 | -0.009 | 0.007 | 0.176 |
| Trimethylamine oxide | -0.017 | 0.006 | 0.009 | -0.018 | 0.007 | 0.007 | -0.018 | 0.007 | 0.006 | 0.001 | 0.005 | 0.780 |
| Choline | 0.004 | 0.007 | 0.603 | 0.001 | 0.008 | 0.893 | -0.002 | 0.007 | 0.754 | 0.004 | 0.006 | 0.492 |
| Carnitine | 0.000 | 0.006 | 0.950 | -0.003 | 0.006 | 0.614 | 0.007 | 0.006 | 0.262 | 0.011 | 0.005 | 0.015 |
| Trimethylpentaaminovaleric acid | -0.004 | 0.005 | 0.390 | 0.004 | 0.005 | 0.457 | -0.005 | 0.005 | 0.257 | 0.002 | 0.004 | 0.583 |
| Butyl betaine | 0.010 | 0.006 | 0.070 | 0.005 | 0.006 | 0.329 | 0.005 | 0.006 | 0.354 | -0.003 | 0.004 | 0.503 |
| tml | -0.014 | 0.008 | 0.107 | 0.004 | 0.008 | 0.649 | -0.015 | 0.008 | 0.081 | -0.001 | 0.007 | 0.819 |

**Table S12d. Multi-variables regression of 38 biomarkers and Delta-NIHSS$_{(admission-discharge)}$**

| | Delta-NIH8 | | | Delta-NIH9 | | | Delta-NIH10 | | | Delta-NIH11 | | |
|---|---|---|---|---|---|---|---|---|---|---|---|---|
| variables | Estimate | SE | P-value | Estimate | SE | P-value | Estimate | SE | P-value | Estimate | SE | P-value |
| (Intercept) | 0.155 | 0.005 | 0.000 | 0.174 | 0.005 | 0.000 | 0.220 | 0.006 | 0.000 | 0.005 | 0.001 | 0.000 |
| Adiponectin | 0.003 | 0.005 | 0.616 | 0.000 | 0.005 | 0.996 | -0.009 | 0.006 | 0.115 | 0.000 | 0.001 | 0.895 |
| ANGPTL3 | 0.005 | 0.006 | 0.387 | -0.010 | 0.006 | 0.102 | -0.002 | 0.006 | 0.804 | 0.000 | 0.001 | 0.817 |
| Vitamin B12 | 0.018 | 0.005 | 0.001 | -0.002 | 0.006 | 0.756 | -0.006 | 0.006 | 0.308 | 0.000 | 0.001 | 0.738 |
| Folic acid | -0.010 | 0.006 | 0.142 | 0.000 | 0.007 | 0.966 | 0.016 | 0.007 | 0.034 | -0.001 | 0.001 | 0.305 |
| Creatinine | -0.005 | 0.009 | 0.601 | -0.005 | 0.010 | 0.629 | 0.005 | 0.011 | 0.615 | 0.002 | 0.002 | 0.318 |
| Cystatin C | -0.004 | 0.009 | 0.664 | -0.019 | 0.009 | 0.048 | -0.003 | 0.010 | 0.785 | -0.001 | 0.002 | 0.427 |
| D-dimer DD | 0.000 | 0.006 | 0.990 | 0.004 | 0.006 | 0.556 | -0.011 | 0.007 | 0.083 | 0.000 | 0.001 | 0.774 |
| Fibrinogen Fib | 0.000 | 0.006 | 0.984 | 0.006 | 0.006 | 0.354 | -0.016 | 0.007 | 0.017 | -0.002 | 0.001 | 0.124 |
| Fasting blood glucose | 0.011 | 0.006 | 0.037 | 0.002 | 0.006 | 0.749 | 0.000 | 0.006 | 0.992 | 0.003 | 0.001 | 0.004 |
| Homocysteine HCY | 0.002 | 0.006 | 0.736 | 0.008 | 0.006 | 0.153 | -0.014 | 0.006 | 0.028 | 0.000 | 0.001 | 0.936 |
| High-density lipoprotein | -0.007 | 0.007 | 0.287 | 0.011 | 0.007 | 0.131 | -0.003 | 0.008 | 0.701 | 0.000 | 0.001 | 0.989 |
| High-sensitivity C-reactive protein | 0.000 | 0.005 | 0.959 | 0.012 | 0.006 | 0.039 | 0.011 | 0.006 | 0.064 | 0.005 | 0.001 | 0.000 |
| Interleukin-1 Receptor Antagonist | 0.001 | 0.006 | 0.862 | -0.003 | 0.006 | 0.683 | -0.006 | 0.007 | 0.394 | 0.001 | 0.001 | 0.257 |
| Interleukin-6 receptor Antagonist | 0.008 | 0.006 | 0.188 | 0.026 | 0.006 | 0.000 | 0.027 | 0.007 | 0.000 | 0.000 | 0.001 | 0.854 |
| Interleukin-6 receptor | 0.009 | 0.005 | 0.085 | 0.004 | 0.005 | 0.463 | -0.007 | 0.006 | 0.262 | 0.001 | 0.001 | 0.184 |
| Low-density lipoprotein | 0.004 | 0.007 | 0.597 | 0.009 | 0.007 | 0.244 | 0.025 | 0.008 | 0.002 | 0.000 | 0.001 | 0.922 |
| LDL Receptor | -0.007 | 0.006 | 0.250 | -0.004 | 0.007 | 0.497 | -0.015 | 0.007 | 0.039 | 0.001 | 0.001 | 0.509 |
| Lipoprotein phospholipase A2-activity | -0.008 | 0.008 | 0.344 | -0.010 | 0.009 | 0.245 | 0.002 | 0.009 | 0.842 | 0.000 | 0.002 | 0.794 |
| Lipoprotein phospholipase A2-MASS | 0.008 | 0.008 | 0.333 | 0.005 | 0.008 | 0.558 | -0.011 | 0.009 | 0.209 | 0.000 | 0.002 | 0.859 |

| | | | | | | | | | | | | |
|---|---|---|---|---|---|---|---|---|---|---|---|---|
| Monocyte Chemoattractant Protein-1 | -0.011 | 0.005 | 0.039 | 0.000 | 0.005 | 0.958 | -0.007 | 0.006 | 0.262 | -0.001 | 0.001 | 0.480 |
| Lipoprotein a | 0.001 | 0.005 | 0.798 | -0.006 | 0.006 | 0.262 | 0.013 | 0.006 | 0.030 | 0.000 | 0.001 | 0.887 |
| Methylmalonic acid | 0.007 | 0.004 | 0.119 | 0.003 | 0.005 | 0.486 | 0.003 | 0.005 | 0.542 | 0.001 | 0.001 | 0.497 |
| Proprotein Convertase Subtilisin 9 | 0.007 | 0.006 | 0.192 | -0.020 | 0.006 | 0.000 | -0.017 | 0.006 | 0.006 | -0.001 | 0.001 | 0.446 |
| Triglyceride | 0.013 | 0.009 | 0.155 | 0.002 | 0.010 | 0.812 | 0.003 | 0.011 | 0.755 | -0.001 | 0.002 | 0.429 |
| Human chitinase 3-like protein 1 | -0.010 | 0.006 | 0.088 | 0.005 | 0.006 | 0.460 | 0.006 | 0.007 | 0.347 | 0.001 | 0.001 | 0.192 |
| Apo-A2 | -0.003 | 0.008 | 0.657 | -0.001 | 0.008 | 0.887 | -0.012 | 0.009 | 0.191 | 0.000 | 0.001 | 0.767 |
| Apo-C3 | -0.012 | 0.014 | 0.376 | -0.008 | 0.014 | 0.597 | -0.037 | 0.016 | 0.018 | 0.003 | 0.003 | 0.252 |
| Apo-E | 0.010 | 0.010 | 0.290 | 0.018 | 0.010 | 0.067 | 0.012 | 0.011 | 0.264 | -0.001 | 0.002 | 0.672 |
| Apo-A2 | 0.011 | 0.008 | 0.176 | -0.013 | 0.008 | 0.127 | 0.001 | 0.009 | 0.952 | 0.001 | 0.002 | 0.698 |
| Apo-B | -0.008 | 0.008 | 0.297 | 0.019 | 0.008 | 0.019 | -0.007 | 0.009 | 0.410 | 0.000 | 0.001 | 0.873 |
| Apo-C2 | -0.003 | 0.013 | 0.788 | -0.011 | 0.013 | 0.386 | 0.027 | 0.014 | 0.056 | -0.004 | 0.002 | 0.108 |
| Betaine | -0.012 | 0.008 | 0.137 | 0.006 | 0.008 | 0.449 | -0.026 | 0.009 | 0.004 | 0.002 | 0.002 | 0.169 |
| Trimethylamine oxide | -0.007 | 0.006 | 0.246 | -0.007 | 0.006 | 0.302 | -0.011 | 0.007 | 0.112 | 0.000 | 0.001 | 0.883 |
| Choline | 0.000 | 0.007 | 0.949 | -0.003 | 0.007 | 0.659 | -0.001 | 0.008 | 0.895 | -0.002 | 0.001 | 0.157 |
| Carnitine | 0.003 | 0.006 | 0.553 | -0.002 | 0.006 | 0.760 | 0.004 | 0.006 | 0.558 | 0.000 | 0.001 | 0.866 |
| Trimethylpentaaminovaleric acid | -0.003 | 0.004 | 0.530 | -0.002 | 0.005 | 0.672 | 0.001 | 0.005 | 0.828 | 0.000 | 0.001 | 0.879 |
| Butyl betaine | 0.010 | 0.005 | 0.052 | 0.001 | 0.006 | 0.800 | 0.002 | 0.006 | 0.785 | 0.000 | 0.001 | 0.842 |
| tml | -0.007 | 0.008 | 0.384 | -0.004 | 0.008 | 0.599 | 0.003 | 0.009 | 0.751 | 0.001 | 0.002 | 0.683 |

**Table S13a. Multi-variables regression of 38 biomarkers and Delta-NIHSS$_{(admission-discharge)}$ in scenario 1**

| | Delta-NIHSS | | | Delta-NIH1A | | | Delta-NIH1B | | | Delta-NIH1C | | |
|---|---|---|---|---|---|---|---|---|---|---|---|---|
| variables | Estimate | SE | P-value | Estimate | SE | P-value | Estimate | SE | P-value | Estimate | SE | P-value |
| (Intercept) | 0.084 | 0.005 | 0.000 | 0.013 | 0.002 | 0.000 | 0.013 | 0.002 | 0.000 | 0.013 | 0.002 | 0.000 |
| Adiponectin | -0.001 | 0.005 | 0.779 | 0.001 | 0.002 | 0.603 | 0.006 | 0.002 | 0.008 | 0.001 | 0.002 | 0.605 |
| ANGPTL3 | -0.006 | 0.006 | 0.302 | -0.003 | 0.002 | 0.193 | -0.002 | 0.002 | 0.337 | 0.001 | 0.002 | 0.598 |
| Vitamin B12 | 0.008 | 0.005 | 0.148 | 0.004 | 0.002 | 0.109 | 0.005 | 0.002 | 0.018 | 0.003 | 0.002 | 0.126 |
| Folic acid | 0.003 | 0.007 | 0.718 | -0.002 | 0.003 | 0.387 | 0.004 | 0.003 | 0.130 | -0.001 | 0.003 | 0.750 |
| Creatinine | -0.007 | 0.009 | 0.466 | -0.002 | 0.004 | 0.520 | -0.005 | 0.004 | 0.193 | 0.001 | 0.004 | 0.845 |
| Cystatin C | -0.005 | 0.009 | 0.585 | -0.003 | 0.004 | 0.419 | -0.003 | 0.004 | 0.480 | -0.003 | 0.004 | 0.469 |
| D-dimer DD | 0.005 | 0.006 | 0.426 | 0.001 | 0.003 | 0.634 | 0.005 | 0.003 | 0.073 | -0.003 | 0.003 | 0.328 |
| Fibrinogen Fib | -0.001 | 0.006 | 0.871 | 0.001 | 0.002 | 0.836 | -0.001 | 0.002 | 0.711 | -0.005 | 0.002 | 0.026 |
| Fasting blood glucose | 0.011 | 0.005 | 0.046 | 0.006 | 0.002 | 0.012 | 0.006 | 0.002 | 0.003 | 0.002 | 0.002 | 0.258 |
| Homocysteine HCY | 0.006 | 0.006 | 0.261 | 0.000 | 0.002 | 0.932 | 0.002 | 0.002 | 0.470 | 0.002 | 0.002 | 0.382 |
| High-density lipoprotein | -0.010 | 0.007 | 0.114 | -0.001 | 0.003 | 0.846 | -0.002 | 0.003 | 0.477 | 0.002 | 0.003 | 0.502 |
| High-sensitivity C-reactive protein | 0.007 | 0.006 | 0.225 | 0.004 | 0.002 | 0.086 | 0.002 | 0.002 | 0.488 | 0.001 | 0.002 | 0.612 |
| Interleukin-1 Receptor Antagonist | 0.007 | 0.006 | 0.267 | 0.001 | 0.003 | 0.735 | 0.003 | 0.003 | 0.299 | 0.002 | 0.003 | 0.399 |
| Interleukin-6 receptor Antagonist | 0.016 | 0.006 | 0.011 | 0.008 | 0.003 | 0.003 | 0.002 | 0.003 | 0.435 | 0.006 | 0.003 | 0.018 |
| Interleukin-6 receptor | -0.005 | 0.005 | 0.370 | -0.001 | 0.002 | 0.674 | 0.000 | 0.002 | 0.961 | 0.003 | 0.002 | 0.125 |
| Low-density lipoprotein | 0.004 | 0.007 | 0.597 | 0.005 | 0.003 | 0.110 | 0.003 | 0.003 | 0.226 | -0.002 | 0.003 | 0.474 |
| LDL Receptor | -0.006 | 0.006 | 0.316 | -0.003 | 0.003 | 0.196 | -0.004 | 0.003 | 0.095 | 0.002 | 0.003 | 0.550 |
| Lipoprotein phospholipase A2-activity | 0.000 | 0.008 | 0.980 | 0.004 | 0.003 | 0.293 | 0.000 | 0.003 | 0.924 | 0.001 | 0.003 | 0.694 |
| Lipoprotein phospholipase A2-MASS | 0.012 | 0.008 | 0.125 | -0.002 | 0.003 | 0.613 | 0.000 | 0.003 | 0.905 | 0.003 | 0.003 | 0.421 |

| | | | | | | | | | | | | |
|---|---|---|---|---|---|---|---|---|---|---|---|---|
| Monocyte Chemoattractant Protein-1 | -0.001 | 0.004 | 0.855 | 0.005 | 0.002 | 0.002 | 0.000 | 0.002 | 0.957 | -0.001 | 0.002 | 0.754 |
| Lipoprotein a | -0.002 | 0.006 | 0.771 | -0.003 | 0.002 | 0.181 | -0.003 | 0.002 | 0.215 | 0.001 | 0.002 | 0.811 |
| Methylmalonic acid | 0.002 | 0.004 | 0.673 | 0.001 | 0.002 | 0.477 | 0.001 | 0.002 | 0.425 | 0.001 | 0.002 | 0.417 |
| Proprotein Convertase Subtilisin 9 | 0.004 | 0.005 | 0.488 | 0.002 | 0.002 | 0.352 | 0.000 | 0.002 | 0.874 | -0.001 | 0.002 | 0.675 |
| Triglyceride | -0.010 | 0.009 | 0.283 | -0.002 | 0.004 | 0.687 | -0.002 | 0.004 | 0.587 | -0.006 | 0.004 | 0.103 |
| Human chitinase 3-like protein 1 | 0.005 | 0.006 | 0.385 | 0.002 | 0.002 | 0.342 | 0.000 | 0.002 | 0.959 | 0.003 | 0.002 | 0.166 |
| Apo-A2 | -0.005 | 0.008 | 0.480 | 0.001 | 0.003 | 0.855 | -0.001 | 0.003 | 0.829 | -0.001 | 0.003 | 0.736 |
| Apo-C3 | 0.004 | 0.014 | 0.777 | 0.002 | 0.006 | 0.765 | -0.002 | 0.006 | 0.679 | -0.006 | 0.006 | 0.257 |
| Apo-E | 0.005 | 0.009 | 0.573 | 0.006 | 0.004 | 0.120 | 0.007 | 0.004 | 0.093 | 0.003 | 0.004 | 0.382 |
| Apo-A2 | 0.002 | 0.008 | 0.767 | 0.001 | 0.003 | 0.717 | 0.001 | 0.003 | 0.809 | 0.001 | 0.003 | 0.807 |
| Apo-B | 0.000 | 0.008 | 0.992 | -0.005 | 0.003 | 0.150 | 0.000 | 0.003 | 0.925 | -0.001 | 0.003 | 0.696 |
| Apo-C2 | -0.013 | 0.013 | 0.308 | -0.008 | 0.005 | 0.124 | -0.002 | 0.005 | 0.694 | 0.000 | 0.005 | 0.955 |
| Betaine | -0.003 | 0.006 | 0.598 | -0.002 | 0.002 | 0.307 | -0.002 | 0.002 | 0.368 | -0.006 | 0.002 | 0.010 |
| Trimethylamine oxide | 0.000 | 0.007 | 0.956 | -0.001 | 0.003 | 0.707 | 0.004 | 0.003 | 0.103 | 0.000 | 0.003 | 0.936 |
| Choline | 0.005 | 0.007 | 0.467 | -0.001 | 0.003 | 0.730 | -0.002 | 0.003 | 0.480 | 0.000 | 0.003 | 0.896 |
| Carnitine | 0.011 | 0.006 | 0.047 | 0.004 | 0.002 | 0.132 | 0.006 | 0.002 | 0.012 | 0.003 | 0.002 | 0.142 |
| Trimethylpentaaminovaleric acid | -0.001 | 0.004 | 0.747 | -0.001 | 0.002 | 0.695 | 0.000 | 0.002 | 0.969 | -0.001 | 0.002 | 0.581 |
| Butyl betaine | 0.002 | 0.005 | 0.643 | 0.002 | 0.002 | 0.447 | 0.000 | 0.002 | 0.824 | 0.002 | 0.002 | 0.352 |
| Trimethyllysine | -0.006 | 0.009 | 0.482 | 0.001 | 0.004 | 0.731 | 0.003 | 0.004 | 0.383 | 0.000 | 0.004 | 0.923 |

**Table S13b. Multi-variables regression of 38 biomarkers and Delta-NIHSS$_{(admission-discharge)}$ in scenario 1**

| | Delta-NIH2 | | | Delta-NIH3 | | | Delta-NIH4 | | | Delta-NIH5A | | |
|---|---|---|---|---|---|---|---|---|---|---|---|---|
| variables | Estimate | SE | P-value | Estimate | SE | P-value | Estimate | SE | P-value | Estimate | SE | P-value |
| (Intercept) | 0.009 | 0.002 | 0.000 | 0.007 | 0.002 | 0.000 | 0.059 | 0.004 | 0.000 | 0.039 | 0.004 | 0.000 |
| Adiponectin | -0.001 | 0.002 | 0.428 | 0.000 | 0.002 | 0.842 | -0.003 | 0.004 | 0.471 | -0.002 | 0.004 | 0.568 |
| ANGPTL3 | -0.001 | 0.002 | 0.749 | -0.001 | 0.002 | 0.727 | 0.005 | 0.005 | 0.317 | -0.002 | 0.004 | 0.599 |
| Vitamin B12 | 0.001 | 0.002 | 0.442 | 0.002 | 0.002 | 0.147 | 0.002 | 0.004 | 0.619 | 0.003 | 0.004 | 0.481 |
| Folic acid | 0.000 | 0.002 | 0.990 | -0.001 | 0.002 | 0.622 | 0.005 | 0.006 | 0.392 | 0.001 | 0.005 | 0.778 |
| Creatinine | -0.001 | 0.003 | 0.830 | 0.003 | 0.003 | 0.374 | 0.001 | 0.008 | 0.864 | -0.007 | 0.006 | 0.306 |
| Cystatin C | -0.004 | 0.003 | 0.247 | -0.003 | 0.003 | 0.337 | -0.007 | 0.008 | 0.345 | 0.001 | 0.006 | 0.861 |
| D-dimer DD | -0.001 | 0.002 | 0.734 | 0.000 | 0.002 | 0.961 | 0.004 | 0.005 | 0.486 | 0.009 | 0.004 | 0.039 |
| Fibrinogen Fib | 0.000 | 0.002 | 0.875 | 0.000 | 0.002 | 0.821 | 0.008 | 0.005 | 0.133 | -0.008 | 0.004 | 0.058 |
| Fasting blood glucose | 0.002 | 0.002 | 0.169 | 0.003 | 0.002 | 0.045 | -0.002 | 0.004 | 0.607 | 0.004 | 0.004 | 0.245 |
| Homocysteine HCY | 0.000 | 0.002 | 0.919 | 0.003 | 0.002 | 0.064 | -0.003 | 0.005 | 0.564 | 0.004 | 0.004 | 0.276 |
| High-density lipoprotein | -0.001 | 0.002 | 0.625 | 0.001 | 0.002 | 0.639 | -0.003 | 0.006 | 0.564 | -0.003 | 0.005 | 0.582 |
| High-sensitivity C-reactive protein | -0.001 | 0.002 | 0.599 | -0.001 | 0.002 | 0.445 | -0.003 | 0.005 | 0.581 | -0.005 | 0.004 | 0.213 |
| Interleukin-1 Receptor Antagonist | 0.004 | 0.002 | 0.085 | -0.003 | 0.002 | 0.173 | 0.012 | 0.005 | 0.018 | -0.002 | 0.004 | 0.642 |
| Interleukin-6 receptor Antagonist | 0.004 | 0.002 | 0.067 | 0.004 | 0.002 | 0.050 | 0.014 | 0.005 | 0.007 | 0.013 | 0.004 | 0.002 |
| Interleukin-6 receptor | 0.004 | 0.002 | 0.039 | 0.001 | 0.002 | 0.634 | 0.007 | 0.004 | 0.095 | -0.001 | 0.004 | 0.705 |
| Low-density lipoprotein | 0.004 | 0.002 | 0.093 | 0.000 | 0.002 | 0.915 | 0.004 | 0.006 | 0.474 | 0.007 | 0.005 | 0.152 |
| LDL Receptor | -0.001 | 0.002 | 0.745 | 0.003 | 0.002 | 0.197 | 0.003 | 0.005 | 0.520 | -0.007 | 0.004 | 0.106 |
| Lipoprotein phospholipase A2-activity | 0.006 | 0.003 | 0.043 | -0.003 | 0.003 | 0.239 | -0.005 | 0.007 | 0.462 | 0.006 | 0.006 | 0.283 |
| Lipoprotein phospholipase A2-MASS | -0.003 | 0.003 | 0.236 | 0.005 | 0.002 | 0.043 | 0.010 | 0.007 | 0.134 | -0.005 | 0.005 | 0.404 |

| | | | | | | | | | | | | |
|---|---|---|---|---|---|---|---|---|---|---|---|---|
| Monocyte Chemoattractant Protein-1 | -0.001 | 0.001 | 0.646 | 0.000 | 0.001 | 0.954 | -0.002 | 0.004 | 0.492 | 0.000 | 0.003 | 0.897 |
| Lipoprotein a | 0.000 | 0.002 | 0.848 | -0.003 | 0.002 | 0.089 | -0.002 | 0.005 | 0.707 | 0.005 | 0.004 | 0.238 |
| Methylmalonic acid | 0.002 | 0.001 | 0.265 | 0.000 | 0.001 | 0.882 | -0.001 | 0.004 | 0.694 | -0.001 | 0.003 | 0.805 |
| Proprotein Convertase Subtilisin 9 | -0.002 | 0.002 | 0.256 | -0.002 | 0.002 | 0.147 | 0.009 | 0.005 | 0.058 | 0.004 | 0.004 | 0.343 |
| Triglyceride | -0.001 | 0.003 | 0.647 | -0.003 | 0.003 | 0.308 | -0.006 | 0.008 | 0.408 | 0.000 | 0.006 | 0.998 |
| Human chitinase 3-like protein 1 | 0.001 | 0.002 | 0.678 | -0.001 | 0.002 | 0.479 | -0.006 | 0.005 | 0.188 | 0.007 | 0.004 | 0.103 |
| Apo-A2 | -0.001 | 0.003 | 0.799 | -0.001 | 0.002 | 0.721 | 0.022 | 0.006 | 0.001 | 0.001 | 0.005 | 0.807 |
| Apo-C3 | 0.002 | 0.005 | 0.602 | -0.001 | 0.004 | 0.819 | -0.017 | 0.012 | 0.137 | 0.006 | 0.010 | 0.511 |
| Apo-E | -0.001 | 0.003 | 0.712 | 0.002 | 0.003 | 0.552 | 0.010 | 0.008 | 0.213 | 0.007 | 0.007 | 0.298 |
| Apo-A2 | 0.000 | 0.003 | 0.881 | -0.001 | 0.002 | 0.732 | -0.003 | 0.007 | 0.661 | -0.008 | 0.006 | 0.162 |
| Apo-B | -0.002 | 0.003 | 0.337 | 0.001 | 0.002 | 0.776 | -0.008 | 0.006 | 0.196 | -0.007 | 0.005 | 0.220 |
| Apo-C2 | -0.004 | 0.004 | 0.312 | 0.002 | 0.004 | 0.592 | 0.008 | 0.011 | 0.446 | -0.010 | 0.009 | 0.277 |
| Betaine | -0.005 | 0.002 | 0.007 | -0.002 | 0.002 | 0.294 | 0.002 | 0.005 | 0.726 | -0.005 | 0.004 | 0.174 |
| Trimethylamine oxide | -0.001 | 0.002 | 0.602 | 0.000 | 0.002 | 0.906 | 0.005 | 0.006 | 0.340 | -0.002 | 0.005 | 0.689 |
| Choline | 0.003 | 0.002 | 0.180 | 0.001 | 0.002 | 0.734 | -0.002 | 0.006 | 0.683 | 0.001 | 0.005 | 0.765 |
| Carnitine | 0.000 | 0.002 | 0.923 | 0.004 | 0.002 | 0.012 | -0.001 | 0.005 | 0.819 | 0.003 | 0.004 | 0.415 |
| Trimethylpentaaminovaleric acid | -0.001 | 0.001 | 0.667 | -0.001 | 0.001 | 0.592 | 0.001 | 0.003 | 0.807 | -0.002 | 0.003 | 0.491 |
| Butyl betaine | 0.000 | 0.002 | 0.951 | -0.001 | 0.002 | 0.541 | 0.000 | 0.005 | 0.929 | -0.002 | 0.004 | 0.583 |
| Trimethyllysine | -0.001 | 0.003 | 0.842 | -0.003 | 0.003 | 0.235 | 0.002 | 0.007 | 0.786 | -0.011 | 0.006 | 0.080 |

**Table S13c. Multi-variables regression of 38 biomarkers and Delta-NIHSS$_{(admission-discharge)}$ in scenario 1**

| | Delta-NIH5B | | | Delta-NIH6A | | | Delta-NIH6B | | | Delta-NIH7 | | |
|---|---|---|---|---|---|---|---|---|---|---|---|---|
| variables | Estimate | SE | P-value | Estimate | SE | P-value | Estimate | SE | P-value | Estimate | SE | P-value |
| (Intercept) | 0.157 | 0.007 | 0.000 | 0.161 | 0.007 | 0.000 | 0.161 | 0.007 | 0.000 | 0.085 | 0.005 | 0.000 |
| Adiponectin | -0.002 | 0.007 | 0.801 | 0.001 | 0.007 | 0.909 | -0.006 | 0.007 | 0.351 | 0.002 | 0.005 | 0.742 |
| ANGPTL3 | -0.016 | 0.007 | 0.026 | -0.012 | 0.007 | 0.108 | -0.012 | 0.007 | 0.106 | -0.023 | 0.006 | 0.000 |
| Vitamin B12 | 0.014 | 0.007 | 0.039 | 0.001 | 0.007 | 0.837 | 0.007 | 0.007 | 0.335 | 0.000 | 0.005 | 0.954 |
| Folic acid | 0.019 | 0.009 | 0.041 | 0.001 | 0.009 | 0.930 | 0.012 | 0.009 | 0.202 | -0.003 | 0.007 | 0.677 |
| Creatinine | -0.004 | 0.012 | 0.724 | -0.033 | 0.012 | 0.008 | -0.003 | 0.012 | 0.779 | -0.020 | 0.009 | 0.030 |
| Cystatin C | 0.011 | 0.012 | 0.339 | 0.011 | 0.012 | 0.362 | 0.024 | 0.012 | 0.042 | 0.013 | 0.009 | 0.142 |
| D-dimer DD | 0.005 | 0.008 | 0.543 | 0.000 | 0.008 | 0.998 | -0.001 | 0.008 | 0.947 | -0.002 | 0.006 | 0.736 |
| Fibrinogen Fib | 0.004 | 0.008 | 0.624 | -0.006 | 0.008 | 0.440 | 0.003 | 0.008 | 0.743 | 0.003 | 0.006 | 0.592 |
| Fasting blood glucose | -0.001 | 0.007 | 0.867 | 0.023 | 0.007 | 0.001 | 0.005 | 0.007 | 0.475 | 0.012 | 0.005 | 0.027 |
| Homocysteine HCY | 0.002 | 0.007 | 0.797 | 0.013 | 0.007 | 0.072 | -0.009 | 0.007 | 0.209 | 0.004 | 0.006 | 0.459 |
| High-density lipoprotein | -0.007 | 0.009 | 0.388 | -0.006 | 0.009 | 0.481 | -0.008 | 0.009 | 0.354 | -0.002 | 0.007 | 0.814 |
| High-sensitivity C-reactive protein | 0.021 | 0.007 | 0.006 | 0.002 | 0.008 | 0.797 | 0.010 | 0.008 | 0.171 | 0.000 | 0.006 | 0.937 |
| Interleukin-1 Receptor Antagonist | 0.000 | 0.008 | 0.965 | 0.010 | 0.008 | 0.232 | -0.001 | 0.008 | 0.940 | 0.008 | 0.006 | 0.175 |
| Interleukin-6 receptor Antagonist | 0.014 | 0.008 | 0.094 | 0.033 | 0.008 | 0.000 | 0.021 | 0.008 | 0.010 | 0.023 | 0.006 | 0.000 |
| Interleukin-6 receptor | 0.004 | 0.007 | 0.511 | -0.009 | 0.007 | 0.171 | 0.007 | 0.007 | 0.305 | 0.006 | 0.005 | 0.275 |
| Low-density lipoprotein | 0.004 | 0.009 | 0.662 | 0.018 | 0.009 | 0.052 | -0.003 | 0.009 | 0.760 | 0.015 | 0.007 | 0.029 |
| LDL Receptor | 0.015 | 0.008 | 0.073 | -0.012 | 0.008 | 0.154 | 0.011 | 0.008 | 0.209 | -0.007 | 0.006 | 0.308 |
| Lipoprotein phospholipase A2-activity | -0.013 | 0.011 | 0.228 | -0.013 | 0.011 | 0.232 | -0.010 | 0.011 | 0.364 | -0.003 | 0.008 | 0.719 |
| Lipoprotein phospholipase A2-MASS | 0.003 | 0.010 | 0.734 | 0.009 | 0.010 | 0.408 | -0.001 | 0.010 | 0.899 | -0.009 | 0.008 | 0.253 |

| | | | | | | | | | | | | |
|---|---|---|---|---|---|---|---|---|---|---|---|---|
| Monocyte Chemoattractant Protein-1 | -0.004 | 0.006 | 0.458 | -0.001 | 0.006 | 0.853 | -0.002 | 0.006 | 0.693 | -0.005 | 0.004 | 0.218 |
| Lipoprotein a | 0.003 | 0.007 | 0.629 | -0.006 | 0.007 | 0.413 | 0.008 | 0.007 | 0.295 | 0.001 | 0.006 | 0.921 |
| Methylmalonic acid | 0.002 | 0.006 | 0.777 | -0.005 | 0.006 | 0.352 | 0.002 | 0.006 | 0.770 | 0.000 | 0.004 | 0.957 |
| Proprotein Convertase Subtilisin 9 | -0.018 | 0.007 | 0.011 | -0.002 | 0.007 | 0.798 | -0.013 | 0.007 | 0.061 | -0.002 | 0.005 | 0.669 |
| Triglyceride | -0.006 | 0.012 | 0.632 | -0.014 | 0.012 | 0.248 | 0.000 | 0.012 | 0.974 | 0.000 | 0.009 | 0.958 |
| Human chitinase 3-like protein 1 | 0.000 | 0.008 | 0.983 | 0.007 | 0.008 | 0.381 | 0.009 | 0.008 | 0.256 | -0.010 | 0.006 | 0.088 |
| Apo-A2 | -0.008 | 0.010 | 0.421 | -0.002 | 0.010 | 0.873 | -0.007 | 0.010 | 0.479 | -0.003 | 0.008 | 0.724 |
| Apo-C3 | -0.008 | 0.018 | 0.670 | 0.006 | 0.018 | 0.730 | -0.026 | 0.018 | 0.148 | -0.007 | 0.014 | 0.618 |
| Apo-E | 0.014 | 0.012 | 0.259 | 0.007 | 0.012 | 0.560 | 0.018 | 0.012 | 0.141 | 0.016 | 0.009 | 0.100 |
| Apo-A2 | 0.007 | 0.010 | 0.507 | -0.008 | 0.010 | 0.421 | 0.011 | 0.010 | 0.289 | 0.003 | 0.008 | 0.708 |
| Apo-B | 0.010 | 0.010 | 0.311 | -0.012 | 0.010 | 0.241 | 0.018 | 0.010 | 0.077 | -0.005 | 0.008 | 0.528 |
| Apo-C2 | -0.018 | 0.017 | 0.274 | 0.008 | 0.017 | 0.641 | -0.012 | 0.017 | 0.454 | 0.002 | 0.013 | 0.846 |
| Betaine | -0.013 | 0.007 | 0.074 | -0.005 | 0.007 | 0.477 | -0.014 | 0.007 | 0.051 | 0.002 | 0.006 | 0.661 |
| Trimethylamine oxide | -0.015 | 0.009 | 0.081 | -0.012 | 0.009 | 0.173 | -0.019 | 0.009 | 0.033 | 0.004 | 0.007 | 0.526 |
| Choline | 0.016 | 0.009 | 0.083 | -0.001 | 0.009 | 0.954 | 0.006 | 0.009 | 0.521 | -0.001 | 0.007 | 0.865 |
| Carnitine | -0.003 | 0.007 | 0.725 | -0.006 | 0.007 | 0.428 | 0.000 | 0.007 | 1.000 | 0.008 | 0.006 | 0.165 |
| Trimethylpentaaminovaleric acid | -0.004 | 0.005 | 0.452 | 0.006 | 0.005 | 0.263 | -0.005 | 0.005 | 0.312 | 0.001 | 0.004 | 0.780 |
| Butyl betaine | 0.003 | 0.007 | 0.620 | 0.010 | 0.007 | 0.176 | 0.013 | 0.007 | 0.072 | -0.003 | 0.005 | 0.571 |
| Trimethyllysine | -0.012 | 0.011 | 0.295 | -0.002 | 0.012 | 0.852 | -0.021 | 0.012 | 0.071 | -0.002 | 0.009 | 0.832 |

**Table S13d. Multi-variables regression of 38 biomarkers and Delta-NIHSS$_{(admission-discharge)}$ in scenario 1**

| | Delta-NIH8 | | | Delta-NIH9 | | | Delta-NIH10 | | | Delta-NIH11 | | |
|---|---|---|---|---|---|---|---|---|---|---|---|---|
| variables | Estimate | SE | P-value | Estimate | SE | P-value | Estimate | SE | P-value | Estimate | SE | P-value |
| (Intercept) | 0.143 | 0.007 | 0.000 | 0.164 | 0.007 | 0.000 | 0.206 | 0.008 | 0.000 | 0.004 | 0.001 | 0.001 |
| Adiponectin | 0.005 | 0.007 | 0.415 | -0.006 | 0.007 | 0.423 | -0.008 | 0.008 | 0.273 | -0.001 | 0.001 | 0.305 |
| ANGPTL3 | 0.003 | 0.007 | 0.642 | -0.011 | 0.007 | 0.147 | 0.001 | 0.008 | 0.936 | 0.000 | 0.001 | 0.971 |
| Vitamin B12 | 0.023 | 0.007 | 0.001 | -0.005 | 0.007 | 0.464 | -0.010 | 0.008 | 0.192 | 0.001 | 0.001 | 0.612 |
| Folic acid | -0.019 | 0.009 | 0.029 | 0.000 | 0.009 | 0.972 | 0.014 | 0.010 | 0.158 | -0.002 | 0.002 | 0.222 |
| Creatinine | -0.015 | 0.012 | 0.202 | -0.017 | 0.012 | 0.153 | 0.002 | 0.013 | 0.857 | 0.002 | 0.002 | 0.422 |
| Cystatin C | 0.011 | 0.012 | 0.328 | -0.012 | 0.012 | 0.316 | -0.010 | 0.013 | 0.431 | -0.002 | 0.002 | 0.306 |
| D-dimer DD | -0.014 | 0.008 | 0.084 | 0.000 | 0.008 | 0.995 | -0.017 | 0.009 | 0.070 | -0.001 | 0.001 | 0.652 |
| Fibrinogen Fib | -0.005 | 0.008 | 0.473 | 0.004 | 0.008 | 0.593 | -0.023 | 0.009 | 0.010 | -0.001 | 0.001 | 0.478 |
| Fasting blood glucose | 0.012 | 0.007 | 0.070 | -0.002 | 0.007 | 0.753 | 0.005 | 0.008 | 0.534 | 0.001 | 0.001 | 0.317 |
| Homocysteine HCY | 0.003 | 0.007 | 0.668 | 0.003 | 0.007 | 0.681 | -0.018 | 0.008 | 0.033 | -0.001 | 0.001 | 0.272 |
| High-density lipoprotein | 0.004 | 0.008 | 0.653 | 0.012 | 0.009 | 0.155 | -0.004 | 0.010 | 0.683 | 0.001 | 0.002 | 0.360 |
| High-sensitivity C-reactive protein | 0.005 | 0.007 | 0.509 | 0.007 | 0.008 | 0.342 | 0.012 | 0.008 | 0.142 | 0.001 | 0.001 | 0.544 |
| Interleukin-1 Receptor Antagonist | -0.001 | 0.008 | 0.889 | 0.002 | 0.008 | 0.780 | 0.002 | 0.009 | 0.792 | 0.000 | 0.001 | 0.897 |
| Interleukin-6 receptor Antagonist | 0.013 | 0.008 | 0.094 | 0.031 | 0.008 | 0.000 | 0.018 | 0.009 | 0.051 | 0.001 | 0.001 | 0.449 |
| Interleukin-6 receptor | 0.009 | 0.007 | 0.161 | 0.014 | 0.007 | 0.047 | -0.002 | 0.008 | 0.814 | 0.001 | 0.001 | 0.339 |
| Low-density lipoprotein | 0.012 | 0.009 | 0.183 | 0.007 | 0.009 | 0.430 | 0.030 | 0.010 | 0.003 | 0.000 | 0.002 | 0.945 |
| LDL Receptor | -0.002 | 0.008 | 0.841 | -0.008 | 0.008 | 0.348 | -0.015 | 0.009 | 0.101 | 0.000 | 0.001 | 0.939 |
| Lipoprotein phospholipase A2-activity | 0.002 | 0.010 | 0.880 | -0.021 | 0.011 | 0.055 | -0.002 | 0.012 | 0.879 | 0.002 | 0.002 | 0.287 |
| Lipoprotein phospholipase A2-MASS | 0.002 | 0.010 | 0.839 | 0.006 | 0.010 | 0.575 | -0.004 | 0.011 | 0.757 | -0.002 | 0.002 | 0.291 |

| | | | | | | | | | | | | |
|---|---|---|---|---|---|---|---|---|---|---|---|---|
| Monocyte Chemoattractant Protein-1 | -0.010 | 0.005 | 0.082 | 0.003 | 0.006 | 0.626 | -0.006 | 0.006 | 0.317 | 0.000 | 0.001 | 0.650 |
| Lipoprotein a | -0.005 | 0.007 | 0.504 | -0.001 | 0.007 | 0.855 | 0.013 | 0.008 | 0.117 | 0.001 | 0.001 | 0.344 |
| Methylmalonic acid | 0.013 | 0.005 | 0.017 | 0.001 | 0.006 | 0.797 | 0.000 | 0.006 | 0.949 | 0.001 | 0.001 | 0.423 |
| Proprotein Convertase Subtilisin 9 | 0.013 | 0.007 | 0.057 | -0.023 | 0.007 | 0.001 | -0.019 | 0.008 | 0.020 | -0.001 | 0.001 | 0.479 |
| Triglyceride | 0.005 | 0.012 | 0.674 | 0.016 | 0.012 | 0.181 | 0.013 | 0.013 | 0.335 | -0.001 | 0.002 | 0.805 |
| Human chitinase 3-like protein 1 | -0.010 | 0.007 | 0.168 | 0.002 | 0.008 | 0.763 | 0.009 | 0.009 | 0.303 | 0.002 | 0.001 | 0.098 |
| Apo-A2 | -0.006 | 0.010 | 0.522 | -0.006 | 0.010 | 0.559 | -0.013 | 0.011 | 0.237 | 0.000 | 0.002 | 0.877 |
| Apo-C3 | 0.000 | 0.018 | 0.985 | -0.036 | 0.018 | 0.050 | -0.054 | 0.020 | 0.007 | 0.001 | 0.003 | 0.695 |
| Apo-E | 0.014 | 0.012 | 0.249 | 0.026 | 0.013 | 0.035 | 0.020 | 0.014 | 0.148 | 0.002 | 0.002 | 0.337 |
| Apo-A2 | 0.005 | 0.010 | 0.598 | -0.013 | 0.010 | 0.218 | 0.010 | 0.012 | 0.386 | 0.000 | 0.002 | 0.845 |
| Apo-B | -0.006 | 0.010 | 0.534 | 0.025 | 0.010 | 0.012 | -0.004 | 0.011 | 0.703 | 0.000 | 0.002 | 0.796 |
| Apo-C2 | -0.008 | 0.016 | 0.637 | -0.001 | 0.017 | 0.941 | 0.020 | 0.018 | 0.271 | -0.004 | 0.003 | 0.179 |
| Betaine | -0.007 | 0.007 | 0.324 | 0.005 | 0.007 | 0.544 | -0.010 | 0.008 | 0.215 | 0.001 | 0.001 | 0.375 |
| Trimethylamine oxide | -0.006 | 0.008 | 0.445 | -0.007 | 0.009 | 0.447 | -0.002 | 0.010 | 0.825 | -0.001 | 0.002 | 0.689 |
| Choline | 0.000 | 0.009 | 0.995 | 0.009 | 0.009 | 0.301 | -0.002 | 0.010 | 0.877 | 0.000 | 0.002 | 0.872 |
| Carnitine | 0.007 | 0.007 | 0.312 | -0.009 | 0.007 | 0.253 | -0.009 | 0.008 | 0.290 | 0.000 | 0.001 | 0.776 |
| Trimethylpentaaminovaleric acid | -0.001 | 0.005 | 0.786 | -0.004 | 0.005 | 0.408 | 0.006 | 0.006 | 0.289 | 0.000 | 0.001 | 0.835 |
| Butyl betaine | 0.007 | 0.007 | 0.274 | 0.001 | 0.007 | 0.837 | -0.001 | 0.008 | 0.933 | 0.000 | 0.001 | 0.785 |
| Trimethyllysine | -0.005 | 0.011 | 0.633 | -0.009 | 0.012 | 0.441 | 0.007 | 0.013 | 0.589 | 0.001 | 0.002 | 0.806 |

**Table S14a. Multi-variables regression of 38 biomarkers and Delta-NIHSS$_{(admission-discharge)}$ in scenario 2**

| | Delta-NIHSS | | | Delta-NIH1A | | | Delta-NIH1B | | | Delta-NIH1C | | |
|---|---|---|---|---|---|---|---|---|---|---|---|---|
| variables | Estimate | SE | P-value | Estimate | SE | P-value | Estimate | SE | P-value | Estimate | SE | P-value |
| (Intercept) | 0.099 | 0.005 | 0.000 | 0.012 | 0.002 | 0.000 | 0.015 | 0.002 | 0.000 | 0.013 | 0.002 | 0.000 |
| Adiponectin | -0.002 | 0.005 | 0.637 | 0.000 | 0.002 | 0.840 | 0.005 | 0.002 | 0.009 | 0.001 | 0.002 | 0.399 |
| ANGPTL3 | -0.009 | 0.005 | 0.066 | -0.002 | 0.002 | 0.264 | -0.001 | 0.002 | 0.598 | 0.001 | 0.002 | 0.453 |
| Vitamin B12 | 0.005 | 0.005 | 0.279 | 0.001 | 0.002 | 0.370 | 0.004 | 0.002 | 0.022 | 0.004 | 0.002 | 0.013 |
| Folic acid | 0.009 | 0.006 | 0.095 | 0.000 | 0.002 | 0.912 | 0.003 | 0.002 | 0.170 | -0.002 | 0.002 | 0.406 |
| Total cholesterol | -0.004 | 0.008 | 0.571 | 0.000 | 0.003 | 0.899 | -0.001 | 0.003 | 0.729 | 0.002 | 0.003 | 0.544 |
| Creatinine | -0.004 | 0.008 | 0.588 | -0.002 | 0.003 | 0.434 | -0.001 | 0.003 | 0.682 | -0.004 | 0.003 | 0.181 |
| Cystatin C | 0.002 | 0.005 | 0.761 | 0.002 | 0.002 | 0.341 | 0.006 | 0.002 | 0.001 | 0.004 | 0.002 | 0.018 |
| D-dimer DD | 0.006 | 0.005 | 0.255 | 0.000 | 0.002 | 0.805 | -0.001 | 0.002 | 0.592 | -0.003 | 0.002 | 0.138 |
| Fibrinogen Fib | 0.007 | 0.005 | 0.135 | 0.002 | 0.002 | 0.184 | 0.004 | 0.002 | 0.045 | 0.001 | 0.002 | 0.447 |
| Fasting blood glucose | 0.003 | 0.005 | 0.490 | 0.001 | 0.002 | 0.533 | 0.001 | 0.002 | 0.773 | 0.002 | 0.002 | 0.298 |
| Homocysteine HCY | -0.008 | 0.006 | 0.181 | 0.001 | 0.002 | 0.538 | 0.000 | 0.002 | 0.967 | 0.000 | 0.002 | 0.931 |
| High-density lipoprotein | 0.007 | 0.005 | 0.127 | 0.001 | 0.002 | 0.379 | 0.004 | 0.002 | 0.025 | -0.001 | 0.002 | 0.431 |
| High-sensitivity C-reactive protein | 0.010 | 0.005 | 0.063 | 0.000 | 0.002 | 0.938 | 0.003 | 0.002 | 0.199 | 0.000 | 0.002 | 0.896 |
| Interleukin-1 Receptor Antagonist | 0.006 | 0.005 | 0.271 | 0.008 | 0.002 | 0.000 | 0.003 | 0.002 | 0.231 | 0.007 | 0.002 | 0.001 |
| Interleukin-6 receptor Antagonist | -0.007 | 0.005 | 0.113 | -0.002 | 0.002 | 0.305 | 0.001 | 0.002 | 0.749 | 0.002 | 0.002 | 0.275 |
| Interleukin-6 receptor | -0.001 | 0.006 | 0.865 | 0.001 | 0.002 | 0.610 | -0.001 | 0.002 | 0.691 | -0.001 | 0.002 | 0.578 |
| Low-density lipoprotein | -0.008 | 0.005 | 0.140 | -0.001 | 0.002 | 0.695 | -0.001 | 0.002 | 0.571 | 0.001 | 0.002 | 0.782 |
| LDL Receptor | -0.008 | 0.007 | 0.267 | 0.002 | 0.003 | 0.341 | 0.001 | 0.003 | 0.863 | 0.004 | 0.003 | 0.186 |
| Lipoprotein phospholipase A2-activity | 0.017 | 0.007 | 0.015 | 0.000 | 0.003 | 0.995 | 0.003 | 0.003 | 0.253 | 0.000 | 0.003 | 0.891 |

| | | | | | | | | | | | | |
|---|---|---|---|---|---|---|---|---|---|---|---|---|
| Lipoprotein phospholipase A2-MASS | -0.003 | 0.004 | 0.447 | 0.004 | 0.002 | 0.015 | 0.000 | 0.002 | 0.810 | 0.000 | 0.002 | 0.816 |
| Monocyte Chemoattractant Protein-1 | 0.000 | 0.005 | 0.981 | -0.003 | 0.002 | 0.102 | -0.002 | 0.002 | 0.415 | 0.002 | 0.002 | 0.158 |
| Lipoprotein a | 0.002 | 0.004 | 0.624 | 0.000 | 0.001 | 0.967 | 0.001 | 0.002 | 0.651 | 0.001 | 0.001 | 0.585 |
| Methylmalonic acid | 0.001 | 0.005 | 0.894 | 0.000 | 0.002 | 0.948 | -0.002 | 0.002 | 0.324 | -0.001 | 0.002 | 0.781 |
| Proprotein Convertase Subtilisin 9 | -0.012 | 0.008 | 0.156 | -0.001 | 0.003 | 0.683 | -0.004 | 0.003 | 0.280 | -0.003 | 0.003 | 0.280 |
| Triglyceride | 0.008 | 0.005 | 0.101 | 0.003 | 0.002 | 0.111 | -0.001 | 0.002 | 0.752 | 0.004 | 0.002 | 0.026 |
| Human chitinase 3-like protein 1 | -0.001 | 0.005 | 0.889 | 0.000 | 0.002 | 0.877 | -0.001 | 0.002 | 0.521 | -0.001 | 0.002 | 0.604 |
| Apo-A2 | 0.006 | 0.012 | 0.610 | 0.001 | 0.004 | 0.740 | 0.003 | 0.005 | 0.549 | -0.001 | 0.005 | 0.797 |
| Apo-C3 | -0.001 | 0.008 | 0.886 | 0.002 | 0.003 | 0.610 | 0.001 | 0.003 | 0.855 | 0.000 | 0.003 | 0.969 |
| Apo-E | 0.005 | 0.007 | 0.432 | -0.001 | 0.002 | 0.644 | 0.000 | 0.003 | 0.884 | 0.001 | 0.003 | 0.568 |
| Apo-A2 | -0.001 | 0.007 | 0.897 | -0.003 | 0.002 | 0.288 | 0.000 | 0.003 | 0.974 | -0.004 | 0.003 | 0.096 |
| Apo-B | -0.007 | 0.011 | 0.527 | -0.003 | 0.004 | 0.490 | -0.002 | 0.004 | 0.593 | -0.002 | 0.004 | 0.623 |
| Apo-C2 | -0.011 | 0.007 | 0.139 | -0.001 | 0.003 | 0.620 | 0.001 | 0.003 | 0.770 | -0.004 | 0.003 | 0.157 |
| Betaine | -0.001 | 0.006 | 0.853 | -0.001 | 0.002 | 0.734 | 0.001 | 0.002 | 0.759 | -0.002 | 0.002 | 0.370 |
| Trimethylamine oxide | 0.003 | 0.006 | 0.573 | 0.000 | 0.002 | 0.907 | -0.002 | 0.002 | 0.445 | -0.001 | 0.002 | 0.771 |
| Choline | 0.012 | 0.005 | 0.012 | 0.003 | 0.002 | 0.091 | 0.004 | 0.002 | 0.044 | 0.004 | 0.002 | 0.021 |
| Carnitine | 0.000 | 0.004 | 0.945 | -0.001 | 0.001 | 0.692 | 0.000 | 0.002 | 0.974 | -0.001 | 0.002 | 0.616 |
| Trimethylpentaaminovaleric acid | 0.001 | 0.005 | 0.785 | 0.000 | 0.002 | 0.978 | -0.001 | 0.002 | 0.775 | 0.002 | 0.002 | 0.332 |
| Butyl betaine | -0.001 | 0.007 | 0.935 | -0.002 | 0.003 | 0.408 | 0.000 | 0.003 | 0.992 | -0.001 | 0.003 | 0.789 |

**Table S14b. Multi-variables regression of 38 biomarkers and Delta-NIHSS$_{(admission-discharge)}$ in scenario 2**

| | Delta-NIH2 | | | Delta-NIH3 | | | Delta-NIH4 | | | Delta-NIH5A | | |
|---|---|---|---|---|---|---|---|---|---|---|---|---|
| variables | Estimate | SE | P-value | Estimate | SE | P-value | Estimate | SE | P-value | Estimate | SE | P-value |
| (Intercept) | 0.008 | 0.001 | 0.000 | 0.009 | 0.001 | 0.000 | 0.063 | 0.004 | 0.000 | 0.047 | 0.003 | 0.000 |
| Adiponectin | 0.000 | 0.001 | 0.988 | 0.002 | 0.001 | 0.165 | -0.004 | 0.004 | 0.331 | -0.002 | 0.003 | 0.548 |
| ANGPTL3 | -0.001 | 0.001 | 0.396 | -0.002 | 0.002 | 0.121 | -0.001 | 0.004 | 0.776 | -0.006 | 0.003 | 0.078 |
| Vitamin B12 | 0.000 | 0.001 | 0.971 | 0.003 | 0.001 | 0.072 | 0.006 | 0.004 | 0.101 | 0.003 | 0.003 | 0.332 |
| Folic acid | -0.001 | 0.002 | 0.410 | -0.001 | 0.002 | 0.647 | 0.009 | 0.005 | 0.045 | -0.002 | 0.004 | 0.691 |
| Total cholesterol | 0.000 | 0.002 | 0.981 | 0.001 | 0.002 | 0.618 | -0.003 | 0.006 | 0.623 | -0.009 | 0.005 | 0.103 |
| Creatinine | -0.001 | 0.002 | 0.728 | -0.003 | 0.002 | 0.301 | 0.001 | 0.006 | 0.875 | 0.008 | 0.005 | 0.136 |
| Cystatin C | 0.001 | 0.001 | 0.656 | -0.001 | 0.002 | 0.548 | -0.003 | 0.004 | 0.512 | 0.004 | 0.003 | 0.225 |
| D-dimer DD | 0.000 | 0.002 | 0.822 | 0.000 | 0.002 | 0.767 | 0.005 | 0.004 | 0.268 | 0.001 | 0.004 | 0.836 |
| Fibrinogen Fib | 0.003 | 0.001 | 0.024 | 0.003 | 0.002 | 0.025 | 0.001 | 0.004 | 0.858 | 0.005 | 0.003 | 0.176 |
| Fasting blood glucose | -0.001 | 0.001 | 0.390 | 0.003 | 0.002 | 0.036 | 0.001 | 0.004 | 0.752 | -0.001 | 0.003 | 0.799 |
| Homocysteine HCY | 0.000 | 0.002 | 0.920 | 0.000 | 0.002 | 0.840 | 0.002 | 0.005 | 0.614 | 0.001 | 0.004 | 0.721 |
| High-density lipoprotein | 0.002 | 0.001 | 0.094 | -0.001 | 0.001 | 0.608 | -0.002 | 0.004 | 0.592 | -0.001 | 0.003 | 0.841 |
| High-sensitivity C-reactive protein | 0.002 | 0.002 | 0.212 | -0.003 | 0.002 | 0.091 | 0.005 | 0.004 | 0.238 | 0.002 | 0.004 | 0.675 |
| Interleukin-1 Receptor Antagonist | 0.002 | 0.002 | 0.173 | 0.004 | 0.002 | 0.033 | 0.010 | 0.004 | 0.024 | 0.008 | 0.004 | 0.045 |
| Interleukin-6 receptor Antagonist | 0.001 | 0.001 | 0.523 | -0.001 | 0.001 | 0.436 | 0.001 | 0.004 | 0.833 | -0.002 | 0.003 | 0.496 |
| Interleukin-6 receptor | 0.002 | 0.002 | 0.363 | -0.002 | 0.002 | 0.328 | 0.000 | 0.005 | 0.996 | 0.003 | 0.004 | 0.541 |
| Low-density lipoprotein | 0.000 | 0.002 | 0.929 | 0.000 | 0.002 | 0.868 | 0.006 | 0.004 | 0.170 | -0.007 | 0.004 | 0.089 |
| LDL Receptor | 0.004 | 0.002 | 0.078 | 0.001 | 0.002 | 0.754 | -0.009 | 0.006 | 0.140 | 0.008 | 0.005 | 0.107 |
| Lipoprotein phospholipase A2-activity | -0.002 | 0.002 | 0.423 | 0.002 | 0.002 | 0.267 | 0.010 | 0.006 | 0.073 | -0.005 | 0.005 | 0.270 |

| | | | | | | | | | | | | |
|---|---|---|---|---|---|---|---|---|---|---|---|---|
| Lipoprotein phospholipase A2-MASS | -0.001 | 0.001 | 0.469 | 0.000 | 0.001 | 0.767 | -0.003 | 0.004 | 0.487 | -0.001 | 0.003 | 0.739 |
| Monocyte Chemoattractant Protein-1 | 0.000 | 0.001 | 0.826 | 0.000 | 0.001 | 0.804 | 0.001 | 0.004 | 0.793 | 0.006 | 0.003 | 0.058 |
| Lipoprotein a | 0.001 | 0.001 | 0.456 | 0.000 | 0.001 | 0.718 | -0.003 | 0.003 | 0.385 | -0.001 | 0.003 | 0.679 |
| Methylmalonic acid | -0.001 | 0.001 | 0.337 | 0.001 | 0.002 | 0.531 | 0.002 | 0.004 | 0.681 | -0.001 | 0.003 | 0.767 |
| Proprotein Convertase Subtilisin 9 | 0.000 | 0.002 | 0.878 | -0.002 | 0.003 | 0.523 | -0.009 | 0.007 | 0.186 | 0.005 | 0.006 | 0.410 |
| Triglyceride | 0.002 | 0.002 | 0.190 | -0.001 | 0.002 | 0.576 | -0.002 | 0.004 | 0.549 | 0.003 | 0.004 | 0.410 |
| Human chitinase 3-like protein 1 | 0.000 | 0.001 | 0.798 | 0.000 | 0.002 | 0.822 | 0.013 | 0.004 | 0.001 | 0.000 | 0.003 | 0.929 |
| Apo-A2 | 0.001 | 0.004 | 0.792 | 0.001 | 0.004 | 0.793 | -0.003 | 0.010 | 0.745 | 0.000 | 0.008 | 0.966 |
| Apo-C3 | -0.002 | 0.003 | 0.441 | -0.002 | 0.003 | 0.455 | 0.003 | 0.007 | 0.710 | -0.001 | 0.006 | 0.931 |
| Apo-E | 0.000 | 0.002 | 0.951 | -0.003 | 0.002 | 0.198 | 0.000 | 0.006 | 0.984 | -0.001 | 0.005 | 0.840 |
| Apo-A2 | -0.002 | 0.002 | 0.387 | 0.001 | 0.002 | 0.788 | -0.004 | 0.006 | 0.508 | -0.009 | 0.005 | 0.072 |
| Apo-B | -0.002 | 0.003 | 0.482 | 0.002 | 0.004 | 0.544 | 0.000 | 0.009 | 0.980 | -0.002 | 0.008 | 0.847 |
| Apo-C2 | -0.005 | 0.002 | 0.030 | -0.005 | 0.002 | 0.027 | -0.002 | 0.006 | 0.752 | -0.012 | 0.005 | 0.014 |
| Betaine | -0.001 | 0.002 | 0.401 | -0.001 | 0.002 | 0.546 | 0.004 | 0.005 | 0.341 | -0.002 | 0.004 | 0.549 |
| Trimethylamine oxide | 0.000 | 0.002 | 0.917 | 0.002 | 0.002 | 0.413 | 0.000 | 0.005 | 0.956 | 0.002 | 0.004 | 0.710 |
| Choline | 0.002 | 0.001 | 0.301 | 0.005 | 0.002 | 0.002 | 0.003 | 0.004 | 0.488 | 0.005 | 0.003 | 0.152 |
| Carnitine | 0.000 | 0.001 | 0.721 | -0.001 | 0.001 | 0.500 | 0.001 | 0.003 | 0.822 | -0.001 | 0.003 | 0.682 |
| Trimethylpentaaminovaleric acid | -0.001 | 0.001 | 0.576 | 0.000 | 0.001 | 0.977 | 0.001 | 0.004 | 0.738 | 0.000 | 0.003 | 0.961 |
| Butyl betaine | 0.000 | 0.002 | 0.920 | -0.001 | 0.002 | 0.628 | 0.004 | 0.006 | 0.439 | -0.009 | 0.005 | 0.066 |

**Table S14c. Multi-variables regression of 38 biomarkers and Delta-NIHSS$_{(admission-discharge)}$ in scenario 2**

| | Delta-NIH5B | | | Delta-NIH6A | | | Delta-NIH6B | | | Delta-NIH7 | | |
|---|---|---|---|---|---|---|---|---|---|---|---|---|
| variables | Estimate | SE | P-value | Estimate | SE | P-value | Estimate | SE | P-value | Estimate | SE | P-value |
| (Intercept) | 0.180 | 0.006 | 0.000 | 0.188 | 0.006 | 0.000 | 0.184 | 0.006 | 0.000 | 0.099 | 0.005 | 0.000 |
| Adiponectin | -0.004 | 0.006 | 0.492 | 0.003 | 0.006 | 0.601 | -0.005 | 0.006 | 0.358 | -0.003 | 0.005 | 0.488 |
| ANGPTL3 | -0.006 | 0.006 | 0.301 | -0.015 | 0.006 | 0.014 | -0.009 | 0.006 | 0.139 | -0.021 | 0.005 | 0.000 |
| Vitamin B12 | 0.010 | 0.006 | 0.099 | 0.011 | 0.006 | 0.072 | 0.003 | 0.006 | 0.575 | 0.003 | 0.005 | 0.464 |
| Folic acid | 0.013 | 0.007 | 0.069 | -0.007 | 0.007 | 0.370 | 0.012 | 0.007 | 0.105 | 0.006 | 0.006 | 0.331 |
| Total cholesterol | 0.010 | 0.010 | 0.292 | -0.034 | 0.010 | 0.001 | 0.005 | 0.010 | 0.583 | -0.001 | 0.008 | 0.861 |
| Creatinine | -0.002 | 0.010 | 0.828 | 0.022 | 0.010 | 0.029 | 0.013 | 0.010 | 0.195 | -0.006 | 0.008 | 0.428 |
| Cystatin C | 0.005 | 0.006 | 0.419 | 0.012 | 0.006 | 0.064 | 0.003 | 0.006 | 0.630 | 0.002 | 0.005 | 0.647 |
| D-dimer DD | 0.014 | 0.006 | 0.028 | 0.003 | 0.007 | 0.628 | 0.018 | 0.007 | 0.005 | 0.008 | 0.005 | 0.120 |
| Fibrinogen Fib | 0.008 | 0.006 | 0.187 | 0.020 | 0.006 | 0.002 | 0.011 | 0.006 | 0.079 | 0.006 | 0.005 | 0.202 |
| Fasting blood glucose | 0.000 | 0.006 | 0.948 | 0.002 | 0.006 | 0.708 | -0.001 | 0.006 | 0.846 | -0.001 | 0.005 | 0.814 |
| Homocysteine HCY | 0.006 | 0.007 | 0.418 | 0.002 | 0.008 | 0.822 | 0.001 | 0.008 | 0.885 | -0.003 | 0.006 | 0.612 |
| High-density lipoprotein | 0.014 | 0.006 | 0.023 | 0.003 | 0.006 | 0.678 | 0.007 | 0.006 | 0.271 | 0.002 | 0.005 | 0.677 |
| High-sensitivity C-reactive protein | 0.008 | 0.007 | 0.216 | 0.015 | 0.007 | 0.026 | 0.005 | 0.007 | 0.507 | 0.010 | 0.005 | 0.058 |
| Interleukin-1 Receptor Antagonist | 0.016 | 0.007 | 0.021 | 0.036 | 0.007 | 0.000 | 0.012 | 0.007 | 0.083 | 0.010 | 0.005 | 0.060 |
| Interleukin-6 receptor Antagonist | -0.005 | 0.006 | 0.435 | -0.011 | 0.006 | 0.060 | -0.003 | 0.006 | 0.661 | 0.004 | 0.005 | 0.422 |
| Interleukin-6 receptor | -0.004 | 0.008 | 0.626 | 0.004 | 0.008 | 0.590 | -0.004 | 0.008 | 0.620 | 0.011 | 0.006 | 0.069 |
| Low-density lipoprotein | 0.022 | 0.007 | 0.002 | -0.008 | 0.007 | 0.232 | 0.010 | 0.007 | 0.154 | -0.015 | 0.005 | 0.006 |
| LDL Receptor | -0.008 | 0.009 | 0.376 | -0.005 | 0.009 | 0.618 | -0.005 | 0.009 | 0.562 | -0.004 | 0.007 | 0.603 |
| Lipoprotein phospholipase A2-activity | 0.008 | 0.009 | 0.393 | 0.004 | 0.009 | 0.668 | 0.001 | 0.009 | 0.919 | -0.001 | 0.007 | 0.881 |

| | | | | | | | | | | | | |
|---|---|---|---|---|---|---|---|---|---|---|---|---|
| Lipoprotein phospholipase A2-MASS | -0.004 | 0.006 | 0.500 | -0.001 | 0.006 | 0.863 | -0.002 | 0.006 | 0.782 | -0.006 | 0.004 | 0.198 |
| Monocyte Chemoattractant Protein-1 | 0.004 | 0.006 | 0.546 | -0.003 | 0.006 | 0.569 | 0.006 | 0.006 | 0.282 | 0.004 | 0.005 | 0.382 |
| Lipoprotein a | 0.001 | 0.005 | 0.762 | -0.002 | 0.005 | 0.661 | 0.000 | 0.005 | 0.921 | 0.006 | 0.004 | 0.132 |
| Methylmalonic acid | -0.022 | 0.006 | 0.000 | 0.005 | 0.006 | 0.436 | -0.017 | 0.006 | 0.007 | -0.004 | 0.005 | 0.462 |
| Proprotein Convertase Subtilisin 9 | -0.005 | 0.011 | 0.623 | -0.010 | 0.011 | 0.372 | -0.013 | 0.011 | 0.230 | -0.014 | 0.008 | 0.090 |
| Triglyceride | 0.008 | 0.006 | 0.219 | 0.000 | 0.007 | 0.965 | 0.022 | 0.007 | 0.001 | -0.003 | 0.005 | 0.565 |
| Human chitinase 3-like protein 1 | -0.006 | 0.006 | 0.316 | -0.004 | 0.006 | 0.545 | -0.003 | 0.006 | 0.598 | 0.000 | 0.005 | 0.947 |
| Apo-A2 | -0.020 | 0.015 | 0.191 | -0.004 | 0.015 | 0.785 | -0.017 | 0.015 | 0.270 | -0.003 | 0.012 | 0.813 |
| Apo-C3 | 0.017 | 0.011 | 0.108 | -0.011 | 0.011 | 0.315 | 0.016 | 0.011 | 0.144 | 0.015 | 0.008 | 0.080 |
| Apo-E | 0.012 | 0.009 | 0.161 | 0.001 | 0.009 | 0.911 | 0.000 | 0.009 | 0.990 | -0.003 | 0.007 | 0.698 |
| Apo-A2 | 0.014 | 0.009 | 0.103 | -0.002 | 0.009 | 0.806 | 0.023 | 0.009 | 0.008 | -0.004 | 0.007 | 0.509 |
| Apo-B | -0.016 | 0.014 | 0.268 | 0.026 | 0.014 | 0.069 | -0.006 | 0.014 | 0.661 | 0.013 | 0.011 | 0.249 |
| Apo-C2 | -0.022 | 0.009 | 0.015 | -0.020 | 0.009 | 0.029 | -0.017 | 0.009 | 0.061 | -0.010 | 0.007 | 0.147 |
| Betaine | -0.016 | 0.007 | 0.028 | -0.017 | 0.007 | 0.015 | -0.017 | 0.007 | 0.018 | 0.005 | 0.006 | 0.387 |
| Trimethylamine oxide | 0.005 | 0.008 | 0.525 | -0.001 | 0.008 | 0.944 | -0.001 | 0.008 | 0.879 | 0.004 | 0.006 | 0.510 |
| Choline | 0.003 | 0.006 | 0.628 | -0.004 | 0.006 | 0.532 | 0.010 | 0.006 | 0.104 | 0.015 | 0.005 | 0.002 |
| Carnitine | -0.005 | 0.005 | 0.365 | 0.006 | 0.005 | 0.237 | -0.006 | 0.005 | 0.243 | 0.002 | 0.004 | 0.618 |
| Trimethylpentaaminovaleric acid | 0.009 | 0.006 | 0.131 | 0.005 | 0.006 | 0.414 | 0.004 | 0.006 | 0.508 | -0.005 | 0.005 | 0.259 |
| Butyl betaine | -0.015 | 0.009 | 0.107 | 0.006 | 0.009 | 0.521 | -0.017 | 0.009 | 0.058 | -0.005 | 0.007 | 0.498 |

**Table S14d. Multi-variables regression of 38 biomarkers and Delta-NIHSS$_{(admission-discharge)}$ in scenario 2**

| | Delta-NIH8 | | | Delta-NIH9 | | | Delta-NIH10 | | | Delta-NIH11 | | |
|---|---|---|---|---|---|---|---|---|---|---|---|---|
| variables | Estimate | SE | P-value | Estimate | SE | P-value | Estimate | SE | P-value | Estimate | SE | P-value |
| (Intercept) | 0.161 | 0.006 | 0.000 | 0.175 | 0.006 | 0.000 | 0.219 | 0.006 | 0.000 | 0.004 | 0.001 | 0.000 |
| Adiponectin | 0.004 | 0.006 | 0.494 | 0.002 | 0.006 | 0.769 | -0.007 | 0.006 | 0.236 | 0.000 | 0.001 | 0.681 |
| ANGPTL3 | 0.006 | 0.006 | 0.344 | -0.009 | 0.006 | 0.151 | -0.003 | 0.007 | 0.655 | 0.000 | 0.001 | 0.715 |
| Vitamin B12 | 0.018 | 0.006 | 0.001 | -0.002 | 0.006 | 0.679 | -0.006 | 0.006 | 0.335 | 0.001 | 0.001 | 0.462 |
| Folic acid | -0.009 | 0.007 | 0.221 | -0.001 | 0.007 | 0.849 | 0.018 | 0.008 | 0.023 | -0.001 | 0.001 | 0.245 |
| Total cholesterol | -0.004 | 0.009 | 0.680 | 0.003 | 0.010 | 0.759 | 0.009 | 0.011 | 0.402 | 0.001 | 0.002 | 0.471 |
| Creatinine | -0.006 | 0.009 | 0.555 | -0.025 | 0.010 | 0.010 | -0.008 | 0.011 | 0.453 | 0.000 | 0.002 | 0.998 |
| Cystatin C | -0.001 | 0.006 | 0.933 | 0.004 | 0.006 | 0.511 | -0.012 | 0.007 | 0.071 | 0.000 | 0.001 | 0.829 |
| D-dimer DD | -0.001 | 0.006 | 0.895 | 0.008 | 0.006 | 0.210 | -0.014 | 0.007 | 0.053 | -0.002 | 0.001 | 0.163 |
| Fibrinogen Fib | 0.010 | 0.006 | 0.088 | 0.003 | 0.006 | 0.645 | -0.002 | 0.007 | 0.789 | 0.002 | 0.001 | 0.041 |
| Fasting blood glucose | 0.003 | 0.006 | 0.594 | 0.008 | 0.006 | 0.185 | -0.015 | 0.007 | 0.027 | 0.000 | 0.001 | 0.888 |
| Homocysteine HCY | -0.007 | 0.007 | 0.308 | 0.010 | 0.007 | 0.160 | -0.003 | 0.008 | 0.750 | 0.000 | 0.001 | 0.715 |
| High-density lipoprotein | 0.000 | 0.006 | 0.950 | 0.009 | 0.006 | 0.122 | 0.011 | 0.006 | 0.077 | 0.004 | 0.001 | 0.000 |
| High-sensitivity C-reactive protein | -0.004 | 0.007 | 0.497 | -0.002 | 0.007 | 0.774 | -0.004 | 0.007 | 0.590 | 0.001 | 0.001 | 0.510 |
| Interleukin-1 Receptor Antagonist | 0.012 | 0.007 | 0.071 | 0.017 | 0.007 | 0.010 | 0.023 | 0.007 | 0.002 | -0.001 | 0.001 | 0.466 |
| Interleukin-6 receptor Antagonist | 0.009 | 0.006 | 0.117 | 0.000 | 0.006 | 0.960 | -0.011 | 0.006 | 0.091 | 0.000 | 0.001 | 0.810 |
| Interleukin-6 receptor | 0.006 | 0.008 | 0.457 | 0.005 | 0.008 | 0.518 | 0.027 | 0.009 | 0.002 | -0.001 | 0.001 | 0.423 |
| Low-density lipoprotein | -0.009 | 0.007 | 0.181 | -0.008 | 0.007 | 0.229 | -0.018 | 0.008 | 0.014 | 0.001 | 0.001 | 0.414 |
| LDL Receptor | -0.004 | 0.009 | 0.677 | -0.005 | 0.009 | 0.569 | 0.005 | 0.010 | 0.638 | 0.000 | 0.002 | 0.888 |
| Lipoprotein phospholipase A2-activity | 0.007 | 0.009 | 0.426 | 0.004 | 0.009 | 0.636 | -0.015 | 0.010 | 0.110 | 0.001 | 0.002 | 0.596 |

| | | | | | | | | | | | | |
|---|---|---|---|---|---|---|---|---|---|---|---|---|
| Lipoprotein phospholipase A2-MASS | -0.010 | 0.006 | 0.076 | 0.001 | 0.006 | 0.887 | -0.006 | 0.006 | 0.344 | 0.000 | 0.001 | 0.755 |
| Monocyte Chemoattractant Protein-1 | -0.003 | 0.006 | 0.590 | -0.007 | 0.006 | 0.215 | 0.014 | 0.006 | 0.032 | 0.001 | 0.001 | 0.476 |
| Lipoprotein a | 0.007 | 0.005 | 0.115 | 0.003 | 0.005 | 0.553 | 0.002 | 0.005 | 0.704 | 0.001 | 0.001 | 0.506 |
| Methylmalonic acid | 0.010 | 0.006 | 0.096 | -0.015 | 0.006 | 0.011 | -0.011 | 0.007 | 0.103 | -0.001 | 0.001 | 0.248 |
| Proprotein Convertase Subtilisin 9 | 0.007 | 0.010 | 0.516 | 0.000 | 0.010 | 0.962 | 0.006 | 0.011 | 0.617 | -0.001 | 0.002 | 0.542 |
| Triglyceride | -0.007 | 0.006 | 0.248 | 0.006 | 0.006 | 0.390 | 0.007 | 0.007 | 0.294 | 0.002 | 0.001 | 0.162 |
| Human chitinase 3-like protein 1 | -0.002 | 0.006 | 0.693 | -0.001 | 0.006 | 0.884 | -0.009 | 0.007 | 0.190 | 0.000 | 0.001 | 0.838 |
| Apo-A2 | -0.013 | 0.015 | 0.380 | -0.010 | 0.015 | 0.519 | -0.039 | 0.016 | 0.018 | 0.003 | 0.003 | 0.283 |
| Apo-C3 | 0.014 | 0.010 | 0.162 | 0.021 | 0.011 | 0.048 | 0.011 | 0.012 | 0.358 | 0.000 | 0.002 | 0.892 |
| Apo-E | 0.010 | 0.008 | 0.243 | -0.018 | 0.009 | 0.038 | -0.002 | 0.009 | 0.843 | 0.001 | 0.002 | 0.522 |
| Apo-A2 | -0.010 | 0.008 | 0.237 | 0.022 | 0.009 | 0.010 | -0.007 | 0.009 | 0.452 | -0.001 | 0.002 | 0.330 |
| Apo-B | 0.003 | 0.014 | 0.834 | -0.008 | 0.014 | 0.555 | 0.030 | 0.015 | 0.050 | -0.003 | 0.002 | 0.182 |
| Apo-C2 | -0.012 | 0.009 | 0.173 | 0.004 | 0.009 | 0.645 | -0.034 | 0.010 | 0.001 | 0.002 | 0.002 | 0.193 |
| Betaine | -0.010 | 0.007 | 0.164 | -0.007 | 0.007 | 0.338 | -0.013 | 0.008 | 0.101 | 0.000 | 0.001 | 0.844 |
| Trimethylamine oxide | -0.003 | 0.008 | 0.700 | -0.004 | 0.008 | 0.653 | -0.002 | 0.008 | 0.814 | -0.002 | 0.001 | 0.243 |
| Choline | 0.003 | 0.006 | 0.581 | 0.003 | 0.006 | 0.615 | 0.010 | 0.007 | 0.134 | 0.000 | 0.001 | 0.653 |
| Carnitine | -0.002 | 0.005 | 0.715 | -0.001 | 0.005 | 0.775 | 0.003 | 0.006 | 0.602 | 0.000 | 0.001 | 0.979 |
| Trimethylpentaaminovaleric acid | 0.011 | 0.006 | 0.042 | 0.000 | 0.006 | 0.961 | -0.002 | 0.006 | 0.714 | 0.000 | 0.001 | 0.968 |
| Butyl betaine | -0.007 | 0.009 | 0.443 | -0.008 | 0.009 | 0.384 | 0.003 | 0.010 | 0.786 | 0.000 | 0.002 | 0.942 |
