## Supplementary material for "DISCO: A Delta-NIHSS-Based Machine Learning Model for Predicting Recurrence, Disability, and Mortality Following Acute Ischemic Stroke": Expanded Methods

### 1. Search strategies for literature review

A systematic literature search identified studies which evaluated the application of AI in predicting recurrence attributed to stroke. The last search was performed on 12 March 2024 using MEDLINE/PubMed and Web of Science databases with no language restrictions. The search terms used were intersections of "stroke", "artificial intelligence", "deep learning", "machine learning", "recurrence", " relapse ", and so on. The complete search strategies can be found below Table M1.

Table M1 The detail searching strategy about prediction model of stroke outcome

| Database | Search strategy |
| --- | --- |
| PubMed | "stroke"[MeSH Terms] OR "stroke"[All Fields] OR "CVA"[All Fields] OR "stroke*"[All Fields] OR ("stroke"[MeSH Terms] OR "stroke"[All Fields] OR ("cerebrovascular"[All Fields] AND "accident"[All Fields]) OR "cerebrovascular accident"[All Fields]) OR ("apoplexies"[All Fields] OR "stroke"[MeSH Terms] OR "stroke"[All Fields] OR "apoplexy"[All Fields]) OR "poststroke"[All Fields] OR ("stroke"[MeSH Terms] OR "stroke"[All Fields] OR ("brain"[All Fields] AND "attack"[All Fields]) OR "brain attack"[All Fields])<br>AND ("artificial intelligence"[Mesh] OR "artificial intelligence"[tiab] OR "deep learning"[tiab] OR "machine learning"[tiab] OR "neural network*"[tiab])<br>AND ("recurrence"[Mesh] OR " patient readmission "[Mesh] OR " recurrence "[tiab] OR " readmission "[tiab] OR " relapse "[tiab]) |
| Web of Science | ("artificial intelligence" OR "deep learning" OR "machine learning" OR "neural network") AND (stroke* OR "CVA" OR "cerebrovascular accident" OR "apoplexy" OR "poststroke" OR "brain attack") AND ("recurrence " OR " relapse ") |

### 2. PRS calculation

Genotype data in the CNSR-III were WGS data described in the STROMICS study (**Figure 1**)[1], constructed by the BGISEQ-500 platform. The average sequencing depth exceeded 30x for each subject, obviating the need for imputation. Based on that, we calculated the PRS for each ischemic stroke patient based on ischemic stroke susceptibility PRS model in Eastern Asia by utilizing the code on Github (https://github.com/ShujiaHuang/genotools/blob/master/scripts/mr.py).

### 3. RRE score calculation in CNSR-III group

Based on the variables described in RRE model, 7,261 ischemic stroke patients in our analysis group had the corresponding MRI data. Based on that data, we standardized our related variables, and calculated the RRE score on the given website (https://www.nmr.mgh.harvard.edu/RRE/). For the remaining ischemic stroke patients without the image data, we based on the history of TIA and CCS etiologic ischemic stroke subtype and calculated the RRE score on the website (https://www.nmr.mgh.harvard.edu/RRE/). In conclusion, 13,940 ischemic stroke patients in our analysis group with or without MRI data calculated the RRE score in the given website (https://www.nmr.mgh.harvard.edu/RRE/).

### 4. Details of follow-up in CNSR-III cohort

#### 4.1 the way of follow-up

Patients were interviewed face-to-face by experienced physicians within the first year, followed by telephone interviews conducted by trained clinical research coordinators (CRCs) thereafter. The transition from face-to-face to telephone follow-up was necessitated by the COVID-19 outbreak in 2019, following the closure of the CNSR-III registry in 2018. Additionally, if patients were unable to attend a face-to-face interview within the first year, telephone follow-ups were conducted instead.

Regardless of the mode of follow-up, patient evaluations adhered to the standardized operational manual of the follow-up case report form (CRF), as conducted by physicians at participating centers or trained CRCs via telephone. For reference, we have included an English version of the three-month follow-up CRF in the end of supplementary method "7. Case Report form in three months follow-up of CNSR-III cohort".

The CNSR-III cohort is one of the largest stroke registries in China, organized by the National Clinical Research Center in China for Neurological Diseases (https://ncrcnd.org.cn/English/). In our study, the integration of this robust dataset with advanced explainable AI methodologies, along with a rigorous validation design, ensures the reliability the DISCO model and underscores the significant predictive value of delta-NIHSS (admission NIHSS minus discharge NIHSS) in stroke prognosis.

Follow-up assessments were conducted within a 14-day window from each scheduled time point (i.e., 3 months [90 days], 6 months [180 days], and annually from 1 to 5 years for all patients). If a patient did not respond within the 14-day window and was not recorded as deceased, the follow-up was classified as missing data. For the primary endpoints of stroke recurrence, disability and death at three months, as well as the 6-month, 1-year follow ups- none of the participants had missing follow-up data. The follow-up completion rates exceeded 80% in subsequent 5 years.

### 5. XGBoost Model construction and SHAP Interpretation

#### 5.1 Framework for XGBoost model construction

XGBoost algorithm was used in the model construction for its scalability and adeptness with nonlinear, sparse, and class-imbalanced classification data. In order to ensure the robustness of the model and avoid overfitting, we employed a nested cross-validation approach to train, validate, and test prediction models.

This methodology involves two key components: an inner loop for hyperparameter tunning and an outer loop for model evaluation. The hyperparameters include max depth and eta (learning rate). In the outer loop, we began by dividing our dataset into two primary segments: 80% for the training set and 20% for the test set. In the inner loop, the optimal combination of hyperparameters is determined (tenfold cross-validation was adopted to ward off overfitting/underfitting, fortify model robustness, and mitigate bias). Specially, after the list of predictor variables is decided, we used a grid search for each classifier, assessing every possible combination of hyperparameters. The search for the selection of the best hyperparameters was based on achieving the maximum of the area under the receiver operating characteristic curve (AUC) on the validation dataset. In the second stage, we trained and evaluated our XGBoost classifiers within the outer loop of the nested cross-validation method, utilizing the best hyperparameters identified in the first step. In this phase, we assessed performance metrics using the test dataset. We repeated this method four times, using one of the remaining test folds as the testing dataset and the other as the training dataset in each iteration.

#### 5.2 Interpretation of the model using SHAP

SHAP values can enhance global and local interpretability for tree-based model[2]. It offers a novel approach to explaining various black-box ML models provides both local and global interpretability and possesses a more robust theoretical foundation compared to other methods. We utilized the SHAP value to gain feature importance, which combined with feature importance given by XGBoost and utilized for comprehensive consideration in feature selection, and deliver personalized explanation of features. During each round of the outer cross-validation, the SHAP values of the selected features are calculated on the test folds.

#### 5.3 Feature Selection for simplified prediction model (DISCO model)

By the framework for XGBoost model construction and SHAP interpretation described above, we constructed a model for all 308 features at first. In order to develop a predictive model that would be useful in clinical practice, we attempted to simplify the model. Based on feature importance rankings using SHAP value in models for the model of 308 features and expert suggestions, eight easy-assessed features, global and domain-specific delta-NIHSS were selected for simplified prediction model. Then, we constructed the Delta-NIHSS Integrated post-Stroke Composite Outcome (DISCO) model for stroke recurrence in 3 months and other secondary outcomes using the same framework for XGBoost model construction.

#### 5.4 Performance of prediction model

We use multiple metrics, including sensitivity, specificity, F1-score, and the area under the receiver operating characteristic curve (AUC) were used to evaluate the model performance. The Brier score was used to evaluate model calibration, and 95% confidence intervals (CI) rescaled Brier score using the 2.5th and 97.5th percentiles from 200 bootstrap samples as boundaries.

#### 5.5 External validation

To provide the evidence of generalizability of the DISCO model we develop, we conducted external validation by testing performance of the model in two extra independent cohorts 1 and 2.
Validation cohort 1 consisted of 2,127 patients from 40 independent hospitals in CNSR-III which were assigned to external in the initial randomization. Validation cohort 2 consisted of 5,158 ischemic stroke patients we selected after removing TIA in the CHANCE-2 cohort. CHANCE-2 is a cohort involved 6,412 patients who were enrolled in a randomized, double-blind, placebo-controlled trial conducted at 202 centers in China.[3] And this cohort utilized similar evaluation methods as CNSR-III with a 100% follow-up rate in 3 months. (**Table S5**) The follow-up data collected in 3 months and 1 year is utilized for external validation. Based on the “pwr" package in R, the sample size in three cohorts is big enough for the prediction model. Data collection processes and informed consent from patients were presented in the original CNSR-III and CHANCE-2 studies.

#### 5.6 Comparison of predictive performance of multiple prediction models

Firstly, we compare different machine learning classifiers with our XGBoost algorithm. Five algorithms including logistic regression, naive Bayes, random forest (RF), and support vector machine (SVM) were used to carry out the construction of predictive models using the same features and model evaluation using the same metrics. Secondly, we compared the predictive models built with different features. Specially, we constructed the prediction model using the all 308 variables, the 8 features of DISCO, the biomarkers, and the top 20 features of SHAP as input features, respectively. In addition, a head-to-head comparison with the RRE model in the CNSR-III cohort was performed (Supplementary Method). Thirdly, to investigate whether PRS improves the prediction of stroke outcome, we compared the performances of three models, including a pure PRS model, DISCO model, and a PRS + PRS model in CNSR-III. And patients in CNSR-III with WGS data were recruited to calculate PRS (**Figure 1**).[1] The Delong test was utilized to evaluate the difference between the two models.[4]

### 6. The R packages utilized in method part

The analysis is based on R version 4.2.0 and the related R packages is described below in Table M1.

Table M1 the R Packages utilized in our analysis

| Package Name | Version | Date | URL |
|---|---|---|---|
| xgboost | 1.6.0.1 | 2022/4/16 | https://github.com/dmlc/xgboost |
| SHAPforxgboost | 0.1.3 | 2023/5/18 | https://github.com/liuyanguu/SHAPforxgboost |
| survival | 3.2-13 | 2021/8/23 | https://github.com/therneau/survival |
| survminer | 0.4.9 | 2021/3/9 | https://rpkhs.datanovia.com/survminer/index.html |
| Ckmeans.1d.dp | 4.3.4 | 2022/1/30 | NA |
| shapviz | 0.9.3 | 2024/1/12 | https://hithub.com/ModelOriented/shapviz |
| ggplot2 | 3.4.2 | 2023/4/3 | https://ggplot2.tidyverse.org |
| caret | 6.0-94 | 2023/3/21 | https://github.com/topepo/caret |
| pROC | 1.18.0 | 2021/9/2 | http://expasy.org/tools/pROC |

### 7. Case report form in 3 months follow-up of CNSR-III cohort

Visit 3: 3-Month Follow-Up (Face-to-Face Visit)

At the 3-month follow-up after the patient's onset, you are required to complete the following sections:

| |
|---|
| M3: Follow-Up Basic Information |
| N3: Follow-Up Events |
| O3: Adherence to Secondary Prevention Medications |
| P3: Clinical Information |

M3. Follow-Up Basic Information

Patient follow-up method:

○1 - Face-to-face visit ○2 - Telephone visit ○3 - Uncompleted visit

If the visit is completed, please proceed with the following questionnaires.

N3. Follow-Up Events

N3 - 1: Has the patient died? ○1 - Yes ○2 - No

N3 - 1.1: If the patient has died, then:

N3 - 1.1.1: Date of death: □□□□ year □□ month □□ day

N3 - 1.1.2: Cause of death:

○1 - Cardiovascular death

○1 - Ischemic stroke

○2 - Hemorrhagic stroke

○3 - Sudden cardiac death

○4 - Death due to acute myocardial infarction

○5 - Death due to heart failure

○6 - Other cardiovascular deaths

○2 - Non-vascular death (e.g., cancer, pneumonia, accident, etc.)

○3 - Unknown cause

Cardiovascular Deaths include deaths caused by stroke, sudden cardiac death, acute myocardial infarction, heart failure, or other cardiovascular reasons. Other cardiovascular-related deaths include arrhythmias unrelated to sudden cardiac death, pulmonary embolism, cardiovascular interventions (unrelated to acute MI), aortic aneurysm rupture, or peripheral artery disease.

Non-Cardiovascular Deaths include deaths caused by bleeding (including gastrointestinal bleeding), pulmonary causes (respiratory failure, pneumonia), malignancies, trauma, suicide, infection/sepsis, or any other well-defined causes (e.g., liver or kidney failure).

Unknown Cause refers to deaths where the cause cannot be determined.

Note: Any death within 30 days of stroke, MI, or a procedure/surgery with an unknown/unclear cause will be attributed to stroke, MI, or the procedure/surgery, respectively.

N3 - 1.2: If the patient is alive, then:

N3 - 1.2.1: Neurological function score (mRS) at follow-up:

○0 - No symptoms at all

○1 - Although there are symptoms, no obvious disability; able to perform all usual duties and activities

○2 - Mild disability; unable to perform all previous activities but can handle personal affairs without assistance

○3 - Moderate disability; requires some assistance but can walk without assistance

○4 - Severe disability; unable to walk or attend to bodily needs without assistance

○5 - Profound disability; bedridden, incontinent, and requiring constant nursing and care

N3 - 1.2.2: Has there been a non - fatal stroke recurrence?

○1 - No ○2 - Yes

If "Yes", the number of stroke recurrences: □ times

Type of the first stroke recurrence:

○1 - Cerebral infarction

○2 - Cerebral hemorrhage

○3 - Subarachnoid hemorrhage

Date of first stroke recurrence: □□□□ year □□ month □□ day

N3-1.2.3. Has a TIA Occurred? ○1 - No ○2 - Yes

If "Yes", the number of TIA episodes: □□ times

Date of the first TIA episode: □□□□ Year □□ Month □□ Day

N3-1.2.4. Has a Non-Fatal Myocardial Infarction Occurred?

○1 - No ○2 - Yes

If "Yes", the date of the first episode: □□□□ Year □□ Month □□ Day

N3-1.2.5. Has Non-Fatal Heart Failure Occurred? ○1 - No ○2 - Yes

If "Yes", the date of the first episode: □□□□ Year □□ Month □□ Day

N3-1.2.6. Has Any Vascular-Related Procedure or Surgery Been Performed?

○1 - No ○2 - Yes

If "Yes", the name of the procedure/surgery:

□1 - Carotid Artery Stenting (CAS)

□2 - Carotid Endarterectomy (CEA)

□3 - Intracranial Artery Stenting

□4 - Coronary Artery Stenting and Balloon Angioplasty

□99 - Other

N3-1.2.7. Has a Systemic Embolic Event Occurred? ○1 - No ○2 - Yes

Systemic Embolic Event: An arterial embolism causing clinical ischemic events outside the central nervous system, coronary, and pulmonary circulation. Evidence of embolism may be supported by surgical specimens, autopsy, angiography, or other objective examinations, such as mesenteric artery embolism, splenic artery embolism, etc.

O3. Secondary Prevention Medication Adherence

O3 - 1: Current medications

O3 - 1.1: Anti - platelet drugs ○1 - No ○2 - Yes

If "Yes", the types of drugs:

□1 - Aspirin

□2 - Clopidogrel

□3 - Dipyridamole

□4 - Cilostazol

□99 - Others

Aspirin dosage: □□□mg/day; Clopidogrel dosage: □□□mg/day

O3 - 1.2: Anticoagulant drugs ○1 - No ○2 - Yes

If "Yes", the types of drugs:

□1 - Warfarin

□2 - Low - molecular - weight heparin

□3 - Unfractionated heparin

□4 - Rivaroxaban

□5 - Dabigatran

□6 - Apixaban

□99 - Others

O3 - 1.3: Lipid - regulating drugs ○1 - No ○2 - Yes

If "Yes", the types of drugs:

□1 - Statins (such as atorvastatin calcium, etc.)

□1 - Atorvastatin: □□mg/day

□2 - Rosuvastatin: □□mg/day

□3 - Simvastatin: □□mg/day

□4 - Pravastatin: □□mg/day

□5 - Lovastatin: □□mg/day

□6 - Fluvastatin: □□mg/day

□7 - Pitavastatin: □□mg/day

□2 - Fibrates (such as fenofibrate, gemfibrozil, etc.)

□3 - Bile acid sequestrants (such as cholestyramine, etc.)

□4 - Niacin and its derivatives (such as acipimox, etc.)

□5 - Zhibituo

□99 - Others

O3 - 1.4: Hypoglycemic treatment ○1 - No ○2 - Yes

If "Yes", the types of drugs:

□1 - Insulin

□2 - Biguanides

□3 - Sulfonylureas (such as glipizide, gliquidone, etc.)

□4 - α - Glucosidase inhibitors (such as acarbose, etc.)

□5 - Thiazolidinediones (such as rosiglitazone, etc.)

□6 - Meglitinides (such as repaglinide, etc.)

□99 - Others

O3 - 1.5: Antihypertensive treatment ○1 - No ○2 - Yes

If "Yes", the types of drugs:

□1 - Calcium channel blockers

□2 - Angiotensin - converting enzyme inhibitors (ACEI)

□3 - Angiotensin II receptor blockers (ARB)

□4 - Diuretics

□5 - β - Blockers

□6 - α - Blockers

□7 - α,β - Blockers

□99 - Others

O3 - 1.6: Anti - lipid oxidants ○1 - No ○2 - Yes

If "Yes", the types of drugs:

□1 - Probucol □2 - Others

If taking "Probucol", the dosage:

○1 - 250mg/day

○2 - 500mg/day

○3 - 750mg/day

○4 - 1000mg/day

If taking "Probucol", the cumulative medication time:

○1 - Less than 1 month

○2 - 1 - 2 months

○3 - More than 2 months

O3 - 2: Adherence to Discharge Medications

| Discharge Medications (Auto-Matched) | Currently Taking | If "Yes", select | If "No", select the reason |
|---|---|---|---|
| ———— | ○ 1 - No<br>○ 2 - Yes | ○ 1 = Never Forget<br>○ 2 = Rarely<br>○ 3 = Sometimes<br>○ 4 = Often | ○ 1 - Doctor Discontinued<br>○ 2 - Severe Side Effects<br>○ 3 - High Cost<br>○ 4 - Perceived Ineffectiveness<br>○ 99 - Other |

| | | | |
|---|---|---|---|
| ________ | ○ 1 - No<br>○ 2 - Yes | ○ 1 = Never Forget<br>○ 2 = Rarely<br>○ 3 = Sometimes<br>○ 4 = Often | ○ 1 - Doctor Discontinued<br>○ 2 - Severe Side Effects<br>○ 3 - High Cost<br>○ 4 - Perceived Ineffectiveness<br>○ 99 - Other |
| ________<br><br>________ | ○ 1 - No<br>○ 2 - Yes | ○ 1 = Never Forget<br>○ 2 = Rarely<br>○ 3 = Sometimes<br>○ 4 = Often | ○ 1 - Doctor Discontinued<br>○ 2 - Severe Side Effects<br>○ 3 - High Cost<br>○ 4 - Perceived Ineffectiveness<br>○ 99 - Other |
| ________ | ○ 1 - No<br>○ 2 - Yes | ○ 1 = Never Forget<br>○ 2 = Rarely<br>○ 3 = Sometimes<br>○ 4 = Often | ○ 1 - Doctor Discontinued<br>○ 2 - Severe Side Effects<br>○ 3 - High Cost<br>○ 4 - Perceived Ineffectiveness<br>○ 99 - Other |

Note: The degree of forgetting is divided into four categories: 1 = Never forget; 2 = Rarely forget 1 time/week; 3 = Sometimes forget 2 - 3 times/week; 4 = Often forget 4 times or more/week.

P3. Clinical Information

P3 - 1: Has blood pressure measurement been completed? ○1 - No ○2 - Yes

Left side: Blood pressure value: □□□/□□□mmHg

Right side: Blood pressure value: □□□/□□□mmHg

P3 - 2: Has the 3 - month blood specimen collection (fasting blood in the morning) been completed? ○1 - No ○2 - Yes

Blood specimen collection time: □□□□ year □□ month □□ day □□ o'clock

P3 - 3: Has the 3 - month urine specimen collection (random time) been completed? ○1 - No ○2 - Yes

Urine specimen collection time: □□□□ year □□ month □□ day □□ o'clock

P3 - 4: Has lipid measurement been completed? ○1 - No ○2 - Yes

| | | | | |
|---|---|---|---|---|
| Triglycerides (TG) | | mmol/L | | mg/dl |
| Total Cholesterol (CHO) | | mmol/L | | mg/dl |
| High-Density Lipoprotein (HDL) | | mmol/L | | mg/dl |
| Low-Density Lipoprotein (LDL) | | mmol/L | | mg/dl |

P3 - 5: Has blood glucose monitoring been completed? ○1 - No ○2 - Yes

□1 - Fasting blood glucose: □□.□mmol/L

□2 - Random blood glucose: □□.□mmol/L

P3 - 6: Has glycated hemoglobin been measured? ○1 - No ○2 - Yes

□.□%

P3 - 7: Living status at follow-up:

○1 - Living alone

○2 - Living with others

If living with others, the co - residents are:

□1 - Children

□2 - Spouse

□3 - Parents

□4 - Relatives

□5 - Nannies or other caregivers

□99 - Others

P3 - 8: Has the patient received professional rehabilitation training since discharge?

○1 - No ○2 - Yes

P3 - 8.1: If "Yes", the person providing rehabilitation training:

□1 - Professional rehabilitation trainers

□2 - Relatives

□3 - Nannies or other caregivers

□99 - Others

P3 - 8.2: If "Yes", the frequency of rehabilitation training:

○1 - Daily

○2 - 2 - 6 times a week

○3 - 1 - 4 times a month

○98 - Unknown

P3 - 8.3: If "Yes", the duration of each rehabilitation training session:

○1 - ≥1 hour

○2 - 30 minutes - 1 hour

○3 - <30 minutes

○98 - Unknown

Signature of the doctor who completed Visit 3: ______________________________

Date of completion of Visit 3: □□□□ year □□ month □□ day

### Reference:

1. Cheng S, Xu Z, Bian S, et al. The STROMICS genome study: deep whole-genome sequencing and analysis of 10K Chinese patients with ischemic stroke reveal complex genetic and phenotypic interplay. *Cell Discov*. 2023;9(1):75. doi:10.1038/s41421-023-00582-8

2. Lundberg SM, Erion G, Chen H, et al. From local explanations to global understanding with explainable AI for trees. *Nat Mach Intell*. 2020;2(1):56-67. doi:10.1038/s42256-019-0138-9

3. Wang Yongjun, Meng Xia, Wang Anxin, et al. Ticagrelor versus Clopidogrel in CYP2C19 Loss-of-Function Carriers with Stroke or TIA. *New England Journal of Medicine*. 2021;385(27):2520-2530. doi:10.1056/NEJMoa2111749

4. DeLong ER, DeLong DM, Clarke-Pearson DL. Comparing the areas under two or more correlated receiver operating characteristic curves: a nonparametric approach. *Biometrics*. 1988;44(3):837-845.
